# Comparing trivalent and quadrivalent seasonal influenza vaccine efficacy in persons 60 years of age and older: A systematic review and network meta-analysis

**DOI:** 10.1101/2023.11.29.23299123

**Authors:** Areti Angeliki Veroniki, Sai Surabi Thirugnanasampanthar, Menelaos Konstantinidis, Jasmeen Dourka, Marco Ghassemi, Dipika Neupane, Paul A. Khan, Vera Nincic, Margarita Corry, Reid Robson, Amanda Parker, Charlene Soobiah, Angela Sinilaite, Pamela Doyon-Plourde, Anabel Gil, Winnie Siu, Nasheed Moqueet, Adrienne Stevens, Kelly English, Ivan D. Florez, Juan J. Yepes-Nuñez, Brian Hutton, Matthew Muller, Lorenzo Moja, Sharon E. Straus, Andrea C. Tricco

**Affiliations:** Knowledge Translation Program, Li Ka Shing Knowledge Institute, St. Michael’s Hospital, 209 Victoria Street, Toronto, ON, Canada; Institute for Health Policy, Management, and Evaluation, University of Toronto, 155 College Street, Toronto, ON, Canada; School of Nursing and Midwifery, Trinity College, 24 D’Olier Street, Dublin 2, Ireland; Public Health Agency of Canada, Ottawa, ON, Canada; Patient Partner, Strategy for Patient Oriented-Research Evidence Alliance (SPOR EA), 209 Victoria Street, Toronto, ON, Canada; Department of Pediatrics, University of Antioquia, Medellin, Colombia; School of Rehabilitation Science, McMaster University, Hamilton, ON, Canada; Pediatric Intensive Care Unit, Clinica Las Américas-AUNA, Medellin, Colombia; School of Medicine, Universidad de los Andes, Bogotá, Colombia; Pulmonology Service, Internal Medicine Section, Fundación Santa Fe de Bogotá University Hospital, Bogotá, Colombia; Ottawa Hospital Research Institute, University of Ottawa, 1053 Carling Ave, Ottawa, Ontario; Department of Medicine, St. Michael’s Hospital, 2-61 Queen St E, Toronto, Ontario; Institute of Medical Sciences, University of Toronto, 1 King’s College Circle, Toronto, Ontario; Department of Biomedical Sciences for Health, University of Milan, Milan, Italy, Via Mangiagalli 31; Department of Geriatric Medicine, University of Toronto, Toronto, ON, Canada; Epidemiology Division & Institute of Health Policy, Management, and Evaluation, Dalla Lana School of Public Health, University of Toronto, Toronto, ON, Canada

**Author notes:** **Corresponding Author:** Dr. Areti Angeliki Veroniki, PhD, MSc Scientist, Knowledge Translation Program Assistant Professor, Institute for Health Policy, Management, and Evaluation, University of Toronto Li Ka Shing Knowledge Institute of St. Michael’s Hospital, Unity Health Toronto 209 Victoria Street, East Building, Toronto, Ontario, M5B 1T8, Canada.

**Keywords:** seasonal influenza, recombinant influenza vaccine, inactivated influenza vaccine, older adults, vaccine, network meta-analysis, patient and public involvement

## Abstract

**Objectives:** To compare the efficacy of influenza vaccines of any valency for adults 60 years and older.

**Methods:** *Design:* Systematic review with network meta-analysis (NMA)

*Information sources:* MEDLINE, EMBASE, JBI Evidence-Based Practice (EBP) Database, PsycINFO, and Cochrane Evidence Based Medicine database from inception to June 20, 2022.

*Eligibility criteria:* Randomized controlled trials (RCTs) including older adults (≥60 years old) receiving an influenza vaccine licensed in Canada or the United States (versus placebo, no vaccine, or any other licensed vaccine), at any dose.

*Outcome measures:* Primary outcomes: Laboratory-confirmed influenza (LCI) and influenza-like illness (ILI). Secondary outcomes: number of vascular adverse events, hospitalization for acute respiratory infection (ARI) and ILI, inpatient hospitalization, emergency room (ER) visit for ILI, outpatient visit, and mortality, among others.

*Data extraction, risk of bias (ROB), and certainty of evidence assessment:* Two reviewers screened, abstracted, and appraised articles (Cochrane ROB 2 tool) independently. We assessed certainty of findings using CINeMA and GRADE approaches.

*Data synthesis:* We performed random-effects meta-analysis and NMA, and estimated odds ratios (ORs) for dichotomous outcomes and incidence rate ratios (IRRs) for count outcomes along with corresponding 95% confidence intervals (95%CI) and prediction intervals.

**Results:** We included 41 RCTs and 15 companion reports comprising eight vaccine types and 206,032 participants. Vaccines prevented LCI compared with placebo, with high-dose trivalent (IIV3-HD) (NMA, nine RCTs, 52,202 participants, OR 0.23, 95%CI [0.11 to 0.51], low certainty of evidence) and RIV (OR 0.25, 95%CI [0.08 to 0.73], low certainty of evidence) among the most efficacious vaccines. Standard dose trivalent inactivated influenza vaccine (IIV3-SD) prevented ILI compared with placebo, but the result was imprecise (meta-analysis, two RCTs, 854 participants, OR 0.39, 95%CI [0.15 to 1.02], low certainty of evidence). Any high dose (HD) prevented ILI compared with placebo (NMA, nine RCTs, 65,658 participants, OR 0.38, 95%CI [0.15 to 0.93]). Adjuvanted quadrivalent inactivated influenza vaccine (IIV4-Adj) was associated with the least vascular adverse events (NMA: eight RCTs, 57,677 participants, IRR 0.18, 95%CI [0.07 to 0.43], very low certainty of evidence). RIV on all-cause mortality was comparable to placebo (NMA: 20 RCTs, 140,577 participants, OR 1.01, 95%CI [0.23 to 4.49], low certainty of evidence).

**Conclusions:** This systematic review demonstrated high efficacy associated with IIV3-HD and RIV vaccines in protecting elderly persons against LCI, and RIV vaccine minimizing all-cause mortality when compared with other vaccines. However, differences in efficacy between these vaccines remain uncertain with very low to moderate certainty of evidence.

**Funding:** Canadian Institutes of Health Research Drug Safety and Effectiveness Network (No. DMC – 166263)

**Systematic review registration:** PROSPERO CRD42020177357

**SUMMARY BOX:** *What is already known on this topic:*

- Seasonal influenza vaccination of older adults (≥60 years old) is an important societal, cost-effective means of reducing morbidity and mortality.
- A multitude of licensed seasonal influenza vaccines for older adults are available in a variety of formulations (such as IIV3, IIV4; prepared in standard and high doses; with and without an adjuvant) relying on production methods including those based on embryonated chicken eggs, or mammalian cell cultures and comprising seasonally selected viral strains or recombinant constructs.
- Lack of high-quality analysis of randomized control trial (RCT) data pertaining to influenza vaccine production and composition poses challenges for public health clinicians and policy makers who are tasked with making evidence-based decisions regarding recommendations about choosing optimally efficacious and safe influenza vaccines for older adults.

*What this study adds:*

- This systematic review and network meta-analysis of RCT data found that recombinant influenza vaccines (RIV) are among the most effective (lowest odds of laboratory-confirmed influenza [LCI]) and safest (lowest odds of all-cause mortality) of any licensed influenza vaccine type administered to older adults.

*How this study might affect research, practice or policy:*

- Our review points to a potential safety concern regarding increased odds of all-cause mortality associated with older adults receiving adjuvanted influenza vaccines (IIV3-adj and IIV4-adj).

## INTRODUCTION

Seasonal influenza is a leading cause of infectious respiratory disease globally causing three to five million cases of severe illness and 650,000 deaths annually.^1^ In Canada, influenza ranks among the top 10 causes of mortality, with approximately 3,500 deaths and 12,200 influenza hospitalizations per year.^2^ Annually, influenza results in an economic burden on average of $14,000 to $20,000 per laboratory-confirmed influenza (LCI) hospitalization.^3^

Older adults, particularly those who are frail bare a disproportionate burden of influenza and its complications. Globally, individuals 75 years of age and older experience the highest rates of influenza-associated respiratory deaths (51.3 to 99.4 deaths per 100,000 vs 13.3 to 27.8 in those 60 to 74 years of age, and 1.0 to 5.1 in those less than 60 years of age).^4^ Annual influenza vaccination is the best way to prevent influenza infection and its complications.^5, 6^

Although there are various influenza vaccines available for older adults, all current vaccines contain two influenza A virus strains and either one (trivalent) or two (quadrivalent) influenza B virus strains.^7^ Existing randomized controlled trials (RCTs) focus on the safety and efficacy of individual vaccines. The lack of direct comparative vaccine efficacy evidence poses challenges to public health clinicians and policy makers to make evidence-based decisions regarding the preferential use of one influenza vaccine over another in older adults regarding vaccine choice. We aimed to systematically assess the comparative efficacy of influenza vaccines (e.g., trivalent and quadrivalent at standard-dose, high-dose or adjuvanted) in adults 60 years of age and older. A systematic review with network meta-analysis (NMA) was conducted to fill this critical knowledge gap in comparative vaccine efficacy.

## METHODS

The protocol was registered in PROSPERO (CRD42020177357), and any deviations are reported in Appendix 1 (Additional file 1). In Canada, national immunization recommendations of vaccine use are made by the National Advisory Committee on Immunization (NACI). Our protocol was revised in collaboration with the NACI Influenza Working Group, to improve the feasibility and relevancy of this review for key knowledge users. Below, we briefly describe the methods used in our review; more details are presented in Appendix 2 (Additional file 1). This systematic review follows: PRISMA (Preferred Reporting Items for Systematic Reviews and Meta-analyses) extension to NMA^8^ and Guidance for Reporting Involvement of Patients and the Public (GRIPP-2) reporting guidelines^9^ (Additional files 2 and 3), as well as the methods outline in the Cochrane Handbook for Systematic Reviews of Interventions.^10^

### Knowledge user engagement

We enhanced systematic review process by employing an integrated knowledge translation approach^11^ from project onset via established partnerships with the NACI Influenza Working Group, who will use these research results to inform the development of guidance for influenza vaccination. Our systematic review included: two clinicians (MM, SES), one patient partner (KE), the NACI Secretariat (AS, PDP, AG, WS, NM, AS), and seven researchers (AAV, MK, IDF, JJYN, BH, LM, ACT). Throughout the research process, all knowledge users actively participated by contributing their insights in various stages, such as developing and refining the research question, reviewing the search parameters, framing the risk groups, prioritizing the outcome measures, and interpreting the study findings.

### Data sources and searches

We searched Ovid MEDLINE, JBI (formerly the Joanna Briggs Institute) Evidence-Based Practice (EBP) Database, Embase, PsycINFO, and Cochrane Evidence-Based Medicine database from inception to March 31, 2022, with an updated search on June 20, 2022. An expert librarian developed the search strategies with input from the research team. Another expert librarian peer-reviewed the search strategy using the Peer Review of Electronic Search Strategies (PRESS) checklist (see Appendix 3 (Additional file 1) for literature search).^12^ Grey literature (i.e., unpublished or difficult-to-locate studies) was searched using the Canadian Agency for Drugs and Technologies in Health (CADTH) guide (see Appendix 4 (Additional file 1)).^13^ We consulted content experts from the NACI Influenza Working Group, and scanned the NACI Practices review,^14^ the National Immunization Technical Advisory Group (NITAG) websites (e.g., SAGE, JCVI, ATAGI, STIKO), reference lists of included studies and related reviews searching for additional relevant studies (Appendix 4 (Additional file 1)). A library technician searched for article corrections and retractions to inform our retrieval of full-text reports.

### Eligibility criteria

We included RCTs comparing any influenza vaccine at any dose for adults 60 years of age and older that are licensed in Canada or the United States, at time of the review and irrespective of commercial name. RCTs comparing an influenza vaccine versus another influenza vaccine, placebo or any other licensed vaccine, were eligible. We included RCTs regardless of publication status, publication date, duration of follow-up, language of publication, geographic region or setting (e.g., community, residential facilities, nursing homes). Studies involving the general population that reported stratified data and results for older adults were also included. If stratified data and results were not reported, authors were contacted to retrieve any additional information with up to three reminders.

Eligible influenza vaccines included but were not limited to egg-based, quadrivalent inactivated influenza vaccine (IIV4) at standard (SD) or high dose (HD) or adjuvanted (Adj), trivalent inactivated influenza vaccine (IIV3) at SD or HD or Adj, mammalian cell culture-based trivalent or quadrivalent inactivated influenza vaccine, and quadrivalent or trivalent recombinant influenza vaccine (RIV4 or RIV3). We determined vaccine license at time of the study based on the details provided by the studies and/or trial protocols. Intramuscular and subcutaneous routes of administration of relevant licensed vaccines were considered for inclusion.

Eligible outcomes were determined in collaboration with the NACI Influenza Working Group and clinician experts based on the GRADE (Grading of Recommendations, Assessment, Development, and Evaluations)^15^ methodology for rating the importance of outcomes for decision-making.^16^ The primary outcomes were LCI and influenza-like illness (ILI). Secondary outcomes were number of vascular adverse events, hospitalization for acute respiratory infection (ARI) and ILI, inpatient hospitalization, emergency room (ER) visit for ILI, outpatient visit, and mortality. All outcomes included binary data apart from vascular adverse events that included count data. Other outcomes of interest included ARI cases, LCI hospitalization, LC-ARI cases, ER visit for ARI or LCI or pneumonia or any cause, hospitalization for pneumonia due to ILI, ARI, LCI, inpatient or outpatient hospital visit, hospitalization due to cardiovascular events, LCI-related mortality, LCI-related healthcare interactions, number of hospitalizations due to cardiovascular events, number of participants with vascular adverse events, and influenza-related vascular adverse events.

We excluded intradermal, intranasal, and AS03-adjuvanted vaccines, which are not licensed for use in Canada or the United States. Pandemic vaccines were excluded along with experimental vaccines that have not been approved for use.

### Selection process, data extraction and risk of bias

The study selection, data extraction and risk of bias processes are described in Appendix 2.

### Synthesis

Study-level information on participant and study characteristics was synthesized descriptively.

#### Pairwise meta-analyses

We synthesized data in a meta-analysis using the odds ratio (OR) for dichotomous outcomes and the incidence rate ratio (IRR) for count outcomes, along with corresponding 95% confidence intervals (95% CI) when at least two studies were available. We performed a random-effects inverse-variance meta-analysis, since we expected the studies to be methodologically and clinically different (e.g., vaccine dose, participant age). We estimated the overall effect size and its 95% CI using the Hartung-Knapp-Sidik-Jonkman (HKSJ) method for meta-analyses with few studies:^17–19^ specifically, when the estimated heterogeneity was positive (>0) among at least three studies in the meta-analysis, we used the HKSJ approach,^20, 21^ otherwise we applied the Wald-type approach.^17–19, 22–24^ We supplemented estimated overall effects and 95% CIs with prediction intervals (PIs) to capture the interval within which we expected the true intervention effect of a new study to fall. We used the restricted maximum likelihood method^25^ to estimate the between-study variance (τ^2^) and the Q-profile approach to calculate its 95%CI.^26, 27^ We also calculated the percentage of variation that can be attributed to heterogeneity rather than chance using the I^2^ statistic.^28^

#### Network meta-analyses

For each outcome and connected network diagram, we conducted a frequentist random-effects NMA when the number of studies exceeded the number of intervention nodes. Alternatively, pairwise meta-analysis was conducted for each intervention comparison separately. A common within-network τ^2^ was assumed for each network, which was a clinically reasonable assumption since networks were composed of influenza-type vaccines; τ^2^ was estimated using the restricted maximum likelihood method.^25^ We estimated summary ORs or IRRs and presented them along with 95% CIs and prediction intervals. Interventions were ranked according to their efficacy using the P-score statistic.^29^

We conducted an NMA under the transitivity assumption, where trial participants are equally likely to be randomized to any of the network interventions and the included trials are sufficiently similar across intervention comparisons with respect to the distribution of effect modifiers. We visually assessed transitivity across intervention comparisons on the *a priori* selected effect modifiers: overall risk of bias, participant age, percentage of female participants, and year of publication, using mean for continuous and mode for discrete variables. The statistical manifestation of intransitivity can cause inconsistency in the network of interventions. We assessed for consistency using both local (node-splitting)^30^ and global (design-by-treatment interaction)^31^ approaches. We planned to assess transitivity for populations with comorbid conditions, frailty, and previous influenza vaccination, but these were rarely reported in the included studies.

#### Presentation of results

Summary ORs/IRRs for all pairwise comparisons, along with their 95% CIs and 95% PIs, were presented in tables. We also calculated and presented vaccine efficacy (VE) for all pairwise comparisons in the LCI and ILI outcomes using the combined mean baseline risk per outcome. P-scores across interventions and outcomes were presented in a rank-heat plot (https://rankheatplot.com/rankheatplot/).^25^ Statistical analyses were conducted using the *meta*^32^ and *netmeta*^33^ packages in R.

### Assessment of the confidence in the evidence

We assessed confidence in NMA estimates for each outcome using CINeMA (Confidence in Network meta-analysis)^34^ and confidence in pairwise meta-analysis estimates for which a NMA could not be performed using the GRADE approach.^35^ More details are presented in Appendix 2.

## RESULTS

Overall, 41 studies (206,032 participants) and 15 companion reports were included,^36–89^ after screening 6,858 citations and 3,425 full-text articles (Figure 1). All of the included studies were written in English. Two of the seven contacted authors responded to our emails asking for additional information, and one provided data for analysis. Of the 41 RCTs, 26 were included in the quantitative analysis; the remaining 15 RCTs did not report relevant data for the analysis or compared the same intervention node (e.g., Agriflu^®^ vs. Fluvirin^®^ ^90^ both of which are trivalent, inactivated, egg-based influenza vaccines). Lists of the studies included in the review and studies excluded after data abstraction can be found in Appendices 8 and 9 (Additional file 1), respectively.

**Figure 1.**
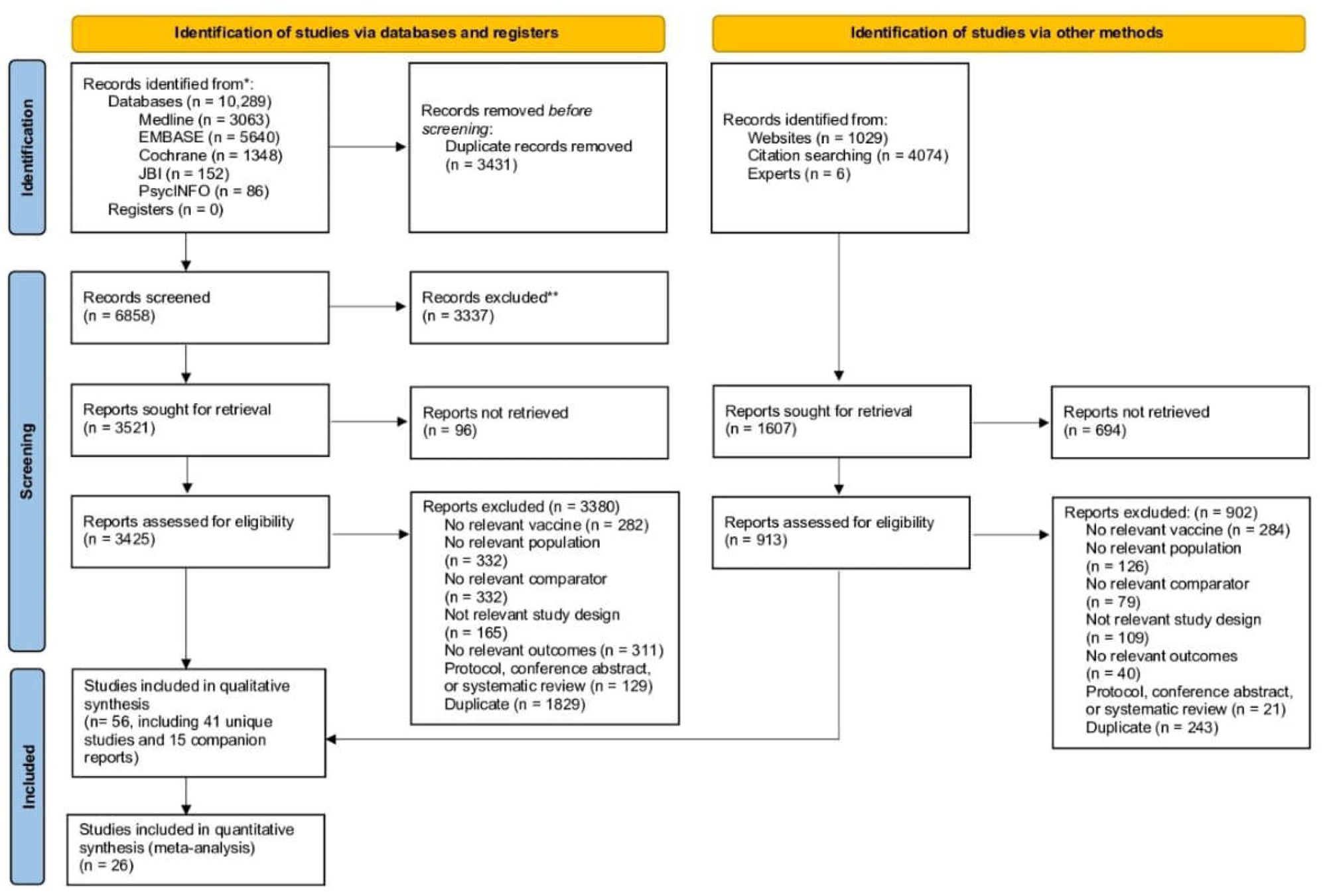
PRISMA flow diagram for identification of eligible studies

### Study, patient, and intervention characteristics

The included RCTs were published between 1994-2021 with most (36%) published between 2016-2020 (Table 1). Most of the studies were conducted in the United States (N=19, 46%) and funded by industry (N= 23, 56%). Three were cluster-RCTs.^51, 52, 56^ Most RCTs (N=9, 22%) compared IIV3-SD versus IIV3-HD, followed by RCTs comparing IIV3-SD versus placebo or versus IIV3-Adj (N=4, 10%) (Table 1, Appendix 10 (Additional file 1).

**Table 1:**
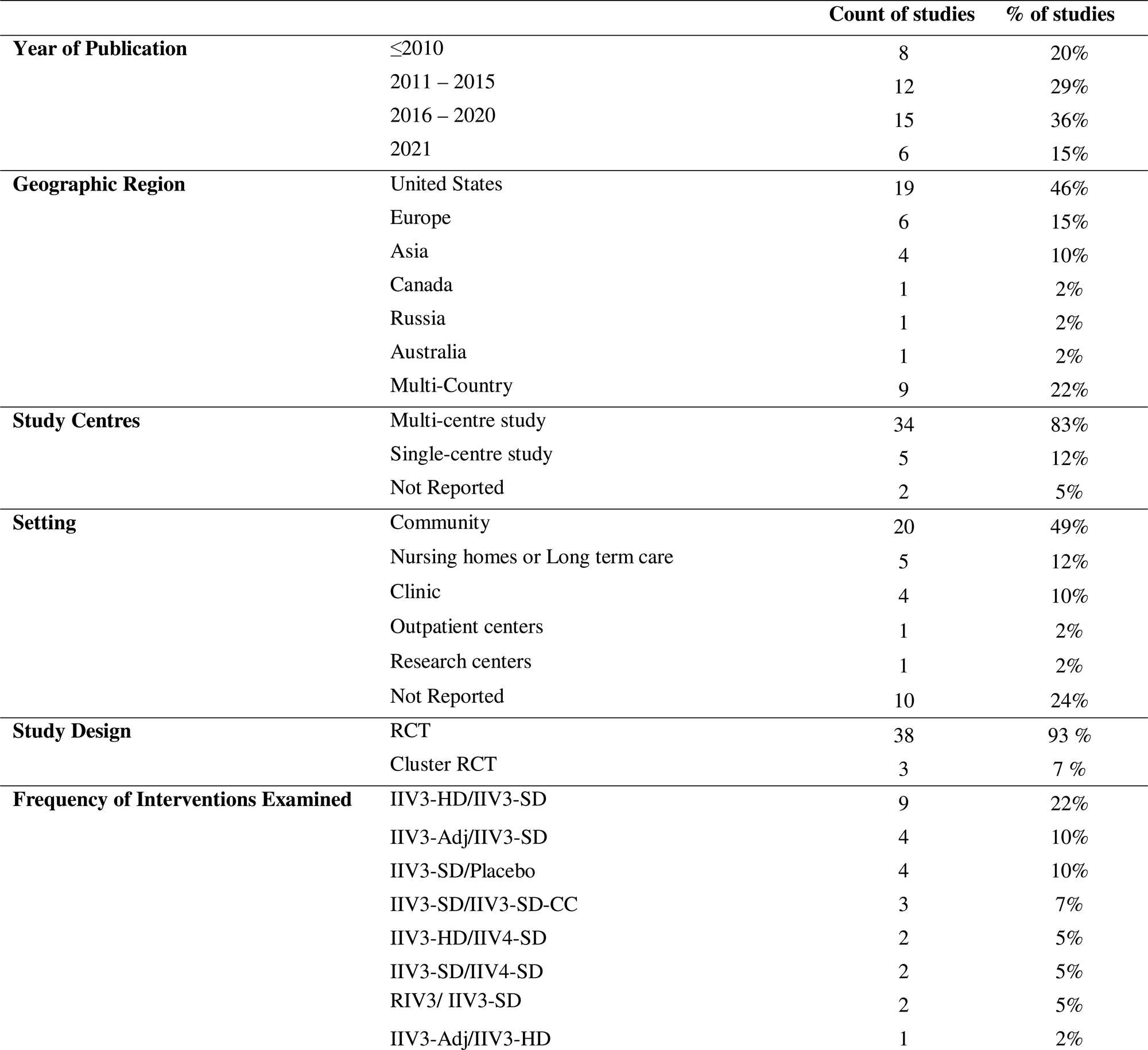
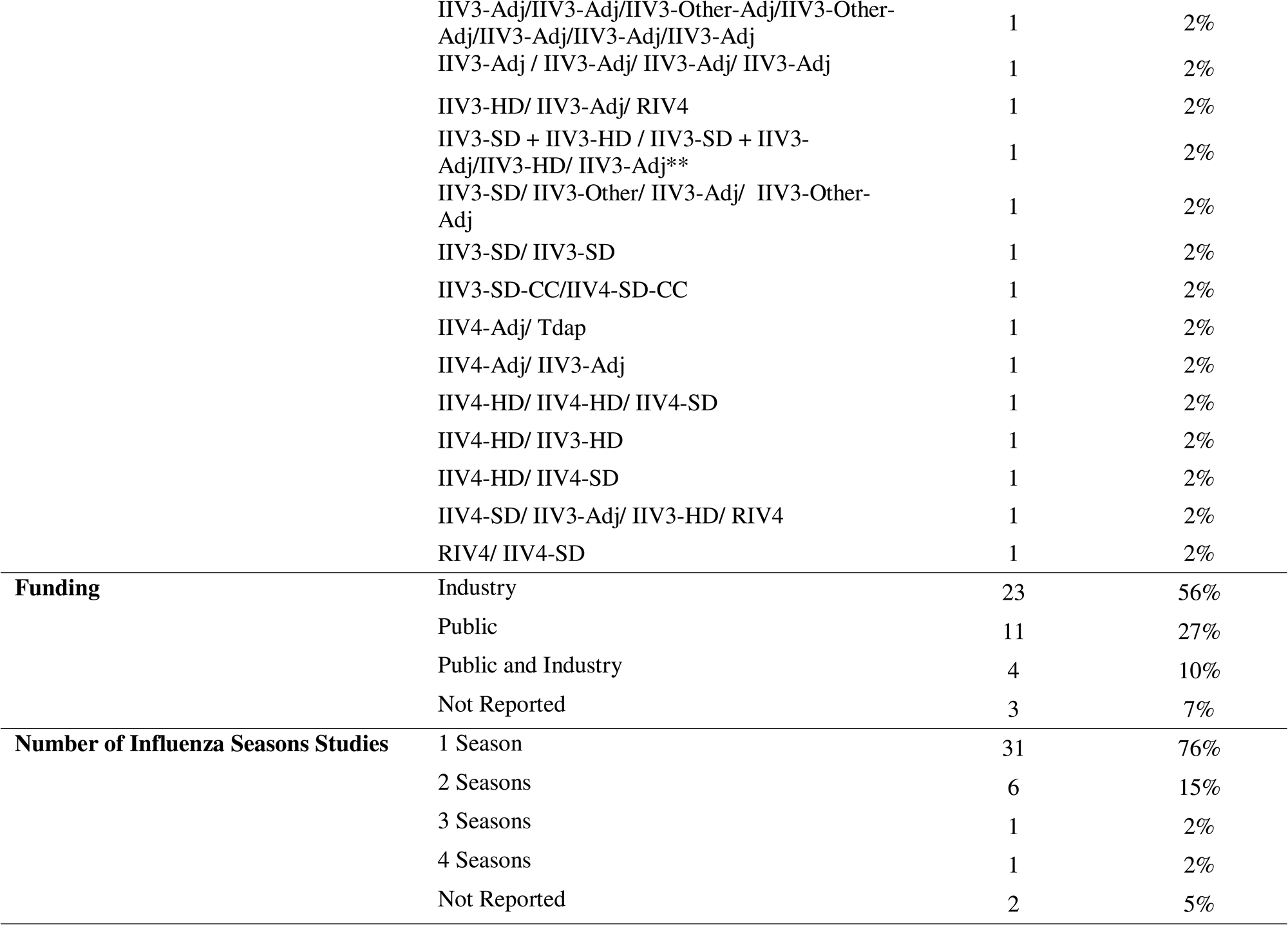
Study characteristic of included studies (N=41)

The reported participant age within most RCTs (N=34, 83%) was ≥65 years; participants reflected racial diversity (N=19, 46%; Table 2, Appendix 11 (Additional file 1).

**Table 2:**
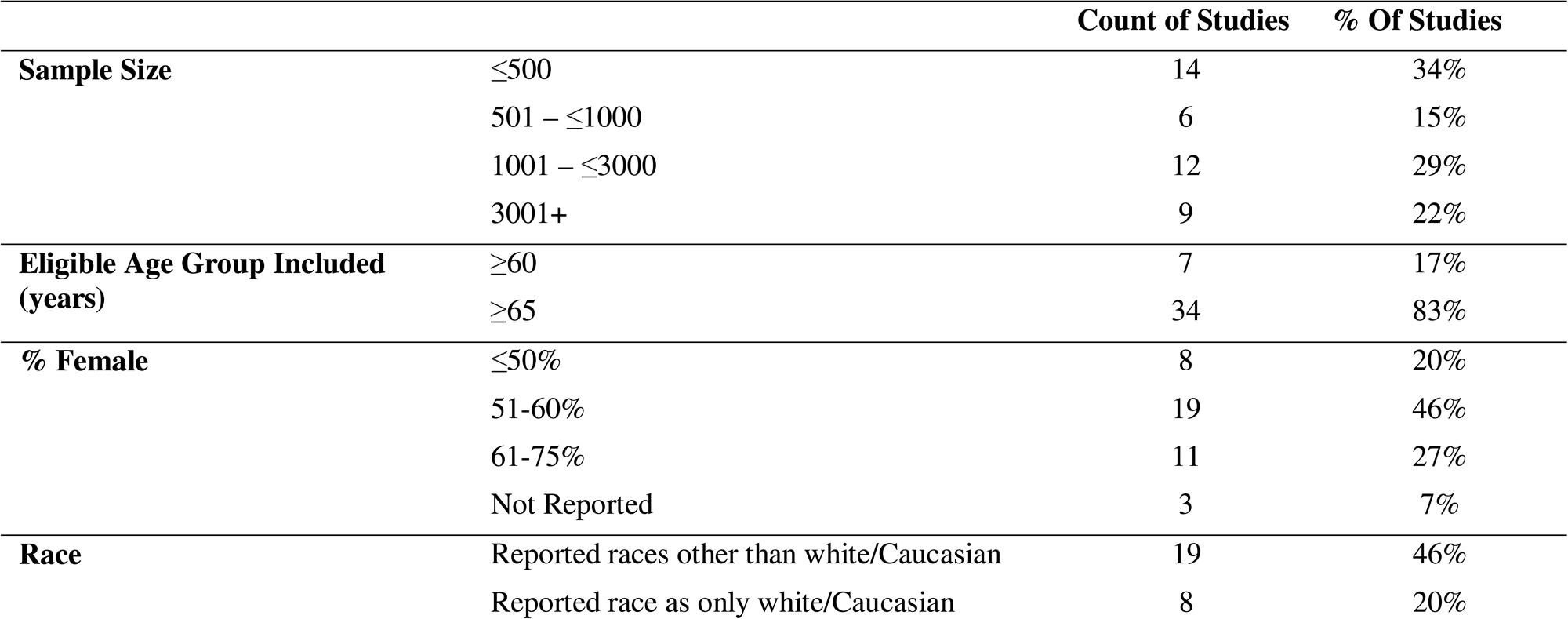
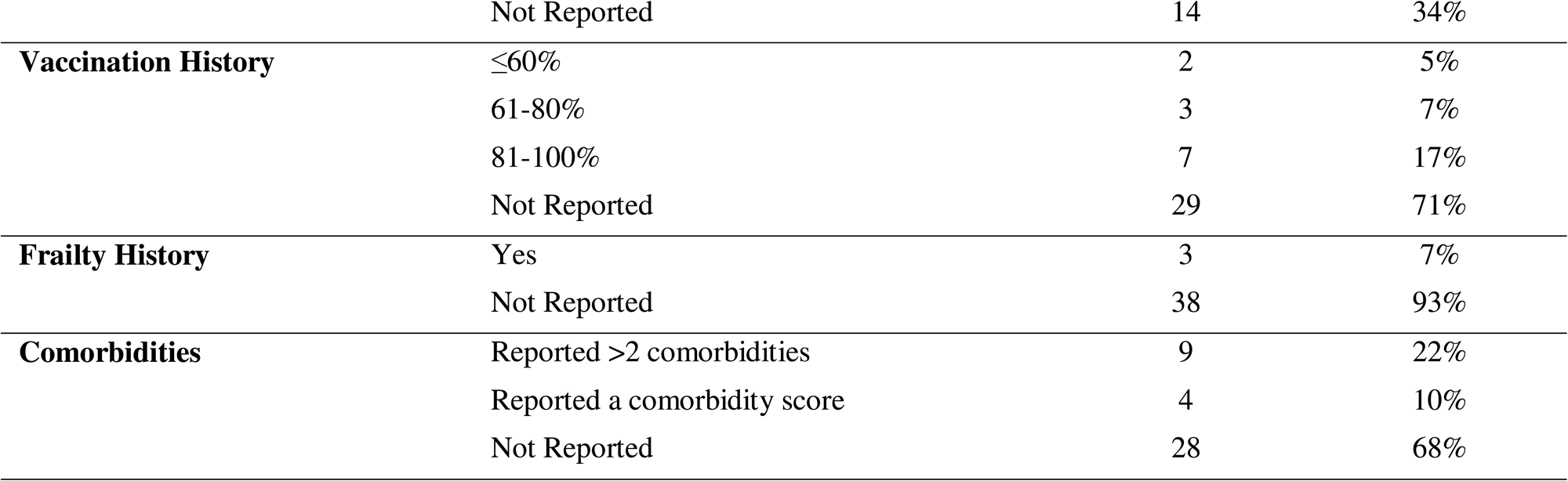
Participant characteristics of included studies (N=41)

|  |  | Count of Studies | % Of Studies |
| --- | --- | --- | --- |
| **Sample Size** | $\leq 500$ | 14 | 34% |
| | 501 – $\leq 1000$ | 6 | 15% |
| | 1001 – $\leq 3000$ | 12 | 29% |
|  | 3001+ | 9 | 22% |
| **Eligible Age Group Included (years)** | $\geq 60$ | 7 | 17% |
| | $\geq 65$ | 34 | 83% |
| **% Female** | $\leq 50\%$ | 8 | 20% |
|  | 51-60% | 19 | 46% |
|  | 61-75% | 11 | 27% |
|  | Not Reported | 3 | 7% |
| **Race** | Reported races other than white/Caucasian | 19 | 46% |
|  | Reported race as only white/Caucasian | 8 | 20% |
|  | Not Reported | 14 | 34% |
| **Vaccination History** | ≤60% | 2 | 5% |
|  | 61-80% | 3 | 7% |
|  | 81-100% | 7 | 17% |
|  | Not Reported | 29 | 71% |
| **Frailty History** | Yes | 3 | 7% |
|  | Not Reported | 38 | 93% |
| **Comorbidities** | Reported >2 comorbidities | 9 | 22% |
|  | Reported a comorbidity score | 4 | 10% |
|  | Not Reported | 28 | 68% |

Overall, eight influenza vaccine types were identified across all RCT interventions: IIV3-Adj, IIV4-Adj, IIV3-HD, IIV4-HD, IIV3-SD, IIV4-SD, RIV3, and RIV4. Most influenza vaccines were egg-based (N=47, 46%), and at standard dose (N=64, 63%). The delay between doses, which, if reported and/or applicable, ranged from three to four weeks (Table 3, Appendix 12 (Additional file 1)).

**Table 3:**
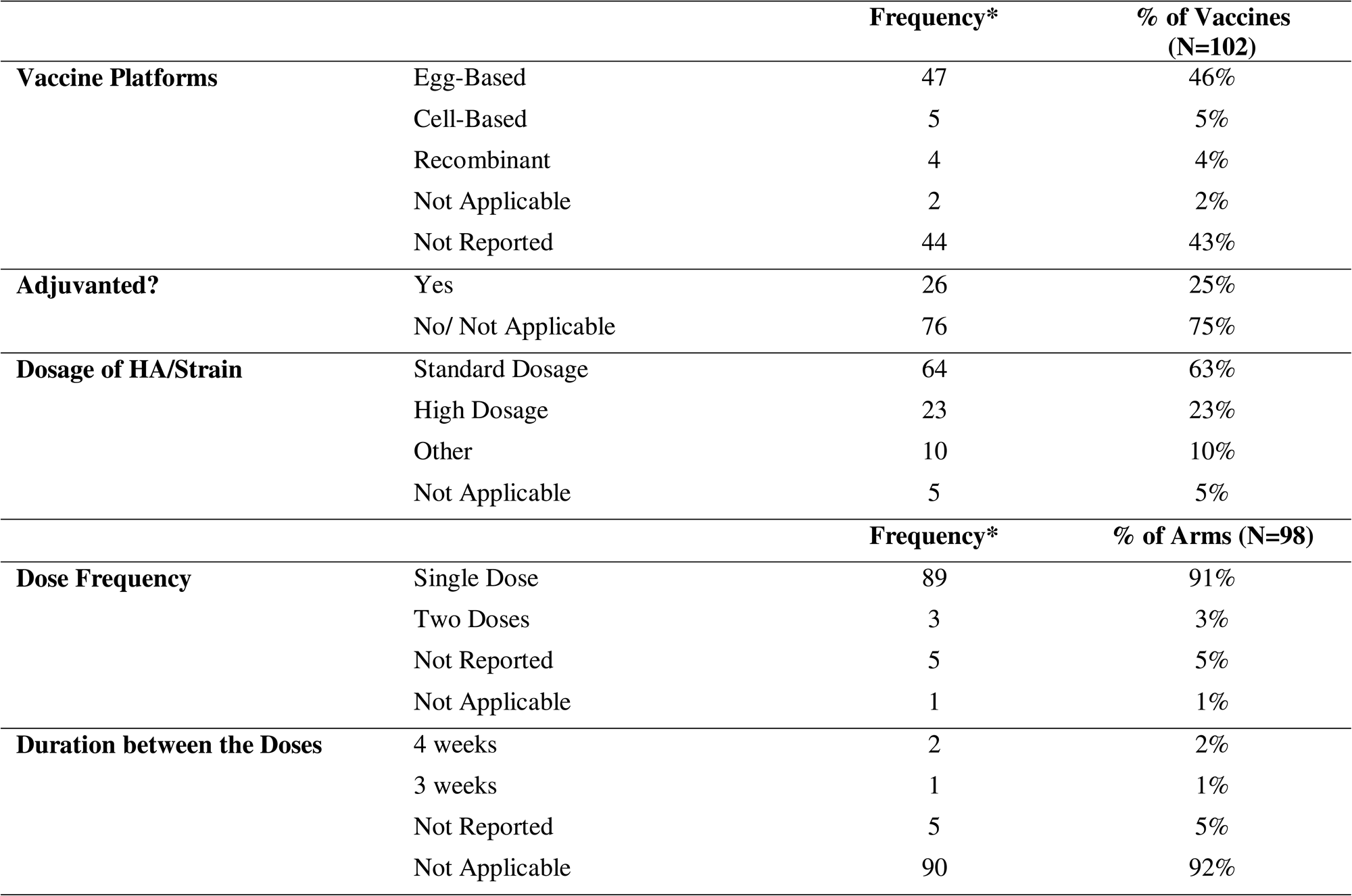
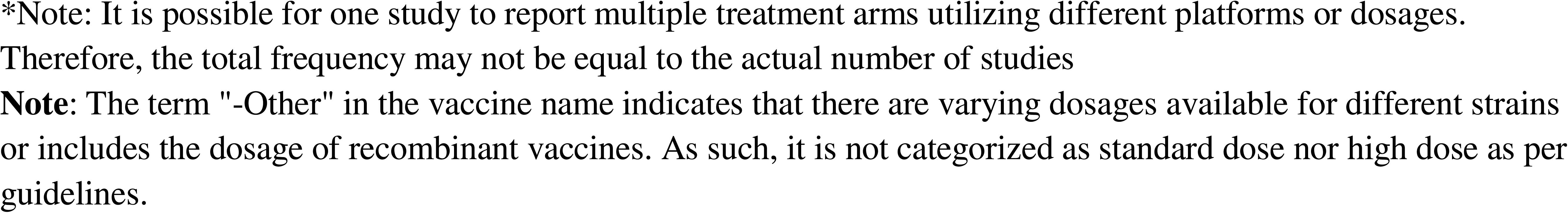
Vaccine characteristics of included studies (N=41)

|  |  | Frequency* | % of Vaccines<br>(N=102) |
| --- | --- | --- | --- |
| **Vaccine Platforms** | Egg-Based | 47 | 46% |
|  | Cell-Based | 5 | 5% |
|  | Recombinant | 4 | 4% |
|  | Not Applicable | 2 | 2% |
|  | Not Reported | 44 | 43% |
| **Adjuvanted?** | Yes | 26 | 25% |
|  | No/ Not Applicable | 76 | 75% |
| **Dosage of HA/Strain** | Standard Dosage | 64 | 63% |
|  | High Dosage | 23 | 23% |
|  | Other | 10 | 10% |
|  | Not Applicable | 5 | 5% |
|  |  | Frequency* | % of Arms (N=98) |
| **Dose Frequency** | Single Dose | 89 | 91% |
|  | Two Doses | 3 | 3% |
|  | Not Reported | 5 | 5% |
|  | Not Applicable | 1 | 1% |
| **Duration between the Doses** | 4 weeks | 2 | 2% |
|  | 3 weeks | 1 | 1% |
|  | Not Reported | 5 | 5% |
|  | Not Applicable | 90 | 92% |

\*Note: It is possible for one study to report multiple treatment arms utilizing different platforms or dosages. Therefore, the total frequency may not be equal to the actual number of studies **Note:** The term "-Other" in the vaccine name indicates that there are varying dosages available for different strains or includes the dosage of recombinant vaccines. As such, it is not categorized as standard dose nor high dose as per guidelines.

Of the a priori selected outcomes, 17 were omitted from both pairwise or network meta-analysis. This is because in 10 of these outcomes, there was a lack of relevant data within the identified studies. In two outcomes only one study reported relevant data, while for five outcomes, there were multiple studies available, but one study was included per intervention comparison. Of the outcomes analyzed in pairwise or network meta-analysis, an additional seven outcomes had studies which were not included in the analyses, because intervention comparisons included a single study or were disconnected from the underlying network. See Appendix 13 (Additional file 1) for more details.

### Within-study risk of bias and across-study reporting bias

Within-study bias appraisal suggested that overall low ROB was present for: two (22%) RCTs reporting LCI, nine (45%) RCTs reporting all-cause mortality, four (50%) RCTs reporting vascular adverse events, three (33%) RCTs reporting ILI, and one RCT (50%) reporting ER-visits (Figure 2, Appendix 14 (Additional file 1)). Overall, ROB for outpatient visits, hospitalization for ARI and ILI, and inpatient hospitalizations outcomes were judged to be at some or high concerns (Appendix 15 (Additional file 1)).

**Figure 2.**
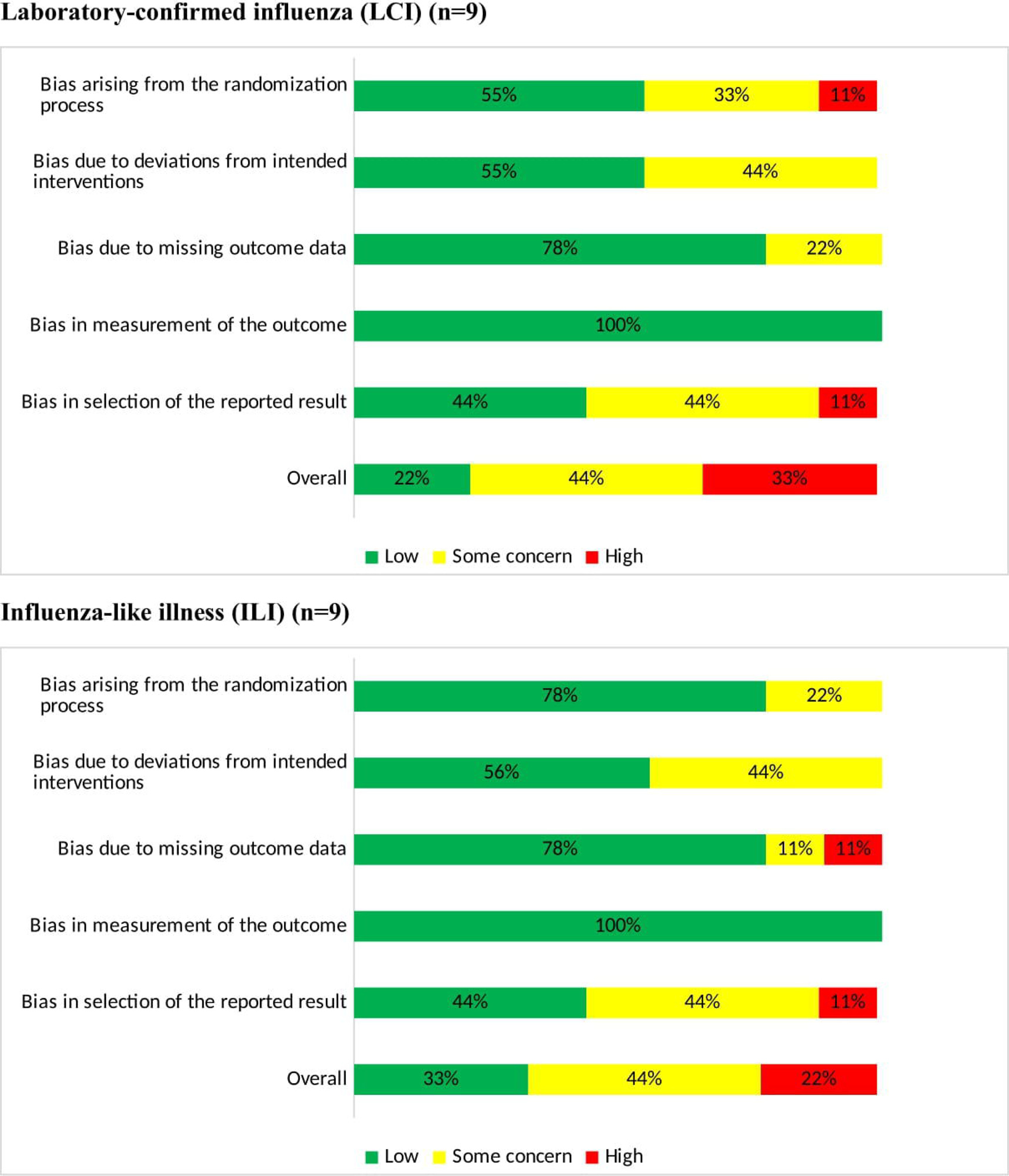
Cochrane Risk of Bias 2 (ROB 2) Summary Results of Primary Outcomes (N=26 RCTs) Laboratory-confirmed influenza (LCI) (n=9) Influenza-like illness (ILI) (n=9)

Reporting bias across studies could be assessed only for the all-cause mortality outcome, due to the small number of studies per outcome. The comparison-adjusted funnel indicated no evidence of publication bias or small-study effects (Appendix 16 (Additional file 1)).

### Results of testing assumptions

Assessment of the consistency assumption was possible for LCI and all-cause mortality outcomes (with available closed loops of evidence), where there was no evidence of statistical inconsistency (design-by-treatment interaction model LCI: χ^2^= 2.35, 2 degrees of freedom (df), p = 0.31; all-cause mortality: χ^2^= 0.23, 3 df, p = 0.98; Appendix 17A (Additional file 1)). Overall, there were no important concerns regarding average age or ROB, but the mean percentage of female participants varied for LCI (range across comparisons: 32% to 59%), outpatient visits (range across comparisons: 26% to 55%) and all-cause mortality (range across comparisons: 26% to 63%) outcomes, which may challenge the reliability of the transitivity assumption (Appendix 17B (Additional file 1)).

### Results of syntheses

Below we present the analysis results for NMA and pairwise meta-analysis for the included vaccines against placebo or against IIV3-SD if a placebo node wasn’t available for the outcome. Additional results and analyses are presented in Appendices 17C and 18 (Additional file 1). The network geometry for each outcome analyzed in an NMA is presented in Figure 3.

**Figure 3.**
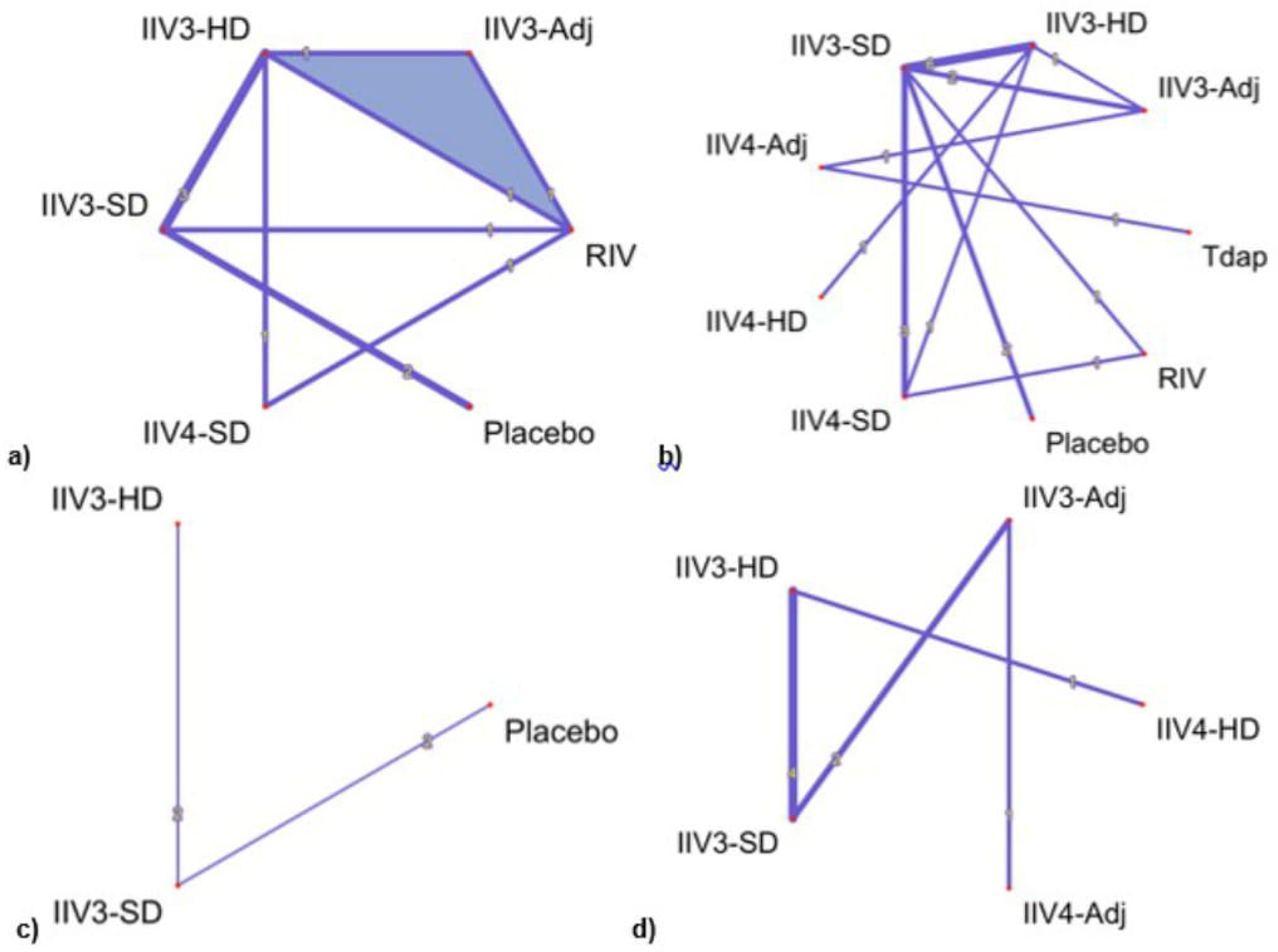
Network-plots of (a) LCI, (b) all-cause mortality, (c) outpatient visits, (d) number of vascular AEs Abbreviations-IIV3: Inactivated Influenza Vaccine Trivalent; IIV4: Inactivated Influenza Vaccine Quadrivalent; Adj: Adjuvanted; SD: Standard dosage; HD: High dosage; RIV: Recombinant influenza vaccine; Tdap: Tetanus, diphtheria, pertussis; LCI: laboratory-confirmed influenza; AEs: adverse events

#### Laboratory confirmed influenza (LCI)

The NMA for LCI included nine RCTs,^38, 45–47, 54, 55, 62, 69, 89^ six interventions (including placebo), and 52,202 participants (see Appendix 19 (Additional file 1) for study definitions). Results suggested that vaccines were associated with reduced incidence of LCI compared to placebo, but with varying degrees of precision of effect estimates, although with no appreciable between-study network heterogeneity (I^2^=0%, τ=0.00). IIV3-HD (OR 0.23, 95%CI [0.11 to 0.51], P-score= 0.78; low certainty of evidence, VE 72.85 95%CI [43.5 to 86.64]) and RIV (OR 0.25, 95%CI [0.08 to 0.73], P-score= 0.73; low certainty of evidence, VE 70.63 95%CI [22.86 to 90.21]) were the most effective vaccines. These were followed by IIV3-Adj vaccine (OR 0.24, 95%CI [0.04 to 1.47], P-score= 0.65; very low certainty of evidence, VE 71.73 95%CI [-34.45 to 95.06]) (Table 4, Appendices 20, 21 (Additional file 1)).

**Table 4.** Summary of results in outcomes with pairwise and network meta-analysis.

| Treatment Comparison (vs. Placebo) | Number of studies | OR: Direct Meta-Analysis estimates (95% CI) | OR: Network Meta-Analysis Estimate (95% CI), (95% PI) | RR: Direct Meta-Analysis estimates (95% CI) | RR: Network Meta-Analysis Estimate (95% CI) | VE: Direct Meta-Analysis estimates (95% CI) | VE: Network Meta-Analysis Estimate (95% CI) | P - Score | Credibility Assessment (CINeMA) |
| --- | --- | --- | --- | --- | --- | --- | --- | --- | --- |
| **Laboratory Confirmed Influenza: OR** |  |  |  |  |  |  |  |  |  |
| IIV3-Adj | 0 |  | 0.24 (0.04, 1.47), (0.02,3.1) |  | 0.28 (0.05,1.34) |  | 71.73 (-34.45,95.06) | 0.65 | Low |
| IIV3-HD | 0 |  | 0.23 (0.11,0.51) |  | 0.27 (0.13,0.56) |  | 72.85 (43.5,86.64) | 0.78 | Low |
| IIV3-SD | 2 | 0.31 (0.13, 0.75) | 0.31 (0.15,0.67), (0.08,0.7) | 0.36 (0.16,0.79) | 0.36 (0.18,0.72) | 64.08 (21.08,84.29) | 64.08 (28.3,81.95) | 0.45 | Low |
| IIV4-SD | 0 |  | 0.35 (0.12,1.07), (0.07,1.70) |  | 0.40 (0.15,1.06) |  | 59.81 (-5.53,85.46) | 0.37 | Low |
| RIV | 0 |  | 0.25 (0.08,0.73), (0.05,1.15) |  | 0.29 (0.10,0.77) |  | 70.63 (22.86,90.21) | 0.73 | Low |
| Placebo |  |  |  |  |  |  |  | 0.02 |  |
| Tau | 0 |  |  |  |  |  |  |  |  |
| I<sup>2</sup> | 0% |  |  |  |  |  |  |  |  |
| **Number of Vascular Adverse Events: IRR (relative to IIV3-SD)** |  |  |  |  |  |  |  |  |  |
| IIV3-Adj | 2 | 0.83 (0.54, 1.27) | 0.90 (0.54, 1.50), (0.23, 3.50) |  |  |  |  | 0.23 | Very Low |
| IIV3-HD | 4 | 0.75 (0.43, 1.29) | 0.69 (0.49, 0.97), (0.20, 2.32) |  |  |  |  | 0.48 | Very Low |
| IIV4-Adj | 0 |  | 0.18 (0.07, 0.43), (0.03, 1.07) |  |  |  |  | 0.98 | Very Low |
| IIV4-HD | 0 |  | 0.46 (0.21, 1.00), (0.09, 2.41) |  |  |  |  | 0.71 | Low |
| IIV3-SD |  |  |  |  |  |  |  | 0.1 |  |
| Tau | 0.34 |  |  |  |  |  |  |  |  |
| I<sup>2</sup> | 97% |  |  |  |  |  |  |  |  |
| Outpatient Visits: OR |  |  |  |  |  |  |  |  |  |
| IIV3-HD | 0 |  | 0.65 (0.35, 1.19), (0.01, 41.95) |  |  |  |  | 0.49 | Very Low |
| IIV3-SD | 2 | 0.4 (0.07, 2.14) | 0.64 (0.36, 1.14), (0.01, 35.50) |  |  |  |  | 0.93 | Very Low |
| Placebo |  |  |  |  |  |  |  | 0.07 |  |
| Tau | 0.11 |  |  |  |  |  |  |  |  |
| I<sup>2</sup> | 60.70% |  |  |  |  |  |  |  |  |
| All-cause Mortality: OR |  |  |  |  |  |  |  |  |  |
| IIV3-Adj | 0 |  | 1.60 (0.44, 5.85), (0.37, 6.87) |  |  |  |  | 0.42 | Low |
| IIV3-HD | 0 |  | 1.40 (0.41, 4.84), (0.35, 5.64) |  |  |  |  | 0.61 | Low |
| IIV3-SD | 2 | 1.47 (0.43, 5.06) | 1.47 (0.43, 5.06), (0.37, 5.89) |  |  |  |  | 0.45 | Very Low |
| IIV4-Adj | 0 |  | 8.00 (0.29, 217.75), (0.20, 326.89) |  |  |  |  | 0.42 | Low |
| IIV4-HD | 0 |  | 1.05 (0.12, 9.29), (0.09, 12.14) |  |  |  |  | 0.66 | Low |
| IIV4-SD | 0 |  | 1.42 (0.41, 4.97), (0.35, 5.80) |  |  |  |  | 0.55 | Very Low |
| RIV | 0 |  | 1.02 (0.23, 4.53), (0.19, 5.44) |  |  |  |  | 0.75 | Low |
| Tdap | 0 |  | 8.24 (0.29, 232.17), (0.19, 350.04) |  |  |  |  | 0.17 | Low |
| Placebo |  |  |  |  |  |  |  | 0.71 |  |
| Tau<sup>2</sup> | 0 |  |  |  |  |  |  |  |  |
| I<sup>2</sup> | 0% |  |  |  |  |  |  |  |  |

Results were imprecise in sensitivity analyses restricting to studies with percentage of female above 50% and matched vs mismatched circulating strains (Appendix 17C (Additional file 1)). When formulations were combined (i.e., Adj, SD, HD, RIV), RIV performed best (OR 0.22, 95%CI [0.10 to 0.49], P-score= 0.78, 95% PI [0.08, 0.63], I^2^=0%, τ=0.00). Precise results were identified for SD (OR 0.31, 95%CI [0.15 to 0.67], P-score= 0.33, 95% PI [0.12 to 0.85]) and HD (OR 0.23, 95%CI [0.11 to 0.50], P-score= 0.71, 95% PI [0.08 to 0.64]).

#### Influenza-like illness (ILI)

NMA with original coding of interventions was not possible for ILI. Pairwise meta-analysis suggested that IIV3-SD was associated with reduced ILI incidence compared with placebo, but was imprecise (two RCTs,^36, 89^ 854 participants; OR 0.39, 95%CI [0.15 to 1.02], I^2^=8%, τ=0.21 low certainty of evidence, VE 57.79 95%CI [-1.75 to 83.22]) (Table 5, Appendix 22 (Additional file 1)).

**Table 5.**
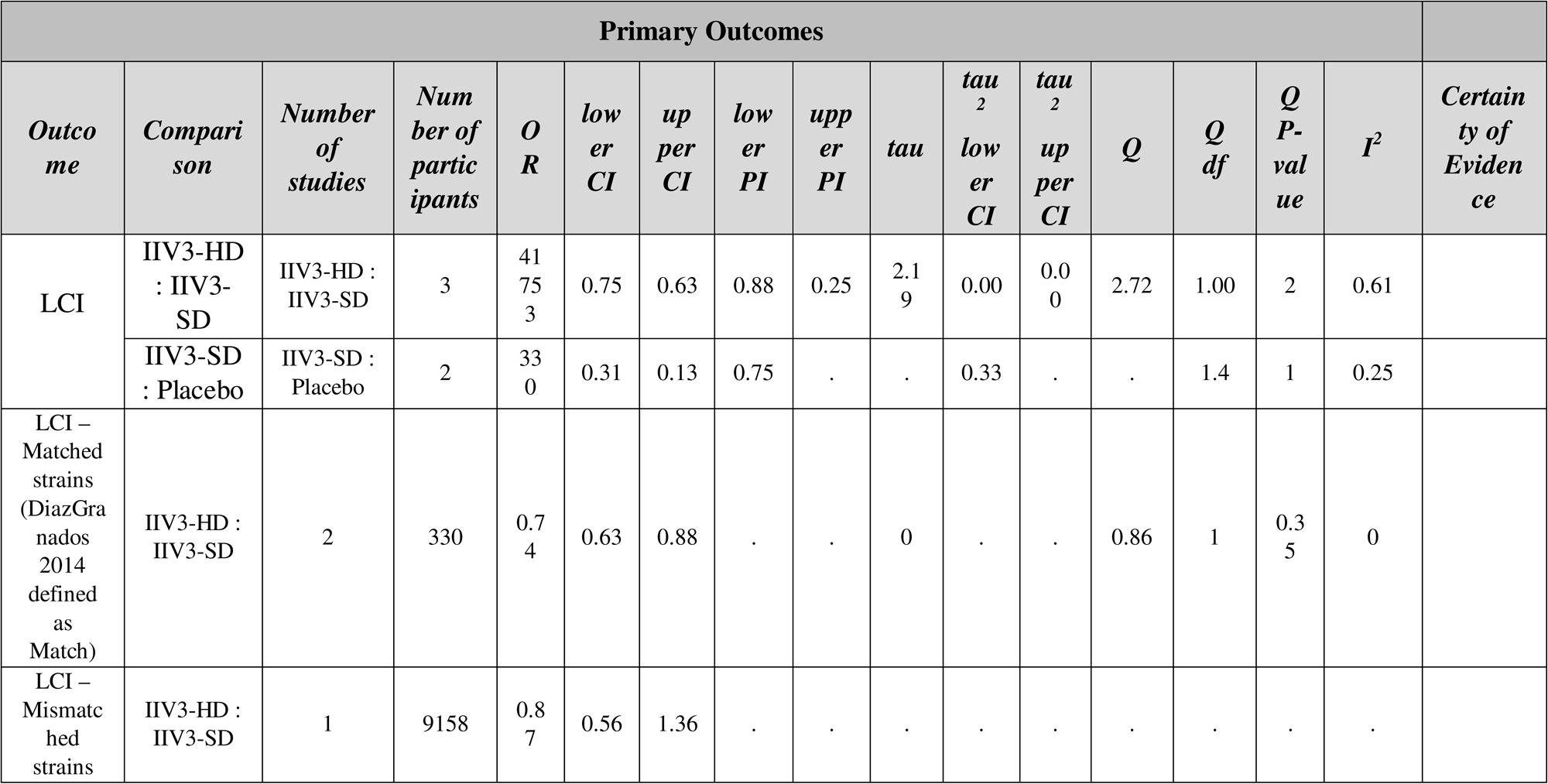
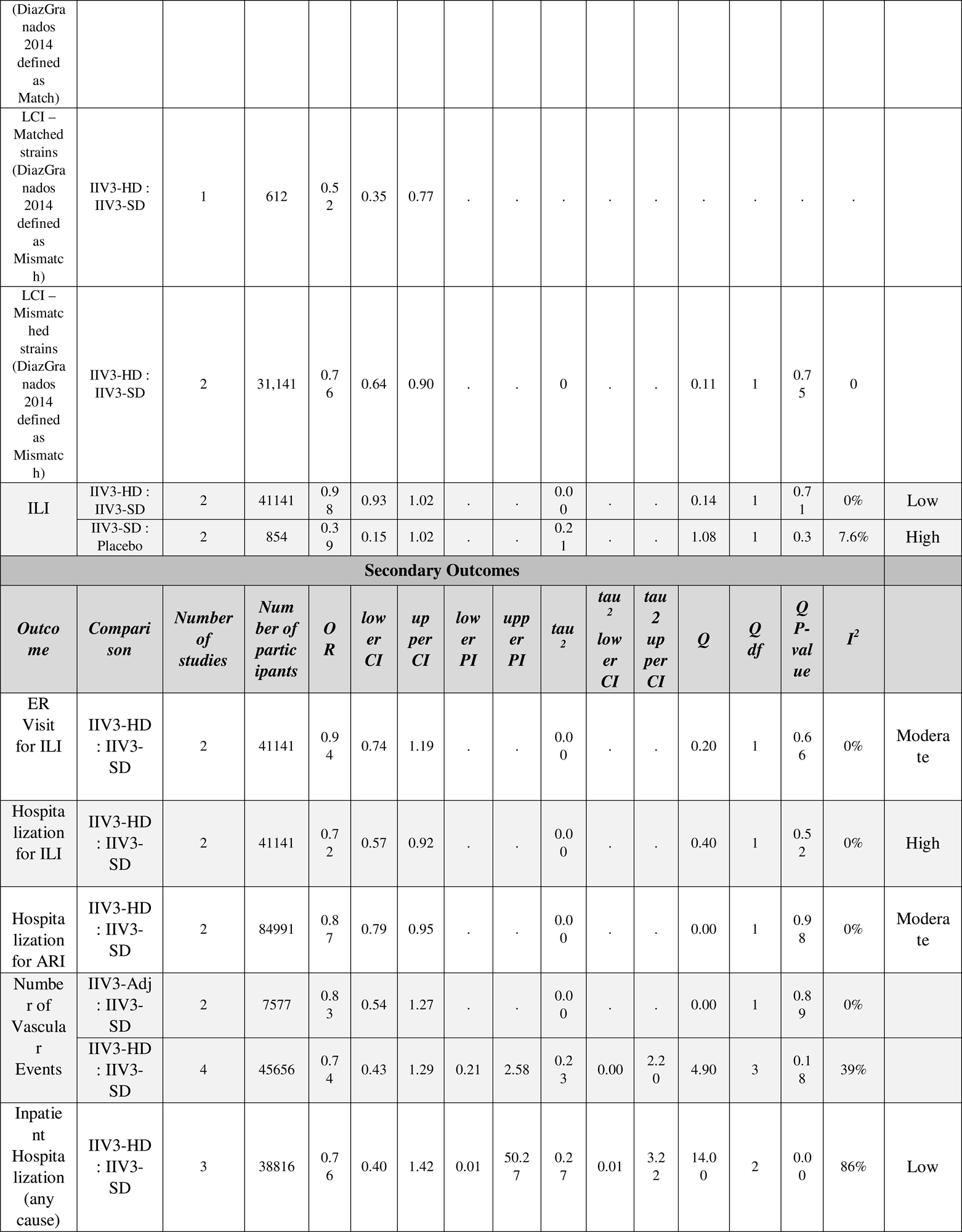
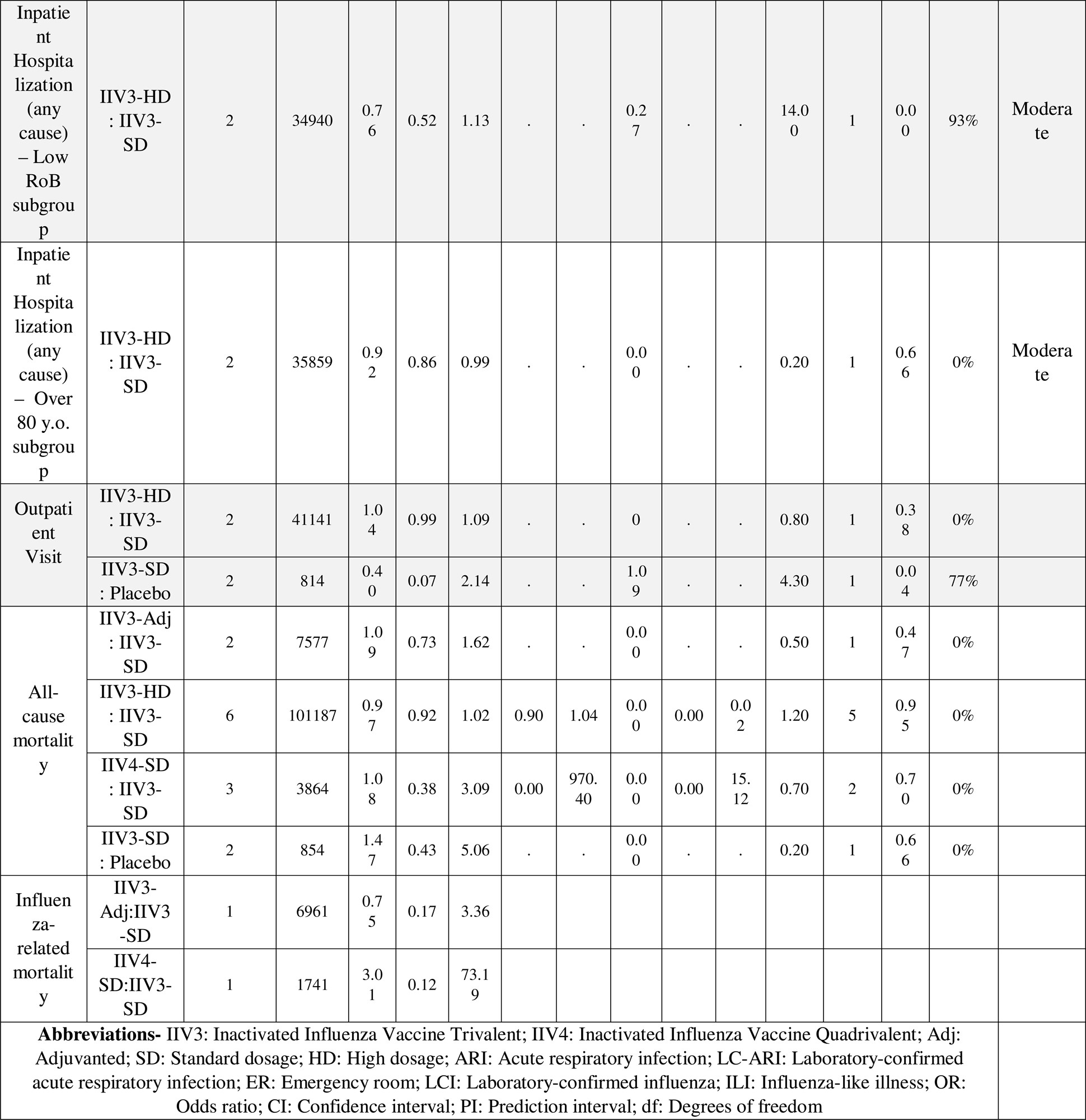
Summary of results in outcomes with pairwise meta-analysis.

When vaccine formulations were combined in a sensitivity analysis, a NMA was possible, including nine RCTs,^36, 39, 45–47, 50, 54, 69, 89^ five interventions (Adj, HD, RIV, SD, and Tdap) compared to placebo, and 65,658 participants. HD performed best, yet the 95% PI suggested that new evidence may change results (OR 0.38, 95%CI [0.15 to 0.93], P-score= 0.77, 95% PI [0.09 to 1.63], I^2^=0%, τ=0.00). Similar precision of the effect estimate was identified for RIV (OR 0.38, 95%CI [0.15 to 0.94], P-score= 0.68, 95% PI [0.09 to 1.66]) and SD (OR 0.38, 95%CI [0.16 to 0.95], P-score= 0.57, 95% PI [0.09 to 1.67]), yet PIs were wide (Appendix 17C (Additional file 1)).

#### Secondary outcomes analyzed in both network and pairwise meta-analysis

##### Number of vascular adverse events

The NMA for the number of vascular adverse events included eight RCTs,^45, 46, 48, 49, 64, 71, 87^ five interventions, and 57,677 participants (see Appendix 24 (Additional file 1) for study outcome definitions). Results suggested that HD and adjuvanted vaccines prevented more vascular adverse events compared with IIV3-SD, but with large dispersion among studies (I^2^=97%, τ=0.34). IIV4-Adj was ranked the most efficacious intervention and was associated the least with vascular adverse events (IRR 0.18, 95%CI [0.07 to 0.43], P-score= 0.98, 95% PI [0.03 to 1.07]; very low certainty of evidence) followed by IIV4-HD (IRR 0.46, 95%CI [0.21 to 1.00], P-score= 0.71, 95% PI [0.09 to 2.41]; low certainty of evidence) compared with IIV3-SD (Table 4, Appendix 17C (Additional file 1)). However, PIs were wide. Results were similar when restricting to RCTs with low overall ROB, and there was low between-study heterogeneity (I^2^=0%, τ=0.00), but more imprecision around the summary effect estimates.

##### Outpatient visits

The NMA for the outpatient visits included four RCTs,^36, 45, 46, 89^ three interventions (including placebo), and 41,995 participants. IIV3-SD (OR 0.654, 95%CI [0.36 to 1.14]) and IIV3-HD (OR 0.665, 95%CI [0.35 to 1.19]) were superior to placebo in being less associated with visits, but with imprecise effect estimates, moderate between-study network heterogeneity (I^2^=61%, τ=0.11), and very low certainty of evidence. Pairwise meta-analysis suggested an imprecise effect for IIV3-SD compared with placebo (OR 0.40, 95%CI [0.07 to 2.14], 2 RCTs, 814 participants, I^2^=77%, τ=1.08) (Table 4).

##### All-cause mortality

The NMA for all-cause mortality included 20 RCTs,^36, 37, 39, 40, 45–52, 54, 58, 60, 65, 86, 88, 89^ 9 interventions (including placebo), and 140,577 participants (Table 4). The effect of RIV on all-cause mortality was comparable to placebo (OR 1.01, 95%CI [0.23 to 4.49], low certainty of evidence) and both ranked among the best interventions (P-score: RIV 0.75, placebo 0.71). Despite the low between-study heterogeneity in the network (I^2^=0%, τ=0.00), effect estimates were highly imprecise with very low to moderate certainty of evidence. Pairwise meta-analysis estimate of IIV3-SD against placebo was highly uncertain (OR 1.47, 95%CI [0.43 to 5.06], 2 RCTs,^36, 89^ 854 participants, I^2^=0%, τ=0).

Inconclusive results were provided in sensitivity analyses restricting to studies with percentage of female above 50% and overall low ROB (Appendix 17C (Additional file 1)). One RCT^50^ (6,961 participants) showed that IIV3-Adj was associated with lower odds of influenza-related deaths compared with IIV3-SD (OR 0.75, 95%CI [0.17 to 3.36]), yet the estimate was imprecise. Another RCT^86^ (1,741 participants) suggested that IIV3-SD was associated with higher odds of influenza related deaths compared with IIV4-SD, yet the result was imprecise (OR 3.01, 95%CI [0.12 to 73.92]) (Table 5).

## DISCUSSION

This systematic review and NMA provides a comprehensive assessment of influenza vaccine efficacy in older adults. We included a total of 41 RCTs, involving over 200,000 participants. Our results indicated that influenza vaccines were effective in preventing LCI compared with placebo. IIV3-HD showed the most promising results in preventing LCI, followed by RIV, IIV3-Adj, and IIV3-SD. However, precision of effect estimates varied among interventions with low overall certainty of evidence, except for the comparison IIV3-HD vs IIV3-SD where certainty was high. The findings highlight the potential benefits of IIV3-HD in preventing hospitalizations for ILI and hospitalization for ARI, and of IIV4-Adj in reducing vascular adverse events among older adults. Overall, few trials were included in the intervention comparisons in LCI, ILI, vascular adverse events, hospitalizations for ILI and ARI, and influenza-related mortality outcomes. Vaccine efficacy may differ across certain participant characteristics, such as age, sex or circulating strains between and within seasons. However, there was insufficient evidence to make definite conclusions or explore heterogeneity in populations with different characteristics (e.g., co-morbidities, frailty) or study characteristics (e.g., circulating strains or between influenza season variability) that could modify the intervention effect. Within-study risk of bias was predominantly at some or high concerns across outcomes, while small-study effects and reporting bias was only possible to assess in all-cause mortality outcome with no overall concerns. Overall, our results should be interpreted with caution, since uncertainty around the point estimates was high and certainty of evidence was primarily low. High certainty of evidence was in LCI (for one of the 15 comparisons), ILI-hospitalization (for the single comparison), and ILI cases (for the single comparison), but moderate in vascular adverse events (for one of the 10 comparisons), all-cause mortality (for 15 of the 36 comparisons), ARI-hospitalization (for the single comparison), and ER-visits (for the single comparison).

Our findings align with the systematic review and NMA conducted by Minozzi et *al.*,^91^ who assessed the efficacy and safety of influenza vaccines in 220 RCTs of 100,677 children, 329,127 adults, including older adults. Overall, NMA results from their review among the older adult population were consistent with our results in the sense that flu vaccines are better than placebo for LCI, albeit with varying levels of precision in effect estimates. Differences in pairwise results between the two reviews were unremarkable. Another review from the Advisory Committee on Immunization Practices (ACIP)^92^ included 51 studies of which 27 were RCTs and 24 were non-randomized studies to assesses the relative efficacy and safety of influenza vaccines in older adults. They did not conduct any RCT-based meta-analyses that could be compared with those in our review. Relative to both reviews, searches in our review were more comprehensive and up to date including 18 additional RCTs with 71,049 participants not previously included in these reviews. We also evaluated credibility of evidence using both GRADE and CINeMA approaches, which to our knowledge has not been done elsewhere.

The strengths of our study lie in its systematic approach, extensive knowledge user engagement, and rigorous analysis using network meta-analysis to compare various influenza vaccines. This integrated knowledge translation approach allowed for valuable insights throughout the research process, from refining the research question to interpreting the findings. We followed the Cochrane Handbook methods for systematic reviews,^22^ and reviewers worked in pairs and independently for screening, data abstraction, and risk of bias appraisal. We reported the results using the PRISMA-NMA^8^ and GRIPP-2 reporting statements.^9^

However, some limitations should be considered. First, the limited number of studies, constrained the analysis for specific outcomes, such as ILI, ER-visit for ILI, hospitalization for ILI and ARI outcomes. The sparsity of networks, in some cases with only one or two studies per intervention comparison, increased the imprecision of effect estimates and reduced statistical power to detect differences in effects. Second, the lack of studies in certain outcomes comparing enhanced influenza vaccines limited the adequate assessment of the NMA assumptions, including exploring sources of heterogeneity (e.g., through meta-regression analysis), small-study effects, and inconsistency. Also, demographic variables that may explain heterogeneity, such as frailty, comorbidities and season intensities, were absent from the primary studies. RCTs usually were limited to one or two influenza seasons and because of the characteristics of influenza viruses (high mutation) and the history of exposure to influenza viruses in the population (herd immunity, effect of repeated vaccination, and protection from previous exposure to influenza viruses) it is very hard to predict the efficacy of influenza vaccines in future seasons. Also, studies usually include relatively healthy populations, and there is a need for high quality RCT among older frail adults. Heterogeneity among studies varied in NMA from low to severe, but in all pairwise meta-analyses it was generally low, except for the inpatient hospitalization outcome. While no major concerns were identified for small-study effects and publication bias for the all-cause mortality outcome, we could not assess this for any other outcome. For LCI and all-cause mortality outcomes where it was possible to assess, there was no evidence of statistical inconsistency. The transitivity assumption was likely satisfied regarding the overall ROB and participants’ age. Lastly, we conducted a subgroup analysis of LCI with respect to whether vaccine strains matched the circulating strains. A more targeted analysis would have looked at specific strains; however, there was an insufficient number of studies to undertake such an analysis.

Our study contributes valuable insights to inform evidence-based public health decisions and immunization guidelines for older adults. Policymakers and healthcare providers should consider these findings when formulating immunization strategies to protect older and vulnerable populations from seasonal influenza and its complications. Annual vaccination with any vaccine is the best way to prevent infection and its complications.

## CONCLUSIONS

Our analysis of high quality RCT data highlights the high efficacy associated with IIV3-HD and RIV vaccines in protecting elderly persons against LCI and RIV vaccine minimizing all-cause mortality when compared with other vaccines we examined. Moreover, our analysis points to a possible association of higher all-cause mortality in elderly persons receiving adjuvant vaccines (IIV3-Adj and IIV4-Adj). The comparative efficacy of individual vaccines regarding LCI, ILI, and secondary outcomes (e.g., influenza-related mortality) remains uncertain. Differences among vaccines are possible and might emerge with accumulation of new evidence.

Our results should be interpreted with caution due to imprecise and heterogeneous summary effects with low certainty of evidence of the RCT studies we included. Vaccine efficacy may vary depending on population (e.g., frailty, comorbidities) virus strain matching and study characteristics. More high quality RCTs are necessary that encompass multiple flu seasons to evaluate the relationship between these factors and vaccine efficacy, such as seasonal vaccines specifically based on RIV and IIV4 formulations.

## LIST OF ABBREVIATIONS

LCI: Laboratory-Confirmed Influenza
RCT: Randomized Control Trial
NMA: Network Meta-Analysis
NACI: National Advisory Committee on Immunization
PRISMA: Preferred Reporting Items for Systematic Reviews and Meta-analyses
GRIPP-2: Guidance for Reporting Involvement of Patients and the Public
EBP: Evidence-Based Practice
PRESS: Peer Review of Electronic Search Strategies
CADTH: Canadian Agency for Drugs and Technologies in Health
NITAG: National Immunization Technical Advisory Group
IIV4: Quadrivalent inactivated influenza vaccine
SD: Standard Dose
HD: High Dose
Adj: Adjuvanted
IIV3: Trivalent inactivated influenza vaccine
RIV4: Quadrivalent recombinant influenza vaccine
RIV3: Trivalent recombinant influenza vaccine
GRADE: Grading of Recommendations, Assessment, Development, and Evaluations
ILI: Influenza-Like Illness
ARI: Acute Respiratory Infection
ER: Emergency Room
ROB: Risk of Bias
OR: Odds Ratios
IRR: Incidence Rate Ratios
CI: Confidence Intervals
HKSJ: Hartung-Knapp-Sidik-Jonkman
PI: Prediction Interval
Tdap: Tetanus-diphtheria-pertussis vaccine
VE: Vaccine Efficacy
CINeMA: Confidence in Network meta-analysis
df: Degrees of freedom
ACIP: Advisory Committee on Immunization Practices

## DECLARATIONS

### Ethics approval and consent to participate

Not required.

### Consent for publication

Not applicable.

### Availability of data and materials

The full dataset and statistical code are available from the corresponding author upon reasonable request.

### Patient and public involvement

A patient partner (KE) was involved in defining the research question, interpreting the results, writing a lay summary (Additional file 4) and disseminating of the article.

### Authors’ Contributions

AAV, SES, and ACT designed the study. ACT and SES obtained funding. SST, MG, and JD coordinated the study. SST, JD, MG, PAK, VN, MC, RR, AP, CS screened articles, abstracted data, and carried out risk of bias assessments. AAV and MK conducted data analyses. IDF and JJYN conducted GRADE certainty of evidence assessments. AAV drafted the first version of the manuscript. All authors contributed to the manuscript’s revision and interpretation of findings. AAV is the guarantor. The corresponding author attests that all listed authors meet authorship criteria and that no others meeting the criteria have been omitted.

## Acknowledgements

The authors wish to thank Huda Ashoor, Areej A. Hezam, Myanca Rodrigues, Ji Yoon Kim, Patricia Rios, Amruta Radhakrishnan, and Kelsey Young for their contributions in screening and data abstracting the studies.

We thank Brahmleen Kaur and Navjot Mann for their support in formatting the paper and creating the EndNote library.

We thank Dr. Jessie McGowan for developing the literature searches, Tamara Rader for peer-reviewing the literature search, and Raymond Daniel for conducting the literature search.

We thank the National Advisory Committee on Immunization Influenza Working Group for their content expertise.

## ADDITIONAL FILES

**File name:** Additional file 1

**File format:** Microsoft Word

**Title of data:** Appendix Supplementary Content

**File name:** Additional file 2

**File format:** Microsoft Word

**Title of data:** PRISMA NMA Checklist of Items to Include When Reporting A Systematic Review Involving a Network Meta-analysis

**File name:** Additional file 3

**File format:** Microsoft Word

**Title of data:** GRIPP2 Checklist – Long Form

**File name:** Additional file 4

**File format:** Microsoft Word

**Title of data:** Comparing trivalent and quadrivalent seasonal influenza vaccine efficacy in persons 60 years of age and older: A systematic review and network meta-analysis

