## Additional file 1 for "Comparing trivalent and quadrivalent seasonal influenza vaccine efficacy in persons 60 years of age and older: A systematic review and network meta-analysis"

Appendix Supplementary Content

### **Appendix 1: Protocol amendments**

**Comparative effectiveness of influenza vaccines in adults 60 years of age and older: a systematic review and network meta-analysis**

| **Section** | **Previous protocol criteria** | **Changed criteria/deviations** | **Justification** |
| --- | --- | --- | --- |
| Review question | Which influenza vaccine available for adults 65 years of age and older is the most effective? | Which influenza vaccine available for adults 60 years of age and older is the most effective? | Some studies include data on older adults by referring to 60+. Therefore, to capture a broader sample of studies, we have updated the age to include 60+ instead of 65+. |
| Types of study to be included | Randomised controlled trials (RCTs), case test-negative studies, NRCTs (e.g., such as quasi-RCTs, non-randomised trials, interrupted time series, controlled before after), and observational studies (e.g., cohort, case control) will be included. Studies must have a control or comparator in order to be eligible for inclusion and as such, cross-sectional, case series, case reports, and qualitative studies will be excluded. | Randomised controlled trials (RCTs) will be included. Case test-negative studies, NRCTs (e.g., such as quasi-RCTs, non-randomised trials, interrupted time series, controlled before after), and observational studies (e.g., cohort, case control) will be excluded. Studies must have a control or comparator in order to be eligible for inclusion and as such, cross-sectional, case series, case reports, and qualitative studies will be excluded. | Due to feasibility reasons and resource concerns, case test-negative studies, NRCTs, and observational studies (e.g., cohort, case control) were excluded. |
| Participants/  population | All adults aged 65 years and older (studies with a median age of participants of 60 years will be included). | All adults aged 60 years and older will be included. | Some studies include data on older adults by referring to 60+. Therefore, to capture a broader sample of studies, we have updated the age to include 60+ instead of 65+. |
| Intervention(s), exposure(s) | Any influenza vaccines for adults 65 years of age and older (e.g. trivalent standard dose influenza vaccine, quadrivalent standard dose influenza vaccine, adjuvanted trivalent influenza vaccine, trivalent high dose influenza vaccine, egg-based vaccines). Both pandemic and seasonal influenza vaccines will be considered for inclusion. | Any influenza vaccines for adults 60 years of age and older licensed for use in Canada and/or the United States (e.g. trivalent standard dose influenza vaccine, quadrivalent standard dose influenza vaccine, adjuvanted trivalent influenza vaccine, trivalent high dose influenza vaccine, egg-based vaccines). | Pandemic vaccines were not considered in this review as they are not relevant to the main objective of this research, as per the guidance received from stakeholders and subject matter experts. |
| Main outcome | Cases of laboratory confirmed influenza and cases of influenza like illness (ILI). | Vaccine efficacy against infection. Both laboratory confirmed symptomatic infection (LCI) as well as influenza like illness (ILI) will be considered for inclusion and analysed separately. | Main outcome defined as vaccine efficacy will be explored. |
| Secondary outcomes | Secondary outcomes include:  - All cause hospitalization  - Hospitalization due to influenza infection  - Hospitalization due to pneumonia infection  - Adverse vascular events (non-fatal myocardial infarction, stroke, cardiovascular death, hospitalization for heart failure)  - Mortality from influenza infection | Secondary outcomes include:   - Acute respiratory illness (ARI) cases - Laboratory confirmed-ARI - Hospitalization for ILI - Emergency room (ER) visit for ILI - Hospitalization for ARI - ER visit for ARI - Hospitalization for LCI - ER visit for LCI - Hospitalization for pneumonia due to ILI - Hospitalization for pneumonia due to ARI - Hospitalization for pneumonia due to LCI - ER visit for pneumonia - Hospitalization/inpatient for any cause - ER Visit (any cause) - Inpatient/outpatient visit - Outpatient visit - LCI-related healthcare interactions - Number of vascular adverse events (AEs) - Number of participants with vascular AEs - Hospitalizations due to cardiovascular events - LCI-related mortality - All-cause mortality | Additional outcomes of interest were identified collaboratively with stakeholders and subject matter experts. |
| Strategy for data synthesis | Random-effects network meta-analysis  (NMA) will be conducted, by vaccine type, to compare the effectiveness of available vaccines for  seasonal and pandemic influenza separately. | The NMA will be conducted in R using the netmeta package.  We assessed small-study effects and publication bias in the outcomes with at least 10 studies and using the comparison-adjusted funnel plot and the following chronological order of the interventions and based on Barberis et al., 2016^1^   - IIV4 Adjuvanted - IIV4 High Dose - IIV4 Standard dose (2012) - IIV3 Adjuvanted (1997) - IIV3 High dose (2009) - IIV3 Standard dose - Recombinant Trivalent (RIV3) (2013) - Recombinant Quadrivalent (RIV4) - Tdap (Tetanus, diphtheria, and pertussis) - Placebo | Random-effects meta-analysis was conducted, by vaccine type due to the reduced scope of the project. |
| Analysis of subgroups or subsets | If feasible (i.e., sufficient amount of studies and data), the following sub-group analyses will be considered:  - Age: 65-79 years of age vs. ≥80 years of age  - Healthy vs. individuals with comorbid conditions (defined as conditions that last for one or more years, require ongoing medical attention and/or limit activities of daily living)  - Sex (female vs. male)  - Previous vaccination with any influenza vaccine vs. vaccine-naïve  - Individuals who are frail vs. those who are not  - Vaccines with matched vs. unmatched strains | We conducted the following analyses:  a.       LCI (NMA): sex  b.      Outpatient visits (NMA): none  c.       All-cause mortality (NMA): sex, rob  d.      Vascular events (NMA): none  e.       Pairwise meta-analysis: LCI (none), outpatient (none), all-cause mortality (rob, age>80 vs age<80), inpatient hospitalizations any cause (low rob, age> 80 vs <80) | We were unable to assess transitivity using healthy versus chronic disease (comorbidities) and previous vaccination, since this was rarely reported in the publications.  We were only able to explore sex (% of females below and above 50%), age (below and above 80 years) and risk of bias (restricting to low risk of bias studies) in pairwise and network meta-analysis.  We also conducted network meta-analyses as a sensitivity analysis for the primary outcomes using a revised categorization of the nodes. Specifically:  IIV3-SD and IIV4-SD were defined as SD  IIV3-HD and IIV4-HD were defined as HD  RIV3 and RIV4 were defined as RIV  IIV3-Adj and IIV4-Adj were defined as Adj |
| Risk of bias (quality) assessment | Risk of bias appraisal will be carried out independently and in duplicate by 2 reviewers using the Cochrane Risk of Bias tool RCTs, the Risk Of Bias In Non-randomised Studies - of Interventions (ROBINS-I) tool for non-randomised intervention studies. | Risk of bias appraisal will be carried out independently and in duplicate by 2 reviewers using the Cochrane Risk of Bias tool RCTs. | Due to the exclusion of NCRTs and observational studies, no risk of bias assessments will be completed using the ROBINS-I tool. |
| Summary of Findings | NA | We assessed credibility of evidence using CINeMA in the outcomes that NMA was possible and GRADE in the outcomes where a pairwise meta-analysis was conducted but no NMA was conducted. | For completeness and to inform decision making we assessed certainty and quality of evidence. |
| 1. Barberis I, Myles P, Ault SK, Bragazzi NL, Martini M. History and evolution of influenza control through vaccination: from the first monovalent vaccine to universal vaccines. J Prev Med Hyg. 2016 Sep;57(3):E115-E120. PMID: 27980374; PMCID: PMC5139605. | | | |

### **Appendix 2: Methods on selection process, data extraction, risk of bias, and synthesis**

#### **Selection process**

A screening form for the title and abstract stage was developed in collaboration with Public Health Agency of Canada and the research team based on pre-defined eligibility criteria. Screening was conducted in two stages using Synthesi.SR:^1^ level one screening of titles and abstracts (Appendix 5 (Additional file 1)) and level two screening of full-text articles (Appendix 6 (Additional file 1)). First, the team conducted one pilot-test at level one of 50 random titles/abstracts resulting in 74% agreement, and another at level two of 25 random full-texts resulting in 70% agreement among the team. Next, two reviewers (MG, PAK, VN, MC, AP) independently screened the identified citations for study eligibility. Conflicts at both screening levels were resolved by a third reviewer (MG).

#### **Data items and data abstraction**

A data abstraction form (Appendix 7 (Additional file 1)) was created to capture data on the study characteristics (e.g. duration of follow-up, study design, country of conduct, multi-center vs. single site), patient characteristics (e.g., mean age, age range, co-morbidities, frailty), intervention characteristics (e.g., type of vaccine, dose, unadjuvanted versus adjuvanted), and outcome characteristics (e.g., number of patients with influenza, acute respiratory infection, vascular adverse events, hospitalizations, and mortality at the longest duration of follow-up). Prior to data abstraction, the form was piloted on a random sample of five included articles. When a high level of agreement was reached, two reviewers SST, MG, PAK, VN, MC, RR, AP, CS) independently abstracted data from each study. Conflicts were resolved by a third reviewer (SST, MG, VN).

#### **Within and across study bias assessment**

Once a pilot-test was carried out for risk of bias assessment on a random sample of five studies, two reviewers (JD, PAK, VN, RR) independently appraised the included RCTs using the Cochrane Risk of Bias (ROB) 2 tool.^2^ Conflicts were resolved by a third reviewer (MK).

We visually inspected small-study effects and reporting bias using the comparison-adjusted funnel plot when at least ten studies were available per outcome.^3^ For this analysis, the interventions were ordered chronologically^4^ according to when they were licensed in Canada (or US where applicable).

#### **Exploring the impact of effect modifiers via subgroup analyses**

For the primary outcomes and outcomes displaying evidence of inconsistency, intransitivity or heterogeneity in any NMA, we performed subgroup and sensitivity analyses considering the predefined potential treatment effect modifiers and monitored changes in heterogeneity and effect estimates, whenever possible. In our primary outcomes, we also compared match and mismatch between the circulating and vaccine strains. To determine matched and mismatched studies we followed previously published methods.^5^ We categorized a study as matched when the influenza strains were antigenically similar to the vaccine stains. Similarly, we categorized a study as mismatched when the influenza strains were antigenically distinct from vaccine strains. When the information on matching was not reported in the study, we determined the matching using surveillance data (Appendix 25 and 26 (Additional file 1)). We combined intervention nodes for placebo and for trivalent and quadrivalent formulation by vaccine type (i.e., adjuvant, standard dose, high dose, and RIV). At the request of the NACI Influenza Working Group, we excluded tetanus-diphtheria-pertussis vaccine (Tdap) in a sensitivity analysis, where Tdap was included as an intervention.

#### **Assessment of the confidence in the evidence**

Confidence in NMA estimates was assessed for each outcome using CINeMA (Confidence in Network meta-analysis)^6^ as very low, low, moderate or high on the basis of six domains (within-study bias, reporting bias, indirectness, imprecision, heterogeneity and incoherence). Each domain was evaluated as ‘no concerns’, ‘some concerns’ or ‘major concerns’. We used the overall risk of bias per study, and for each treatment comparison we applied the average risk of bias over the studies. Similarly, for all treatment comparisons we used the average for indirectness. We assessed reporting bias based on the comparison-adjusted funnel plot, where possible. Downgrading of the confidence rating was conducted whenever any of the domains except for reporting bias was rated as ‘some concerns’ or ‘major concerns’.

For imprecision, we considered the null effect (e.g., OR=1) for the evaluation of the clinically important size of effect, where statistical significance and clinical importance would coincide. Heterogeneity and incoherence (i.e., inconsistency) were assessed by following the standard CINeMA approach.^6^ As a complimentary approach in the NMA outcomes, and based on previous publications and content expert advice, we considered the following to be clinically important effect sizes: risk ratio at 67% in LCI (or OR at 63.5%), risk ratio at 85% in vascular adverse events^7^ (or IRR at 85%), risk ratio at 75% in outpatient visits (or OR at 72%), risk ratio at 5% in all-cause mortality^8^ (or OR at 34%).

Confidence in pairwise meta-analysis estimates for which a NMA could not be performed was assessed by two reviewers (IVF, JJYN) independently using the GRADE approach^9^. Disagreements were resolved by discussion.

### **Appendix 3: Database search strategies**

**March 30, 2022**

**Ovid MEDLINE(R) ALL <1946 to March 22, 2022>**

--------------------------------------------------------------------------------
1     influenza, human/ or exp influenzavirus a/ or exp influenzavirus b/ or influenzavirus c/ or Hemagglutinin Glycoproteins, Influenza Virus/

2     (flu or flue or influenza* or grippe).tw,kf.
3     1 or 2
4     exp Vaccines/ or Immunization/
5     (vaccin* or immuni* or inocula* or shot or jab).tw,kf.
6     4 or 5
7     3 and 6
8     exp Adjuvants, Immunologic/
9     exp Antibodies, Viral/
10     Hemagglutination Inhibition Tests/ or Plant Proteins/im or (adjuvant* or squalene* or emulsion*).mp.

11     or/8-10
12     3 and 11
13     influenza vaccines/
14     influenza vaccine*.tw,kf.

15     13 or 14
16     7 or 12 or 15

**Core set 1**
17     (trivalent or quadrivalent or TIV or aTIV or QIV or aQIV).tw,kf.
18     16 and 17

**Core set 2 (limited to trivalent or quadrivalent)**
19     (flulaval or fluzone or fluad* or fluzone or agriflu or fluviral or influvac or alfuria or fluarix or flucelvax or flublok).tw,kf,rn.
20     (MF59* or MF?59* or aTIV or aQIV or chiromas or gripguard or Influpozzi Adiuvato or aIIV3* or aIIV4*).tw,kf.
21     19 or 20
22     18 or 21

**Core set 3 (core set 3 plus all known drug names)**
23     limit 22 to "all aged (65 and over)"
24     exp Aged/ or geriatrics/ or aging/
25     (geriatric* or elder* or old* or ageing or aging or senior* or older adult* or retired or retiree* or elder* or pensioner* or nursing home* or older people or older patient* or gerontology or Sexagenarian*OR septuagenarian* or octogenarian or nonagenarian*OR centenarian* or sixties or seventies or eighties or nineties).tw,kf.
26     22 and (24 or 25)
27     23 or 26
28     animals/ not humans/
**29     27 not 28**
30     21 not 28

**31     29 or 30 – Final set with all named drug articles**

**Database: Embase Classic+Embase <1947 to 2022 March 29>**
--------------------------------------------------------------------------------
1     exp Influenza virus/ or exp influenza/ or exp influenzavirus a/ or Hemagglutinin Glycoproteins, Influenza Virus/
2     (flu or flue or influenza*or grippe).tw.
3     1 or 2
4     exp vaccine/ or exp immunization/
5     (vaccin* or immuni* or inocula* or shot or jab).tw.
6     4 or 5
7     3 and 6
8     immunological adjuvant/
9     exp virus antibody/
10     hemagglutination inhibition test/ or exp plant protein/ or (adjuvant* or squalene* or emulsion*).mp.
11     or/8-10
12     3 and 11
13     influenza vaccine/
14     influenza vaccine*.tw.
15     13 or 14
16     7 or 12 or 15
17     (trivalent or quadrivalent or TIV or aTIV or QIV or aQIV).tw.
18     16 and 17
19     (flulaval or fluzone or fluad* or fluzone or agriflu or fluviral or influvac or alfuria or fluarix or flucelvax or flublok).tw.
20     (MF59* or MF?59* or aTIV or aQIV or chiromas or gripguard or Influpozzi Adiuvato or aIIV3* or aIIV4*).tw.
21     19 or 20
22     18 or 21
23     limit 22 to aged <65+ years>
24     exp aged/ or exp aging/ or exp geriatrics/
25     (geriatric* or elder* or old* or ageing or aging or senior* or older adult* or retired or retiree* or elder* or pensioner* or nursing home* or older people or older patient* or gerontology or Sexagenarian*OR septuagenarian* or octogenarian or nonagenarian*OR centenarian* or sixties or seventies or eighties or nineties).tw.
26     22 and (24 or 25)
27     23 or 26
28     animals/ not humans/
29     27 not 28
30     21 not 28
31     29 or 30

**Database: JBI EBP Database <Current to March 23, 2022>
Search Strategy:**
--------------------------------------------------------------------------------
1     (flu or flue or influenza* or grippe or hemagglutinin).tw.
2     (vaccin* or immuni* or inocula* or shot or jab).tw.
3     (adjuvant* or squalene* or emulsion*).mp.
4     1 and (2 or 3)
5     (trivalent or quadrivalent or TIV or aTIV or QIV or aQIV).tw.
6     (flulaval or fluzone or fluad* or fluzone or agriflu or fluviral or influvac or alfuria or fluarix or flucelvax or flublok).tw.
7     (MF59* or MF?59* or aTIV or aQIV or chiromas or gripguard or Influpozzi Adiuvato or aIIV3* or aIIV4*).tw.
8     or/4-7

*Note: no age limit

**Database: APA PsycInfo <1806 to March Week 3 2022>**

--------------------------------------------------------------------------------
1     exp influenza/
2     (flu or flue or influenza*or grippe or Hemagglutinin).tw.
3     1 or 2
4     immunization/
5     (vaccin* or immuni* or inocula* or shot or jab).tw.
6     (adjuvant* or squalene* or emulsion*).tw.
7     or/4-6
8     3 and 7
9     (trivalent or quadrivalent or TIV or aTIV or QIV or aQIV).tw.
10     8 and 9
11     (flulaval or fluzone or fluad* or fluzone or agriflu or fluviral or influvac or alfuria or fluarix or flucelvax or flublok).tw.
12     (MF59* or MF?59* or aTIV or aQIV or chiromas or gripguard or Influpozzi Adiuvato or aIIV3* or aIIV4*).tw.
13     10 or 11 or 12

14	animals/ not humans/

15	13 not 14

*Note: no age limit

**Database: EBM Reviews** - Cochrane Database of Systematic Reviews <2005 to March 24, 2022>, EBM Reviews - ACP Journal Club <1991 to March 2022>, EBM Reviews - Database of Abstracts of Reviews of Effects <1st Quarter 2016>, EBM Reviews - Cochrane Clinical Answers <March 2022>, EBM Reviews - Cochrane Central Register of Controlled Trials <January 2022>, EBM Reviews - Cochrane Methodology Register <3rd Quarter 2012>, EBM Reviews - Health Technology Assessment <4th Quarter 2016>, EBM Reviews - NHS Economic Evaluation Database <1st Quarter 2016>
Search Strategy:
--------------------------------------------------------------------------------
1     (flu or flue or influenza* or grippe or hemagglutinin).tw.
2     (vaccin* or immuni* or inocula* or shot or jab).tw.
3     (adjuvant* or squalene* or emulsion*).mp.
4     1 and (2 or 3)
5     (trivalent or quadrivalent or TIV or aTIV or QIV or aQIV).tw.
6     4 and 5
7     (geriatric* or elder* or old* or ageing or aging or senior* or older adult* or retired or retiree* or elder* or pensioner* or nursing home* or older people or older patient* or gerontology or Sexagenarian*OR septuagenarian* or octogenarian or nonagenarian*OR centenarian* or sixties or seventies or eighties or nineties).tw.
8     6 and 7
9     (flulaval or fluzone or fluad* or fluzone or agriflu or fluviral or influvac or alfuria or fluarix or flucelvax or flublok).tw.
10     (MF59* or MF?59* or aTIV or aQIV or chiromas or gripguard or Influpozzi Adiuvato or aIIV3* or aIIV4*).tw.
11     9 or 10
12     8 or 11

*Note: no need to exclude animals

**UPDATED SEARCHES - June 17, 2022**

**Database: Ovid MEDLINE(R) ALL <1946 to June 16, 2022>**

--------------------------------------------------------------------------------
1     influenza, human/ or exp influenzavirus a/ or exp influenzavirus b/ or influenzavirus c/ or Hemagglutinin Glycoproteins, Influenza Virus/
2     (flu or flue or influenza* or grippe).tw,kf.
3     1 or 2
4     exp Vaccines/ or Immunization/
5     (vaccin* or immuni* or inocula* or shot or jab).tw,kf.
6     4 or 5
7     3 and 6
8     exp Adjuvants, Immunologic/
9     exp Antibodies, Viral/
10     Hemagglutination Inhibition Tests/ or Plant Proteins/im or exp Recombinant Proteins/ or (adjuvant* or squalene* or emulsion*).mp.
11     or/8-10
12     3 and 11
13     influenza vaccines/
14     influenza vaccine*.tw,kf.
15     13 or 14
16     7 or 12 or 15

**Core set 1**
17     (trivalent or quadrivalent or TIV or aTIV or QIV or aQIV).mp.
18     16 and 17

**Core set 2 (limited to trivalent or quadrivalent)**
19     (flulaval or fluzone or fluad* or fluzone or agriflu or fluviral or influvac or alfuria or fluarix or flucelvax or flublok or Supemtek).tw,kf,rn.
20     (MF59* or MF?59* or aTIV or aQIV or chiromas or gripguard or "Influpozzi Adiuvato" or aIIV3* or aIIV4*).tw,kf.
21     (RIV or RIV3 or RIV4 or IIV3* or SD-IIV3 or HD-IIV3 or ccIIV3 or cc-IIV3 or cIIV3 or eIIV4 or IIV4* or SD-IIV4 or HD-IIV4 or ccIIV4 or cc-IIV4 or cIIV4 or eIIV4).tw,kf.
22     or/19-21
23     18 or 22

**Core set 3: core set 3 PLUS all known drug names and abbreviations – Note that the known drug names and abbreviations have NOT been combined with terms for influenza or vaccines**

24     limit 23 to "all aged (65 and over)"
25     exp Aged/ or geriatrics/ or aging/
26     (geriatric* or elder* or old* or ageing or aging or senior* or "older adult*" or retired or retiree* or elder* or pensioner* or "nursing home*" or "older people" or "older patient*" or gerontology or Sexagenarian*OR septuagenarian* or octogenarian or nonagenarian* or centenarian* or sixties or seventies or eighties or nineties).tw,kf.
27     23 and (25 or 26)
28     24 or 27
29     animals/ not humans/
30     28 not 29
31     22 not 29
32     30 or 31 * **– Final set with all named drugs/abbreviations**

*RIV abbreviation includes rivaroxaban, so these articles were removed using “NOT rivaroxaban.tw,kf.”

**Database: EBM Reviews - Cochrane Database of Systematic Reviews <2005 to June 15, 2022>, EBM Reviews - ACP Journal Club <1991 to May 2022>, EBM Reviews - Database of Abstracts of Reviews of Effects <1st Quarter 2016>, EBM Reviews - Cochrane Clinical Answers <May 2022>, EBM Reviews - Cochrane Central Register of Controlled Trials <May 2022>, EBM Reviews - Cochrane Methodology Register <3rd Quarter 2012>, EBM Reviews - Health Technology Assessment <4th Quarter 2016>, EBM Reviews - NHS Economic Evaluation Database <1st Quarter 2016>**
--------------------------------------------------------------------------------
1 (flu or flue or influenza* or grippe).tw.
2 (vaccin* or immuni* or inocula* or shot or jab).tw.
3 (adjuvant* or squalene* or emulsion*).mp.
4 1 and (2 or 3)
5 (trivalent or quadrivalent or TIV or aTIV or QIV or aQIV).tw.
6 4 and 5
7 (geriatric* or elder* or old* or ageing or aging or senior* or older adult* or retired or retiree* or elder* or pensioner* or nursing home* or older people or older patient* or gerontology or Sexagenarian*OR septuagenarian* or octogenarian or nonagenarian*OR centenarian* or sixties or seventies or eighties or nineties).tw.
8 6 and 7
9 (flulaval or fluzone or fluad* or fluzone or agriflu or fluviral or influvac or alfuria or fluarix or flucelvax or flublok or Supemtek).tw.
10 (MF59* or MF?59* or aTIV or aQIV or chiromas or gripguard or Influpozzi Adiuvato or aIIV3* or aIIV4*).tw.
11 (RIV or RIV3 or RIV4 or IIV3* or SD-IIV3 or HD-IIV3 or ccIIV3 or cc-IIV3 or cIIV3 or eIIV4 or IIV4* or SD-IIV4 or HD-IIV4 or ccIIV4 or cc-IIV4 or cIIV4 or eIIV4).tw.
12 or/9-11
13 7 and 12
14 8 or 13

**Database: JBI EBP Database <Current to June 08, 2022>**
--------------------------------------------------------------------------------
1     (flu or flue or influenza* or grippe).tw.
2     (vaccin* or immuni* or inocula* or shot or jab).tw.
3     (adjuvant* or squalene* or emulsion*).mp.
4     1 and (2 or 3)
5     (trivalent or quadrivalent or TIV or aTIV or QIV or aQIV).tw.
6     (flulaval or fluzone or fluad* or fluzone or agriflu or fluviral or influvac or alfuria or fluarix or flucelvax or flublok).tw.
7     (MF59* or MF?59* or aTIV or aQIV or chiromas or gripguard or Influpozzi Adiuvato or aIIV3* or aIIV4*).tw.
8     (RIV or RIV3 or RIV4 or IIV3* or SD-IIV3 or HD-IIV3 or ccIIV3 or cc-IIV3 or cIIV3 or eIIV4 or IIV4* or SD-IIV4 or HD-IIV4 or ccIIV4 or cc-IIV4 or cIIV4 or eIIV4).tw.
9     or/4-8

**Database: APA PsycInfo <1806 to June Week 2 2022>**
--------------------------------------------------------------------------------
1 exp influenza/
2 (flu or flue or influenza*or grippe or Hemagglutinin).tw.
3 1 or 2
4 immunization/
5 (vaccin* or immuni* or inocula* or shot or jab).tw.
6 (adjuvant* or squalene* or emulsion*).tw.
7 or/4-6
8 3 and 7
9 (trivalent or quadrivalent or TIV or aTIV or QIV or aQIV).tw.
10 8 and 9
11 (flulaval or fluzone or fluad* or fluzone or agriflu or fluviral or influvac or alfuria or fluarix or flucelvax or flublok or Supemtek).tw.
12 (MF59* or MF?59* or aTIV or aQIV or chiromas or gripguard or Influpozzi Adiuvato or aIIV3* or aIIV4*).tw.
13 (RIV or RIV3 or RIV4 or IIV3* or SD-IIV3 or HD-IIV3 or ccIIV3 or cc-IIV3 or cIIV3 or eIIV4 or IIV4* or SD-IIV4 or HD-IIV4 or ccIIV4 or cc-IIV4 or cIIV4 or eIIV4).tw.
14 or/10-13
15 animals/ not humans/
16 14 not 15

**Database: Embase Classic+Embase <1947 to 2022 June 16>**
--------------------------------------------------------------------------------
1     exp Influenza virus/ or exp influenza/ or exp influenzavirus a/ or Hemagglutinin Glycoproteins, Influenza Virus/
2     (flu or flue or influenza*or grippe).tw.
3     1 or 2
4     exp vaccine/ or exp immunization/
5     (vaccin* or immuni* or inocula* or shot or jab).tw.
6     4 or 5
7     3 and 6
8     immunological adjuvant/
9     exp virus antibody/
10     hemagglutination inhibition test/ or exp plant protein/ or exp recombinant protein/ or (adjuvant* or squalene* or emulsion*).mp.
11     or/8-10
12     3 and 11
13     influenza vaccine/
14     influenza vaccine*.tw.
15     13 or 14
16     7 or 12 or 15
17     (trivalent or quadrivalent or TIV or aTIV or QIV or aQIV).tw.
18     16 and 17
19     (flulaval or fluzone or fluad* or fluzone or agriflu or fluviral or influvac or alfuria or fluarix or flucelvax or flublok or Supemtek).tw.
20     (MF59* or MF?59* or aTIV or aQIV or chiromas or gripguard or "Influpozzi Adiuvato" or aIIV3* or aIIV4*).tw.
21     (RIV or RIV3 or RIV4 or IIV3* or SD-IIV3 or HD-IIV3 or ccIIV3 or cc-IIV3 or cIIV3 or eIIV4 or IIV4* or SD-IIV4 or HD-IIV4 or ccIIV4 or cc-IIV4 or cIIV4 or eIIV4).tw.
22     or/19-21
23     18 or 22
24     limit 23 to aged <65+ years>
25     exp aged/ or exp aging/ or exp geriatrics/
26     (geriatric* or elder* or old* or ageing or aging or senior* or older adult* or retired or retiree* or elder* or pensioner* or nursing home* or older people or older patient* or gerontology or Sexagenarian*OR septuagenarian* or octogenarian or nonagenarian*OR centenarian* or sixties or seventies or eighties or nineties).tw.
27     23 and (25 or 26)
28     24 or 27
29     animals/ not humans/
30     28 not 29
31     22 not 29
32     30 or 31*

*RIV abbreviation includes rivaroxaban, so these articles were removed using “NOT rivaroxaban.tw.”

### **Appendix 4: Grey literature sources**

**Trial registries**

- CenterWatch Clinical Trials Listing Service: <https://www.centerwatch.com/clinical-trials/listings/>
- Clinicaltrials.gov: <https://clinicaltrials.gov/ct/screen/AdvancedSearch>
  - New trial in 2022 - Flublok or Fluzone With Advax-CpG55.2 or AF03 <https://clinicaltrials.gov/ct2/show/NCT03945825>
- EU Clinical Trials Register: <https://www.clinicaltrialsregister.eu/ctr-search/>
- Health Canada: <https://www.canada.ca/en/health-canada/services/drugs-health-products/drug-products/health-canada-clinical-trials-database.html>
- ISRCTN: <http://www.isrctn.com>
- UKCRN: <http://www.ukcrc.org/research-infrastructure/clinical-research-networks/uk-clinical-research-network-ukcrn/>
- WHO: <https://www.who.int/ictrp/en/>

**COVID databases**

- Cochrane: <https://covid-19.cochrane.org>
- CAMRADES COVID-19-SOLES: <https://camarades.shinyapps.io/COVID-19-SOLES/>
- COVID-NMA: <https://covid-nma.com>
- COVID-19 L*ove:
- <https://app.iloveevidence.com/loves/5e6fdb9669c00e4ac072701d?population=5e7fce7e3d05156b5f5e032a&intervention_variable=603b9fe03d05151f35cf13dc&classification=all>
- LitCOVID: <https://www.ncbi.nlm.nih.gov/research/coronavirus/>
- WHO COVID-19: <https://covid19.who.int>

**Pre-print databases**

- bioRxiv (biology): [www.biorxiv.org](http://www.biorxiv.org)
- JMIR Publications:  <https://preprints.jmir.org/>
- medRxiv: <https://www.medrxiv.org/>
- Open Science Framework: <https://osf.io>
- Preprints.org: <https://www.preprints.org>
- Research Square: <https://www.researchsquare.com>

**CDC**

- Quadrivalent Influenza Vaccine: <https://www.cdc.gov/flu/prevent/quadrivalent.htm>

Brand names: AFLURIA Quadrivalent, Fluarix Quadrivalent, FluLaval Quadrivalent, Flucelvax Quadrivalent and Fluzone Quadrivalent, FluMist Quadrivalent

- TABLE. Influenza vaccines — United States, 2021–22 influenza season* - <https://www.cdc.gov/flu/professionals/acip/2021-2022/acip-table.htm>
- Summary: ‘Prevention and Control of Seasonal Influenza with Vaccines: Recommendations of the Advisory Committee on Immunization Practices (ACIP)—United States, 2021-22’ - <https://www.cdc.gov/flu/professionals/acip/summary/summary-recommendations.htm>

**Alfuria**

- BCCDC: <http://www.bccdc.ca/resource-gallery/Documents/Guidelines%20and%20Forms/Guidelines%20and%20Manuals/Epid/CD%20Manual/Chapter%202%20-%20Imms/Part4/Influenza_AfluriaTetra.pdf>
- FDA: <https://www.fda.gov/vaccines-blood-biologics/vaccines/afluria-afluria-southern-hemisphere>
- Government of Canada: <https://www.canada.ca/en/public-health/services/publications/healthy-living/supplemental-statement-afluria-tetra.html>
- Public Health ON: <https://www.publichealthontario.ca/-/media/documents/f/2020/fact-sheet-influenza-vaccine-2020-2021.pdf?la=en>
- Seqirus: https://www.seqirus.ca/-/media/seqirus-canada/docs-en/afluria-tetra-product-monograph--februray-19th-2021.pdf

**Adiuvato**

- Sequirus: <https://www.seqirus.it/notizie/approvato-dall-unione-europea-il-primo-vaccino-antinfluenzale-quadrivalente-adiuvato>
- <https://www.ema.europa.eu/en/documents/product-information/fluad-tetra-epar-product-information_it.pdf>
- <https://www.aulss2.veneto.it/documents/6017636/7745924/Fluad/10cad3c6-abc9-4ed6-991c-6bd0eddf6e63>

**Agriflu**

- FDA: <https://www.fda.gov/vaccines-blood-biologics/vaccines/agriflu> AND related: <https://www.drugs.com/history/agriflu.html>
- Government of Canada: <http://healthycanadians.gc.ca/recall-alert-rappel-avis/hc-sc/2012/15577a-eng.php>
- Government of Canada:<https://www.canada.ca/en/public-health/services/publications/healthy-living/canadian-immunization-guide-statement-seasonal-influenza-vaccine-2017-2018.html>
- Novartis:<https://www.novartis.ca/sites/www.novartis.ca/files/agriflu_patient_e_0.pdf>
- NPOA: <https://npao.org/health-canada-suspends-use-of-novartis-flu-vaccine-agriflu-fluad/>
- ON public drug programs: <http://www.health.gov.on.ca/en/pro/programs/drugs/opdp_eo/notices/exec_office_20171117.pdf>
- Peterborough Public Health: <https://www.peterboroughpublichealth.ca/wp-content/uploads/2012/05/121027-ALERT-Vaccine-Suspensions.pdf>
- Sequirus: <https://www.seqirus.ca/products/AGRIFLU>

**Flulaval / Fluviral**

- BCCDC: <http://www.bccdc.ca/resource-gallery/Documents/Guidelines%20and%20Forms/Guidelines%20and%20Manuals/Epid/CD%20Manual/Chapter%202%20-%20Imms/Part4/Influenza_FluLavalTetra.pdf> - updated
- FDA: <https://www.fda.gov/media/74537/download>
- GSK: <https://ca.gsk.com/media/590283/flulaval-tetra.pdf>
- GSK: <https://ca.gsk.com/media/1352353/fluviral.pdf>
- NLM: <https://dailymed.nlm.nih.gov/dailymed/drugInfo.cfm?setid=a9806027-8323-4abf-849f-46fb10984f13>

**Fluzone**

- BCCDC:<http://www.bccdc.ca/resource-gallery/Documents/Guidelines%20and%20Forms/Guidelines%20and%20Manuals/Immunization/Vaccine%20Info/FluzoneHD_QandA.pdf>
- Fluzone.ca: <https://www.fluzone.ca/>
- CDC: <https://www.cdc.gov/flu/prevent/qa_fluzone.htm>
- FDA: <https://www.fda.gov/vaccines-blood-biologics/vaccines/fluzone-fluzone-high-dose-and-fluzone-intradermal>
- Fluzone® High-Dose Influenza Vaccine with a Booster Is Associated with Low Rates of Influenza Infection in Patients with Plasma Cell Disorders - <http://www.bloodjournal.org/content/126/23/3058?sso-checked=true>
- Sanofi: <http://products.sanofi.ca/en/fluzone-qiv.pdf>
- Sanofiflu: <https://sanofiflu.com/fluzone-quadrivalent-influenza-vaccine.html>

**Fluad**

- CDC: <https://www.cdc.gov/flu/prevent/adjuvant.htm>
- CentreWatch: <https://www.centerwatch.com/clinical-trials/listings/213945/flu-fluad-vs-fluzone-hd/>
- Fluad.com: <https://flu.seqirus.com/Formulation/Fluad/p/FluadIn?seasonCategoryCode=Seqirus_InSeason>
- FDA: <https://www.fda.gov/vaccines-blood-biologics/vaccines/fluad>
- Government of Canada: Literature review update on the efficacy and effectiveness of high-dose (Fluzone® High-Dose) and MF59-adjuvanted (Fluad®) trivalent inactivated influenza vaccines in adults 65 years of age and older: <http://publications.gc.ca/site/eng/9.852907/publication.html>
- Government of Canada: <http://healthycanadians.gc.ca/recall-alert-rappel-avis/hc-sc/2012/15577a-eng.php>
- Gripguard (Fluad France)- <https://www.has-sante.fr/upload/docs/application/pdf/2010-06/gripguard_-_ct-6733.pdf>
- Novartis: file:///Users/jessiemcgowan/Downloads/Fluad%20Slide%20Deck_E_Sep2014.pdf
- Peel Public Health: <http://www.peelregion.ca/health/professionals/pdfs/2014/uiip-fluad-fact-sheet.pdf>
- Peterborough Public Health: <https://www.peterboroughpublichealth.ca/wp-content/uploads/2012/09/120925-FLU-Monograph-Fluad-2012.pdf>
- PubMED CENTRAL: <https://www.ncbi.nlm.nih.gov/pmc/articles/PMC3780956/>
- Seqirus: <https://www.seqirus.ca/products/FLUAD.htm>

**Fluarix**

- FDA: <https://www.fda.gov/vaccines-blood-biologics/vaccines/fluarix-quadrivalent>
- GSK: <https://www.gskflu.com/fluarix-quadrivalent/>
- <https://www.precisionvaccinations.com/vaccines/fluarix-quadrivalent-influenza-vaccine>

**Flucelvax**

- <https://flucelvax.ca>
- European medicine agency: <https://www.ema.europa.eu/en/medicines/human/EPAR/flucelvax-tetra>
- FDA: https://www.fda.gov/vaccines-blood-biologics/vaccines/flucelvax-quadrivalent
- Immunize.org: <https://www.immunize.org/askexperts/experts_inf.asp>
- Precision vaccinations: <https://www.precisionvaccinations.com/vaccines/flucelvax-quadrivalent-influenza-vaccine>
- Seqirus: https://www.seqirus.us/news/first-shipment-2021-22

**Flublok**

- CDC: Recombinant Influenza (Flu) Vaccine - <https://www.cdc.gov/flu/prevent/qa_flublok-vaccine.htm>
- FDA: <https://www.fda.gov/vaccines-blood-biologics/vaccines/flublok-quadrivalent>

<https://www.fluzone.com/flu-vaccines/flublok-quadrivalent>

- <https://www.sanofiflu.com/flublok-quadrivalent-influenza-vaccine/>
- Precision vaccinations: https://www.precisionvaccinations.com/vaccines/flublok-quadrivalent-influenza-vaccine

**Influvac**

- BGP Pharma: <https://www.mylan.ca/-/media/mylanca/documents/english/product-pdf/influvac-pm-2017-05-30.pdf>
- <https://pdf.hres.ca/dpd_pm/00045022.PDF>
- <https://www.scribd.com/document/347239541/Influvac-Insert-2017>

**Canada/USA**

AHRQ: <https://www.ahrq.gov/>

Alberta: <https://www.albertahealthservices.ca/assets/info/hp/cdc/if-hp-cdc-influenza-qiv-fluzone-bio-pg-07-265.pdf>

Blue Cross and Blue Shield Association's Technology Evaluation Center (TEC): <https://app.evidencestreet.com/>

British Columbia:

- <https://www.healthlinkbc.ca/health-feature/flu-season>
- <https://immunizebc.ca/>
- <http://www.bccdc.ca/resource-gallery/Documents/Guidelines%20and%20Forms/Guidelines%20and%20Manuals/Immunization/Vaccine%20Info/Archived_Quadrivalent_Influenza_Vaccine_Q_A_2015-16.pdf>
- [Health Quality Council of Alberta](http://www.hqca.ca/about) (HQCA). Completed Reviews
- <http://hqca.ca/studies-and-reviews/completed-reviews/>

Canadian Pharmacy Association/Immunize Canada : <https://www.pharmacists.ca/advocacy/issues/influenza/> - updated

CADTH: [www.CADTH.ca](http://www.CADTH.ca) –

- Pan-Canadian HTA collaborative: <https://www.cadth.ca/resources/hta-database-canadian-search-interface>
- High dose influenza vaccine for adults: <https://www.cadth.ca/high-dose-influenza-vaccine-adults-review-clinical-effectiveness-cost-effectiveness-and-guidelines>
- Influenza Vaccinations for the Prevention of Hospital Admissions: Clinical Effectiveness: <https://www.cadth.ca/influenza-vaccinations-prevention-hospital-admissions-clinical-effectiveness>
- Combined SARS-CoV-2 and Influenza Tests - <https://www.cadth.ca/combined-sars-cov-2-and-influenza-tests>
- Healthy Aging Interventions, Programs, and Initiatives: An Environmental Scan - https://www.cadth.ca/healthy-aging-interventions-programs-and-initiatives-environmental-scan
- Influenza Vaccination and Risk of Subsequent Non-Influenza Respiratory Viruses: Safety - <https://covid.cadth.ca/prevention/influenza-vaccination-and-risk-of-subsequent-non-influenza-respiratory-viruses-safety-2/>
- Point-of-Care Testing for Influenza - <https://www.cadth.ca/point-care-testing-influenza>

CDC (see updates above)

- <https://www.cdc.gov/flu/about/qa/vaxadmin.htm>
- MMWR Reports: <https://www.cdc.gov/mmwr/index.html>
- FastStats: <https://www.cdc.gov/nchs/fastats/>
- National Centre for Health Statistics: <https://www.cdc.gov/nchs/>

CIHI: <https://www.cihi.ca/en/covid-19-and-other-common-conditions-comparing-hospital-costs> - updated

CMA Infobase: <https://joulecma.ca/cpg/homepage>

CMS.gov: <https://www.cms.gov/medicare-coverage-database/indexes/technology-assessments-index.aspx?TAId=85&bc=AAAQAAAAAAAA&>

Department of Veteren Affairs (US): Updated

- <https://www.prevention.va.gov/flu/>

ECRI: <https://www.ecri.org>

FDA

- <https://www.fda.gov/vaccines-blood-biologics/vaccines/influenza-virus-vaccine-quadrivalent-types-and-types-b>
- <https://www.fda.gov/vaccines-blood-biologics/vaccines/influenza-virus-vaccine-trivalent-types-and-b>

Government of Canada:

- https://publications.gc.ca/site/eng/home.html (general)
- <https://www.canada.ca/content/dam/phac-aspc/documents/services/publications/vaccines-immunization/canadian-immunization-guide-statement-seasonal-influenza-vaccine-2021-2022/naci-2021-2022-statement.pdf>
- <https://www.canada.ca/en/public-health/services/diseases/flu-influenza/influenza-surveillance/weekly-influenza-reports.html>
- <https://www.canada.ca/en/public-health/services/canadian-immunization-guide.html>
- <https://www.canada.ca/en/public-health/services/diseases/flu-influenza.html>

ICER: <https://icer.org> - updated

Immunize Canada: <https://immunize.ca/>

IPAC: <https://ipac-canada.org/influenza-resources.php>

Johns Hopkins: <https://www.hopkinsguides.com/hopkins/view/Johns_Hopkins_ABX_Guide/540287/all/Influenza_vaccine>

PEI:

- <https://www.princeedwardisland.ca/en/information/health-and-wellness/universal-influenza-program-frequently-asked-questions-immunizers>

PHAC:

- <https://www.canada.ca/en/public-health.html>
- <https://www.canada.ca/en/public-health/services/reports-publications/disease-prevention-control-guidelines.html>
- Notifiable disease online: <https://dsol-smed.phac-aspc.gc.ca/notifiable/>
- Reports and publication: <https://www.canada.ca/en/public-health/services/reports-publications.html>
- Surveillance: <https://www.canada.ca/en/public-health/services/surveillance.html>

Manitoba:

- <https://www.gov.mb.ca/health/publichealth/cdc/div/manual/docs/msiipp.pdf>
- <http://mchp-appserv.cpe.umanitoba.ca/deliverablesList.html>

National guidelines clearinghouse: <https://www.ahrq.gov/gam/index.html>

New Brunswick: https://www.nbms.nb.ca/flu-shot/ - updated

Newfoundland:

- <https://www.gov.nl.ca/hcs/publichealth/cdc/flu-information/> - updated
- NLCAHR: <https://www.nlcahr.mun.ca/CHRSP/CompletedCHRSP.php>’

Nova Scotia: <https://novascotia.ca/flu/> - updated

Nunavit: <https://nrbhss.ca/en/departments/public-health/infectious-diseases/vaccination>

Ontario

- <http://www.health.gov.on.ca/en/pro/programs/publichealth/flu/uiip/default.aspx>
- <http://www.health.gov.on.ca/en/pro/programs/publichealth/flu/uiip/default09212017.aspx>
- HQO - <https://www.hqontario.ca/about-us>
- PATH HTA: https://www.path-hta.ca

Quebec:

- INESSS - <https://www.inesss.qc.ca/en/about-us/about-the-institut.html>
- <https://www.quebec.ca/en/health/advice-and-prevention/vaccination/flu-vaccine> - updated

Saskatchewan:

- Influenza reports: <https://www.saskatchewan.ca/government/government-structure/ministries/health/other-reports/influenza-reports>
- Rxfiles: <https://www.rxfiles.ca/rxfiles/modules/druginfoindex/druginfo.aspx>
- <https://www.saskhealthauthority.ca> Updated
- <https://publications.saskatchewan.ca/#/home>

Stats Can:

- <https://www150.statcan.gc.ca/n1/pub/82-624-x/2015001/article/14218-eng.htm>
- <https://www150.statcan.gc.ca/n1/pub/82-003-x/2018010/article/00003-eng.htm>

Vaccine Choice Canada: <https://vaccinechoicecanada.com/specific-vaccines/fluad-influenza-vaccine-for-seniors/>

Yukon: <http://www.hss.gov.yk.ca/2599.php>

**International**

WHO

- <https://www.who.int/health-topics/influenza-seasonal#tab=tab_1> - Updated

INAHTA: <http://www.inahta.org/publications/>

**General Grey**

- GreyNet International: <http://www.greylit.org>
- National Technical Information Service (NTIS): <http://www.ntis.gov>/

**Search engines**

- TRIP database: <http://www.tripdatabase.com/>
- Google: <https://www.google.ca/advanced_search>
- Google Scholar: <https://scholar.google.com/intl/en/scholar/about.html>

**Thesis**

- Center for Research Libraries Foreign Dissertation: <https://www.crl.edu/collections/topics/dissertations>
- DART-Europe E-theses Portal: <http://www.dart-europe.eu/basic-search.php>
- Electronic Theses Online Service (ETHOS) | British Library: <http://ethos.bl.uk/Home.do;jsessionid=D96E9CF245B0FE0199DDDB94FF4BD2A7>
- Open access dissertations: <https://oatd.org>
- Thesis Canada Portal: <http://www.bac-lac.gc.ca/eng/services/theses/Pages/theses-canada.aspx>

**Specific articles**

- Effectiveness of the MF59‐adjuvanted trivalent or quadrivalent seasonal influenza vaccine among adults 65 years of age or older, a systematic review and meta‐analysis: <https://www.ncbi.nlm.nih.gov/pmc/articles/PMC8542957/>
- Recombinant HA-based vaccine outperforms split and subunit vaccines in elicitation of influenza-specific CD4 T cells and CD4 T cell-dependent antibody responses in humans - https://www.nature.com/articles/s41541-020-00227-

**Clinical Trial Databases – full list - new**

- AMIS (German): <https://www.dimdi.de/dynamic/de/arzneimittel/arzneimittel-recherchieren/amis/>
- Australian New Zealand Clinical Trials Registry (ANZCTR): <http://www.anzctr.org.au>
- Be Part of Research (UK Clinical Trials): <https://sites.google.com/a/york.ac.uk/yhectrialsregisters/home/clinicaltrials/uk-clinical-trials>
- Brazilian Clinical Trials Registry: <http://www.ensaiosclinicos.gov.br/>
- China Drug Trials (in Chinese only): <http://www.chinadrugtrials.org.cn/eap/main>
- Chinese Clinical Trial Registry (ChiCTR): <http://www.chictr.org.cn/enIndex.aspx>
- ClinicalStudyDataRequest: <https://clinicalstudydatarequest.com>
- ClinicalTrials.gov: <https://www.clinicaltrials.gov>
- CentreWatch: <https://www.centerwatch.com/clinical-trials/listings/>
- Chinese Clinical Trial Registry (ChiCTR): <http://www.chictr.org.cn/abouten.aspx>
- Clinical Research Information Service (CRiS) (Korea): <https://cris.nih.go.kr/cris/en/>
- Cuban Registry of Clinical Trials: <http://registroclinico.sld.cu/en/home>
- DRKS - German Clinical Trials Register: <https://www.drks.de/drks_web/>
- Drugs@FDA: <https://www.accessdata.fda.gov/scripts/cder/daf/>
- EORTC Clinical Trials Database: <https://www.eortc.org/clinical-trials/>
- EORTC Clinical Trials tools: <https://www.eortc.org/tools/>
- EU Clinical Trials Register: <https://www.clinicaltrialsregister.eu/ctr-search/>
- EudraCT (European Union Drug Regulating Authorities Clinical Trials Database): <https://eudract.ema.europa.eu>
- Health Canada Clinical Trials Database: <https://health-products.canada.ca/ctdb-bdec/index-eng.jsp>
- Hong Kong UCT Register: <http://www.hkuctr.com>
- ISRCTN (UK): <http://www.isrctn.com/page/about>
- India (Clinical Trials Registry India): <http://ctri.nic.in/Clinicaltrials/pubview.php>
- Iranian Registry of Clinical Trials (IRCT): <https://www.irct.ir>
- Italian Medicines Agency - AIFA – Agenzia Italiaiana del Farmaco: <https://www.aifa.gov.it>
- OpenTrialsFDA: https://opentrials.net/opentrialsfda.1.html
- Philippine Health Research Registry:
  <http://registry.healthresearch.ph/>
- Japan Primary Registries Network (JPRN): <https://rctportal.niph.go.jp/en/>
- Netherland Trial Register: <https://www.trialregister.nl/trialreg/index.asp>
- Pan African Clinical Trials Registry: <https://pactr.samrc.ac.za/>
- PharmNet.Bund Clinical Trials: <https://www.pharmnet-bund.de/static/en/clinical-trials/index.html>
- Peruvian Registry of Clinical Trials: <https://www.ins.gob.pe/ensayosclinicos/>
- Research Registry: <https://www.researchregistry.com>
- South African National Clinical Trials Register: <http://www.sanctr.gov.za/>
- Spanish Registry of Clinical Studies (REec): <https://reec.aemps.es/reec/public/web.html>
- SNCTP (Swiss National Clinical Trials Portal): <https://www.kofam.ch/en/snctp-portal/searching-for-a-clinical-trial/>
- Sri Lanka Clinical Trials Registry: <https://slctr.lk/>
- Tanzania Clinical Trial Registry: <https://www.trialassure.com/resources/blog/registry-snapshot-tanzania-clinical-trials-registry-tzctr/>
- Thai Clinical Trials Registry: <http://www.clinicaltrials.in.th/>
- Trials tracker: <https://fdaaa.trialstracker.net>
- University hospital Medical Information Network (UMIN) Clinical Trials Registry (for Japan): <https://www.umin.ac.jp/ctr/>
- WHO: [WHO International Clinical Trials Registry Platform](http://www.who.int/ictrp/en/)
- YODA Yale University Open Data Access: <https://yoda.yale.edu>

### **Appendix 5: Level 1 screening guidelines**

**Question 1: Does this study examine a relevant influenza vaccine? [Mandatory]**

| **Response Options:** | □ Yes  □ No  □ Unclear |
| --- | --- |
| **Notes:** | **INCLUDE** the citation if the study mentions influenza vaccines licensed in Canada/United States for adults 60 years of age and older or similar, which may include but is not limited to the following relevant vaccines and associated acronyms:   - Afluria Tetra (Afluria Quadrivalent) - Agriflu/Agrippal - ccIIV3/IIV3-cc - Cell-Culture Inactivated Influenza Vaccine, Trivalent (Flucelvax) - ccIIV4/IIV4-cc - Cell-cultured inactivated quadrivalent vaccine - Fluad - Fluad Quadrivalent - Flucelvax Tetra (Flucelvax Quadrivalent) - Flulaval Tetra - Fluviral - Fluvirin - Fluzone High-Dose - Fluzone High-Dose Quadrivalent - Fluzone Quadrivalent - IIV3- Adj - Inactivated trivalent influenza vaccine - adjuvanted - IIV3 - Inactivated Influenza Vaccine, Trivalent - IIV4 – SD - Inactivated quadravalent influenza vaccine - standard dose - IIV4 –HD - Inactivated quadravalent influenza vaccine - high dose - IIV4-Adj - Inactivated quadravalent influenca vaccine - adjuvanted - IIV3-HD - Inactivated trivalent influenza vaccine - high dose - IIV3-SD - Inactivated trivalent influenza vaccine - standard dose - Influvac - Influvac Tetra - QIV - quadravalent inactivated vaccine - RIV3 - Recombinant Influenza Vaccine, Trivalent (Flublok) - TIV - Trivalent (Inactivated) Influenza Vaccine (replaced by the term IIV) - Recombinant influenza vaccine (RIV) - RIV4 – Recombinant Influenza Vaccine, Quadrivalent (Flublok quadrivalent) - Supemtek - 2009 H1N1 pandemic vaccine   **EXCLUDE** the citation if the following vaccines are mentioned:   - Live attenuated influenza vaccine (LAIV) - Virosomal, ASO3-adjuvanted, and intradermal vaccines - Monovalent influenza vaccines **UNLESS** it is the H1N1 pandemic vaccine from 2009 - Bivalent influenza vaccines - Experimental vaccines that have not yet moved to development or ever been approved - Any other influenza vaccine not licensed in Canada/United States (e.g. Vaxigrip) |

**Question 2: Does this study include adult patients aged 60 years and older?**

| **Response Options:** | □ Yes  □ No  □ Unclear |
| --- | --- |
| **Notes:** | **INCLUDE** the citation if:   - A range is given with only the minimum, e.g., ‘18 and over’ or it says only ‘adults’ - Only the mean or median age is given and it is >60 years   **EXCLUDE** the citation if:   - A range is given where the maximum age is less than 60 years, e.g., ’18-45 years’ |

**Question 3: Is this an eligible study design?**

| **Response Options:** | □ Yes  □ No  □ Unclear |
| --- | --- |
| **Notes:** | **INCLUDE** all primary research studies. Study designs include:   - Experimental studies such as   - Randomized controlled trials (RCTs), including patient randomized, cluster-randomized, and cross-over randomized trials   - Quasi-randomized clinical trial   **EXCLUDE** citations with the following study designs   - Cross-sectional - Case series - Case reports - Registry reports without comparator or denominator info - Qualitative studies - Opinion pieces (editorials, commentaries, letters) - Economic evaluations (cost-effectiveness, cost benefit analysis, etc.) - Clinical practice guideline - Consensus conference paper - Health technology assessment report - Non-RCTs (non-randomized controlled studies)* - Quasi-experimental studies* such as   - Interrupted time series   - Controlled before after studies - Observational studies*   - Cohort studies (both retrospective and prospective)   - Case control studies (traditional and test-negative designs) |
| *Study designs were excluded in a post-protocol decision | |

**Question 4: Is this a potentially relevant study with the following formats? [Flagging question]**

| **Response Options:** | □ Conference Abstract  □ Non-English article  □ Protocol  □ Systematic Review  □ Unclear |
| --- | --- |

### **Appendix 6: Level 2 screening guidelines**

**Question 1: Does this study examine a relevant influenza vaccine? [Mandatory]**

| **Response Options:** | □ Yes  □ No  □ Unclear |
| --- | --- |
| **Notes:** | **INCLUDE** the citation if the study mentions influenza vaccines licensed in Canada/United States for adults 65 years of age and older or similar, which may include but is not limited to the following relevant vaccines and associated acronyms:   - Afluria Tetra (Afluria Quadrivalent) - Agriflu/Agrippal - ccIIV3/IIV3-cc - Cell-Culture Inactivated Influenza Vaccine, Trivalent (Flucelvax) - ccIIV4/IIV4-cc - Cell-cultured inactivated quadrivalent vaccine - Fluad - Fluad Quadrivalent - Flucelvax Tetra (Flucelvax Quadrivalent) - Flulaval Tetra - Fluviral - Fluvirin - Fluzone High-Dose - Fluzone High-Dose Quadrivalent - Fluzone Quadrivalent - IIV3- Adj - Inactivated trivalent influenza vaccine - adjuvanted - IIV3 - Inactivated Influenza Vaccine, Trivalent - IIV4 – SD - Inactivated quadravalent influenza vaccine - standard dose - IIV4 –HD - Inactivated quadravalent influenza vaccine - high dose - IIV4-Adj - Inactivated quadravalent influenca vaccine - adjuvanted - IIV3-HD - Inactivated trivalent influenza vaccine - high dose - IIV3-SD - Inactivated trivalent influenza vaccine - standard dose - Influvac - Influvac Tetra - QIV - quadravalent inactivated vaccine - RIV3 - Recombinant Influenza Vaccine, Trivalent (Flublok) - TIV - Trivalent (Inactivated) Influenza Vaccine (replaced by the term IIV) - Recombinant influenza vaccine (RIV) - RIV4 – Recombinant Influenza Vaccine, Quadrivalent (Flublok quadrivalent) - Supemtek   **EXCLUDE** the citation if the following vaccines are mentioned:   - Live attenuated influenza vaccine (LAIV) - Virosomal, ASO3-adjuvanted, and intradermal vaccines - Monovalent influenza vaccines - Bivalent influenza vaccines - Experimental vaccines that have not yet moved to development or ever been approved - Any other influenza vaccine not licensed in Canada/United States (e.g. Vaxigrip) - Intradermal vaccines |

**Question 2: Does this study include adult patients aged 60 years and older?**

| **Response Options:** | □ Yes  □ No  □ Unclear |
| --- | --- |
| **Notes:** | **INCLUDE** the citation if:   - A range is given with only the minimum, e.g., ‘18 and over’ or it says only ‘adults’ - Only the mean or median age is given and it is >60 years   **EXCLUDE** the citation if:   - A range is given where the maximum age is less than 60 years, e.g., ’18-45 years’ |

**Question 3: Does this study include an eligible comparator group?**

| **Response Options:** | □ Yes  □ No  □ Unclear |
| --- | --- |
| **Notes:** | **INCLUDE** the citation if the comparator is:   - Any trivalent or quadrivalent influenza vaccines licensed in Canada/United States for adults ages 60 years and over - May consider comparisons of products similar to a Canadian or US-licensed vaccine in terms of dosage, formulation, and route of administration (e.g. Vaxigrip is similar to Fluzone) - Any vaccine for influenza or non-influenza control vaccines for any other condition (i.e., meningococcal or pneumococcal vaccines) - The same vaccine as the treatment arm but with a different dose and/or formulation - Placebo or no vaccine |

**Question 4: Is this an eligible study design?**

| **Response Options:** | □ Yes  □ No  □ Unclear |
| --- | --- |
| **Notes:** | **INCLUDE** all primary research studies. Study designs include:   - Experimental studies such as   - Randomized controlled trials (RCTs), including patient randomized, cluster-randomized, and cross-over randomized trials   - Quasi-randomized clinical trial   **EXCLUDE** citations with the following study designs   - Cross-sectional - Case series - Case reports - Registry reports without comparator or denominator info - Qualitative studies - Opinion pieces (editorials, commentaries, letters) - Economic evaluations (cost-effectiveness, cost benefit analysis, etc.) - Clinical practice guideline - Consensus conference paper - Health technology assessment report - Non-RCTs (non-randomized controlled studies)* - Quasi-experimental studies* such as - Interrupted time series - Controlled before after studies - Observational studies* - Cohort studies (both retrospective and prospective) - Case control studies (traditional and test-negative designs) |
| *Study designs were excluded in a post-protocol decision | |

**Question 5: Does this study include relevant outcomes?**

| **Response Options:** | □ Yes  □ No  □ Unclear |
| --- | --- |
| **Notes:** | Relevant outcomes include:  **Laboratory confirmed influenza** –Incidence (Newly diagnosed cases) of laboratory confirmed influenza with or without symptoms.  **Influenza-like-Illness (ILI)** - Incidence (Newly diagnosed cases) of influenza like illness diagnosed based on symptoms only or based on symptoms + laboratory confirmed.  **Hospitalization**- All causes of hospitalization, including but not limited to laboratory-confirmed influenza, influenza like illness, pneumonia etc.  **Mortality**- Deaths recorded due to influenza, influenza like illness, cardiovascular disease or all-cause mortality  **Adverse vascular events**- Adverse events include non-fatal myocardial infarction, stroke, cardiovascular death and heart failure |

**Question 6: Is this a potentially relevant study with the following formats? [Flagging question]**

*Only flag studies if the responses to Q1-Q4 are YES or UNCLEAR

| **Response Options:** | □ Conference Abstract  □ Non-English article  □ Protocol  □ Systematic Review  □ Unclear |
| --- | --- |

### **Appendix 7: Data abstraction form guidelines**

NOTES

Please enter not applicable (NA) or not reported (NR) as needed instead of leaving cells blank.

The form has been designed to accommodate six arm studies, for studies with 7 or more arms copy and paste additional columns as needed. Please ensure they are numbered properly.

**Tab 1. Study Characteristics**

|  | **STUDY CHARACTERISTICS** | |
| --- | --- | --- |
|  | **Excel column** | **Description** |
|  | RefID | [Will already be filled in] |
|  | Reviewer | [Will already be filled in] |
|  | Last name of first author | Enter the last name of the first author.  **Example:** Smith |
|  | Year of publication | Enter the year the study was published.  **Example:** 2015 |
|  | Publishing source name | Enter the journal name or name of the publishing source (if the journal name is not available). |
|  | Publication type | Select the publication type.   - Journal article - Report - Conference abstract - Results from trial registry - Dissertation/Thesis |
|  | Funding source name | Enter the source of funding support for the research.  **Example:** CIHR; Bayer. Enter ‘None’ for no funding source. |
|  | Funding source type | Select the category of the research sponsor type.   - Public - Industry - Public & Industry - NR - Unclear |
| Trial/Study name | | Enter the name of the trial or study.  **Example:** Flublok Study |
| Trial identifier | | Enter the clinicaltrials.gov trial number or any other trial identification number if applicable. If details of linked trials are provided please indicate this in the **Comment** column.  **Example:** NCT00007501, EudraCT Number: 2007-000744-28 |
| Study design* | | Select the study design from the dropdown.   - Randomized controlled trials (RCTs) - Cluster RCTs - Cross-over RCTs - Quasi-RCTs - Unclear   **Note:** Studies might not always explicitly state the design in which case select the best option based on the methods section. If unclear, please include any relevant information in the comment column at the end of the tab.  *****The following study designs were excluded in a post-protocol decision: Non-RCTs, Interrupted time series, Controlled before after, Prospective cohort, Retrospective cohort, Case control, Case test-negative, Nested case- control, Case-cohort |
| Country | | Enter the country of conduct.  If the trial is a multi-site trial, ensure all the countries are listed (separated by commas). If the country of conduct is not clear, use the country from the first author’s affiliation. If the study doesn’t mention the country but mentions a region, enter the region (e.g., Sub-Saharan Africa).  **Example:** USA, Canada, Netherlands |
| Single or multicenter | | From the dropdown, select whether the study is single center, multicenter, unclear or NR (not reported). |
| No. of centers in total | | Enter the total number of centers partaking in the study. |
|  | Influenza season | Enter the relevant influenza season(s) during which the study was conducted. If multiple seasons are reported, separate using a semicolon.  Specify northern or southern hemisphere  **Example: N** 2017/2018 or **S** 2018 |
|  | Companion report | Please select from the dropdown the following options:   - YES - NO - Unclear - NR |
|  | Industry conflict declared? | Please indicate if authors declare any industry conflict using the dropdown options.   - YES - NO - Unclear - NR   Example of declaration statement: *Employees may hold stock and/or stock options in the company.* |
|  | Conflict details | Copy-paste the description of the conflict. |
|  | Comment | Please enter any comments. |

**Tab 2. Participant Characteristics**

|  | **PARTICIPANT CHARACTERISTICS** | | |
| --- | --- | --- | --- |
|  | **Excel column** | | **Description** |
|  | RefID | | [Auto-populated based on previous entry] |
|  | Reviewer | | [Auto-populated based on previous entry] |
|  | Patient inclusion criteria | | Copy-paste the patient inclusion criteria for the study. |
|  | Patient exclusion criteria | | Copy-paste the patient exclusion criteria for the study. |
|  | Setting | | Copy and paste the description of the study setting.  **Example:** Long term care home, Family doctor, Flu clinic |
|  | Overall sample size | | Enter the total number of participants enrolled in the study for randomization. |
|  | % Female | | Enter in the overall percent of females in the study (in all treatment groups). If necessary, calculate using the following:  Calculation: [(#Tx1Females+#Tx2Females)/(Tx1Sample+Tx2Sample)]*100 |
|  | % Male | | Enter in the overall percent of males in the study (in all treatment groups).  If necessary, calculate using the following: |
|  | | Calculation: [(#Tx1Males+#Tx2Males)/(Tx1Sample+Tx2Sample)]*100 | |
| % Other | | Enter the overall percent of participants with other specified gender or no specified gender in the study (in all treatment groups). If multiple other genders are reported, separate using a semicolon.  If necessary, calculate using the following:  Calculation: [(#Tx1Other+#Tx2Other)/(Tx1Sample+Tx2Sample)]*100 | |
| Race | | Copy and paste the breakdown of participants’ race if provided. | |
| Age value | | Enter the age of the overall sample. If age is reported as a bracket (e.g., 65- 70 years), enter the relevant bracket and accompanying value with a colon in between (e.g., 65-75: 50%). If there are multiple brackets, separate using a semicolon (e.g., 65-75: 50%; 76-85: 50%). | |
| Age measure | | Select how age is reported for the overall sample:   - Mean - Median - Range - Bracket - Unclear - NR | |
| Age variance value | | Enter the value of the variance reported.  **Example:** SD: 14.3; Range: 59-85 | |
| Age variance type | | Select from the dropdown menu:   - Standard deviation (SD) - Standard error (SE) - Range - Interquartile range (IQR) - 95% CI - Unclear - NR | |
| Comorbidities | | Enter details of any comorbid conditions present in the entire study sample. If a comorbidity scale/index was used report the results. If comorbid conditions are presented per treatment group, enter NA here, and abstract per treatment group below.  **Example:** Heart disease (45%), COPD (13%), Cancer (8%); Charlson  Comorbidity Index (1.2) | |
| Vaccination history | | Copy and paste any information about the participants’ history of influenza vaccination for the entire sample. | |
| Frailty (%) | | Enter the percentage of participants who are described as frail for the entire sample. | |
| Frailty Definition | | Enter or copy and paste how frailty was defined for the entire sample. | |
| Frailty Scale | | Enter the name of the scale used to measure frailty for the entire sample. | |
| Frailty Score | | Enter the participants’ score on the frailty scale along with the variance for the entire sample. | |
| Tx1_Name | | Enter the name of the intervention allocated to the treatment arm. Be sure to include mention of type (monovalent, trivalent, tetravalent) and whether it is adjuvanted.  **Example:** Agriflu (tetravalent adjuvanted); Flublok (trivalent) | |
| Tx1_Comorbidities | | Enter details of any comorbid conditions present in the treatment group. If a comorbidity scale/index was used report the results.  **Example:** Heart disease (45%), COPD (13%), Cancer (8%); Charlson  Comorbidity Index (1.2) | |
| Tx1_Vaccination history | | Copy and paste any information about the participants’ history of influenza vaccination. | |
| Tx1_Frailty (%) | | Enter the percentage of participants who are described as frail. | |
| Tx1_Frailty Definition | | Enter or copy and paste how frailty was defined. | |
| Tx1_Frailty Scale | | Enter the name of the scale used to measure frailty. | |
| Tx1_Place of residence | | Copy and paste the breakdown of places of residence (e.g., rural, urban, suburban, long-term care, congregate setting etc). | |
| Tx1_Race | | Copy and paste the breakdown of races. | |
| Tx1_Occupation | | Copy and paste the breakdown of occupations. | |
| Tx1_% Female | | Enter the percent of females. | |
| Tx1_% Male | | Enter the percent of males. | |
| Tx1_% Other gender | | Enter the percent of other specified gender or no specified gender. If there are multiple genders reported, separate using a semicolon. | |
| Tx1_ Religion | | Copy and paste the breakdown of religious backgrounds. | |
| Tx1_Education | | Copy and paste the breakdown of educational backgrounds. | |
| Tx1_ Socioeconomic status | | Copy and paste the breakdown of participants’ socioeconomic status. | |
| Tx1_Social capital | | Copy and paste the breakdown of participants’ social capital categories. | |
| Tx1_Disability | | Copy and paste the breakdown of participants’ disability categories. | |
| Tx1_Features of relationships | | Copy and paste any information on the participants’ features of relationships (e.g., smoking parents, excluded from school). | |
| Tx1_Time dependent relationships | | Copy and paste any information on the participants’ time dependent relationships (e.g., leaving the hospital, respite care, other instances where a person may be temporarily at a disadvantage). | |
| Comment | | Enter any relevant comments for the patient characteristics that are not captured elsewhere. | |
| *Please add more columns if there are additional arms.* | | | |

**Tab 3. Intervention Characteristics**

|  | **INTERVENTION CHARACTERISTICS** | | |
| --- | --- | --- | --- |
|  | **Excel column** | | **Description** |
|  | RefID | | [Auto-populated based on previous entry] |
|  | Reviewer | | [Auto-populated based on previous entry] |
|  | Description of treatment arms | | Enter or copy and paste the description of the treatment arms in the study.  **Example:** Patients randomized into following groups: Aged 65-75 years to placebo, aged 65-75 years to Flublok |
|  | Tx1_name | | Enter the name of the intervention allocated to the treatment arm. Be sure to include mention of type (monovalent, trivalent, tetravalent) and whether it is adjuvanted.  NOTE:  For observational studies, Tx1_name would be those vaccinated or unvaccinated, and NOT cases or controls.  Cases/controls should be noted in the outcomes tab as appropriate (e.g., cases are LCI).  **Example:** Agriflu (tetravalent adjuvanted); Flublok (trivalent) |
|  |  | | From the dropdown indicate at what level vaccines were assigned: |
|  | Tx1_Vaccine | | - Clinic/facility |
|  | assignment | | - Patient |
|  |  | | - Other - UNCLEAR - NR |
| Tx1_platform | | Copy and paste any description of the platform used for virus growth (e.g. mammalian cell culture based, egg based, plant based, recombinant, etc.).  **Select from the drop-down:**   - Egg-based - Cell-based - Recombinant - NR | |
| Tx1_virus strains | | Enter the virus strains contained in the intervention for this treatment arm. If there are multiple years with different strains, enter the year first, colon and strains. Use a semicolon to separate years.  The virus strains are limited to the following: A/H1N1, A/H3N2, B/Yam, B/Vic | |
| Tx1_concentration | | Enter the concentration of Hemagglutinin per strain in the intervention for this treatment arm.  **Example:** 15 microgram/strain | |
| Tx1_adjuvant | | Enter the name and composition of the adjuvant used in the intervention for this treatment arm. If the vaccine is unadjuvanted enter ‘NA’.  **Example:** MF59 formulated with 9.75 mg squalene, 1.18 mg polysorbate 80, 1.18 mg sorbitan trioleate, 0.66 mg sodium citrate dihydrate, and 0.04 mg citricacid monohydrate | |
| Tx1_volume | | Enter the volume of the injection for this treatment arm.  **Example:** 0.5mL | |
| Tx1_administration | | Enter the route of administration and location of the intervention.  **Example:** Intramuscular, deltoid | |
| Tx1_frequency | | Enter number of doses administered.  **Example:** single dose | |
| Tx1_duration | | Enter the duration between doses if applicable. If a single dose was given enter ‘NA’. | |
| Tx1_sample size | | Enter the number of patients randomized to this study arm that received the intervention. | |
| Tx1_lost to follow up | | Enter the number of participants lost to follow up in this study arm | |
| Tx1_age value | | Enter the overall age of participants in this intervention arm. If age is reported as a bracket (e.g., 65-70 years), enter the relevant bracket and accompanying value with a colon in between (e.g., 65-75: 50%). If there are multiple brackets, separate using a semicolon (e.g., 65-75: 50%; 76-85: 50%)) | |
| Tx1_age measure | | Select how age is reported:   - Mean - Median - Range - Bracket - Unclear - NR | |
| Tx1_ age variance value | | Enter the value of the variance reported.  **Example:** SD: 14.3; Range: 59-85 | |
| Tx1_ age variance type | | Select from the dropdown menu:   - Standard deviation (SD) - Standard error (SE) - Range - Interquartile range (IQR) - 95% CI - NR | |
| Predominant circulating strain | | Enter the name(s) of the predominant circulating strain(s).  **Example:** A/Victoria/7/83 (H1N1), B/USSR/100/83 | |
| Other circulating strain | | Enter the name(s) of the other circulating strain(s). | |
| Mismatched strain? | | Enter whether the study reported a mismatch between the strains included in the vaccine and the circulating strains for the given time period (s). | |
| Comments | | Enter any relevant comments that are not captured elsewhere. | |
| *Please add more columns if there are additional arms.* | | | |

**Tab 4. Outcomes**

|  | **OUTCOMES** | | |
| --- | --- | --- | --- |
|  | **Excel column** | | **Description** |
|  | RefID | | [Auto-populated based on previous entry] |
|  | Reviewer | | [Auto-populated based on previous entry] |
|  | Incidence of ILI and Influenza | | |
|  | ILI without testing | | Enter or copy and paste how cases of Influenza-like-illness (ILI) without testing were defined in the study.  **Example:** ILI was defined as temperature of ≥37.2◦C or feverishness and at least two of the following symptoms: headache, myalgia, cough, or sore throat. |
|  | Acute respiratory illness | | Enter or copy and paste how cases of acute respiratory illness (ARI) were diagnosed and/or confirmed in the study. |
|  | Laboratory confirmed -ARI | | Enter or copy and paste how cases of laboratory confirmed-ARI were diagnosed and/or confirmed in the study. |
|  | Laboratory confirmed influenza | | Enter or copy and paste how laboratory confirmed influenza (LCI) infections were diagnosed and/or confirmed in the study. Make sure to report the methods used to confirm LCI.  **Example:** Respiratory illnesses detected by active and passive surveillance triggered the collection of nasopharyngeal swab for influenza confirmation…Laboratory confirmation of influenza in nasopharyngeal specimens was defined as a positive result on tissue culture and/or polymerase chain reaction (PCR). |
|  | Duration of follow-up | | Enter the duration of follow-up for the outcome measure using the unit reported in the paper.  **Example:** 3 years, 24 months, 365 days, etc. |
|  | Timing of assessment | | Enter the timing of outcome assessment using the unit reported in the paper. Also, indicate if timing of assessment was part of eligibility criteria.  **Example:** started 2 weeks post vaccination and throughout the winter |
|  | Tx1_name | | Name is pre-populated using entry from the intervention characteristics tab. |
|  | Tx1_sample size | | Enter the number of participants included in the outcome analysis (e.g., ITT, Per Protocol, or LOCF). |
|  | Tx1_lost to follow up | | Enter the number of patients lost to follow up in this study arm. |
|  | Tx1_reasons for lost to follow up | | Copy-paste the reasons participants were lost to follow up. |
|  | Tx1_method for dealing with missing data | | Enter the method authors used to deal with missing data (e.g., Intention- to-treat, last observation carried forward, per protocol). |
|  | Tx1_ILI cases | | Enter the number of or percentage of ILI cases reported (if reported as a proportion make sure you include the % symbol). |
| Tx1_ARI cases | | Enter the number of or percentage of acute respiratory infection (ARI) cases reported (if reported as a proportion make sure you include the % symbol). | |
| Tx1_LC-ARI cases | | Enter the number of or percentage of LC-ARI cases reported (if reported as a proportion make sure you include the % symbol). | |
| Tx1_LCI cases | | Enter the number of or percentage of confirmed influenza infections (if reported as a proportion make sure you include the % symbol). | |
| HOSPITALIZATIONS | | | |
| Tx1_Hospitalization for ILI | | Enter the number of or percentage of hospitalizations due to ILI (if reported as a proportion make sure you include the % symbol). | |
| Tx1_Hospitalization/ER visit for ILI | | Enter the number of or percentage of composite hospitalizations/emergency room (ER) visits due to ILI (if reported as a proportion make sure you include the % symbol). | |
| Tx1_Hospitalization for ARI | | Enter the number of or percentage of hospitalizations due to acute respiratory infections (ARI) (if reported as a proportion make sure you include the % symbol). | |
| Tx1_Hospitalization/ER for ARI | | Enter the number of or percentage of composite hospitalizations/ER visits due to ARI (if reported as a proportion make sure you include the % symbol). | |
| Tx1_Hospitalization for LCI | | Enter the number of or percentage of hospitalizations due to LCI (if reported as a proportion make sure you include the % symbol). | |
| Tx1_Hospitalization/ER for LCI | | Enter the number of or percentage of composite hospitalizations/ER visits due to LCI (if reported as a proportion make sure you include the % symbol). | |
| Tx1_Hospitalization for pneumonia due to ILI | | Enter the number of or percentage of hospitalizations due to ILI-related pneumonia (if reported as a proportion make sure you include the % symbol). | |
| Tx1_Hospitalization for pneumonia due to ARI | | Enter the number of or percentage of hospitalizations due to ARI-related pneumonia (if reported as a proportion make sure you include the % symbol). | |
| Tx1_Hospitalization for pneumonia due to LCI | | Enter the number of or percentage of hospitalizations due to LCI-related pneumonia (if reported as a proportion make sure you include the % symbol). | |
| Tx1_Hospitalization/ER for pneumonia | | Enter the number of or percentage of composite hospitalizations/ER visits due to ILI/ARI- or LCI-related pneumonia (if reported as a proportion make sure you include the % symbol). | |
| Tx1_Hospitalization/ Inpatient visit for any cause | | Enter the number of or percentage of hospitalizations due to any cause (if reported as a proportion make sure you include the % symbol). | |
| Tx1_Hospitalization/ER for any cause | | Enter the number of or percentage of composite hospitalizations/ER visits due to any cause (if reported as a proportion make sure you include the % symbol). | |
| Tx1_Inpatient/outpatient visit | | Enter the number of or percentage of composite inpatient/outpatient visits (if reported as a proportion make sure you include the % symbol). | |
| Tx1_Reasons for hospitalization | | Enter or copy and paste the reasons for hospitalization as reported in the study. | |
| Tx1_Outpatient visit | | Enter the number of or percentage of outpatient visits (if reported as a proportion make sure you include the % symbol). | |
| Tx1_ LCI-related health care interactions | | Enter the number of or percentage of LCI-related health care interactions (if reported as a proportion make sure you include the % symbol). | |
| ADVERSE VASCULAR EVENTS AND MORTALITY | | | |
| Definition of Vascular AEs | | Enter or copy and paste the types of vascular adverse events reported in the study (e.g., non-fatal myocardial infarction, stroke, cardiovascular death, heart failure). | |
| Tx1_# of Vascular adverse events | | Enter the count of adverse vascular events including non-fatal myocardial infarction, stroke, cardiovascular death, and heart failure. | |
| Tx1_# participants with Vascular adverse events | | Enter the count of participants with adverse vascular events including non- fatal myocardial infarction, stroke, cardiovascular death, and heart failure. | |
| Tx1_# of Hospitalizations due to cardiovascular events | | Enter the count of hospitalizations due to cardiovascular events. | |
| Tx1_# of participants hospitalized due to cardiovascular events | | Enter the count of participants hospitalized due to cardiovascular events. | |
| Tx1_# of LCI-related deaths | | Enter the count of LCI-related deaths. | |
| Tx1_ # of All-cause deaths | | Enter the count of all-cause deaths during the course of the study | |
| Vaccine Related? | | Enter or copy and paste whether any adverse vascular events or deaths were assessed as vaccine related. | |
| Comments | | Enter any relevant comments for the outcomes that are not captured elsewhere. | |
| *Please add more columns if there are additional arms.* | | | |

**Tab 5. Statistical Model Data**

Notes: This tab is only for non-randomized studies.

| **Excel column** | **Description** |
| --- | --- |
| RefID | Enter the refID of the study being abstracted |
| Reviewer | Enter your initials |
| Tx Comparison 1 | Enter the name of the treatment group/vaccine being compared. Be sure to include the type (monovalent, trivalent, tetravalent) and whether it is adjuvanted.  **Example**: Agriflu (tetravalent adjuvanted); Flublok (trivalent) |
| Tx Comparison 2 | Enter the name of the treatment group/vaccine being compared to “Tx Comparison 1”. Be sure to include the type (monovalent, trivalent, tetravalent) and whether it is adjuvanted. For additional comparisons,  insert data in rows below. Ensure to include the refid and reviewer name. |
| Outcome measure | Enter the relevant outcomes  **Example:** ILI, hospitalization |
| Measure of effect/association | Enter the name of the measure of effect/association (e.g., adjusted odds  ratio, adjusted rate ratio). If adjusted values are not reported, then enter the unadjusted effect size. |
| Adjusted? | Select if the measure of effect was adjusted or not from the dropdown menu:   - Yes - No - Unclear - Not reported |
| Measure of  effect/association value | Enter the value for the measure of effect/association. |
| 95% CI | Enter the 95% CI. |
| Analysis method | Enter in the analysis methods used.  **Example:** Multiple logistic/log binomial/Cox proportional hazards regression |
| Confounders / variables  controlled for in model | List any confounders or variables which were controlled for in the model.  Copy and paste any description of confounders/controls from the paper. |
| Number of participants | Enter the total number of participants included in the model. |
| Description of propensity scores or other advanced  analysis | Please provide a description of propensity scores or any other advanced analysis that was used. |
| Qualitative Evidence | Report any results collected narratively. |
| Comments | Enter any relevant comments that are not captured elsewhere. For  example, if matching was used instead of adjustment. |

### **Appendix 8: List of 41 included studies and 15 companion reports**

(‘*’ denotes the studies included in quantitative synthesis)

**Unique studies (n = 41)**

1. *Allsup S, Haycox A, Regan M, Gosney M. Is influenza vaccination cost effective for healthy people between ages 65 and 74 years? A randomised controlled trial. Vaccine. Dec 16 2004;23(5):639-645.
2. *Bart S, Cannon K, Herrington D, Mills R, Forleo-Neto E, Lindert K, Abdul Mateen A. Immunogenicity and safety of a cell culture-based quadrivalent influenza vaccine in adults: a phase III, double-blind, multicenter, randomized, non-inferiority study. Human vaccines & immunotherapeutics. 2016 Sep 1;12(9):2278-88.
3. *Belongia EA, Levine MZ, Olaiya O, Gross FL, King JP, Flannery B, McLean HQ. Clinical trial to assess immunogenicity of high-dose, adjuvanted, and recombinant influenza vaccines against cell-grown A (H3N2) viruses in adults 65 to 74 years, 2017–2018. Vaccine. 2020 Mar 30;38(15):3121-8.
4. *Beran J, Reynales H, Poder A, Charles YY, Pitisuttithum P, Yuan LL, Vermeulen W, Verhoeven C, Leav B, Zhang B, Sawlwin D. Prevention of influenza during mismatched seasons in older adults with an MF59-adjuvanted quadrivalent influenza vaccine: a randomised, controlled, multicentre, phase 3 efficacy study. The Lancet Infectious Diseases. 2021 Jul 1;21(7):1027-37.
5. *Chang LJ, Meng Y, Janosczyk H, Landolfi V, Talbot HK. Safety and immunogenicity of high-dose quadrivalent influenza vaccine in adults ≥65 years of age: A phase 3 randomized clinical trial. Vaccine. 2019;37(39):5825-34.
6. *Cowling BJ, Perera RAPM, Valkenburg SA, Leung NHL, Iuliano AD, Tam YH, et al. Comparative immunogenicity of several enhanced influenza vaccine options for older adults: A randomized, controlled trial. Clinical Infectious Diseases. 2020;71(7):1704-14.
7. *David W. Scheifele, Shelly A. McNeil, Brian J. Ward, Marc Dionne, Curtis Cooper, Brenda Coleman, Mark Loeb, Ethan Rubinstein, Janet McElhaney, Todd Hatchette, Yan Li, Emanuele Montomoli, Amy Schneeberg, Julie A. Bettinger, Scott A. Halperin & PHAC/CIHR Influenza Research Network (2013) Safety, immunogenicity, and tolerability of three influenza vaccines in older adults, Human Vaccines & Immunotherapeutics, 9:11, 2460-2473, DOI: 10.4161/ hv.25580
8. De Bruijn IA, Nauta J, Gerez L, Palache AM. The virosomal influenza vaccine Invivac®: Immunogenicity and tolerability compared to an adjuvanted influenza vaccine (Fluad®) in elderly subjects. Vaccine. 2006 Nov 10;24(44-46):6629-31.
9. Della Cioppa G, Nicolay U, Lindert K, Leroux-Roels G, Clement F, Castellino F, Galli C, Groth N, Levin Y, Del Giudice G. A dose-ranging study in older adults to compare the safety and immunogenicity profiles of MF59®-adjuvanted and non-adjuvanted seasonal influenza vaccines following intradermal and intramuscular administration. Human vaccines & immunotherapeutics. 2014 Jun 4;10(6):1701-10.
10. Della Cioppa G, Nicolay U, Lindert K, Leroux-Roels G, Clement F, Castellino F, Galli G, Groth N, Del Giudice G. Superior immunogenicity of seasonal influenza vaccines containing full dose of MF59® adjuvant: results from a dose-finding clinical trial in older adults. Human vaccines & immunotherapeutics. 2012 Feb 1;8(2):216-27.
11. *DiazGranados CA, Dunning AJ, Jordanov E, Landolfi V, Denis M, Talbot HK. High-dose trivalent influenza vaccine compared to standard dose vaccine in elderly adults: safety, immunogenicity and relative efficacy during the 2009–2010 season. Vaccine. 2013 Jan 30;31(6):861-6.
12. *DiazGranados CA, Dunning AJ, Kimmel M, Kirby D, Treanor J, Collins A, Pollak R, Christoff J, Earl J, Landolfi V, Martin E. Efficacy of high-dose versus standard-dose influenza vaccine in older adults. New England Journal of Medicine. 2014 Aug 14;371(7):635-45.
13. *Dunkle, Lisa M.; Izikson, Ruvim; Patriarca, Peter; Goldenthal, Karen L.; Muse, Derek; Callahan, Janice; Cox, Manon M. J.; P. S. C. Study Team. Efficacy of Recombinant Influenza Vaccine in Adults 50 Years of Age or Older. The New England journal of medicine. 2017; 376(25):2427-2436.
14. *Essink B, Fierro C, Rosen J, Figueroa AL, Zhang B, Verhoeven C, et al. Immunogenicity and safety of MF59-adjuvanted quadrivalent influenza vaccine versus standard and alternate B strain MF59-adjuvanted trivalent influenza vaccines in older adults. Vaccine. 2020;38(2):242-50.
15. Euctr, C. Z. Clinical trial to evaluate safety and immune response to flu vaccines in individuals 50 years of age and older.
16. *Falsey AR, Treanor JJ, Tornieporth N, Capellan J, Gorse GJ. Randomized, double-blind controlled phase 3 trial comparing the immunogenicity of high-dose and standard-dose influenza vaccine in adults 65 years of age and older. The Journal of infectious diseases. Jul 15 2009;200(2):172-180.
17. *Frey SE, Aplasca-De Los Reyes MR, Reynales H, Bermal NN, Nicolay U, Narasimhan V, Forleo-Neto E, Arora AK. Comparison of the safety and immunogenicity of an MF59®-adjuvanted with a non-adjuvanted seasonal influenza vaccine in elderly subjects. Vaccine. 2014 Sep 3;32(39):5027-34.
18. *Gravenstein S, Davidson HE, Han LF, Ogarek JA, Dahal R, Gozalo PL, Taljaard M, Mor V. Feasibility of a cluster-randomized influenza vaccination trial in US nursing homes: Lessons learned. Human vaccines & immunotherapeutics. 2018 Mar 4;14(3):736-43.
19. *Gravenstein S, Davidson HE, Taljaard M, Ogarek J, Gozalo P, Han L, Mor V. Comparative effectiveness of high-dose versus standard-dose influenza vaccination on numbers of US nursing home residents admitted to hospital: a cluster-randomised trial. The Lancet Respiratory Medicine. 2017 Sep 1;5(9):738-46.
20. Izikson R, Leffell DJ, Bock SA, Patriarca PA, Post P, Dunkle LM, Cox MM. Randomized comparison of the safety of Flublok® versus licensed inactivated influenza vaccine in healthy, medically stable adults≥ 50 years of age. Vaccine. 2015 Nov 27;33(48):6622-8.
21. *Keitel WA, Treanor JJ, El Sahly HM, Gilbert A, Meyer AL, Patriarca PA, Cox MM. Comparative immunogenicity of recombinant influenza hemagglutinin (rHA) and trivalent inactivated vaccine (TIV) among persons≥ 65 years old. Vaccine. 2009 Dec 11;28(2):379-85.
22. *Loeb N, Andrew MK, Loeb M, Kuchel GA, Haynes L, McElhaney JE, Verschoor CP. Frailty is associated with increased hemagglutination-inhibition titers in a 4-year randomized trial comparing standard-and high-dose influenza vaccination. InOpen Forum Infectious Diseases 2020 May (Vol. 7, No. 5, p. ofaa148). US: Oxford University Press.
23. McConeghy KW, Davidson HE, Canaday DH, Han L, Saade E, Mor V, Gravenstein S. Cluster-randomized trial of adjuvanted versus nonadjuvanted trivalent influenza vaccine in 823 US nursing homes. Clinical Infectious Diseases. 2021 Dec 1;73(11):e4237-43.
24. McLean HQ, Levine MZ, King JP, Flannery B, Belongia EA. Serologic response to sequential vaccination with enhanced influenza vaccines: Open label randomized trial among adults aged 65–74 years. Vaccine. 2021 Dec 3;39(49):7146-52.
25. *Nace DA, Lin CJ, Ross TM, Saracco S, Churilla RM, Zimmerman RK. Randomized, controlled trial of high-dose influenza vaccine among frail residents of long-term care facilities. The Journal of infectious diseases. 2015 Jun 15;211(12):1915-24.
26. Nct, Study to Assess the Immuno Response and the Safety Profile of a High-Dose Quadrivalent Influenza Vaccine (QIV-HD) Compared to a Standard-Dose Quadrivalent Influenza Vaccine (QIV-SD) in Japanese Adults 60 Years of Age and Older
27. Nct, T-cell And General Immune Response to Seasonal Influenza Vaccine (SLVP018) Year 3, 2011
28. Otten G, Matassa V, Ciarlet M, Leav B. A phase 1, randomized, observer blind, antigen and adjuvant dosage finding clinical trial to evaluate the safety and immunogenicity of an adjuvanted, trivalent subunit influenza vaccine in adults≥ 65 years of age. Vaccine. 2020 Jan 16;38(3):578-87.
29. *Pépin S, Donazzolo Y, Jambrecina A, Salamand C, Saville M. Safety and immunogenicity of a quadrivalent inactivated influenza vaccine in adults. Vaccine. 2013 Nov 12;31(47):5572-8.
30. *Rudenko LG, Arden NH, Grigorieva E, Naychin A, Rekstin A, Klimov AI, Donina S, Desheva J, Holman RC, DeGuzman A, Cox NJ. Immunogenicity and efficacy of Russian live attenuated and US inactivated influenza vaccines used alone and in combination in nursing home residents. Vaccine. 2000 Sep 15;19(2-3):308-18.
31. Sanchez L, Matsuoka O, Inoue S, Inoue T, Meng Y, Nakama T, Kato K, Pandey A, Chang LJ. Immunogenicity and safety of high-dose quadrivalent influenza vaccine in Japanese adults≥ 65 years of age: a randomized controlled clinical trial. Human Vaccines & Immunotherapeutics. 2020 Apr 2;16(4):858-66.
32. *Schmader KE, Liu CK, Harrington T, Rountree W, Auerbach H, Walter EB, Barnett ED, Schlaudecker EP, Todd CA, Poniewierski M, Staat MA. Safety, reactogenicity, and health-related quality of life after trivalent adjuvanted vs trivalent high-dose inactivated influenza vaccines in older adults: a randomized clinical trial. JAMA Network Open. 2021 Jan 4;4(1):e2031266-.
33. Szymczakiewicz-Multanowska A, Groth N, Bugarini R, Lattanzi M, Casula D, Hilbert A, Tsai T, Podda A. Safety and immunogenicity of a novel influenza subunit vaccine produced in mammalian cell culture. The Journal of infectious diseases. 2009 Sep 1;200(6):841-8.
34. Szymczakiewicz-Multanowska A, Lattanzi M, Izu A, Casula D, Sparacio M, Kovacs C, Groth N. Safety assessment and immunogenicity of a cell-culture-derived influenza vaccine in adults and elderly subjects over three successive influenza seasons. Human Vaccines & Immunotherapeutics. 2012 May 1;8(5):645-52.
35. Szymczakiewicz-Multanowska A, Lattanzi M, Izu A, Casula D, Sparacio M, Kovacs C, Groth N. Safety assessment and immunogenicity of a cell-culture-derived influenza vaccine in adults and elderly subjects over three successive influenza seasons. Human Vaccines & Immunotherapeutics. 2012 May 1;8(5):645-52.
36. *Teh BW, Leung VK, Mordant FL, Sullivan SG, Joyce T, Harrison SJ, Khvorov A, Barr IG, Subbarao K, Slavin MA, Worth LJ. A randomized trial of two 2-Dose influenza vaccination strategies for patients following autologous hematopoietic stem cell transplantation. Clinical Infectious Diseases. 2021 Dec 1;73(11):e4269-77.
37. Treanor J, Dumyati G, O'Brien D, et al. Evaluation of cold-adapted, reassortant influenza B virus vaccines in elderly and chronically ill adults. Journal of infectious diseases. 1994;169(2):402-407.
38. *Treanor JT, Albano FR, Sawlwin DC, et al. Immunogenicity and safety of a quadrivalent inactivated influenza vaccine compared with two trivalent inactivated influenza vaccines containing alternate B strains in adults: a phase 3, randomized noninferiority study. Vaccine. 2017;35(15):1856-1864.
39. *Tsang P, Gorse GJ, Strout CB, et al. Immunogenicity and safety of Fluzone intradermal and high-dose influenza vaccines in older adults >65 years of age: a randomized, controlled, phase II trial. Vaccine. 2014;32(21):2507-2517.
40. *Vardeny O, Kim K, Udell JA, Joseph J, Desai AS, Farkouh ME, Hegde SM, Hernandez AF, McGeer A, Talbot HK, Anand I. Effect of high-dose trivalent vs standard-dose quadrivalent influenza vaccine on mortality or cardiopulmonary hospitalization in patients with high-risk cardiovascular disease: a randomized clinical trial. JAMA. 2021 Jan 5;325(1):39-49.
41. *Wongsurakiat P, Maranetra KN, Wasi C, Kositanont U, Dejsomritrutai W, Charoenratanakul S. Acute respiratory illness in patients with COPD and the effectiveness of influenza vaccination: a randomized controlled study. Chest. 2004 Jun 1;125(6):2011-20.

**Companion reports (n = 15)**

1. DiazGranados, C. A.; Robertson, C. A.; Talbot, H. K.; Landolfi, V.; Dunning, A. J.; Greenberg, D. P. Prevention of serious events in adults 65 years of age or older: A comparison between high-dose and standard-dose inactivated influenza vaccines. Vaccine. 2015; 33(38):4988-4993.
2. Euctr, D. E. Safety and Immunogenicity of a Quadrivalent Influenza Vaccine Administered via the Intramuscular Route in Adult and Elderly Subjects. https://trialsearch. who. int/Trial2. aspx?TrialID=EUCTR2011-001976-21-DE. 2011.
3. Nct, A Phase 3, Randomized, Double-Blind, Controlled, Multicenter, Clinical Study to Evaluate Safety and Immunogenicity of an MF59-Adjuvanted Quadrivalent Subunit Influenza Vaccine in Comparison With an MF59-Adjuvanted Trivalent Subunit Influenza Vaccine and an MF59-Adjuvanted Trivalent Subunit Influenza Vaccine Containing the Alternate B Strain, in Adults Aged 65 Years and Above. 2017.
4. Nct, A Phase III, Observer-Blind, Randomized, Multi-Center Study to Evaluate Safety, Tolerability and Immunogenicity (in a Subset) Following a Single Intramuscular Dose of a Trivalent Subunit Influenza Vaccine Produced Either in Mammalian Cell Culture or in Embryonated Hen Eggs, in Healthy Adult and Elderly Subjects Who Received Either One or the Other Vaccine One Year Before in the V58P4 Study. 2006.
5. Nct, A Phase III, Observer-Blind, Randomized, Multi-Center Study to Evaluate Safety, Tolerability and Immunogenicity of a Single Intramuscular Dose of a Trivalent Subunit Influenza Vaccine Produced in Mammalian Cell Culture and of a Trivalent Subunit Influenza Vaccine Produced in Embryonated Hen Eggs, in Healthy Adult and Elderly Subjects. 2007.
6. Nct, A Phase III, Randomized, Controlled, Observer-Blind, Multicenter Study to Evaluate the Safety and Immunogenicity and the Consistency of Three Consecutive Lots of a MF59C.1 Adjuvanted Trivalent Subunit Influenza Vaccine in Elderly Subjects Aged 65 Years and Older. 2010.
7. Nct, A Phase III, Randomized, Observer-Blind, Controlled, Multicenter Clinical Study to Evaluate the Efficacy, Safety and Immunogenicity of an MF59-Adjuvanted Quadrivalent Influenza Vaccine Compared to Non-influenza Vaccine Comparator in Adults ≥ 65 Years of Age. 2015.
8. Nct, A Phase III, Single-Blind, Multi-Center, Extension Study to Evaluate Safety and Tolerability of a Trivalent Subunit Influenza Vaccine Produced Either in Mammalian Cell Culture or in Embryonated Hen Eggs in Adult and Elderly Subjects Who Participated in Study V58P4, With Subset Analyses of Immunogenicity and Evaluation of Concomitant Polysaccharide Pneumococcal Vaccine (Elderly). 2007.
9. Nct, Comparison of the Protective Efficacy of Flublok® Quadrivalent Versus Licensed Inactivated Influenza Vaccine (IIV4) in Healthy, Medically Stable Adults ≥50 Years of Age. 2014.
10. Nct, Efficacy Study of Fluzone® High-Dose Vaccine Compared With Fluzone® Vaccine In Elderly Adults. 2011.
11. Nct, Immunogenicity and Safety of Two Dosages of the Split, Inactivated, Trivalent Influenza Vaccine Administered by Intradermal Route in the Elderly Compared With Standard Fluzone® in Adults and Elderly Subjects. 2007.
12. Nct, Multi-Year Efficacy Study of Fluzone High-Dose Trivalent Vaccine Compared With Fluzone® Vaccine In Adults ≥ 65 Years of Age. 2009.
13. Nct, Phase III Lot Consistency, Immunogenicity and Safety Study of Three Lots of Fluzone High Dose Vaccine Compared With One Lot of Standard Fluzone® in Adults ≥ 65 Years of Age. 2006.
14. Nct, Safety and Immunogenicity of Adjuvanted Versus High-Dose Inactivated Influenza Vaccines in Older Adults. 2017.
15. Nct, Safety and Immunogenicity of High-Dose Quadrivalent Influenza Vaccine Administered by Intramuscular Route in Participants Aged 65 Years and Older. 2017.

### **Appendix 9: List of excluded studies post-data abstraction**

| **Reference (Author, Year)** | **Reason for Exclusion from Review (Post-Hoc Exclusions)** |
| --- | --- |
| **Chuaychoo, 2016** | Intradermal doses were excluded from the study as they lack authorization in Canada. The study was excluded as there was no other relevant comparator available. |
| **Levin, 2016** | Inflexal V is not authorized for use in Canada. Additionally, Intradermal doses were excluded from the study as they lack authorization in Canada. No other relevant comparator, thus study was excluded. Thus, both study arms were excluded. The study was excluded as there was no other relevant comparator available. |
| **Govaert, 1994** | QIV (Evans Medical Ltd, UK) does not appear to be a vaccine authorized for use in US/Canada. The study was excluded as there was no other relevant comparator available. |
| **Gravenstein, 1994** | TIV conjugated with diphtheria toxoid is not licensed in Canada. The study was excluded as there was no other relevant comparator available. |
| **Pregliasco, 2001** | Inflexal V (virosomal adjuvanted influenza vaccine) and Inflexal (whole virus vaccine) are not available in Canada. The study was excluded as there was no other relevant comparator available. |
| **Chi, 2010** | ID doses are excluded since not authorized in Canada. The study was excluded as there was no other relevant comparator available. |
| **Forrest, 2011** | IIV3 from Aventis Pasteur not authorized for use in Canada nor US. The study was excluded as there was no other relevant comparator available. |
| **McElhaney, 2013** | The study arm utilizing AS03 is excluded, since it is not licensed in Canada. The study was excluded as there was no other relevant comparator available. |
| **Greenberg, 2013** | The candidate or investigational vaccine, which has not yet obtained licensure, is not deemed eligible for inclusion in this review. The study was excluded as there was no other relevant comparator available. |
| **Rumke, 2013** | AS03 is not licensed for use in Canada. The study was excluded as there was no other relevant comparator available. |
| **Hung, 2013** | Intradermal doses were excluded from the study as they lack authorization in Canada. The study was excluded as there was no other relevant comparator available. |
| **Couch, 2014** | AS03-adjuvanted A(H1N1) pandemic vaccine (not investigational) was not licensed for use in Canada. The study was excluded as there was no other relevant comparator available. |
| **Greenberg, 2017** | The candidate or investigational vaccine, which has not yet obtained licensure, is not deemed eligible for inclusion in this review. The study was excluded as there was no other relevant comparator available. |
| **Choi, 2017** | Canada has not authorized SK Chemicals vaccines. Therefore, both treatment arms were excluded. Due to the lack of a relevant vaccine, the study was excluded. |
| **van de Witte, 2018** | The candidate or investigational vaccine, which has not yet obtained licensure, is not deemed eligible for inclusion in this review. The study was excluded as there was no other relevant comparator available. |
| **Young, 2019** | The comparison between bi-annual and annual vaccination was not deemed relevant. |
| **Ward, 2020** | Plant-derived influenza vaccine not authorized in Canada or US. The study was excluded as there was no other relevant comparator available. |
| **Izikson, 2022** | Pandemic vaccines are not considered for inclusion in this review. The study was excluded as there was no other relevant comparator available. |

### **Appendix 10: Study characteristics**

| Author, Year | Country | Publication details | Study design | Single or multicenter | Number of centers in total | Influenza season with Hemisphere | Intervention vs. comparator | Outcomes |
| --- | --- | --- | --- | --- | --- | --- | --- | --- |
| Studies included in quantitative and descriptive analysis (n = 26) | | | | | | | | |
| Rudenko, 2001^10^ | Russia | Journal Name: Elsevier Vaccine  Publication Type: Journal article  Funding Source (Type): NR  Conflict of Interest Declared: NR | RCT | Multicenter | 9 | N 1996/1997 | IIV3-SD vs. Placebo | LCI Cases |
| Wongsurakiat, 2004^11^ | Thailand | Journal Name: Chest Journal  Publication Type: Journal article  Funding Source (Type): Public  Conflict of Interest Declared: No | RCT | Single | 1 | N 1997/1998 | IIV3-SD vs. Placebo | Hospitalization for LCI, ILI Cases, Outpatient Visit, LCI Cases, All Cause Death |
| Allsup, 2004^12^ | UK | Journal Name:  Vaccine  Publication Type:  Journal article  Funding Source (Type): Public  Conflict of Interest Declared: NR | RCT | Multicenter | 20 | N 1999/2000 | IIV3-SD vs. Placebo | ILI Cases, Outpatient Visit, All Cause Death |
| Falsey, 2009^13^ | USA | Journal Name:  Journal of Infectious Diseases  Publication Type:  Journal article  Funding Source (Type): Industry  Conflict of Interest Declared: Yes | RCT | Multicenter | 30 | N 2006/2007 | IIV3-HD vs. IIV3-SD | Inpatient Hospitalization (any cause), All Cause Death, Number of Vascular Adverse Events |
| Keitel, 2010^14^ | USA | Journal Name: Vaccine  Publication Type: Journal article  Funding Source (Type): NR  Conflict of Interest Declared: NR | RCT | Multicenter | NR | N 2006/2007 | RIV3 vs. IIV3-SD | ILI Cases, LCI Cases, All Cause Death |
| Author, Year | **Country** | **Publication details** | **Study design** | **Single or multicenter** | **Number of centers in total** | **Influenza season with Hemisphere** | **Intervention vs. comparator** | **Outcomes** |
| DiazGranados 2013^15^ | USA | Journal Name: Vaccine  Publication Type: Journal article  Funding Source (Type): Industry  Conflict of Interest Declared: Yes | RCT | Multicenter | 99 | N 2009/2010 | IIV3-HD vs. IIV3-SD | Inpatient or Outpatient Hospital Visit, ER Visit for ILI, ILI Cases, Outpatient Visit, LCI Cases, All Cause Death, Number of Vascular Adverse Events |
| Scheifele, 2013^16^ | Canada | Journal Name:  Human Vaccines & Immunotherapeutics  Publication Type: Journal article  Funding Source (Type): Public and industry  Conflict of Interest Declared: Yes | RCT | Multicenter | 8 | N 2011/2012 | IIV3-Adj vs. IIV3-SD | Number of Participants with Vascular Adverse Events, All Cause Death, Number of Vascular Adverse Events |
| Pepin, 2013^17^ | France, Germany | Journal Name: Vaccine  Publication Type: Journal article  Funding Source (Type): Industry  Conflict of Interest Declared: Unclear | RCT | Multicenter | 18 | N 2011/2012 | IIV4-SD vs. IIV3-SD | All Cause Death |
| Tsang, 2014^18^ | USA | Journal Name: Vaccine  Publication Type: Journal article  Funding Source (Type): Industry  Conflict of Interest Declared: Yes | RCT | Multicenter | 31 | N 2007/2008 | IIV3-SD vs. IIV3-HD | Number of Vascular Adverse Events |
| Frey, 2014^19^ | Colombia, Panama, Philippines, USA | Journal Name: Vaccine  Publication Type: Journal article  Funding Source (Type): Industry  Conflict of Interest Declared: Yes | RCT | Multicenter | 38 | N 2010/2011 | IIV3-Adj vs. IIV3-SD | ILI Cases, All Cause Death, Number of Vascular Adverse Events |
| DiazGranados, 2014^20^ | USA, Canada | Journal Name: The New England Journal of Medicine  Publication Type: Journal article  Funding Source (Type): Industry  Conflict of Interest Declared: Yes | RCT | Multicenter | 126 | N 2011/2012 and N 2012/2013 | IIV3-HD vs. IIV3-SD | ARI Cases, ER Visit for LCI, Hospitalization for LCI, ER Visit for ILI, Hospitalization for ILI, Inpatient Hospitalization (any cause), ILI Cases, Hospitalization for ARI, Outpatient Visit, LCI Cases, All Cause Death, Number of Vascular Adverse Events |
| Author, Year | **Country** | **Publication details** | **Study design** | **Single or multicenter** | **Number of centers in total** | **Influenza season with Hemisphere** | **Intervention vs. comparator** | **Outcomes** |
| Nace, 2014^21^ | USA | Journal Name: The Journal of Infectious Diseases  Publication Type: Journal article  Funding Source (Type): Public and Industry  Conflict of Interest Declared: Yes | RCT | Multicenter | 15 | N 2011/2012 and N 2012/2013 | IIV3-HD vs. IIV3-SD | All Cause Death |
| Bart, 2016^22^ | USA | Journal Name:  Human Vaccines & Immunotherapeutics  Publication Type:  Journal article  Funding Source (Type): Industry  Conflict of Interest Declared: Yes | RCT | Multicenter | 40 | N 2013/2014 | IIV3-CC-SD vs. IIV4-CC-SD | All Cause Death |
| Gravenstein, 2017^23^ | USA | Journal Name: Lancet Respiratory Medicine  Publication Type: Journal article  Funding Source (Type): Industry  Conflict of Interest Declared: Yes | Cluster RCT | Multicenter | 823 | N 2013/2014 | IIV3-HD vs. IIV3-SD | ER Visit (any cause), ER Visit for Pneumonia (any cause), Hospitalization for ARI, All Cause Death |
| Dunkle, 2017^24^ | USA | Journal Name: The New England Journal of Medicine  Publication Type: Journal article  Funding Source (Type): Public  Conflict of Interest Declared: Yes | RCT | Multicenter | 40 | N 2014/2015 | RIV4 vs. IIV4-SD | Hospitalization for LCI, ILI Cases, LCI Cases, All Cause Death, Number of Vascular Adverse Events |
| Treanor, 2017^25^ | USA | Journal Name:  Vaccine  Publication Type:  Journal article  Funding Source (Type): Industry  Conflict of Interest Declared: Yes | RCT | Multicenter | 31 | N 2014/2015 | IIV4-SD vs. IIV3-SD | All Cause Death |
| Author, Year | **Country** | **Publication details** | **Study design** | **Single or multicenter** | **Number of centers in total** | **Influenza season with Hemisphere** | **Intervention vs. comparator** | **Outcomes** |
| Gravenstein, 2018^26^ | USA | Journal Name: Human Vaccines & Immunotherapeutics  Publication Type: Journal article  Funding Source (Type): Industry  Conflict of Interest Declared: Yes | Cluster RCT | Multicenter | 39 | N 2012/2013 | IIV3-HD vs. IIV3-SD | Inpatient Hospitalization (any cause), All Cause Death |
| Chang, 2019^27^ | USA | Journal Name:  Vaccine  Publication Type:  Journal article  Funding Source (Type): Industry  Conflict of Interest Declared: Yes | RCT | Multicenter | 35 | N 2017/2018 | IIV4-HD vs. IIV3-HD | All Cause Death, Number of Vascular Adverse Events |
| Loeb, 2020^28^ | USA, Canada | Journal Name:  Open Forum Infectious Diseases  Publication Type: Journal article  Funding Source (Type): Public  Conflict of Interest Declared: Yes | RCT | Multicenter | 2 | N 2014/2015, N 2015/2016, N 2016/2017 and N 2017/2018 | IIV3-SD vs. IIV3-HD | LCI Cases |
| Belongia, 2020^29^ | USA | Journal Name: Vaccine  Publication Type: Journal article  Funding Source (Type): Public  Conflict of Interest Declared: Yes | RCT | Single | 1 | N 2017/2018 | IIV3-HD vs. IIV3-Adj vs.RIV4 | LCI Cases |
| Cowling, 2020^30^ | China | Journal Name:  Clinical Infectious Diseases  Publication Type:  Journal article  Funding Source (Type): Public  Conflict of Interest Declared: Yes | RCT | Multicenter | NR | N 2017/2018 | IIV4-SD vs. IIV3-Adj vs. IIV3-HD vs. RIV4 | Inpatient Hospitalization (any cause) |
| Author, Year | **Country** | **Publication details** | **Study design** | **Single or multicenter** | **Number of centers in total** | **Influenza season with Hemisphere** | **Intervention vs. comparator** | **Outcomes** |
| Essink, 2020^31^ | USA | Journal Name:  Vaccine  Publication Type:  Journal article  Funding Source (Type): Industry  Conflict of Interest Declared: Yes | RCT | Multicenter | 20 | N 2017/2018 | IIV4-Adj vs. IIV3-Adj | All Cause Death, Number of Vascular Adverse Events |
| Beran, 2021^32^ | Bulgaria, Colombia, Czech Republic, Estonia, Latvia, Lithuania, Malaysia, Philippines, Poland, Romania, Thailand, and Turkey | Journal Name: Lancet Infectious Diseases  Publication Type: Journal article  Funding Source (Type): Industry  Conflict of Interest Declared: Yes | RCT | Multicenter | 89 | N 2016/2017 and  S 2017 | IIV4-Adj vs. Tdap | Number of Participants with Vascular Adverse Events, All Cause Death |
| Schmader, 2021^33^ | USA | Journal Name:  JAMA Network Open  Publication Type: Journal article  Funding Source (Type): Public  Conflict of Interest Declared: Yes | RCT | Multicenter | 3 | N 2017/2018 and N 2018/2019 | IIV3-Adj vs IIV3-HD | Number of Participants with Vascular Adverse Events, All Cause Death |
| Teh, 2021^34^ | Australia | Journal Name: Clinical Infectious Diseases  Publication Type: Journal article  Funding Source (Type): Public and Industry  Conflict of Interest Declared: Yes | RCT | Single | 1 | S 2019 | IIV3-HD vs. IIV4-SD | Hospitalization for LCI, ILI Cases, LCI Cases |
| Vardeny, 2021^35^ | USA, Canada | Journal Name: Journal of the American Medical Association  Publication Type: Journal article  Funding Source (Type): Public & Industry  Conflict of Interest Declared: Yes | RCT | Multicenter | 157 | N 2016/2017; N 2017/2018 and N 2018/2019 | IIV3-HD vs.IIV4-SD | All Cause Death |
| Author, Year | **Country** | **Publication details** | **Study design** | **Single or multicenter** | **Number of centers in total** | **Influenza season with Hemisphere** | **Intervention vs. comparator** | **Outcomes** |
| Studies included only in descriptive analysis (n= 15) | | | | | | | | |
| Treanor, 1994^36^ | USA | Journal Name:  The Journal of Infectious Diseases  Publication Type:  Journal article  Funding Source (Type): Public  Conflict of Interest Declared: No | RCT | Multicenter | 2 | NR | IIV3-SD + Placebo | ER Visit for Pneumonia (any cause) |
| de Bruijn, 2006^37^ | Netherlands | Journal Name: Vaccine  Publication Type: Journal article  Funding Source (Type):NR  Conflict of Interest Declared: NR | RCT | NR | NR | N 2004/2005 | IIV3-Adj vs. IIV3-SD | All Cause Death |
| Szymczakiewicz-Multanowska,  2009^38^ | Poland | Journal Name:  The Journal of Infectious Diseases  Publication Type: Journal article  Funding Source (Type): Industry  Conflict of Interest Declared: Yes | RCT | Multicenter | 5 | N 2004/2005 | IIV3-SD vs. IIV3-SD-CC | All Cause Death, Number of Vascular Adverse Events |
| Szymczakiewicz-Multanowska,  2012^39^ | Poland | Journal Name:  Human Vaccines & Immunotherapeutics  Publication Type: Journal article  Funding Source (Type): Industry  Conflict of Interest Declared: Yes | RCT | Multicenter | 5 | N 2005/2006 | IIV3-SD-CC vs. IIV3-SD | All Cause Death, Number of Vascular Adverse Events |
| Szymczakiewicz-Multanowska,  2012^40^ | Poland | Journal Name:  Human Vaccines & Immunotherapeutics  Publication Type: Journal article  Funding Source (Type): Industry  Conflict of Interest Declared: Yes | RCT | Multicenter | 5 | N 2007/2008 | IIV3-SD-CC vs. IIV3-SD | All Cause Death, Number of Vascular Adverse Events |
| Author, Year | **Country** | **Publication details** | **Study design** | **Single or multicenter** | **Number of centers in total** | **Influenza season with Hemisphere** | **Intervention vs. comparator** | **Outcomes** |
| Della Cioppa, 2012^41^ | Poland, Belgium, Germany | Journal Name:  Human Vaccines & Immunotherapeutics  Publication Type: Journal article  Funding Source (Type): Industry  Conflict of Interest Declared: Yes | RCT | Multicenter | 6 | N 2008/2009 | IIV3-Adj vs. IIV3-Adj vs. IIV3-Adj vs IIV3-Adj | All Cause Death |
| Della Cioppa, 2014^42^ | Germany, Poland, Belgium | Journal Name:  Human Vaccines & Immunotherapeutics  Publication Type:  Journal article  Funding Source (Type): Industry  Conflict of Interest Declared: Yes | RCT | Multicenter | 6 | N 2008/2009 | IIV3-SD vs. IIV3-Other vs. IIV3-Adj vs. IIV3-Other-Adj | All Cause Death |
| Izikson, 2015^43^ | USA | Journal Name:  Vaccine  Publication Type:  Journal article  Funding Source (Type): Public  Conflict of Interest Declared: Yes | RCT | Multicenter | NR | N 2012/2013 | RIV3 vs. IIV3-SD | All Cause Death |
| Novartis Vaccines and Diagnostics, 2016^44^ | Thailand, Philippines, South Africa, Czech Republic. | Journal Name:  EU Clinical Trials Register  Publication Type:  Results from trial registry  Funding Source (Type): Industry  Conflict of Interest Declared: NR | RCT | Multicenter | 24 | NR | IIV3-SD vs. IIV3-SD | All Cause Death, Number of Vascular Adverse Events |
| Trial registry, 2017^45^ | USA | Journal Name:  ClinicalTrials.gov  Publication Type:  Results from trial registry  Funding Source (Type): Public  Conflict of Interest Declared: NR | RCT | NR | NR | N 2011 | IIV3-SD vs. IIV3-HD | All Cause Death |
| Author, Year | **Country** | **Publication details** | **Study design** | **Single or multicenter** | **Number of centers in total** | **Influenza season with Hemisphere** | **Intervention vs. comparator** | **Outcomes** |
| Otten, 2020^46^ | Germany | Journal Name:  Vaccine  Publication Type: Journal article  Funding Source (Type): Industry  Conflict of Interest Declared: Yes | RCT | Single | 1 | N 2014/2015 | IIV3-Adj vs. IIV3-Adj vs. IIV3-Other-Adj vs.IIV3-Other-Adj – bilateral vs. IIV3-Adj-bilataeral + saline vs. IIV3-Adj- bilateral + saline vs. IIV3-Adj-bilateral | All Cause Death, Inpatient or Outpatient Hospital Visit |
| McConeghy, 2020^47^ | USA | Journal Name: Clinical Infectious Diseases  Publication Type: Journal article  Funding Source (Type): Industry  Conflict of Interest Declared: Yes | Cluster RCT | Multicenter | 823 | N 2016/2017 | IIV3-Adj vs. IIV3-SD | ER Visit (any cause), Hospitalization for ARI |
| Sanchez, 2020^48^ | Japan | Journal Name:  Human Vaccines & Immunotherapeutics  Publication Type:  Journal article  Funding Source (Type):  Industry  Conflict of Interest Declared: Yes | RCT | Multicenter | 2 | N 2017/2018 | IIV4-HD vs. IIV4-HD vs. IIV4-SD | All Cause Death |
| McLean, 2021^49^ | USA | Journal Name: Vaccine  Publication Type: Journal article  Funding Source (Type): Public  Conflict of Interest Declared: Yes | RCT | Single | 1 | N 2016/2017 and N 201  7/2018 | IIV3-HD (2015/2016, 2016/2017) vs. IIV3-Adj (2015/2016, 2016/2017) vs. IIV3-SD (2015/2016) + IIV3-HD (2016/2017) vs. IIV3-SD (2015/2016) + IIV3-Adj (2016/2017) | All Cause Death, LCI Cases |
| Author, Year | **Country** | **Publication details** | **Study design** | **Single or multicenter** | **Number of centers in total** | **Influenza season with Hemisphere** | **Intervention vs. comparator** | **Outcomes** |
| Trial registry, 2021^50^ | Japan | Journal Name:  ClinicalTrials.gov  Publication Type:  Results from trial registry  Funding Source (Type): Industry  Conflict of Interest Declared: Yes | RCT | Multicenter | 10 | N 2020/2021 | IIV4-SD vs. IIV4-HD | All Cause Death |
| Note: The term "-Other" in the vaccine name indicates that there are varying dosages available for different strains. As such, it is not categorized as standard dose nor high dose as per guidelines.  Abbreviations- NR: Not reported; N: Northern hemisphere; S: Southern hemisphere; IIV3: Trivalent inactivated influenza vaccine; IIV4: Quadrivalent inactivated influenza vaccine; Tdap: Tetanus, diphtheria, pertussis; Adj: Adjuvanted; SD: Standard dosage; HD: High dosage; CC: Cell-cultured; RIV: Recombinant influenza vaccine; USA: United States of America; UK: United Kingdom; RCT: Randomized controlled trial; ARI: Acute respiratory infection; LC-ARI: Laboratory-confirmed acute respiratory infection; ER: Emergency room; LCI: Laboratory-confirmed influenza; ILI: Influenza-like illness; AE: Adverse event | | | | | | | | |

### **Appendix 11: Participant characteristics**

| **Author,**  **Year** | **Setting** | **Overall sample size** | **% Female** | **%**  **Male** | **Race** | **Mean age, years (SD)** | **Most Common Comorbidities*** | **Influenza Vaccination history** | **Frailty (%)** | **Frailty**  **scale** | **Frailty score** |
| --- | --- | --- | --- | --- | --- | --- | --- | --- | --- | --- | --- |
| **Studies included in quantitative and descriptive analysis (n = 26)** | | | | | | | | | | | |
| **Rudenko, 2001^10^** | Nursing home | 319 | 69.3 | 30.7 | NR | 72.5 (5.5) | NR | NR | NR | NR | NR |
| **Wongsurakiat, 2004^11^** | Clinic | 125 | 5.6 | 94.4 | NR | 68.4 (7.5) | Chronic lung disease: 100%; Comorbid diseases (hypertension, coronary artery diseases, or diabetes): 32.8% | NR | NR | NR | NR |
| **Allsup, 2004^12^** | Community | 729 | 46.8 | 53.2 | NR | 68.9 (NR) | NR | NR | NR | NR | NR |
| **Falsey, 2009^13^** | Community | 3876 | 52.4 | 47.6 | White (92%), Hispanic (4.4%), Black (2.7%), Asian (0.4%), American Indian or Alaskan Native (0.08%), Native Hawaiian or Pacific Islander (0.05%), Other (0.3%) | 73.0 (5.6) | NR | 81.9% had receipt of vaccination in 2005 | NR | NR | NR |
| **Keitel, 2010^14^** | Community | 870 | 53.2 | 46.8 | White (98%); Black (1%); Hispanic (0.1%); Asian (0.2%); Other (0.6%). | 72.9 (6.1) | NR | 83.1% vaccinated in the previous season | NR | NR | NR |
| **DiazGranados, 2013^15^** | Clinic | 9172 | 53.7 | 46.3 | Asian  (0.7%); Black: (4.9%); Caucasian: (85%); Hispanic (8.7%); American Indian or Alaska native: (0.3%); Native Hawaiian or other Pacific Islander: (0.07%); Other: (0.25%) | 72.8 (6) | Cardiac disorders: 24.2%; Eye disorders: 50.3%; Immune system disorders: 44.3%; Metabolic disorders: 62.5%; Musculoskeletal disorders: 54.8%; Neoplasms: 21.3%; Psychiatric disorders: 24.2%; Vascular disorders: 65.1% | 88.7% had a previous history of seasonal influenza vaccination | NR | NR | NR |
| **Scheifele, 2013^16^** | Community or assisted living facility | 616 | 59.0 | 41.0 | White/  Caucasian (95%) | 73.8 (NR) | Well with co-morbidity: 19% | 86% vaccinated with IIV3 in the past 2 years | 4% | NR | NR |
| **Pepin, 2013^17^** | Community | 785 | 54 | 46 | NR | NR** | NR | NR | NR | NR | NR |
| **Tsang, 2014^18^** | NR | 1912 | 56.0 | 44.0 | Asian (0.6%), Black (2.5%), Caucasian (93.9%), Hispanic (2.4%), Other (1%) | NR (NR) | NR | 86.5% vaccinated in the past year | NR | NR | NR |
| **Frey, 2014^19^** | NR | 7104 | 65.1 | 34.9 | Asian (53%), Black (1%), Caucasian (28%), Hispanic (18%), Native American/Hawaii (<1%), Other (<1%) | 71.9 (5.3) | NR | NR | NR | NR | NR |
| **DiazGranados, 2014^20^** | Research centers | 31989 | 56.6 | 43.4 | White (94.6%), Asian (0.7%), Black (4%), Hispanic (6.1%), Other (0.6%) | 73.3 (5.8) | Diabetes: 22.6%; Hypothyroidism: 20.1%; | 73.6 | NR | NR | NR |
| **Nace, 2014^21^** | Long term care home | 205 | 68.0 | 32.0 | White, Non-Hispanic (99%) Other (1%) | 87.0 (6) | NR | NR | 100% | The standard ADL and IADL assessed functional status using 7 items each, scored from 0 to 2. The maximum score is 14 per scale, with higher scores indicating better function. Gait speed is measured through a timed 4-meter walk, with scores ≥1 m/s considered normal and scores ≤0.8 m/s indicating significant frailty and higher risk of mortality. | Gait speed, m/s, mean (SD) = 0.7 (0.3); ADL score, mean (SD) = 11.4 (3.7); IADL score, mean (SD) = 7.9 (4.2) |
| **Bart, 2016^22^** | Community | 1340 | NR | NR | NR | NR (NR) | NR | NR | NR | NR | NR |
| **Gravenstein, 2017**^23^ | Nursing home | 53008 | 72.2 | 27.8 | African American (14.8%) White (75.5%), Hispanic (5.1%) | 83.6 (8.5) | Heart failure: 20.5%; Stroke/cerebrov-ascular accident: 20.0%; Hypertension: 79%; Diabetes mellitus: 34.4%: Chronic lung disease: 20.2%; Dementias: 64.0% | NR | NR | NR | NR |
| **Dunkle, 2017^24^** | Outpatient centers | 9003 | 58.5 | 41.5 | Black (17.6%), White (80.3%), Other (2.1%); Hispanic ethnic group (4.9%), non-Hispanic (95.1%), Other (<1%) | 64.1 (NA) | Cardiovascular disease: 30.4%; Condition requiring statin lipid-lowering therapy: 27.7% | NR | NR | NR | NR |
| **Treanor, 2017^25^** | NR | 1743 | NR | NR | NR | NR (NR) | NR | NR | NR | NR | NR |
| **Gravenstein, 2018^26^** | Nursing Homes | 2957 | 74.6 | 25.3 | African American, (12.4%); White, (79.2%); Hispanic, (5.9%); Other (includes: American Indians, Alaskan Natives, Native Americans, Pacific Islanders, and Asians) (2.2%). | 84.0 (8.6) | Hypertension: 74.8%; Diabetes: 29.3%; Chronic lung disease: 19.7%; | NR | NR | NR | NR |
| **Chang, 2019^27^** | Community | 2670 | 54.9 | 39.9 | American Indian or Alaska Native (0.6%), Asian (0.7%), Black or African American (7.2%), White (90.7%), Multiple (0.4%), Not reported (0.4%) | 73.0 (5.6) | NR | 78.2% vaccinated in the past year | NR | NR | NR |
| **Loeb, 2020^28^** | Community | 612 | 67.0 | 33.0 | NR | 77.0 (7.5) | NR | 100% vaccinated in the previous influenza season | 9% | FI was determined based on 40 validated items related to influenza outcomes. Participants were classified as frail (FI > 0.21), pre-frail (0.1 < FI ≤ 0.21), or robust (FI ≤ 0.1) using established thresholds. | Robust: 50%;  Pre-frail: 40.8%;  Frail: 8.8%; Unknown: 0.3% |
| **Belongia, 2020^29^** | Clinic | 89 | 56.2 | 43.8 | White (100%) | 70.0 (2.5) | NR | NR | NR | NR | NR |
| **Cowling, 2020^30^** | Community | 1861 | 60.8 | 39.2 | NR | 71.6 (NR) | NR | 66.5% vaccinated in the 2016/2017 season | NR | NR | NR |
| **Essink, 2020^31^** | NR | 1778 | 56.6 | 43.4 | White (91.6%), Black (7%), Native Hawaiian or Pacific Islander (0.7%), Native American (0.1%), Other (0.2%) | 72.5 (5.5) | NR | 86.7% vaccinated in the past 5 years | NR | NR | NR |
| **Beran, 2021^32^** | Community | 6790 | 61.8 | 38.2 | American Indian or Alaska Native, (1.8%); Asian, (33.8%); Black or African American, (0.01%); White, (48.2%); Other(16.2%) | 71·9 (5.4) | NR | 29.6% vaccinated in the past 5 year | NR | NR | NR |
| **Schmader, 2021^33^** | Community | 757 | 55.5 | 44.5 | White (77.8%), Black (17%), Asian (1.2%), Other (4%) | 76.5 (4.6) | NR | NR | NR | NR | NR |
| **Teh, 2021^34^** | Community | 68 | 32.0 | 68.0 | NR | 60.0 (11.1) | Myeloma: 74%; Lymphoma: 22% | NR | NR | NR | NR |
| **Vardeny, 2021^35^** | NR | 5260 | 28.3 | 71.7 | White (78%), Black (14.9%), Asian (2.9%), First Nations/  American Indian (0.9%), Other (3%) | 65.5 (12.6) | Diabetes: 37.2%; Chronic kidney disease: 30.3%; Myocardial infarction: 14.2%; Ischemic stroke: 8.3%; Peripheral artery disease: 4.4% | NR | NR | NR | NR |
| **Studies included only in descriptive analysis (n= 15)** | | | | | | | | | | | |
| **Treanor, 1994^36^** | Community | 41 | 65.4 | 34.6 | NR | NR (NR) | Chronic pulmonary conditions: 18%; | NR | NR | NR | NR |
| **de Bruijn, 2006^37^** | Community | 386 | NR | NR | NR | 70.1 (NR) | NR | NR | NR | NR | NR |
| **Szymczakiewicz-Multanowska, 2009^38^** | Community | 1354 | 56.4 | 43.6 | NR | 69.0 (5.7) | NR | 59% had a previous history of seasonal influenza vaccination | NR | NR | NR |
| **Szymczakiewicz-Multanowska, 2012^39^** | Community | 1168 | 56.6 | 43.4 | Caucasian (100%) | 69.6 (5.7) | NR | NR | NR | NR | NR |
| **Szymczakiewicz-Multanowska, 2012^40^** | Community | 147 | 51.1 | 42.9 | Caucasian (100%) | 71.5 (5.5) | NR | NR | NR | NR | NR |
| **Della Cioppa, 2012^41^** | NR | 180 | 46.1 | 53.9 | NR | 69.2 (4.1) | NR | NR | NR | NR | NR |
| **Della Cioppa, 2014^42^** | NR | 270 | 53.0 | 47.0 | Caucasian (99.3%), Asian (0.7%) | 69.0 (3.8) | NR | NR | NR | NR | NR |
| **Izikson, 2015^43^** | NR | 1295 | 54.0 | 46.0 | White (91%), Black/African American: (8%), Asian (1%), American Indian/Alaska Native (<1%), Native Hawaiian/  Other Pacific Islander (<1%), Multiple (<1%) | 71.5 (5.1) | NR | NR | NR | NR | NR |
| **Novartis Vaccines and Diagnostics, 2016^44^** | NR | 2902 | 63.3 | 36.7 | NR | 64.2 (8.9) | NR | NR | NR | NR | NR |
| **Trial registry, 2017^45^** | NR | 10 | 60 | 40 | White/Not Hispanic or Latino (100%) | 70.1 (3.8) | NR | NR | NR | NR | NR |
| **Otten, 2020^46^** | Community | 196 | 46.7 | 53.3 | White, non-Hispanic (100%) | 70.5 (NR) | NR | NR | NR | NR | NR |
| **McConeghy, 2020^47^** | Nursing home | 50012 | 69.8 | 30.2 | White (72.3%), Black (16.7%), Hispanic (6.2%) | 78.7 (5.3) | Dementia: 61.9%; Congestive heart failure: 19.7%; Chronic respiratory illness: 21.8% | NR | NR | NR | NR |
| **Sanchez, 2020^48^** | Clinic | 175 | 45.7 | 54.3 | Asian (100%) | 70.3 (3.6) | NR | NR | NR | NR | NR |
| **McLean, 2021^49^** | Community | 152 | 53.3 | 46.7 | White (100%) | 69.8 (2.7) | NR | NR | NR | NR | NR |
| **Trial registry, 2021^50^** | Community | 2100 | 47.1 | 52.9 | Asian (100%) | 68.3 (4.9) | NR | NR | NR | NR | NR |
| **For brevity, the most common comorbidities are listed in the summary table.*  **** Enrollment in Pepin, 2013 was stratified by age at each site into adults 18–60 and >60 years of age. However, only data from the >60 age group was included in our review.  **Abbreviations-** LAIV: live attenuated influenza vaccine; FI: frailty index; ADL: activities of daily living; IADL: instrumental activities of daily living; m/s: meters/second; NR: not reported; SD: standard deviation | | | | | | | | | | | |

### **Appendix 12: Intervention characteristics**

| **Author, Year** | **Setting** | **Tx** | **Vaccine Type** | **Platform** | **Dosage of HA/ strain** | **Adjuvant** | **Administration** | **Frequency** |
| --- | --- | --- | --- | --- | --- | --- | --- | --- |
| **Studies included in quantitative and descriptive analysis (n = 26)** | | | | | | | | |
| **Rudenko, 2001^10^** | Nursing home | Tx1 | IIV3-SD | Egg-based | 15 µg/ strain | NA | Intramuscular | Single dose |
|  |  | Tx2 | Placebo | NA | NA | NA | Intramuscular; intranasal | Two doses |
| **Wongsurakiat, 2004^11^** | Clinic | Tx1 | IIV3-SD | NR | 15 µg/ strain | NA | Intramuscular | Two doses |
|  |  | Tx2 | Placebo | NA | NA | NA | Intramuscular | Two doses |
| **Allsup, 2004^12^** | Community | Tx1 | IIV3-SD | NR | 15 µg/ strain | NA | Intramuscular | Single dose |
|  |  | Tx2 | Placebo | NA | NA | NA | Intramuscular | NA |
| **Falsey, 2009^13^** | Community | Tx1 | IIV3-HD | Egg-based | 60 µg/ strain | NA | Intramuscular | Single dose |
|  |  | Tx2 | IIV3-SD | Egg-based | 15 µg/ strain | NA | Intramuscular | Single dose |
| **Keitel, 2010^14^** | Community | Tx1 | RIV3 | Egg-based | 45 µg/ strain | NA | Intramuscular | Single dose |
|  |  | Tx2 | IIV3-SD | Egg-based | 15 µg/ strain | NA | Intramuscular | Single dose |
| **DiazGranados, 2013^15^** | Clinic | Tx1 | IIV3-HD | Egg-based | 60 µg/ strain | NA | Intramuscular | Single dose |
|  |  | Tx2 | IIV3-sD | Egg-based | 15 µg/ strain | NA | Intramuscular | Single dose |
| **Scheifele, 2013^16^** | Community or assisted living facility | Tx1 | IIV3-Adj | Egg-based | 15 µg/ strain | MF59 | Intramuscular | Single dose |
|  |  | Tx2 | IIV3-SD | Egg-based | 15 µg/ strain | NA | Intramuscular | Single dose |
| **Pepin, 2013^17^** | Community | Tx1 | IIV4-SD | NR | 15 µg/ strain | NA | Intramuscular injection into deltoid muscle or deep SC tissue | Single dose |
|  |  | Tx2 | IIV3-SD | NR | 15 µg/ strain | NA | Intramuscular injection into deltoid muscle or SC tissue | Single dose |
| **Tsang, 2014^18^** | NR | Tx1 | IIV3-SD | NR | 15 µg/ strain | NA | Intramuscular | Single dose |
|  |  | Tx2 | IIV3-HD | NR | 60 µg/ strain | NA | Intramuscular | Single dose |
| **Frey, 2014^19^** | NR | Tx1 | IIV3-Adj | Egg-based | 15 µg/ strain | MF59 formulated with 9.75 mg squalene, 1.18 mg polysorbate 80, 1.18 mg sorbitan trioleate, 0.66 mg sodium citrate dihydrate, and 0.04 mg citric acid monohydrate. | Intramuscular | Single dose |
|  |  | Tx2 | IIV3-SD | Egg-based | 15 µg/ strain | NA | Intramuscular | Single dose |
| **DiazGranados, 2014^20^** | Research centers | Tx1 | IIV3-HD | Egg-based | 60 µg/ strain | NA | Intramuscular | Single dose |
|  |  | Tx2 | IIV3-SD | Egg-based | 15 µg/ strain | NA | Intramuscular | Single dose |
| **Nace, 2014^21^** | Long term care home | Tx1 | IIV3-HD | NR | 60 µg/ strain | NA | Intramuscular | Single dose |
|  |  | Tx2 | IIV3-SD | Egg-based | 15 µg/ strain | NA | Intramuscular | Single dose |
| **Bart, 2016^22^** | Community | Tx1 | IIV3-CC-SD | Cell-based | 15 µg/ strain | NA | Intramuscular | Single dose |
|  |  | Tx2 | IIV4-CC-SD | Cell-based | 15 µg/ strain | NA | Intramuscular | Single dose |
| **Gravenstein, 2017^23^** | Nursing home | Tx1 | IIV3-HD | NR | 60 µg/ strain | NA | NR | Single dose |
|  |  | Tx2 | IIV3-SD | NR | 15 µg/ strain | NA | NR | Single dose |
| **Dunkle, 2017^24^** | Outpatient centers | Tx1 | RIV4 | Recombinant | 45 µg/ strain | NA | Intramuscular | Single dose |
|  |  | Tx2 | IIV4-SD | Egg-based | 15 µg/ strain | NA | Intramuscular | Single dose |
| **Treanor, 2017^25^** | NR | Tx1 | IIV4-SD | Egg-based | 15 µg/ strain | NA | Intramuscular | Single dose |
|  |  | Tx2 | IIV3-SD | Egg-based | 15 µg/ strain | NA | Intramuscular | Single dose |
| **Gravenstein, 2018^26^** | Nursing homes | Tx1 | IIV3-HD | NR | 60 µg/ strain | NA | NR | NR |
|  |  | Tx2 | IIV3-SD | NR | 15 µg/ strain | NA | NR | NR |
| **Chang, 2019^27^** | Community | Tx1 | IIV4-HD | NR | 60 µg/ strain | NA | Intramuscular | Single dose |
|  |  | Tx2 | IIV3-HD | NR | 60 µg/ strain | NA | Intramuscular | Single dose |
| **Loeb, 2020^28^** | Community | Tx1 | IIV3-SD | NR | 15 µg/ strain | NA | NR | NR |
|  |  | Tx2 | IIV3-HD | NR | 60 µg/ strain | NA | NR | NR |
| **Belongia, 2020^29^** | Clinic | Tx1 | IIV3-HD | Egg-based | 60 µg/ strain | NA | Intramuscular | Single dose |
|  |  | Tx2 | IIV3-Adj | Egg-based | 15 µg/ strain | MF59 | Intramuscular | Single dose |
|  |  | Tx3 | RIV4 | Recombinant | 45 µg/ strain | NA | Intramuscular | Single dose |
| **Cowling, 2020^30^** | Community | Tx1 | IIV4-SD | NR | 15 µg/ strain | NA | Intramuscular | Single dose |
|  |  | Tx2 | IIV3-Adj | NR | 15 µg/ strain | MF59, a squalene-based emulsion | Intramuscular | Single dose |
|  |  | Tx3 | IIV3-HD | NR | 60 µg/ strain | NA | Intramuscular | Single dose |
|  |  | Tx4 | RIV4 | Recombinant | 45 µg/ strain | NA | Intramuscular | Single dose |
| **Essink, 2020^31^** | NR | Tx1 | IIV4-Adj | Egg-based | 15 µg/ strain | MF59, squalene based | Intramuscular | Single dose |
|  |  | Tx2 | IIV3-Adj | Egg-based | 15 µg/ strain | MF59, squalene based | Intramuscular | Single dose |
| **Beran, 2021^32^** | Community | Tx1 | IIV4-Adj | Egg-based | 15 µg/ strain | MF59 | Intramuscular | Single dose |
|  |  | Tx2 | Tdap | NA | NA | NA | Intramuscular | Single dose |
| **Schmader, 2021^33^** | Community | Tx1 | IIV3-Adj | Egg-based | 15 µg/ strain | MF59 squalane adjuvant | Intramuscular | Single dose |
|  |  | Tx2 | IIV3-HD | Egg-based | 60 µg/ strain | NA | Intramuscular | Single dose |
| **Teh, 2021^34^** | Community | Tx1 | IIV3-HD | Egg-based | 60 µg/ strain | NA | NR | Single dose |
|  |  | Tx2 | IIV4-SD | Egg-based | 15 µg/ strain | NA | NR | Single dose |
| **Vardeny, 2021^35^** | NR | Tx1 | IIV3-HD | Egg-based | 60 µg/ strain | NA | Intramuscular | Single dose |
|  |  | Tx2 | IIV4-SD | Egg-based | 15 µg/ strain | NA | Intramuscular | Single dose |
| **Studies included only in descriptive analysis (n= 15)** | | | | | | | | |
| **Treanor, 1994^36^** | Community | Tx1 | IIV3-SD | NR | 15 µg/ strain | NA | Intramuscular | Single dose |
|  |  | Tx2Tx2 | Placebo | NA | NA | NA | Intramuscular, intranasal | Single dose |
| **de Bruijn, 2006^37^** | Community | Tx1 | IIV3-Adj | NR | 15 µg/ strain | Yes - but adjuvant is not described in this report | Intramuscular | Single dose |
|  |  | Tx2 | IIV3-SD | NR | 15 µg/ strain | NA | Intramuscular | Single dose |
| **Szymczakiewicz-Multanowska, 2009^38^** | Community | Tx1 | IIV3-SD | Egg-based | 15 µg/ strain | NA | Intramuscular | Single dose |
|  |  | Tx2 | IIV3-SD-CC | Cell-based | 15 µg/ strain | NA | Intramuscular | Single dose |
| **Szymczakiewicz-Multanowska, 2012 (a)^39^** | Community | Tx1 | IIV3-SD-CC | Cell-based | 15 µg/ strain | NA | Intramuscular | Single dose |
|  |  | Tx2 | IIV3-SD | Egg-based | 15 µg/ strain | NA | Intramuscular | Single dose |
| **Szymczakiewicz-Multanowska, 2012 (b)^40^** | Community | Tx1 | IIV3-SD-CC | Cell-based | 15 µg/ strain | NA | Intramuscular | Single dose |
|  |  | Tx2 | IIV3-SD | Egg-based | 15 µg/ strain | NA | Intramuscular | Single dose |
| **Della Cioppa, 2012^41^** | NR | Tx1 | IIV3-Adj | NR | 15 µg/ strain | MF59 (25%) | Intramuscular | Single dose |
|  |  | Tx2 | IIV3-Other-Adj | NR | 15 µg/ A/H1N1 strain; 30 µg/ H3N2 strain; 15 µg/ B strain (total HA is 60µg) | MF59 (25%) | Intramuscular | Single dose |
|  |  | Tx3 | IIV3-Adj | NR | 15 µg/ strain | MF59 (50%) | Intramuscular | Single dose |
|  |  | Tx4 | IIV3-Other-Adj | NR | 15 µg A/H1N1 strain; 30 µg/ H3N2 strain; 15 µg HA for B strain (total HA is 60µg) | MF59 (50%) | Intramuscular | Single dose |
| **Della Cioppa, 2014^42^** | NR | Tx1 | IIV3-SD | NR | 15 µg/ strain | NA | Intramuscular | Single dose |
|  |  | Tx2 | IIV3-Other | NR | 15 µg/ A/H1N1 strain; 30 µg/ H3N2 strain; 15 µg/ B strain (total HA is 60 µg) | NA | Intramuscular | Single dose |
|  |  | Tx3 | IIV3-Adj | NR | 15 µg/ strain | MF59 - A standard/full dose (100%) | Intramuscular | Single dose |
|  |  | Tx4 | IIV3-Other-Adj | NR | 15 µg/ A/H1N1 strain; 30 µg/ H3N2 strain; 15 µg/ for B strain (total HA is 60 µg) | MF59 - A standard/full dose (100%) | Intramuscular | Single dose |
| **Izikson, 2015^43^** | NR | Tx1 | RIV3 | Recombinant | 45 µg/ strain | NA | Intramuscular | Single dose |
|  |  | Tx2 | IIV3-SD | Egg-based | 15 µg/ strain | NA | Intramuscular | Single dose |
| **Novartis Vaccines and Diagnostics, 2016^44^** | NR | Tx2Tx2 | IIV4-CC-SD | Cell-based | 15 µg/ strain | NA | Intramuscular | Single dose |
|  |  | Tx2 | IIV3-SD | Egg-based | 15 µg/ strain | NA | Intramuscular | Single dose |
| **Trial registry, 2017^45^** | NR | Tx1 | IIV3-SD | Egg-based | 15 µg/ strain | NA | Intramuscular | Single dose |
|  |  | Tx2 | IIV3-HD | Egg-based | 60 µg/ strain | NA | Intramuscular | Single dose |
| **Otten, 2020^46^** | Community | Tx1 | IIV3-Adj | NR | 15 µg/ strain | 9.75 µg MF59 | Intramuscular | Single dose |
|  |  | Tx2 | IIV3-Adj | NR | 15 µg/ strain | 19.5 µg MF59 | Intramuscular | Single dose |
|  |  | Tx3 | IIV3-Other-Adj | NR | 30 µg/ strain | 9.75 µg MF59 | Intramuscular | Single dose |
|  |  | Tx4 | IIV3-Other-Adj – bilateral | NR | 30 µg/ strain | 19.5 µg MF59 | Two bilateral intramuscular injections in left (active vaccine) and right (saline) deltoid | Single dose |
|  |  | Tx5 | IIV3-Adj-bilataeral + saline | NR | 15 µg/ strain | 9.75 µg MF59 | Two bilateral intramuscular injections in left (active vaccine) and right (saline) deltoid | Single dose |
|  |  | Tx6 | IIV3-Adj- bilateral + saline | NR | 15 µg/ strain | 29.25 µg MF59 | Two bilateral intramuscular injections in left (active vaccine) and right (saline) deltoid | Single dose |
|  |  | Tx7 | IIV3-Adj-bilateral | NR | 15 µg/ strain | 9.75 µg MF59/ arm | Two bilateral intramuscular injections in left (active vaccine) and right (saline) deltoid | Single dose |
| **McConeghy, 2020^47^** | Nursing home | Tx1 | IIV3-Adj | NR | 15 µg/ strain | MF59 | NR | Single dose |
|  |  | Tx2 | IIV3-SD | Egg-based | 15 µg/ strain | NA | NR | Single dose |
| **Sanchez, 2020^48^** | Clinic | Tx1 | IIV4-HD | NR | 60 µg/ strain | NA | Intramuscular | Single dose |
|  |  | Tx2 | IIV4-HD | NR | 60 µg/ strain | NA | Subcutaneous | NR |
|  |  | Tx3 | IIV4-SD | NR | 15 µg/ strain | NA | Subcutaneous | Single dose |
| **McLean, 2021^49^** | Community | Tx1 | IIV3-HD (2015/2016, 2016/2017) | Egg-based | Year 1+2: 60 µg/ strain | NA | NR | Single dose |
|  |  | Tx2 | IIV3-Adj (2015/2016, 2016/2017) | Egg-based | Year 1+2: 15 µg/ strain | Year 1: MF59; Year 2: MF59 | NR | Single dose |
|  |  | Tx3 | IIV3-SD (2015/2016) + IIV3-HD (2016/2017) | Egg-based | Year 1:15 µg/ strain; Year 2: 60 µg/ strain | NA | NR | Single dose |
|  |  | Tx4 | IIV3-SD (2015/2016) + IIV3-Adj (2016/2017) | Egg-based | Year 1+2: 15 µg/ strain | Year 1: NA; Year 2: MF59 | NR | Single dose |
| **Trial registry, 2021^50^** | Community | Tx1 | IIV4-HD | Egg-based | 60 µg/ strain | NA | Intramuscular | Single dose |
|  |  | Tx2 | IIV4-SD | Egg-based | 15 µg/strain | NA | Subcutaneous | Single dose |
| Note: **The** term "-Other" in the vaccine name indicates that there are varying dosages available for different strains**.** As such, it is not categorized as standard dose nor high dose as per guidelines.  **Abbreviations-** NR: Not reported; N: Northern hemisphere; S: Southern hemisphere; IIV3: Trivalent **i**nactivated influenza vaccine; IIV4: Quadrivalent **i**nactivated influenza vaccine; Tdap: Tetanus, diphtheria, pertussis; Adj: Adjuvanted; SD: Standard dosage; HD: High dosage; CC: Cell-cultured; RIV: Recombinant influenza vaccine; LAIV: Live attenuated influenza vaccine; USA: United States of America; UK: United Kingdom; RCT: Randomized controlled trial; ARI: Acute respiratory infection; LC-ARI: Laboratory-confirmed acute respiratory infection; ER: Emergency room; LCI: Laboratory-confirmed influenza; ILI: Influenza-like illness; AE: Adverse event; μg: micrograms; NA: Not applicable; Tx: treatment arm | | | | | | | | |

### **Appendix 13: List of Outcomes and Relevant Studies in Review**

| **Outcome Name** | **Number of Studies** | **Comparison** | **Number of Participants** | **Effect Estimates** | **Upper CI** | **Lower CI** | **References of Included Studies** |
| --- | --- | --- | --- | --- | --- | --- | --- |
| ARI Cases | 1 | IIV3-HD:IIV3-SD | 31,983 | 0.98 | 0.93 | 1.02 | DiazGranados, 2014 ^20^ |
| LC-ARI Cases | 0 |  |  |  |  |  |  |
| ER Visit for ARI | 0 |  |  |  |  |  |  |
| ER Visit for LCI | 1 | IIV3-HD:IIV3-SD | 31,983 | 1.29 | 0.48 | 3.45 | DiazGranados, 2014 ^20^ |
| Hospitalization for Pneumonia due to ILI | 0 |  |  |  |  |  |  |
| Hospitalization for Pneumonia due to ARI | 0 |  |  |  |  |  |  |
| Hospitalization for Pneumonia due to LCI | 0 |  |  |  |  |  |  |
| Inpatient or Outpatient Hospital Visit | 2 | IIV3-HD:IIV3-SD | 9158 | 0.97 | 0.83 | 1.14 | DiazGranados, 2013; Otten, 2020 ^15, 46^ |
| Hospitalization due to Cardiovascular Events | 0 |  |  |  |  |  |  |
| LCI Related Death | 0 |  |  |  |  |  |  |
| LCI Related Healthcare Interactions | 0 |  |  |  |  |  |  |
| Number of Hospitalization events due to cardiovascular events | 0 |  |  |  |  |  |  |
| Vascular AE related to influenza | 0 |  |  |  |  |  |  |
| ER Visit (any cause) | 2 | IIV3-HD:IIV3-SD | 53,008 | 0.9 | 0.86 | 0.95 | Gravenstein, 2017; McConeghy, 2020 ^23, 47^ |
|  |  | IIV3-Adj vs. IIV3-SD | 50,012 | 0.93 | 0.89 | 0.97 |  |
| ER Visit for Pneumonia (any cause) | 2 | IIV3-HD:IIV3-SD | 556 | 0.79 | 0.67 | 0.93 | Treanor, 1994; Gravenstein, 2017 ^23, 36^ |
|  |  | IIV3-SD:Placebo | 41 | 1.17 | 0.04 | 30.58 |  |
| Number of participants with vascular AE | 3 | IIV4-Adj:Tdap | 6761 | 0.74 | 0.49 | 1.19 | Scheifele, 2013; Beran, 2021; Schmader, 2021; ^16, 32, 33^ |
|  |  | IIV3-Adj:IIV3-HD | 757 | 5.04 | 0.24 | 16.5 |  |
|  |  | IIV3-Adj:IIV3-SD | 616 | 1.01 | 0.06 | 16.27 |  |
| Hospitalization for LCI | 4 | RIV:IIV4-SD | 9003 | 0.33 | 0.04 | 3.21 | Wongsurakiat, 2004; Dunkle, 2017; Teh, 2021; DiazGranados, 2014 ^11, 20, 24, 34^ |
|  |  | IIV3-SD:Placebo | 125 | 0.39 | 0.07 | 2.07 |  |
|  |  | IIV3-HD:IIV4-SD | 68 | 1.00 | 0.06 | 16.7 |  |
|  |  | IIV3-HD:IIV3-SD | 31,983 | 0.6 | 0.22 | 1.65 |  |
| Inpatient Hospitalization (any cause) | 1 | IIV4-SD:IIV3-Adj | 1016 | 0.86 | 0.56 | 1.34 | Cowling, 2020 ^30^ |
|  |  | IIV4-SD*:IIV3-HD* | 1018 | 1.16 | 0.73 | 1.84 |  |
|  |  | IIV4-SD*:RIV* | 843 | 1.25 | 0.73 | 2.14 |  |
|  |  | IIV3-Adj*:IIV3-HD* | 1018 | 1.34 | 0.85 | 2.11 |  |
|  |  | IIV3-Adj*:RIV* | 843 | 1.45 | 0.86 | 2.46 |  |
|  |  | IIV3-HD:RIV | 845 | 1.08 | 0.82 | 1.43 |  |
| ILI Cases | 5 | IIV4-Adj:Tdap | 6761 | 0.98 | 0.88 | 1.1 | Beran, 2021; Dunkle, 2017; Keitel 2009; Teh, 2021; Frey, 2014 ^14, 19, 24, 32, 34^ |
|  |  | RIV:IIV4-SD | 9003 | 0.99 | 0.89 | 1.09 |  |
|  |  | RIV:IIV3-SD | 870 | 0.96 | 0.55 | 1.65 |  |
|  |  | IIV3-HD:IIV4-SD | 68 | 1.21 | 0.36 | 4.07 |  |
|  |  | IIV3-Adj:IIV3-SD | 6961 | 1.03 | 0.87 | 1.21 |  |
| Hospitalization for ARI | 1 | IIV3-Adj:IIV30SD | 50,012 | 0.93 | 0.83 | 1.04 | McConeghy, 2021 ^47^ |
| LCI Cases | 2 | IIV4-Adj:Tdap | 6761 | 0.8 | 0.63 | 1.01 | Beran, 2021; McLean, 2021 ^32, 49^ |
| All Cause Mortality | 14 | IIV3-Adj:IV3-SD | 9150 | 1.05 | 0.99 | 1.1 | McConeghy, 2021; de Bruijn, 2006; Szymczakiewicz-Multanowska,  2009; Szymczakiewicz-Multanowska,  2012; Szymczakiewicz-Multanowska,  2012; Della Cioppa, 2012; Della Cioppa, 2014; Novartis Vaccines and Diagnostics, 2016; Otten, 2020; Trial registry, 2017; Sanchez, 2020; McLean, 2021; Trial Registry, 2021 ^37-50^ |
| Influenza-related Mortality | 2 | IIV3-Adj:IIV3-SD | 6961 | 0.75 | 0.17 | 3.36 | Treanor, 2017; Frey, 2014 ^19, 25^ |
|  |  | IIV4-SD:IIV3-SD | 1741 | 3.01 | 0.12 | 73.91 |  |
| Number of Vascular Adverse Events | 5 | RIV:IIV4-SD | 9003 | 0.89 | 0.3 | 2.6 | Dunkle, 2017; Szymczakiewicz-Multanowska, 2009; Szymczakiewicz-Multanowska, 2012; Novartis Vaccines and Diagnostics, 2016; ^24, 38, 39, 44^ |

**Abbreviations**- CI: confidence intervals, ARI: acute respiratory infections, LC-ARI: laboratory-confirmed acute respiratory infections, ER: emergency room, ILI: influenza-like illness, LCI: laboratory-confirmed influenza, AE: adverse events

*Within the scope of the current review, these studies and their reported outcomes but were excluded from the analyses for the following reasons: 1) coding was not possible as it would result in pooling of the arms with no resultant comparator, 2) data was reported but no counts were present, 3) the study was disconnected from the network, and 4) the study did not contribute to any pairwise meta-analyses.

### **Appendix 14: Aggregate Cochrane Risk-of-bias appraisal results (N=26)**

**Primary Outcomes**

**Laboratory-confirmed influenza (LCI) (n= 9)**

|  | Low | Some concern | High |
| --- | --- | --- | --- |
| Overall | 22% | 44% | 33% |
| Bias in selection of the reported result | 44% | 44% | 11% |
| Bias in measurement of the outcome | 100% |  |  |
| Bias due to missing outcome data | 78% | 22% |  |
| Bias due to deviations from intended interventions | 55% | 44% |  |
| Bias arising from the randomization process | 55% | 33% | 11% |

**Influenza-like illness (ILI) (n=9)**

|  | Low | Some concern | High |
| --- | --- | --- | --- |
| Overall | 33% | 44% | 22% |
| Bias in selection of the reported result | 44% | 44% | 11% |
| Bias in measurement of the outcome | 100% |  |  |
| Bias due to missing outcome data | 78% | 11% | 11% |
| Bias due to deviations from intended interventions | 56% | 44% |  |
| Bias arising from the randomization process | 78% | 22% |  |

**Secondary Outcomes**

**Emergency Room (ER) Visit for ILI (n=2)**

|  | Low | Some concern | High |
| --- | --- | --- | --- |
| Overall | 50% |  | 0.5 |
| Bias in selection of the reported result | 100% |  |  |
| Bias in measurement of the outcome | 100% |  |  |
| Bias due to missing outcome data | 50% | 0.5 |  |
| Bias due to deviations from intended interventions | 50% |  | 0.5 |
| Bias arising from the randomization process | 100% |  |  |

**Hospitalization for ILI (n=2)**

|  | Low | Some concern | High |
| --- | --- | --- | --- |
| Overall |  | 50% | 0.5 |
| Bias in selection of the reported result | 50% | 50% |  |
| Bias in measurement of the outcome | 100% |  |  |
| Bias due to missing outcome data | 50% | 50% |  |
| Bias due to deviations from intended interventions | 50% |  | 0.5 |
| Bias arising from the randomization process | 100% |  |  |

**Hospitalization for ARI (n=2)**

|  | Low | Some concern | High |
| --- | --- | --- | --- |
| Overall |  | 50% | 0.5 |
| Bias in selection of the reported result |  | 100% |  |
| Bias in measurement of the outcome | 1 |  |  |
| Bias due to missing outcome data | 0.5 | 50% |  |
| Bias due to deviations from intended interventions | 0.5 | 50% |  |
| Bias arising from the timing of identification or recruitment of participants in a cluster trial* |  | 100% |  |
| Bias arising from the randomization process | 1 |  |  |

**Number of vascular adverse events (n=8)**

|  | Low | Some concern | High |
| --- | --- | --- | --- |
| Overall | 50% | 38% | 0.12 |
| Bias in selection of the reported result | 50% | 50% |  |
| Bias in measurement of the outcome | 100% |  |  |
| Bias due to missing outcome data | 88% | 12% |  |
| Bias due to deviations from intended interventions | 88% | 12% |  |
| Bias arising from the randomization process | 75% | 25% |  |

**Inpatient hospitalization (any cause) (n=3)**

|  | Low | Some concern | High |
| --- | --- | --- | --- |
| Overall |  | 67% | 0.33 |
| Bias in selection of the reported result |  | 100% |  |
| Bias in measurement of the outcome | 1 |  |  |
| Bias due to missing outcome data | 0.67 |  | 0.33 |
| Bias due to deviations from intended interventions | 0.67 | 33% |  |
| Bias arising from the timing of identification or recruitment of participants in a cluster trial* | 1 |  |  |
| Bias arising from the randomization process | 0.67 | 33% |  |

**Outpatient visits (n=4)**

|  | Low | Some concern | High |
| --- | --- | --- | --- |
| Overall |  | 75% | 0.25 |
| Bias in selection of the reported result | 25% | 75% |  |
| Bias in measurement of the outcome | 100% |  |  |
| Bias due to missing outcome data | 75% | 25% |  |
| Bias due to deviations from intended interventions | 75% |  | 0.25 |
| Bias arising from the randomization process | 75% | 25% |  |

**All-cause mortality (n=20)**

|  | Low | Some concern | High |
| --- | --- | --- | --- |
| Overall | 45% | 40% | 0.15 |
| Bias in selection of the reported result | 70% | 30% |  |
| Bias in measurement of the outcome | 100% |  |  |
| Bias due to missing outcome data | 95% | 5% |  |
| Bias due to deviations from intended interventions | 65% | 30% | 0.05 |
| Bias arising from the timing of identification or recruitment of participants in a cluster trial* | 50% | 50% |  |
| Bias arising from the randomization process | 70% | 30% |  |

### **Appendix 15: Cochrane Risk-of-bias appraisal results (N=26)**

| **LCI** | | | | | | |
| --- | --- | --- | --- | --- | --- | --- |
| Study (Author, Year) | Bias arising from the randomization process | Bias due to deviations from intended interventions | Bias due to missing outcome data | Bias in measurement of the outcome | Bias in selection of the reported result | Overall |
| DiazGranados, 2013^15^ | Low | Some Concerns | Some Concern | Low | Low | Some Concerns |
| DiazGranados, 2014^20^ | Low | Low | Low | Low | Low | Low |
| Dunkle, 2017^24^ | Low | Some Concerns | Low | Low | High | High |
| Keitel, 2010^14^ | Some Concerns | Low | Low | Low | Some Concerns | Some Concerns |
| Wongsurakiat, 2004^11^ | Some Concerns | Low | Low | Low | Some Concerns | Some Concerns |
| Rudenko, 2001^10^ | Some Concerns | Some Concerns | Low | Low | Some Concerns | High |
| Teh, 2021^34^ | Low | Low | Low | Low | Low | Low |
| Loeb, 2020^28^ | Low | Low | Some Concern | Low | Some Concerns | Some Concerns |
| Belongia, 2020^29^ | High | Some Concerns | Low | Low | Low | High |

| **ILI** | | | | | | |
| --- | --- | --- | --- | --- | --- | --- |
| Study (Author, Year) | Bias arising from the randomization process | Bias due to deviations from intended interventions | Bias due to missing outcome data | Bias in measurement of the outcome | Bias in selection of the reported result | Overall |
| DiazGranados, 2013^15^ | Low | Some Concerns | Some Concerns | Low | Low | Some Concerns |
| DiazGranados, 2014^20^ | Low | Low | Low | Low | Low | Low |
| Frey, 2014^19^ | Low | Some Concerns | Low | Low | Some Concerns | Some Concerns |
| Beran, 2021^32^ | Low | Low | Low | Low | Low | Low |
| Dunkle, 2017^24^ | Low | Some Concerns | Low | Low | High | High |
| Allsup, 2004^12^ | Low | Some Concerns | High | Low | Some Concerns | High |
| Teh, 2021^34^ | Low | Low | Low | Low | Low | Low |
| Keitel, 2010^14^ | Some Concerns | Low | Low | Low | Some Concerns | Some Concerns |
| Wongsurakiat, 2004^11^ | Some Concerns | Low | Low | Low | Some Concerns | Some Concerns |

| **ER Visit for ILI** | | | | | | |
| --- | --- | --- | --- | --- | --- | --- |
| Study (Author, Year) | Bias arising from the randomization process | Bias due to deviations from intended interventions | Bias due to missing outcome data | Bias in measurement of the outcome | Bias in selection of the reported result | Overall |
| DiazGranados, 2014^20^ | Low | Low | Low | Low | Low | Low |
| DiazGranados, 2013^15^ | Low | High | Some Concerns | Low | Low | High |

| **Hospitalization for ILI** | | | | | | |
| --- | --- | --- | --- | --- | --- | --- |
| Study (Author, Year) | Bias arising from the randomization process | Bias due to deviations from intended interventions | Bias due to missing outcome data | Bias in measurement of the outcome | Bias in selection of the reported result | Overall |
| DiazGranados, 2014^20^ | Low | Low | Low | Low | Some Concerns | Some Concerns |
| DiazGranados, 2013^15^ | Low | High | Some Concerns | Low | Low | High |

| **Hospitalization for ARI** | | | | | | | |
| --- | --- | --- | --- | --- | --- | --- | --- |
| Study (Author, Year) | Bias arising from the randomization process | Bias arising from the timing of identification or recruitment of participants in a cluster trial* | Bias due to deviations from intended interventions | Bias due to missing outcome data | Bias in measurement of the outcome | Bias in selection of the reported result | Overall |
| DiazGranados, 2014^20^ | Low | NA | Low | Low | Low | Some Concerns | Some Concerns |
| Gravenstein, 2017^23^ | Low | Some Concerns | Some Concerns | Some Concerns | Low | Some Concerns | High |
| *Only applicable to Cluster-RCTs | | | | | | | |

| **Number of vascular adverse events** | | | | | | |
| --- | --- | --- | --- | --- | --- | --- |
| Study (Author, Year) | Bias arising from the randomization process | Bias due to deviations from intended interventions | Bias due to missing outcome data | Bias in measurement of the outcome | Bias in selection of the reported result | Overall |
| Essink, 2020^31^ | Low | Low | Low | Low | Low | Low |
| Chang, 2019^27^ | Low | Low | Low | Low | Low | Low |
| Tsang, 2014^18^ | Low | Low | Low | Low | Some Concerns | Some Concerns |
| Loeb, 2020^28^ | Low | Low | Low | Low | Low | Low |
| Falsey, 2009^13^ | Some Concerns | Low | Low | Low | Some Concerns | Some Concerns |
| DiazGranados, 2013^15^ | Low | Low | Some Concerns | Low | Some Concerns | Some Concerns |
| DiazGranadoz, 2014^20^ | Low | Low | Low | Low | Low | Low |
| Frey, 2014^19^ | Some Concerns | Some Concerns | Low | Low | Some Concerns | High |

| **Inpatient hospitalization (any cause)** | | | | | | | |
| --- | --- | --- | --- | --- | --- | --- | --- |
| Study | Bias arising from the randomization process | Bias arising from the timing of identification or recruitment of participants in a cluster trial* | Bias due to deviations from intended interventions | Bias due to missing outcome data | Bias in measurement of the outcome | Bias in selection of the reported result | Overall |
| Falsey, 2009^13^ | Some Concerns | NA | Low | Low | Low | Some Concerns | Some Concerns |
| DiazGranados, 2014^20^ | Low | NA | Low | High | Low | Some Concerns | High |
| Gravenstein, 2018^26^ | Low | Low | Some Concerns | Low | Low | Some Concerns | Some Concerns |
| *Only applicable to Cluster-RCTs | | | | | | | |

| **Outpatient visits** | | | | | | |
| --- | --- | --- | --- | --- | --- | --- |
| Study (Author, Year) | Bias arising from the randomization process | Bias due to deviations from intended interventions | Bias due to missing outcome data | Bias in measurement of the outcome | Bias in selection of the reported result | Overall |
| DiazGranados, 2014^20^ | Low | Low | Low | Low | Some Concerns | Some Concerns |
| DiazGranados, 2013^15^ | Low | High | Some Concerns | Low | Low | High |
| Allsup, 2004^12^ | Low | Low | Low | Low | Some Concerns | Some Concerns |
| Wongsurakiat, 2004^11^ | Some Concerns | Low | Low | Low | Some Concern | Some Concerns |

| **All-cause death** | | | | | | | |
| --- | --- | --- | --- | --- | --- | --- | --- |
| Study (Author, Year) | Bias arising from the randomization process | Bias arising from the timing of identification or recruitment of participants in a cluster trial* | Bias due to deviations from intended interventions | Bias due to missing outcome data | Bias in measurement of the outcome | Bias in selection of the reported result | Overall |
| Allsup, 2004^12^ | Low | NA | Low | Low | Low | Some Concerns | Some Concerns |
| Nace, 2014^21^ | Some Concerns | NA | Some Concerns | Low | Low | Some Concerns | High |
| Beran, 2021^32^ | Low | NA | Low | Low | Low | Low | Low |
| Schmader, 2021^33^ | Low | NA | Low | Low | Low | Low | Low |
| Vardeny, 2021^35^ | Low | NA | Low | Low | Low | Low | Low |
| Essink, 2020^31^ | Low | NA | Low | Low | Low | Low | Low |
| Chang, 2019^27^ | Low | NA | Low | Low | Low | Low | Low |
| Dunkle, 2017^24^ | Low | NA | Some Concerns | Low | Low | Low | Some Concerns |
| Treanor, 2017^25^ | Low | NA | Low | Low | Low | Low | Low |
| Pepin, 2013^17^ | Some Concerns | NA | Low | Low | Low | Low | Some Concerns |
| Scheifele, 2013^16^ | Low | NA | Low | Low | Low | Low | Low |
| Keitel, 2010^14^ | Some Concerns | NA | Low | Low | Low | Low | Some Concerns |
| Falsey, 2009^13^ | Some Concerns | NA | Low | Low | Low | Some Concerns | Some Concerns |
| Wongsurakiat, 2004^11^ | Some Concerns | NA | Some Concerns | Low | Low | Some Concerns | High |
| DiazGranados, 2013^15^ | Low | NA | High | Some Concerns | Low | Some Concerns | High |
| DiazGranados, 2014^20^ | Low | NA | Low | Low | Low | Low | Low |
| Frey, 2014^19^ | Some Concerns | NA | Some Concerns | Low | Low | Low | Some Concerns |
| Bart, 2016^22^ | Low | NA | Low | Low | Low | Low | Low |
| Gravenstein, 2017^23^ | Low | Some Concerns | Some Concerns | Low | Low | Low | Some Concerns |
| Gravenstein, 2018^26^ | Low | Low | Some Concerns | Low | Low | Some Concerns | Some Concerns |
| *Only applicable to Cluster-RCTs | | | | | | | |

### **Appendix 16: Small-study effects and publication bias assessment**

**Figure 1.** Comparison-adjusted funnel plot for the assessment of small-study effects and publication bias for all-cause mortality.

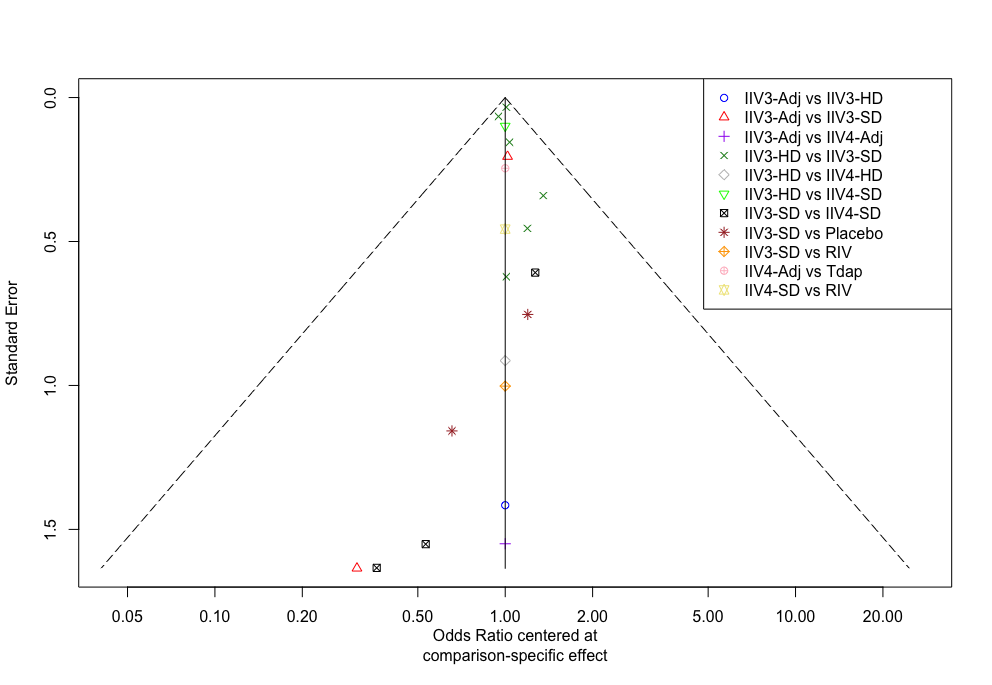

*Hierarchy of interventions based on clinical expertise as follows: 1) RIV4, 2) RIV3, 3) IIV4-HD, 4) IIV4-Adj, 5) IIV4-SD, 6) IIV3-HD, 7) IIV3-Adj, 8) IIV3-SD, 9) Tdap, 10) Placebo

**Abbreviations-** IIV3: Trivalent **i**nactivated influenza vaccine; IIV4: Quadrivalent **i**nactivated influenza vaccine; Adj: Adjuvanted; SD: Standard dosage; HD: High dosage; RIV: Recombinant influenza vaccine; Tdap: Tetanus, diphtheria, pertussis

### **Appendix 17: Network Meta-Analyses**

**Figure 1.** Network plots for the network meta-analyses with coding definition of interventions of a) laboratory-confirmed influenza, b) all-cause mortality, c) outpatient visits, d) number of vascular adverse events

**a)**
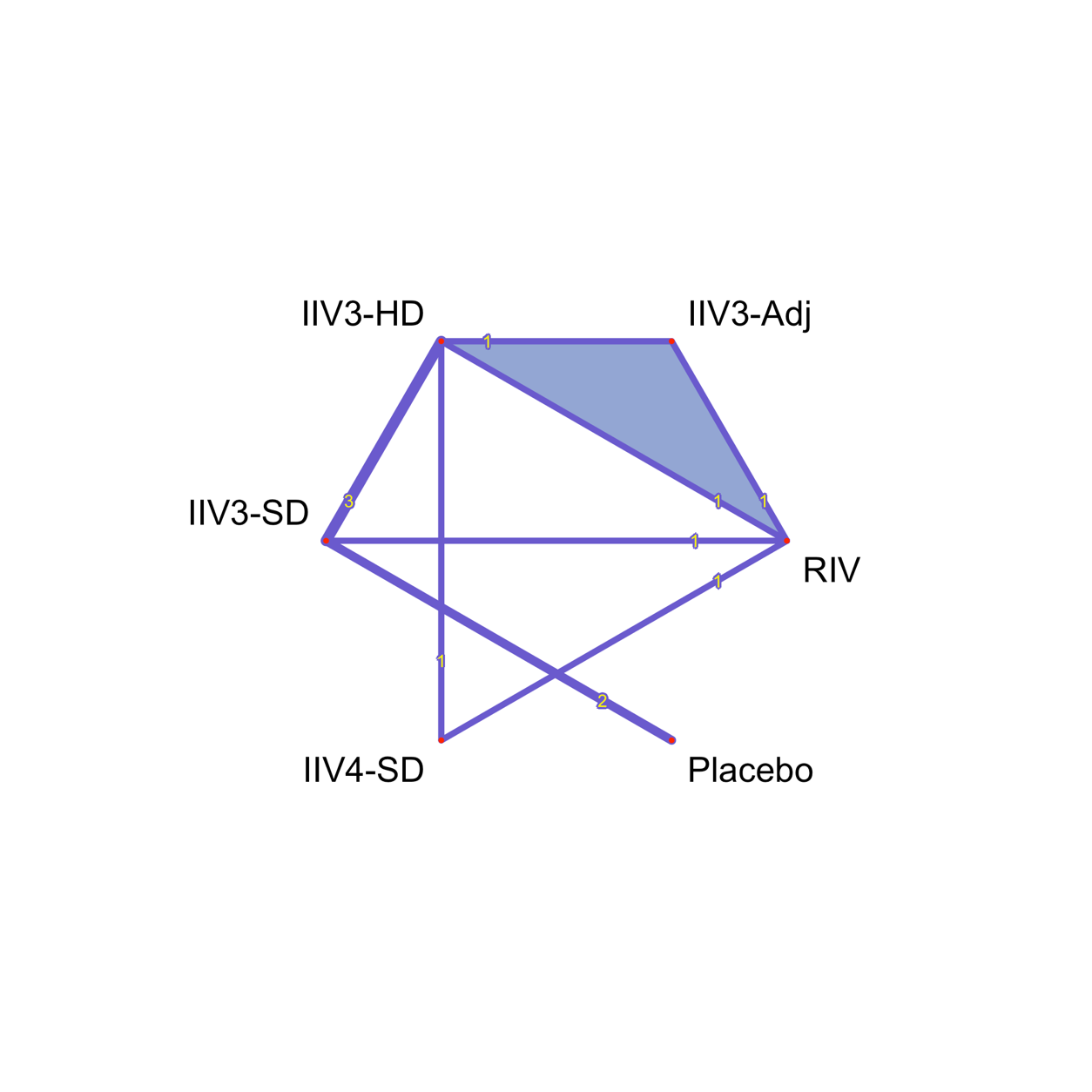
**b)**
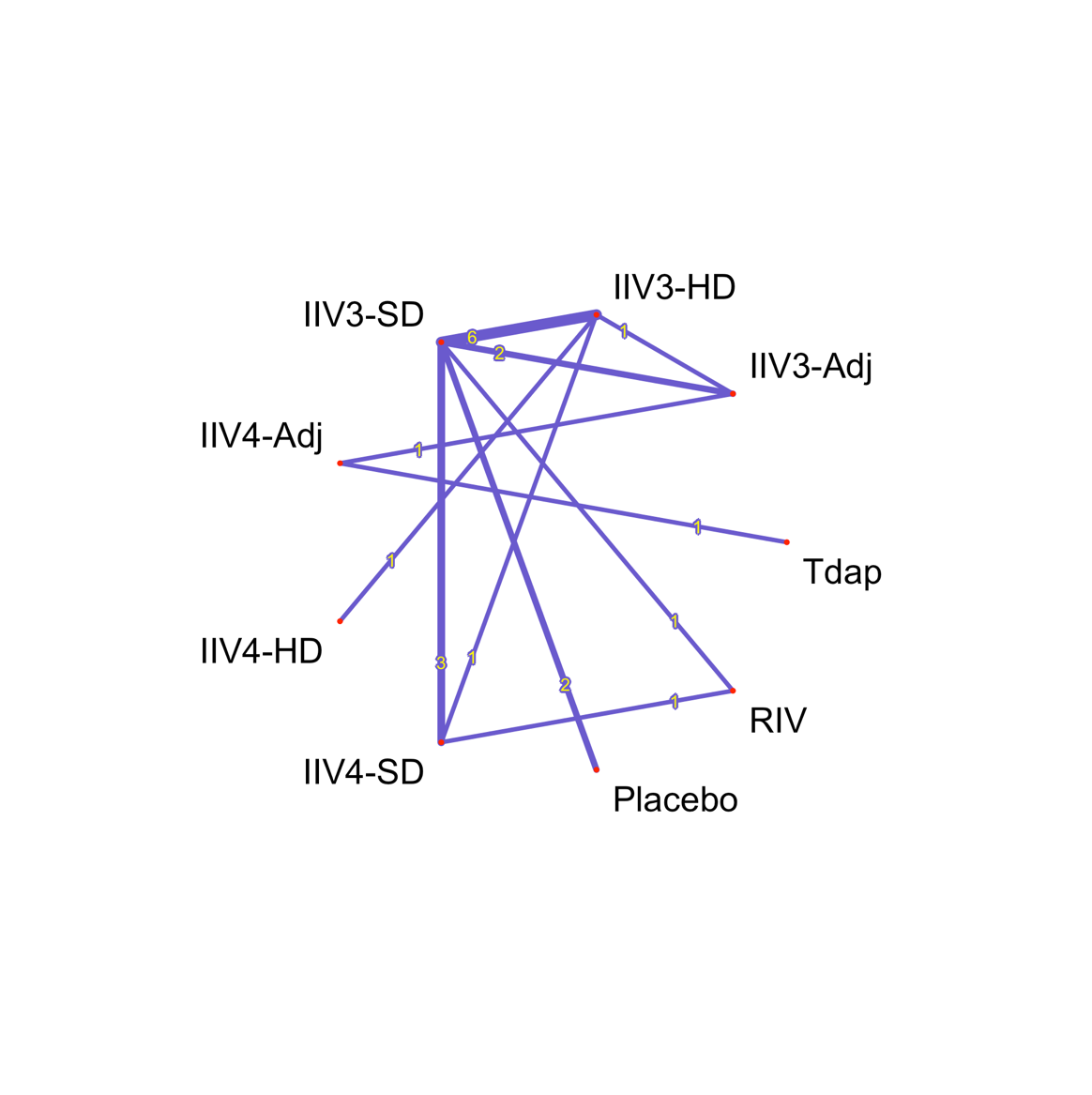

**c)**
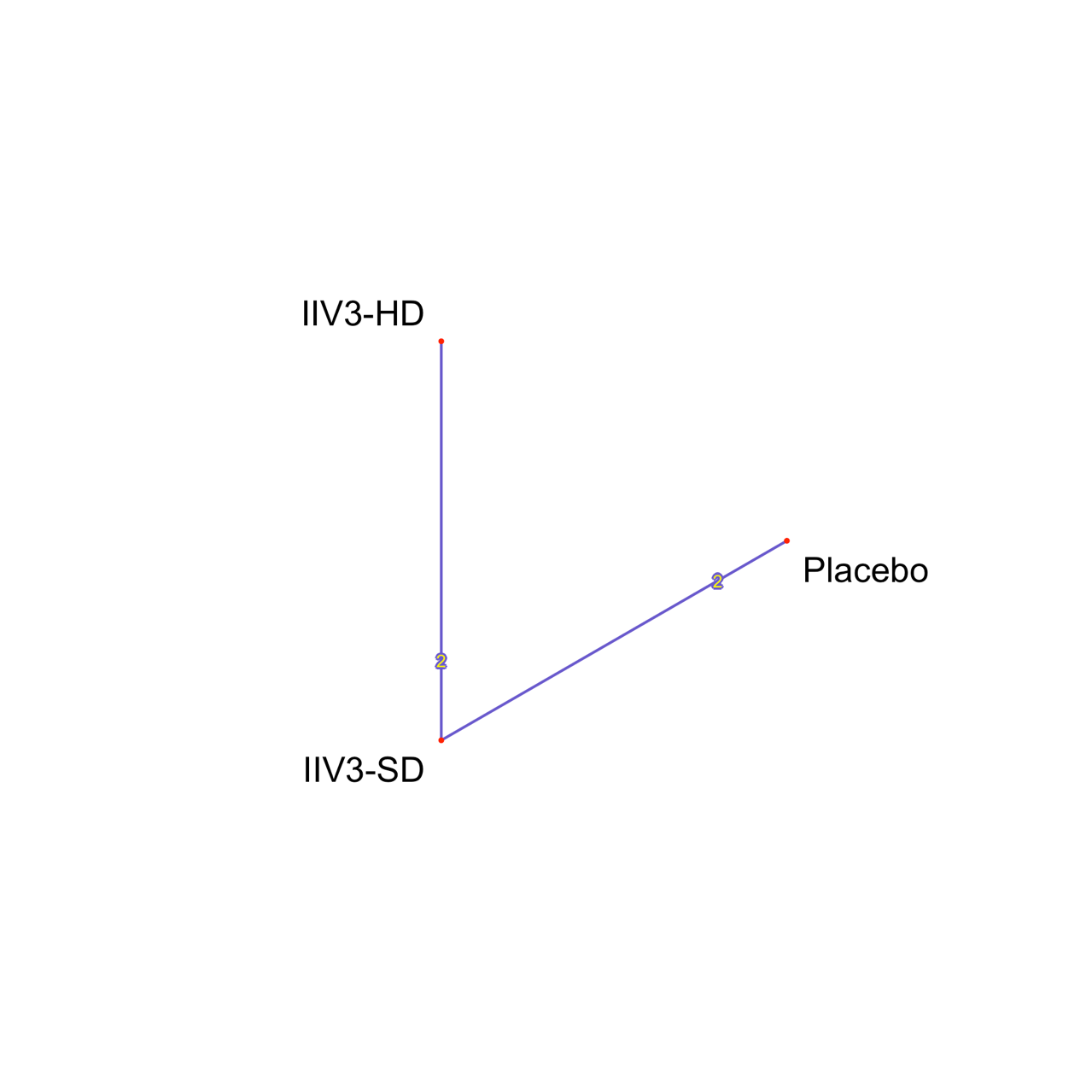
 **d)**
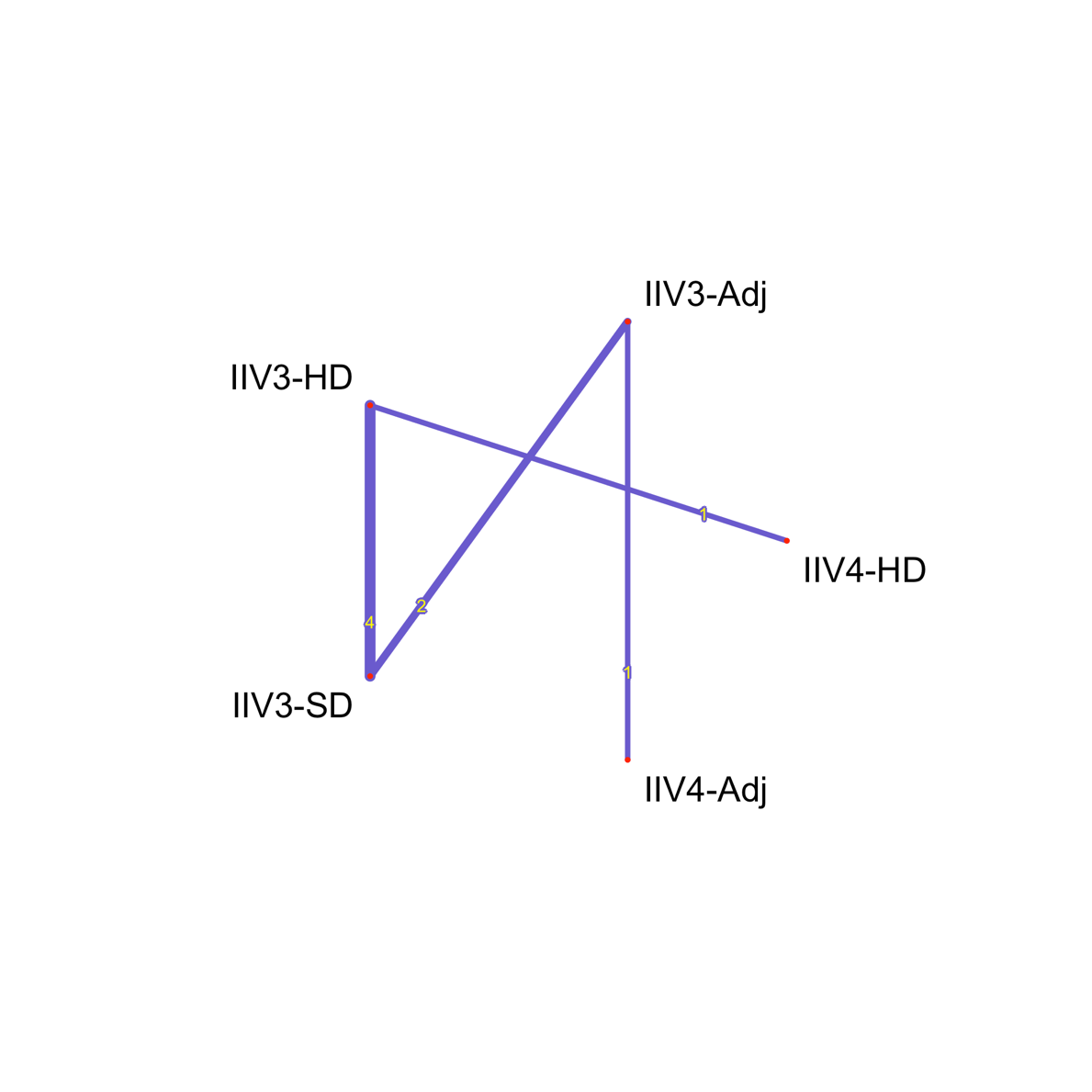

**Abbreviations-** IIV3: Trivalent **i**nactivated influenza vaccine; IIV4: Quadrivalent **i**nactivated influenza vaccine; Adj: Adjuvanted; SD: Standard dosage; HD: High dosage; RIV: Recombinant influenza vaccine; Tdap: Tetanus, diphtheria, pertussis

#### **Appendix 17A: Consistency**

**Table 1.** Summary of design-by-treatment interaction model test for global consistency across the four outcomes on which an NMA was conducted: laboratory-confirmed influenza (LCI), all-cause mortality, outpatient visits, and number of vascular adverse events.

| ***Design-by-treatment interaction model test for global consistency*** | | | | |
| --- | --- | --- | --- | --- |
| **Outcome** | **Chi-square-test** | **Degrees of Freedom** | **P-value** | **Heterogeneity SD** |
| LCI | 2.35 | 2 | 0.31 | 0.00 |
| All-cause mortality | 0.23 | 3 | 0.98 | 0.00 |
| Inpatient visits | Inconsistency cannot be assessed because there are no closed loops | | | |
| Number of vascular adverse events | Inconsistency cannot be assessed because there are no closed loops | | | |
| **Abbreviations-** LCI: Laboratory-confirmed influenza; SD: Standard deviation | | | | |

*P-value > 0.05 indicates no evidence of statistically significant global inconsistency

**Figure 1.** Forest plot for direct, indirect, and network estimates, along with 95% prediction interval where applicable for laboratory confirmed influenza.

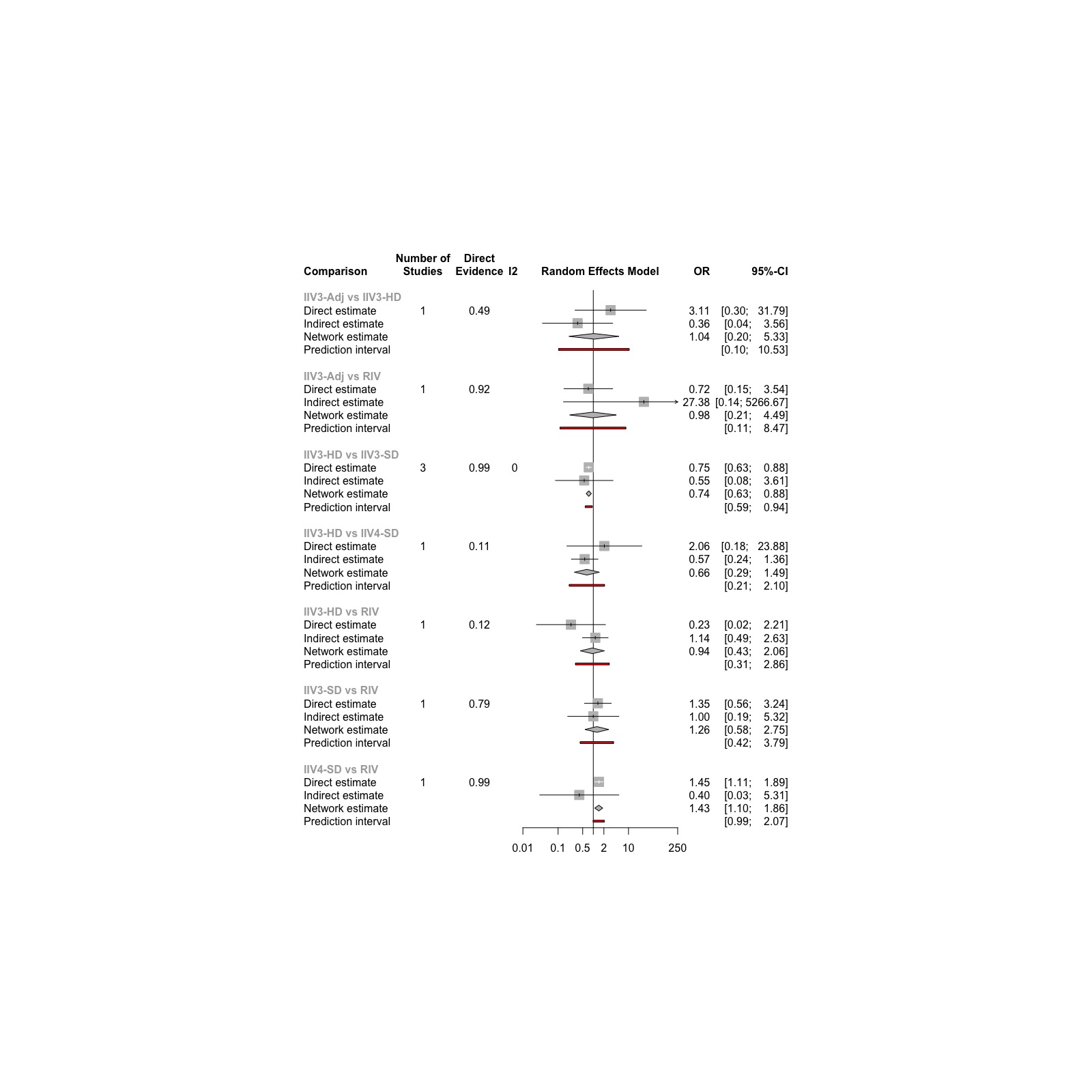

*Direct Evidence (%) column in forest plot indicates percentage contribution of direct evidence to network meta-analysis

**Abbreviations-** IIV3: Trivalent **i**nactivated influenza vaccine; IIV4: Quadrivalent **i**nactivated influenza vaccine; Adj: Adjuvanted; SD: Standard dosage; HD: High dosage; RIV: Recombinant influenza vaccine; OR: Odds ratio; CI: Confidence interval

**Figure 2.** Forest plot illustrating the ratio of odds ratios between direct and indirect estimates as estimated using the node-splitting approach for laboratory-confirmed influenza.

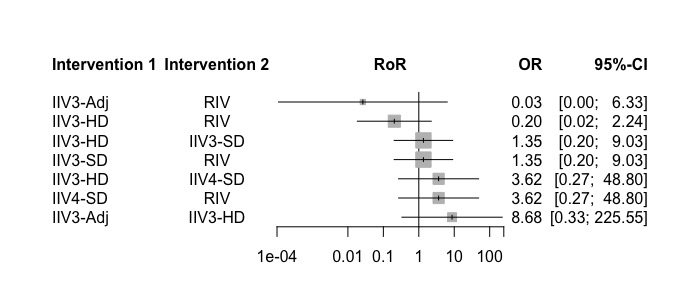

*If the confidence interval for RoR does not include the value 1, it suggests there is evidence of statistically significant inconsistency

**Abbreviations-** IIV3: Trivalent **i**nactivated influenza vaccine; IIV4: Quadrivalent **i**nactivated influenza vaccine; Adj: Adjuvanted; SD: Standard dosage; HD: High dosage; RIV: Recombinant influenza vaccine; RoR; Ratio of odds ratio; OR: Odds ratio; CI: Confidence interval

**Figure 3.** Forest plot for direct, indirect, and network estimates, along with 95% prediction interval where applicable for all-cause mortality

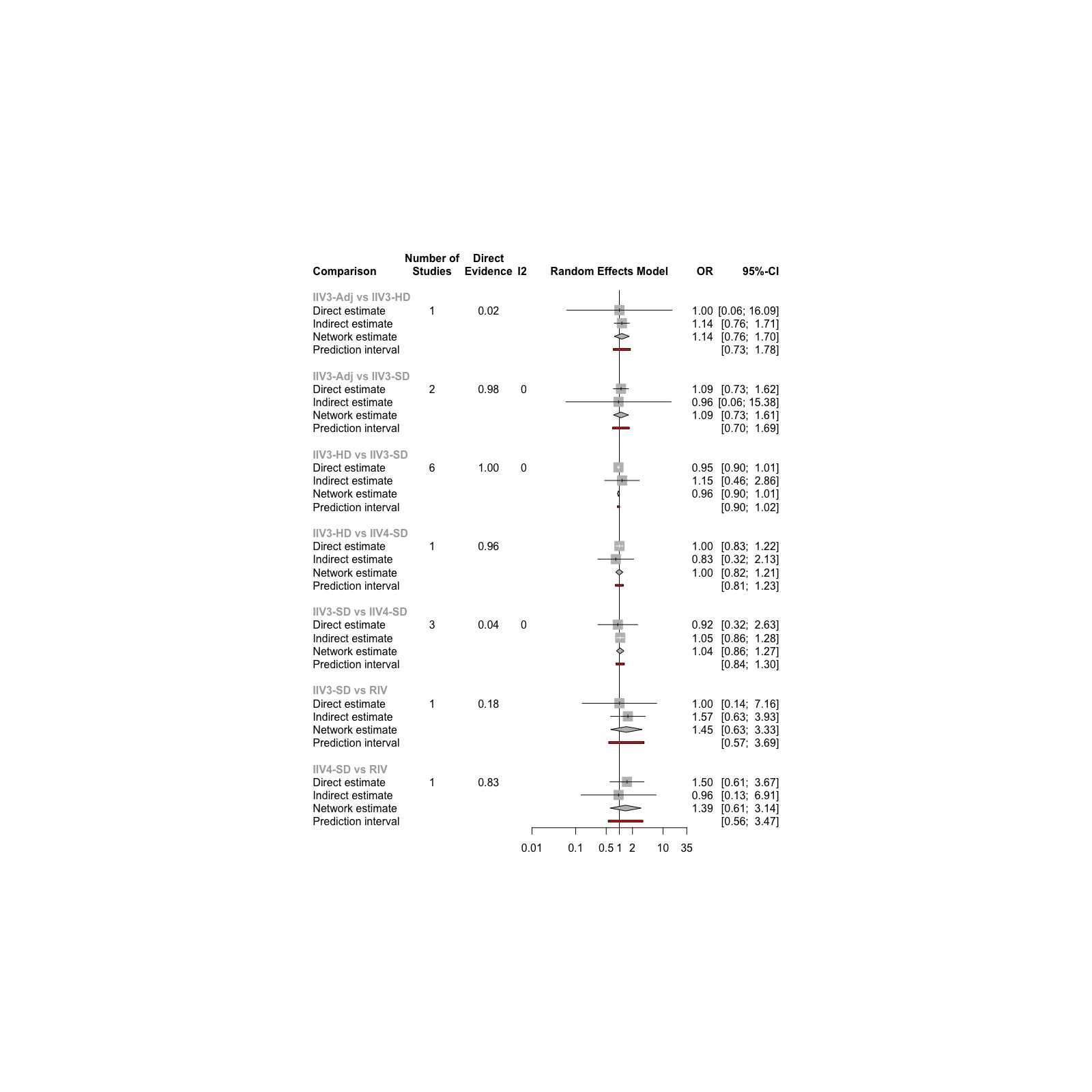

*Direct Evidence (%) column in forest plot indicates percentage contribution of direct evidence to network meta-analysis

**Abbreviations-** IIV3: Trivalent **i**nactivated influenza vaccine; IIV4: Quadrivalent **i**nactivated influenza vaccine; Adj: Adjuvanted; SD: Standard dosage; HD: High dosage; OR: Odds ratio; CI: Confidence interval

**Figure 4.** Forest plot illustrating the ratio of odds ratios between direct and indirect estimates as estimated using the node-splitting approach for all-cause mortality

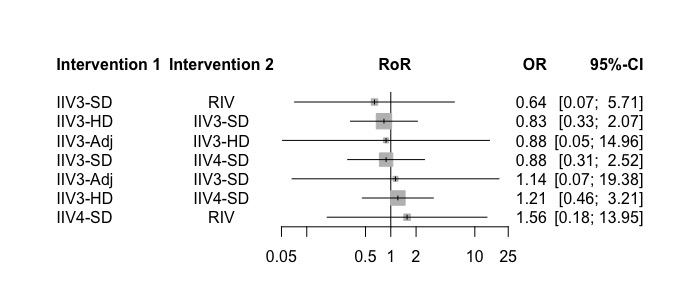

*If the confidence interval for RoR does not include the value 1, it suggests there is evidence of statistically significant inconsistency

**Abbreviations-** IIV3: Trivalent **i**nactivated influenza vaccine; IIV4: Quadrivalent **i**nactivated influenza vaccine; Adj: Adjuvanted; SD: Standard dosage; HD: High dosage; RoR; Ratio of odds ratio; OR: Odds ratio; CI: Confidence interval

#### **Appendix 17B: Transitivity Tables**

**Table 1.** Transitivity table for laboratory-confirmed influenza across age, sex, and overall risk of bias.

| **Transitivity for laboratory-confirmed influenza** | | | | | | | | |
| --- | --- | --- | --- | --- | --- | --- | --- | --- |
| **Comparison** | **Number of Studies** | **Age – mean** | **Sex (F%) – mean** | **Low RoB – n (%)** | **Some Concern RoB – n (%)** | **High RoB – n (%)** | **Mode RoB (n)** | **Year of Publication – mean** |
| IIV3-Adj vs. IIV3-HD | 1 | 70.00 | 56.20 % | . | . | 1 (100%) | High (1) | 2020 |
| IIV3-Adj vs. RIV | 1 | 70.00 | 56.20 % | . | . | 1 (100%) | High (1) | 2020 |
| IIV3-HD vs. IIV3-SD | 3 | 74.37 | 59.08 % | 1 (33.33%) | 2 (66.67%) | . | Some Concern (2) | 2015.67 |
| IIV3-HD vs. IIV4-SD | 1 | 60.00 | 32.00 % | 1 (100%) | . | . | Low (1) | 2021 |
| IIV3-HD vs. RIV | 1 | 70.00 | 56.20 % | . | . | 1 (100%) | High (1) | 2020 |
| IIV3-SD vs. Placebo | 2 | 70.45 | 37.47 % | . | 1 (50%) | 1 (50%) | Some concerns/high (1) | 2002.5 |
| IIV3-SD vs. RIV | 1 | 72.90 | 53.20 % | . | 1 (100%) | . | Some Concern (1) | 2010 |
| RIV4 vs. IIV4-SD | 1 | 64.10 | 58.50 % | . | . | 1 (100%) | High (1) | 2017 |
| **Abbreviations-** IIV3: Trivalent **i**nactivated influenza vaccine; IIV4: Quadrivalent **i**nactivated influenza vaccine; Adj: Adjuvanted; SD: Standard dosage; HD: High dosage; RIV: Recombinant influenza vaccine; RoB: Risk of bias | | | | | | | | |

**Table 2.** Transitivity table for number of vascular adverse events across age, sex, and overall risk of bias.

| **Transitivity for number of vascular adverse events** | | | | | | | | |
| --- | --- | --- | --- | --- | --- | --- | --- | --- |
| **Comparison** | **Number of Studies** | **Age – mean** | **Sex (F%) – mean** | **Low RoB – n (%)** | **Some Concern RoB – n (%)** | **High RoB – n (%)** | **Mode RoB (n)** | **Year of Publication – mean** |
| IIV3-Adj vs. IIV4-SD | 2 | 72.85 | 62.07 % | 1 (50%) | . | 1 (50%) | Low/High (1) | 2013.5 |
| IIV3-Adj vs. IIV4-Adj | 1 | 72.50 | 56.60 % | 1 (100%) | . | . | Low (1) | 2020 |
| IIV3-HD vs. IIV3-SD | 4 | 73.03 | 54.66 % | 1 (25%) | 3 (75%) | . | Some Concern (3) | 2012.5 |
| IIV3-HD vs. IIV4-HD | 1 | 73.00 | 54.90 % | 1 (100%) | . | . | Low (1) | 2019 |
| **Abbreviations-** IIV3: Trivalent **i**nactivated influenza vaccine; IIV4: Quadrivalent **i**nactivated influenza vaccine; Adj: Adjuvanted; SD: Standard dosage; HD: High dosage; RoB: Risk of bias | | | | | | | | |

**Table 3.** Transitivity table for outpatient visits across age, sex, and overall risk of bias.

| **Transitivity for outpatient visits** | | | | | | | | |
| --- | --- | --- | --- | --- | --- | --- | --- | --- |
| **Comparison** | **Number of Studies** | **Age – mean** | **Sex (F%) – mean** | **Low RoB – n (%)** | **Some Concern RoB – n (%)** | **High RoB – n (%)** | **Mode RoB (n)** | **Year of Publication – mean** |
| IIV3-HD vs. IIV3-SD | 2 | 73.05 | 55.12 % | . | 2 (50%) | 1 (50%) | Some Concern/High (1) | 2013.5 |
| IIV3-SD vs. Placebo | 2 | 68.67 | 26.20 % | . | 2 (100%) | . | Some Concern (2) | 2004 |
| **Abbreviations-** IIV3: Trivalent **i**nactivated influenza vaccine; IIV4: Quadrivalent **i**nactivated influenza vaccine; SD: Standard dosage; HD: High dosage; RoB: Risk of bias | | | | | | | | |

**Table 4.** Transitivity table for all-cause mortality across age, sex, and overall risk of bias.

| **Transitivity for all-cause mortality** | | | | | | | | |
| --- | --- | --- | --- | --- | --- | --- | --- | --- |
| **Comparison** | **Number of Studies** | **Age – median** | **Sex (F%) – median** | **Low RoB – n (%)** | **Some Concern RoB – n (%)** | **High RoB – n (%)** | **Mode RoB (n)** | **Year of Publication – mean** |
| IIV3-Adj vs. IIV3-HD | 1 | 76.50 | 55.50 % | 1 (100%) | . | . | Low (1) | 2021 |
| IIV3-Adj vs. IIV3-SD | 2 | 72.85 | 62.07 % | 1 (50%) | 1 (50%) | . | Low/Some Concern (1) | 2013.5 |
| IIV3-Adj vs. IIV4-Adj | 1 | 72.50 | 56.60 % | 1 (100%) |  | . | Low (1) | 2020 |
| IIV3-HD vs. IIV3-SD | 6 | 78.95 | 62.91 % | 1 (16.7%) | 3 (50%) | 2 (33.33%) | Some Concern (3) | 2014.17 |
| IIV3-HD vs. IIV4-HD | 1 | 73.00 | 54.90 % | 1 (100%) | . | . | Low (1) | 2019 |
| IIV3-HD vs. IIV4-HD | 1 | 65.50 | 28.30 % | 1 (100%) | . | . | Low (1) | 2021 |
| IIV3-SD vs. IIV4-SD | 3 | NR | 54.00 % | 2 (66.67%) | 1 (33.33%) | . | Low (2) | 2015.33 |
| IIV3-SD vs. Placebo | 2 | 68.67 | 26.20 % | . | 2 (100%) | . | Some Concern (2) | 2004 |
| IIV3-SD vs. RIV | 1 | 79.20 | 53.20 % | . | 1 (100%) | . | Some Concern (1) | 2010 |
| IIV4-Adj vs. Tdap | 1 | 71.90 | 61.80 % | 1 (100%) | . | . | Low (1) | 2021 |
| IIV4-SD vs. RIV | 1 | 64.10 | 58.50 % | . | 1 (100%) | . | Some Concern (1) | 2017 |
| ***Note:** Enrollment in one of the studies in this intervention comparison (Pepin, 2013) was stratified by age at each site into adults 18–60 and >60 years of age. However, only data from the >60 age group was used in the analysis. The remaining two studies did not report the median/mean age.  **Abbreviations-** IIV3: Trivalent **i**nactivated influenza vaccine; IIV4: Quadrivalent **i**nactivated influenza vaccine; Adj: Adjuvanted; SD: Standard dosage; HD: High dosage; RIV: Recombinant influenza vaccine; Tdap: Tetanus, diphtheria, pertussis; RoB: Risk of bias | | | | | | | | |

#### **Appendix 17C: Network Results**

**Laboratory-confirmed influenza**

**Table 1.** Summary of network meta-analysis results for laboratory-confirmed influenza with original coding of interventions.

| ***Network Meta-analysis Results; 9 studies, 52,202 participants, 7 treatments*** | | | | | | | | |
| --- | --- | --- | --- | --- | --- | --- | --- | --- |
| ***Comparison*** | ***OR*** | ***seOR*** | ***Lower CI*** | ***Upper CI*** | ***Statistic*** | ***p-value*** | ***Lower PI*** | ***Upper PI*** |
| IIV3-Adj-IIV3-HD | 1.04 | 2.30 | 0.20 | 5.33 | 0.05 | 0.96 | 0.10 | 10.53 |
| IIV3-Adj:IIV3-SD | 0.77 | 2.30 | 0.15 | 3.96 | -0.31 | 0.76 | 0.08 | 7.81 |
| IIV3-Adj:IIV4-SD | 0.69 | 2.20 | 0.15 | 3.21 | -0.48 | 0.63 | 0.08 | 6.11 |
| IIV3-Adj:Placebo | 0.2424 | 2.50 | 0.04 | 1.47 | -1.54 | 0.12 | 0.02 | 3.10 |
| IIV3-Adj:RIV | 0.98 | 2.17 | 0.21 | 4.49 | -0.03 | 0.98 | 0.08 | 8.47 |
| IIV3-HD:IIV3-SD | 0.74 | 1.09 | 0.63 | 0.88 | -3.51 | 0.00 | 0.59 | 0.94 |
| IIV3-HD: IIV4-SD | 0.66 | 1.52 | 0.29 | 1.49 | -1.00 | 0.32 | 0.21 | 2.10 |
| IIV3-HD: Placebo | 0.23 | 1.48 | 0.11 | 0.51 | -3.69 | 0.00 | 0.08 | 0.70 |
| IIV3-HD: RIV | 0.94 | 1.49 | 0.43 | 2.06 | -0.15 | 0.88 | 0.31 | 2.86 |
| IIV3-SD: IIV4-SD | 0.88 | 1.51 | 0.39 | 1.99 | -0.30 | 0.77 | 0.28 | 2.79 |
| IIV3-SD: Placebo | 0.31 | 1.47 | 0.15 | 0.67 | -3.01 | 0.00 | 0.11 | 0.92 |
| IIV3-SD: RIV | 1.26 | 1.49 | 0.58 | 2.75 | 0.59 | 0.55 | 0.42 | 3.79 |
| IIV4-SD: Placebo | 0.35 | 1.76 | 0.12 | 1.07 | -1.83 | 0.07 | 0.07 | 1.70 |
| IIV4-SD: RIV | 1.43 | 1.14 | 1.10 | 1.86 | 2.67 | 0.01 | 0.99 | 2.07 |
| RIV: Placebo | 0.25 | 1.74 | 0.08 | 0.73 | -2.52 | 0.01 | 0.05 | 1.15 |
| Common within-network between-study SD (tau) | <0.0001 |  |  |  |  |  |  |  |
| I-square | 0% |  |  |  |  |  |  |  |
| **Abbreviations-** IIV3: Trivalent **i**nactivated influenza vaccine; IIV4: Quadrivalent **i**nactivated influenza vaccine; Adj: Adjuvanted; SD: Standard dosage; HD: High dosage; RIV: Recombinant influenza vaccine; OR: Odds ratio; CI: Confidence interval; PI: Prediction interval; se: Standard error | | | | | | | | |

**Figure 1.** Treatment ranking of interventions using P-score for laboratory-confirmed influenza with original coding of interventions.

| ***Treatment Ranking*** | |
| --- | --- |
| ***Treatment*** | ***P-score*** |
| IIV3-HD | 0.78 |
| RIV | 0.73 |
| IIV3-Adj | 0.65 |
| IIV3-SD | 0.45 |
| IIV4-SD | 0.37 |
| Placebo | 0.02 |

**Abbreviations-** IIV3: Trivalent **i**nactivated influenza vaccine; IIV4: Quadrivalent **i**nactivated influenza vaccine; Adj: Adjuvanted; SD: Standard dosage; HD: High dosage; RIV: Recombinant influenza vaccine

**Table 2**. Vaccine efficacy from network meta-analysis results for laboratory-confirmed influenza with original coding of interventions

| ***Comparison*** | ***OR*** | ***RR*** | ***VE*** |
| --- | --- | --- | --- |
| IIV3-Adj-IIV3-HD | 1.04 (0.2,5.33) | 1.03 (0.24,2.87) | -3.18 (-186.6,76.22) |
| IIV3-Adj:IIV3-SD | 0.77 (0.15,3.96) | 0.81 (0.18,2.49) | 19.32 (-149.41,81.95) |
| IIV3-Adj:IIV4-SD | 0.69 (0.15, 3.21) | 0.74 (0.18,2.23) | 26.47 (-123.1,81.95) |
| IIV3-Adj:Placebo | 0.24 (0.04, 1.47) | 0.28 (0.05,1.34) | 71.73 (-34.45,95.06) |
| IIV3-Adj:RIV | 0.98 (0.21,4.49) | 0.98 (0.25,2.65) | 1.61 (-165.22,75.09) |
| IIV3-HD:IIV3-SD | 0.74 (0.63,0.88) | 0.78 (0.68,0.90) | 21.97 (9.85,32) |
| IIV3-HD: IIV4-SD | 0.66 (0.29,1.49) | 0.71 (0.34,1.36) | 28.22 (-35.79,66.24) |
| IIV3-HD: Placebo | 0.23 (0.11,0.51) | 0.27 (0.13,0.56) | 72.85 (43.5,86.64) |
| IIV3-HD: RIV | 0.94 (0.43,2.06) | 0.95 (0.48,1.70) | 4.67 (-70.18,51.51) |
| IIV3-SD: IIV4-SD | 0.88 (0.39,1.99) | 0.90 (0.44,1.66) | 9.85 (-66.31,55.63) |
| IIV3-SD: Placebo | 0.31 (0.15,0.67) | 0.36 (0.18,0.72) | 64.08 (28.3,81.95) |
| IIV3-SD: RIV | 1.26 (0.58,2.75) | 1.20 (0.63,2.04) | -19.81 (-104.09,36.72) |
| IIV4-SD: Placebo | 0.35 (0.12,1.07) | 0.40 (0.15,1.06) | 59.81 (-5.53,85.46) |
| IIV4-SD: RIV | 1.43 (1.1,1.85) | 1.32 (1.08,1.59) | -31.75 (-58.87,-7.86) |
| RIV:Placebo | 0.25 (0.08,0.73) | 0.29 (0.10,0.77) | 70.63 (22.86,90,21) |
| **Abbreviations-** IIV3: Trivalent **i**nactivated influenza vaccine; IIV4: Quadrivalent **i**nactivated influenza vaccine; Adj: Adjuvanted; SD: Standard dosage; HD: High dosage; RIV: Recombinant influenza vaccine; OR: odds ratio; RR: relative risk; VE: vaccine efficacy | | | |

**Number of Vascular Adverse Events**

**Table 3.** Summary of network meta-analysis results for number of vascular adverse events with original coding of interventions.

| ***Network Meta-analysis results; 8 studies, 57,677 patients, 5 treatments*** | | | | | | | | |
| --- | --- | --- | --- | --- | --- | --- | --- | --- |
| ***Comparison*** | ***IRR*** | ***seOR*** | ***Lower CI*** | ***Upper CI*** | ***Statistic*** | ***p-value*** | ***Lower PI*** | ***Upper PI*** |
| IIV3-Adj: IIV3-HD | 1.31 | 1.37 | 0.71 | 2.43 | 0.85 | 0.39 | 0.30 | 5.71 |
| IIV3-Adj: IIV3-SD | 0.90 | 1.30 | 0.54 | 1.50 | -0.41 | 0.69 | 0.23 | 3.50 |
| IIV3-Adj: IIV4-Adj | 5.00 | 1.44 | 2.46 | 10.18 | 4.44 | 0.00 | 1.03 | 24.22 |
| IIV3-Adj: IIV4-HD | 1.96 | 1.61 | 0.77 | 4.99 | 1.42 | 0.16 | 0.31 | 12.57 |
| IIV3-HD: IIV3-SD | 0.69 | 1.19 | 0.48 | 0.97 | -2.11 | 0.03 | 0.20 | 2.32 |
| IIV3-HD: IIV4-Adj | 3.82 | 1.62 | 1.49 | 9.80 | 2.79 | 0.01 | 0.59 | 24.79 |
| IIV3-HD: IIV4-HD | 1.50 | 1.43 | 0.75 | 3.01 | 1.14 | 0.25 | 0.31 | 7.14 |
| IIV4-Adj: IIV3-SD | 0.18 | 1.56 | 0.07 | 0.43 | -3.84 | 0.00 | 0.03 | 1.07 |
| IIV4-HD: IIV3-SD | 0.46 | 1.49 | 0.21 | 1.00 | -1.96 | 0.05 | 0.09 | 2.41 |
| IIV4-Adj: IIV4-HD | 0.39 | 1.82 | 0.12 | 1.27 | -1.56 | 0.12 | 0.04 | 3.50 |
| Common within-network between-study SD | 0.34 |  |  |  |  |  |  |  |
| I-square | 97.00% |  |  |  |  |  |  |  |
| **Abbreviations-** IIV3: Trivalent inactivated influenza vaccine; IIV4: Quadrivalent inactivated influenza vaccine; Adj: Adjuvanted; SD: Standard dosage; HD: High dosage; IRR: Incidence rate ratio; CI: Confidence interval; PI: Prediction interval; se: Standard error | | | | | | | | |

**Figure 2.** Treatment ranking of interventions using P-score for number of vascular adverse events with original coding of interventions.

| ***Treatment Ranking*** | |
| --- | --- |
| ***Treatment*** | ***P-score*** |
| IIV4-Adj | 0.98 |
| IIV4-HD | 0.71 |
| IIV3-HD | 0.48 |
| IIV3-Adj | 0.23 |
| IIV3-SD | 0.10 |

**Abbreviations-** IIV3: Trivalent inactivated influenza vaccine; IIV4: Quadrivalent inactivated influenza vaccine; Adj: Adjuvanted; SD: Standard dosage; HD: High dosage

**Outpatient Visits**

**Table 4.** Summary of network meta-analysis results for outpatient visits with original coding of interventions.

| ***Network Meta-analysis results; 4 studies, 41,995 patients, 3 treatments*** | | | | | | | | |
| --- | --- | --- | --- | --- | --- | --- | --- | --- |
| ***Comparison*** | ***OR*** | ***seOR*** | ***Lower CI*** | ***Upper CI*** | ***Statistic*** | ***p-value*** | ***Lower PI*** | ***Upper PI*** |
| IIV3-HD: IIV3-SD | 1.01 | 1.09 | 0.85 | 1.21 | 0.16 | 0.87 | 0.17 | 6.09 |
| IIV3-HD: Placebo | 0.65 | 1.36 | 0.35 | 1.19 | -1.40 | 0.16 | 0.01 | 41.95 |
| IIV3-SD: Placebo | 0.64 | 1.34 | 0.36 | 1.14 | -1.51 | 0.13 | 0.01 | 35.50 |
| Common within-network between-study SD | 0.11 |  |  |  |  |  |  |  |
| I-square | 60.70% |  |  |  |  |  |  |  |
| **Abbreviations-** IIV3: Trivalent **i**nactivated influenza vaccine; SD: Standard dosage; HD: High dosage; OR: Odds ratio; CI: Confidence interval; PI: Prediction interval; se: Standard error | | | | | | | | |

**Figure 3.** Treatment ranking of interventions using P-score for outpatient visits with original coding of interventions.

| ***Treatment Ranking*** | |
| --- | --- |
| ***Treatment*** | ***P-score*** |
| IIV3-SD | 0.75 |
| IIV3-HD | 0.68 |
| Placebo | 0.07 |

**Abbreviations-** IIV3: Trivalent **i**nactivated influenza vaccine; SD: Standard dosage; HD: High dosage;

**All-cause Mortality**

**Table 5.** Summary of network meta-analysis results for all-cause mortality with original coding of interventions.

| ***Network Meta-analysis results; 20 studies, 140,577 patients, 9 treatments*** | | | | | | | | |
| --- | --- | --- | --- | --- | --- | --- | --- | --- |
| ***Comparison*** | ***OR*** | ***seOR*** | ***Lower CI*** | ***Upper CI*** | ***Statistic*** | ***p-value*** | ***Lower PI*** | ***Upper PI*** |
| IIV3-Adj: IIV3-HD | 1.14 | 1.23 | 0.76 | 1.70 | 0.64 | 0.52 | 0.73 | 1.78 |
| IIV3-Adj: IIV3-SD | 1.09 | 1.22 | 0.73 | 1.61 | 0.42 | 0.67 | 0.70 | 1.69 |
| IIV3-Adj: IIV4-Adj | 0.20 | 4.71 | 0.01 | 4.16 | -1.04 | 0.30 | 0.01 | 6.05 |
| IIV3-Adj: IIV4-HD | 1.52 | 2.55 | 0.24 | 9.51 | 0.45 | 0.66 | 0.19 | 11.92 |
| IIV3-Adj: IIV4-SD | 1.14 | 1.25 | 0.73 | 1.77 | 0.57 | 0.57 | 0.69 | 1.86 |
| IIV3-Adj: Placebo | 1.60 | 1.94 | 0.44 | 5.85 | 0.71 | 0.48 | 0.37 | 6.87 |
| IIV3-Adj:RIV | 1.58 | 1.60 | 0.63 | 3.96 | 0.97 | 0.33 | 0.56 | 4.43 |
| IIV3-Adj:Tdap | 0.19 | 4.80 | 0.01 | 4.20 | -1.05 | 0.30 | 0.01 | 6.13 |
| IIV3-HD: IIV3-SD | 0.96 | 1.03 | 0.90 | 1.01 | -1.57 | 0.12 | 0.90 | 1.02 |
| IIV3-HD: IIV4-Adj | 0.18 | 4.77 | 0.01 | 3.75 | -1.11 | 0.27 | 0.01 | 5.47 |
| IIV3-HD: IIV4-HD | 1.33 | 2.49 | 0.22 | 7.99 | 0.31 | 0.75 | 0.18 | 9.96 |
| IIV3-HD: IIV4-SD | 1.00 | 1.10 | 0.82 | 1.21 | -0.03 | 0.98 | 0.81 | 1.23 |
| IIV3-HD: Placebo | 1.40 | 1.88 | 0.41 | 4.84 | 0.53 | 0.59 | 0.35 | 5.63 |
| IIV3-HD:RIV | 1.38 | 1.53 | 0.60 | 3.18 | 0.77 | 0.44 | 0.55 | 3.52 |
| IIV3-HD:Tdap | 0.17 | 4.87 | 0.01 | 3.78 | -1.12 | 0.26 | 0.01 | 5.54 |
| IIV3-SD: IIV4-Adj | 0.18 | 4.77 | 0.01 | 3.92 | -1.09 | 0.28 | 0.01 | 5.72 |
| IIV3-SD: IIV4-HD | 1.40 | 2.49 | 0.23 | 8.38 | 0.36 | 0.72 | 0.19 | 10.44 |
| IIV3-SD: IIV4-SD | 1.04 | 1.11 | 0.86 | 1.27 | 0.42 | 0.67 | 0.84 | 1.30 |
| IIV3-SD: Placebo | 1.47 | 1.88 | 0.43 | 5.06 | 0.61 | 0.54 | 0.37 | 5.89 |
| IIV3-SD:RIV | 1.45 | 1.53 | 0.63 | 3.33 | 0.88 | 0.38 | 0.57 | 3.69 |
| IIV3-SD:Tdap | 0.18 | 4.87 | 0.01 | 3.96 | -1.09 | 0.28 | 0.01 | 5.79 |
| IIV4-Adj: IIV4-HD | 7.61 | 6.11 | 0.22 | 264.64 | 1.12 | 0.26 | 0.14 | 409.44 |
| IIV4-Adj: IIV4-SD | 5.69 | 4.79 | 0.26 | 122.55 | 1.11 | 0.27 | 0.18 | 178.75 |
| IIV4-Adj: Placebo | 8.00 | 5.40 | 0.29 | 217.74 | 1.23 | 0.22 | 0.20 | 326.88 |
| IIV4-Adj:RIV | 7.90 | 5.05 | 0.33 | 188.95 | 1.28 | 0.20 | 0.22 | 279.17 |
| IIV4-Adj:Tdap | 0.97 | 1.28 | 0.60 | 1.57 | -0.12 | 0.91 | 0.57 | 1.67 |
| IIV4-HD: IIV4-SD | 0.75 | 2.51 | 0.12 | 4.53 | -0.32 | 0.75 | 0.10 | 5.65 |
| IIV4-HD: Placebo | 1.05 | 3.04 | 0.12 | 9.28 | 0.04 | 0.96 | 0.09 | 12.13 |
| IIV4-HD:RIV | 1.04 | 2.74 | 0.14 | 7.48 | 0.04 | 0.97 | 0.11 | 9.53 |
| IIV4-HD:Tdap | 0.13 | 6.22 | 0.00 | 4.58 | -1.13 | 0.26 | 0.00 | 7.12 |
| IIV4-SD: Placebo | 1.41 | 1.90 | 0.40 | 4.92 | 0.53 | 0.59 | 0.34 | 5.74 |
| IIV4-SD:RIV | 1.39 | 1.52 | 0.61 | 3.14 | 0.79 | 0.43 | 0.56 | 3.47 |
| IIV4-SD:Tdap | 0.17 | 4.88 | 0.01 | 3.82 | -1.12 | 0.26 | 0.01 | 5.59 |
| RIV:Placebo | 1.01 | 2.14 | 0.23 | 4.49 | 0.02 | 0.99 | 0.19 | 5.40 |
| Tdap:Placebo | 8.24 | 5.49 | 0.29 | 232.17 | 1.24 | 0.22 | 0.19 | 350.03 |
| RIV:Tdap | 0.12 | 5.14 | 0.00 | 3.05 | -1.28 | 0.20 | 0.00 | 4.52 |
| Common within-network between-study SD | <0.0001 |  |  |  |  |  |  |  |
| I-square | 0.00% |  |  |  |  |  |  |  |
| **Abbreviations-** IIV3: Trivalent **i**nactivated influenza vaccine; IIV4: Quadrivalent **i**nactivated influenza vaccine; Adj: Adjuvanted; SD: Standard dosage; HD: High dosage; RIV: Recombinant influenza vaccine; Tdap: Tetanus, diphtheria, pertussis; OR: Odds ratio; CI: Confidence interval; PI: Prediction interval; se: Standard error | | | | | | | | |

**Figure 4.** Treatment ranking of interventions using P-score for all-cause mortality with original coding of interventions.

| ***Treatment Ranking*** | |
| --- | --- |
| ***Treatment*** | ***P-score*** |
| RIV | 0.75 |
| Placebo | 0.71 |
| IIV4-HD | 0.66 |
| IIV3-HD | 0.60 |
| IIV4-SD | 0.56 |
| IIV3-SD | 0.45 |
| IIV3-Adj | 0.42 |
| IIV4-Adj | 0.18 |
| Tdap | 0.17 |

**Abbreviations-** IIV3: Trivalent **i**nactivated influenza vaccine; IIV4: Quadrivalent **i**nactivated influenza vaccine; Adj: Adjuvanted; SD: Standard dosage; HD: High dosage; RIV: Recombinant influenza vaccine; Tdap: Tetanus, diphtheria, pertussis

**Forest Plots**

**Laboratory-confirmed influenza**

**Figure 5.** Forest Plots of network estimates relative to placebo for laboratory-confirmed influenza with original coding of interventions

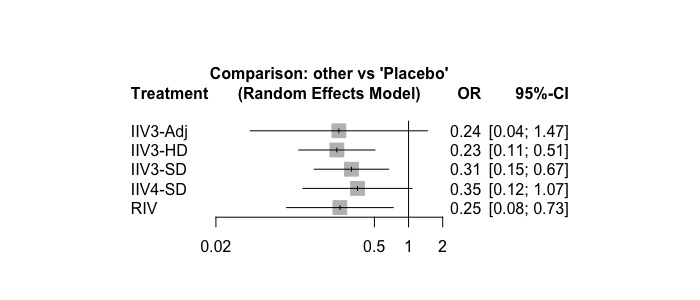

*OR>1 favours placebo; OR<1 favours intervention

**Abbreviations-** IIV3: Trivalent **i**nactivated influenza vaccine; IIV4: Quadrivalent **i**nactivated influenza vaccine; Adj: Adjuvanted; SD: Standard dosage; HD: High dosage; RIV: Recombinant influenza vaccine; OR: Odds ratio; CI: Confidence interval

**Number of Vascular Adverse Events**

**Figure 7.** Forest Plots of network estimates relative to IIV3-SD for number of vascular adverse events with original coding of interventions

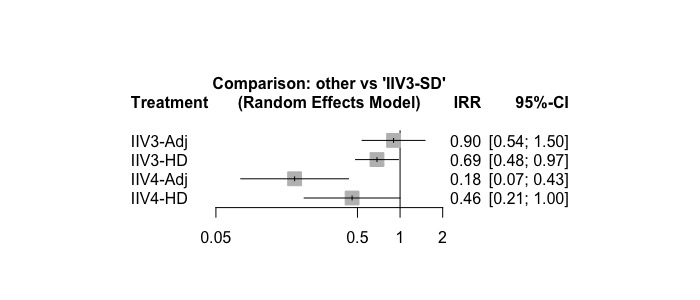

*IRR>1 favours IIV3-SD; IRR<1 favours intervention

**Abbreviations-** IIV3: Trivalent **i**nactivated influenza vaccine; IIV4: Quadrivalent **i**nactivated influenza vaccine; Adj: Adjuvanted; SD: Standard dosage; HD: High dosage; IRR: Incidence rate ratio; CI: Confidence interval

**Outpatient Visits**

**Figure 8.** Forest Plots of network estimates relative to placebo for outpatient visits with original coding of interventions

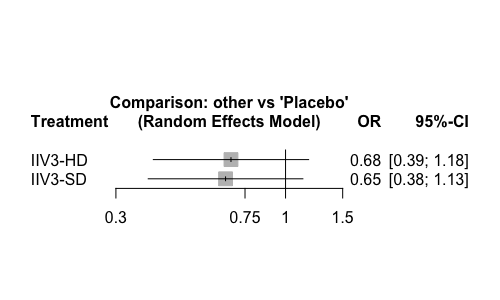

*OR>1 favours placebo; OR<1 favours intervention

**Abbreviations-** IIV3: Trivalent **i**nactivated influenza vaccine; IIV4: Quadrivalent **i**nactivated influenza vaccine; SD: Standard dosage; HD: High dosage; OR: Odds ratio; CI: Confidence interval

**All-cause mortality**

**Figure 9.** Forest Plots of network estimates relative to placebo for all-cause mortality with original coding of interventions

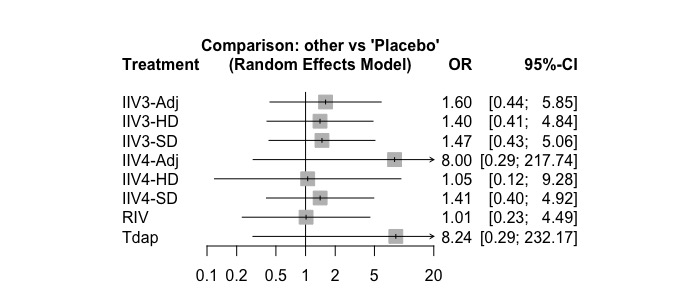

*OR>1 favours placebo; OR<1 favours intervention

**Abbreviations-** IIV3: Trivalent **i**nactivated influenza vaccine; IIV4: Quadrivalent **i**nactivated influenza vaccine; Adj: Adjuvanted; SD: Standard dosage; HD: High dosage; RIV: Recombinant influenza vaccine; Tdap: Tetanus, diphtheria, pertussis; OR: Odds ratio; CI: Confidence interval

**Figure 10.** Rank heat plot across 4 outcomes analyzed in a network meta-analysis

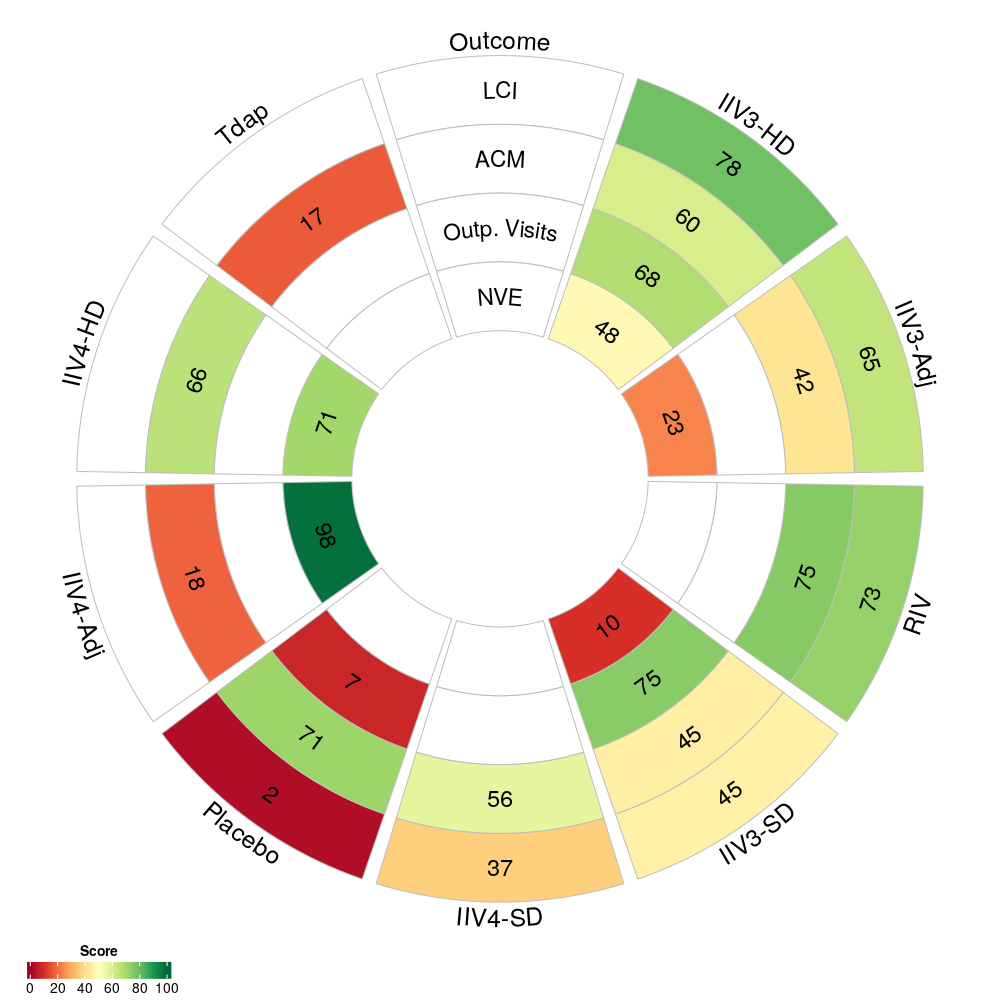

**Abbreviations –** IIV3: Trivalent inactivated influenza vaccine; IIV4: Quadrivalent inactivated influenza vaccine; Adj: Adjuvanted; SD: Standard dosage; HD: High dosage; RIV: Recombinant influenza vaccine; Tdap: Tetanus, diphtheria, pertussis; LCI: Laboratory confirmed influenza; ACM: All-cause mortality; Outp. Visit: Outpatient Visit; NVE: Number of Vascular Adverse Events

### **Appendix 18: Results on Secondary outcomes analyzed in a pairwise meta-analysis only**

Pairwise meta-analysis suggested that IIV3-HD was comparable to IIV3-SD (2 RCTs ^15, 20^, 41,141 participants; OR 0.94, 95%CI [0.74 to 1.19], I^2^=0%, τ=0.00; moderate certainty of evidence) for the prevention of ER visits for ILI. IIV3-HD was superior to IIV3-SD for preventing hospitalizations for ILI (2 RCTs ^15, 20^, 41,141 participants; OR 0.72, 95%CI [0.57 to 0.92], I^2^=0%, τ=0.00; high certainty of evidence) and hospitalizations for ARI (2 RCTs ^20, 23^, 84,991 participants; OR 0.87, 95%CI [0.79 to 0.95], I^2^=0%, τ=0.00; moderate certainty of evidence). For IIV3-HD versus IIV3-SD, inconclusive results were identified for the prevention of inpatient hospitalizations (3 RCTs ^13, 20, 23, 50^, 38,816 participants; OR 0.76, 95%CI [0.40 to 1.42], I^2^=86%, τ=0.26; low certainty of evidence), which were in agreement when the analysis was restricted to low ROB studies (2 RCTs ^20, 26, 50^, 34,940 participants; OR 0.76, 95%CI [0.52 to 1.13], I^2^=93%, τ=0.28; moderate certainty of evidence). Precision increased for IIV3-HD versus IIV3-SD when restricting to studies with participants over 80 years of age (2 RCTs ^20, 26^, 35,859 participants; OR 0.92, 95%CI [0.86 to 0.99], I^2^=0%, τ=0.00; moderate certainty of evidence; Table 5).

### **Appendix 19: Methods for lab-confirmed influenza diagnoses**

| Author, Year | Source of Diagnosis | Influenza Diagnosis |
| --- | --- | --- |
| DiazGranados, 2013^15^ | Determined by Investigator | “If a participant met the criteria for ILI, the study site was to arrange for a nasopharyngeal (NP) swab to be taken within 5 days of ILI episode onset. NP samples underwent tissue culture and molecular testing (Polymerase-Chain-Reaction [PCR]-based assays) for laboratory confirmation of influenza.” |
| DiazGranados, 2014^20^ | Determined by Investigator | “If a participant met the criteria for any respiratory illness, staff members at the study site were to collect a nasopharyngeal swab within 5 days after onset of the illness. Laboratory confirmation of influenza in nasopharyngeal swabs was accomplished by a positive result on culture, a polymerase-chain-reaction (PCR) assay, or both.” |
| Beran, 2021^32^ | Determined by Investigator | “By two commercial lab tests: RT-PCR; viral expansion in Madin Darby Canine Kidney cell culture. Viral strain antigenic typing was determined by means of HA inhibition or microneutralization assays.” |
| Belongia, 2020^29^ | Determined by Investigator | “Nasal and oropharyngeal swabs were combined and tested for influenza type and subtype using real time reverse transcription polymerase chain reaction (RT-PCR) at the Marshfield Clinic Research Institute using CDC primers and probes.” |
| Dunkle, 2017^24^ | NR | NR |
| Keitel, 2010^14^ | Determined by Investigator | “By cell culture and then treatment with virus A and virus B strain specific fluorescent antibodies." “Virus isolation was carried out using standard cell culture methods in primary rhesus monkey kidney (PRhMK) cells. Briefly, clinical samples were absorbed onto PRhMK monolayers for approximately 60 min, and then monitored daily for cytopathic effect up to 14 days. If a cytopathic effect score of 2+ was reached, the presence (or absence) of either influenza A or B was determined by fluorescent antibody testing with monoclonal antibodies.” |
| Wongsurakiat, 2004^11^ | Determined by Investigator | “A fourfold HI titer increase in convalescent serum compared to acute serum with a titer of 40 and/or demonstration of influenza antigen with or without positive culture finding was considered as meeting the criteria for influenza virus infection.” |
| Rudenko, 2001^10^ | Determined by Investigator | “The presence of influenza-like illness in association with isolation of influenza virus, or a >= 4 fold rise in serum HI antibody between acute- and convalescent- phase serum samples, or both.” |
| Teh, 2021^34^ | Determined by Investigator | “Patients who reported ILI were asked to present to their general practitioner or study center for medical review and multiplex polymerase chain reaction (PCR) testing for respiratory viruses including influenza.” |
| Loeb, 2020^28^ | Determined by Investigator | “Influenza illness was documented by PCR detection of influenza during an ARI or seroconversion (4-fold rise in antibody titers) in association with an ARI.” |
| McLean, 2021^49^ | Determined by Investigator | “Respiratory samples were tested for influenza using reverse transcription polymerase chain reaction (RT-PCR) with primers and probes provided by CDC at MCRI.” |
| Abbreviations: ARI = Acute respiratory infection; CDC = Centers for Disease Control and Prevention ; HA = hemagglutinin; ILI = Influenza-Like Illness; MCRI = Marshfield Clinic Research Institute; RT-PCR = real time reverse transcription polymerase chain reaction; NR = not reported. | | |

### **Appendix 20: Confidence in Network Meta-Analysis (CINeMA) Assessments**

CINeMA assessments were conducted using the CINeMA tool.^6^ Using the tool, all comparisons are graded across six domains (i.e., within-study bias, reporting bias, indirectness, imprecision, heterogeneity, and incoherence). Within study bias is obtained from the overall risk of bias assessments where were conducted using the Cochrane RoB2 tool or the RoB2-CRT for cluster trials. Reporting bias was informed by funnel plots where possible, and otherwise was set to “some concern”. A overall confidence rating for each comparison is provided. The rating starts off at “High” and may be downgraded if there are concerns in any of the domains. For our purposes, to be conservative, we treated “some concerns” and “serious concerns” equally. That is, the overall confidence rating was downgraded by one level for each domain (except within-study bias since there was limited information) that had “some concern” or “serious concern”, up to a overall confidence rating of very low. CINeMA assessments are provided for two different minimally important differences, namely, a null effect, and a literature/expert informed important effect. Both results are presented, with minor differences in the overall assessments.

**Laboratory-confirmed Influenza**

**Figure 1.** Contribution Matrix plot illustrating the risk of bias for each treatment comparison for laboratory-confirmed influenza

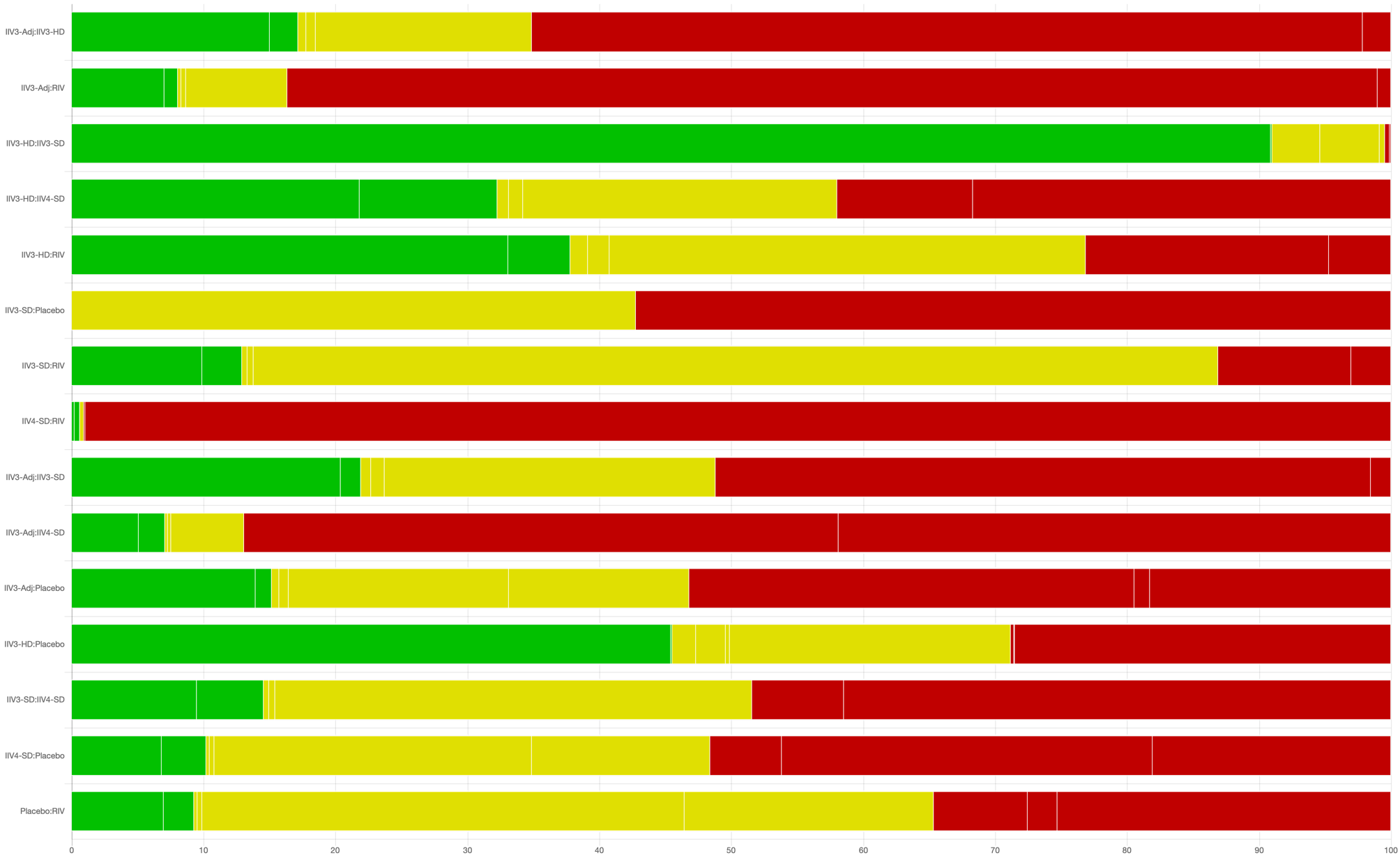

**Abbreviations-** IIV3: Trivalent **i**nactivated influenza vaccine; IIV4: Quadrivalent **i**nactivated influenza vaccine; Adj: Adjuvanted; SD: Standard dosage; HD: High dosage; RIV: Recombinant influenza vaccine

**Table 1.** CINeMA assessment providing credibility for each treatment comparison for laboratory-confirmed influenza under null minimally important differences.

| **Treatment**  **Comparison** | **Number of studies** | **Within-study bias** | **Reporting bias** | **Indirectness** | **Imprecision** | **Heterogeneity** | **Incoherence** | **Confidence rating** | **Reason(s) for downgrading** |
| --- | --- | --- | --- | --- | --- | --- | --- | --- | --- |
| **Direct Evidence** | | | | | | | | | |
| IIV3-Adj:IIV3-HD | 1 | Some concerns | Some concerns | No concerns | Major concerns | No concerns | No concerns | Low | ["Within-study bias","Imprecision"] |
| IIV3-Adj:RIV | 1 | Major concerns | Some concerns | No concerns | Major concerns | No concerns | No concerns | Low | ["Within-study bias","Imprecision"] |
| IIV3-HD:IIV3-SD | 3 | No concerns | Some concerns | No concerns | No concerns | No concerns | No concerns | High | Not applicable |
| IIV3-HD:IIV4-SD | 1 | Some concerns | Some concerns | No concerns | Major concerns | No concerns | No concerns | Low | ["Within-study bias","Imprecision"] |
| IIV3-HD:RIV | 1 | Some concerns | Some concerns | No concerns | Major concerns | No concerns | No concerns | Low | ["Within-study bias","Imprecision"] |
| IIV3-SD:Placebo | 2 | Major concerns | Some concerns | Some concerns | No concerns | No concerns | No concerns | Low | ["Within-study bias","Indirectness"] |
| IIV3-SD:RIV | 1 | Some concerns | Some concerns | No concerns | Major concerns | No concerns | No concerns | Low | ["Within-study bias","Imprecision"] |
| IIV4-SD:RIV | 1 | Major concerns | Some concerns | No concerns | No concerns | Major concerns | No concerns | Low | ["Within-study bias","Heterogeneity"] |
| **Indirect Evidence** | | | | | | | | | |
| IIV3-Adj:IIV3-SD | 0 | Some concerns | Some concerns | No concerns | Major concerns | No concerns | No concerns | Low | ["Within-study bias","Imprecision"] |
| IIV3-Adj:IIV4-SD | 0 | Major concerns | Some concerns | No concerns | Major concerns | No concerns | No concerns | Low | ["Within-study bias","Imprecision"] |
| IIV3-Adj:Placebo | 0 | Some concerns | Some concerns | No concerns | Major concerns | No concerns | No concerns | Low | ["Within-study bias","Imprecision"] |
| IIV3-HD:Placebo | 0 | Some concerns | Some concerns | Some concerns | No concerns | No concerns | No concerns | Low | ["Within-study bias","Indirectness"] |
| IIV3-SD:IIV4-SD | 0 | Some concerns | Some concerns | No concerns | Major concerns | No concerns | No concerns | Low | ["Within-study bias","Imprecision"] |
| IIV4-SD:Placebo | 0 | Some concerns | Some concerns | No concerns | Major concerns | No concerns | No concerns | Low | ["Within-study bias","Imprecision"] |
| Placebo:RIV | 0 | Some concerns | Some concerns | No concerns | No concerns | Major concerns | No concerns | Low | ["Within-study bias","Heterogeneity"] |
| **Abbreviations-** IIV3: Trivalent **i**nactivated influenza vaccine; IIV4: Quadrivalent **i**nactivated influenza vaccine; Adj: Adjuvanted; SD: Standard dosage; HD: High dosage; RIV: Recombinant influenza vaccine | | | | | | | | | |

**Number of Vascular Adverse Events**

**Figure 2.** Contribution Matrix plot illustrating the risk of bias for each treatment comparison for number of vascular adverse events

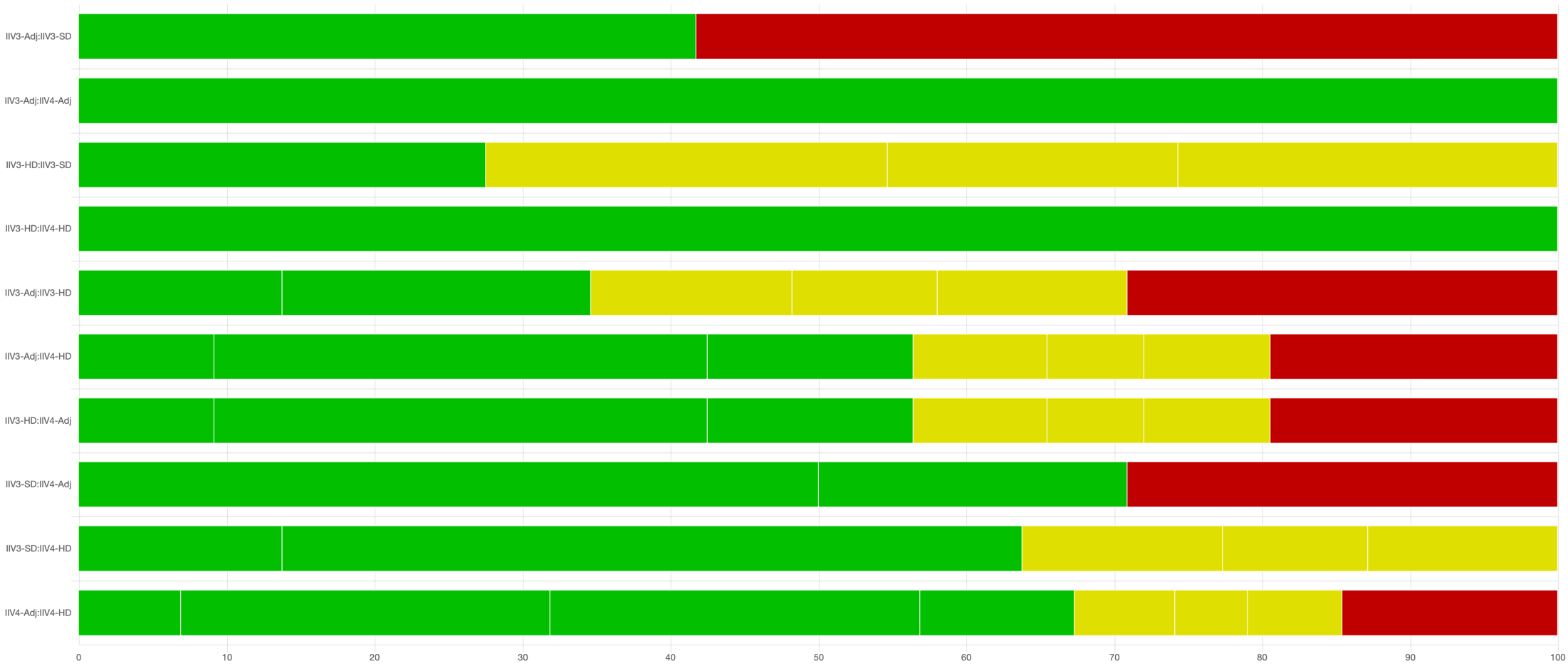

**Abbreviations-** IIV3: Trivalent **i**nactivated influenza vaccine; IIV4: Quadrivalent **i**nactivated influenza vaccine; Adj: Adjuvanted; SD: Standard dosage; HD: High dosage

**Table 2.** CINeMA assessment providing credibility for each treatment comparison for number of vascular adverse events under null minimally important differences.

| **Treatment Comparison** | **Number of studies** | **Within-study bias** | **Reporting bias** | **Indirectness** | **Imprecision** | **Heterogeneity** | **Incoherence** | **Confidence rating** | **Reason(s) for downgrading** |
| --- | --- | --- | --- | --- | --- | --- | --- | --- | --- |
| **Direct Evidence** | | | | | | | | | |
| IIV3-Adj:IIV3-SD | 2 | Some concerns | Some concerns | No concerns | Major concerns | No concerns | Major concerns | Very low | ["Within-study bias","Imprecision","Incoherence"] |
| IIV3-Adj:IIV4-Adj | 1 | No concerns | Some concerns | No concerns | No concerns | No concerns | Major concerns | Moderate | ["Incoherence"] |
| IIV3-HD:IIV3-SD | 4 | Some concerns | Some concerns | No concerns | No concerns | Major concerns | Major concerns | Very low | ["Within-study bias","Heterogeneity","Incoherence"] |
| IIV3-HD:IIV4-HD | 1 | No concerns | Some concerns | No concerns | Major concerns | No concerns | Major concerns | Low | ["Imprecision","Incoherence"] |
| **Indirect Evidence** | | | | | | | | | |
| IIV3-Adj:IIV3-HD | 0 | Some concerns | Some concerns | No concerns | Major concerns | No concerns | Major concerns | Very low | ["Within-study bias","Imprecision","Incoherence"] |
| IIV3-Adj:IIV4-HD | 0 | Some concerns | Some concerns | No concerns | Major concerns | No concerns | Major concerns | Very low | ["Within-study bias","Imprecision","Incoherence"] |
| IIV3-HD:IIV4-Adj | 0 | Some concerns | Some concerns | No concerns | No concerns | Major concerns | Major concerns | Very low | ["Within-study bias","Heterogeneity","Incoherence"] |
| IIV3-SD:IIV4-Adj | 0 | Some concerns | Some concerns | No concerns | No concerns | Major concerns | Major concerns | Very low | ["Within-study bias","Heterogeneity","Incoherence"] |
| IIV3-SD:IIV4-HD | 0 | No concerns | Some concerns | No concerns | Major concerns | No concerns | Major concerns | Low | ["Imprecision","Incoherence"] |
| IIV4-Adj:IIV4-HD | 0 | No concerns | Some concerns | No concerns | Major concerns | No concerns | Major concerns | Low | ["Imprecision","Incoherence"] |
| **Abbreviations-** IIV3: Trivalent **i**nactivated influenza vaccine; IIV4: Quadrivalent **i**nactivated influenza vaccine; Adj: Adjuvanted; SD: Standard dosage; HD: High dosage | | | | | | | | | |

**Outpatient Visits**

**Figure 3.** Contribution Matrix plot illustrating the risk of bias for each treatment comparison for outpatient visits

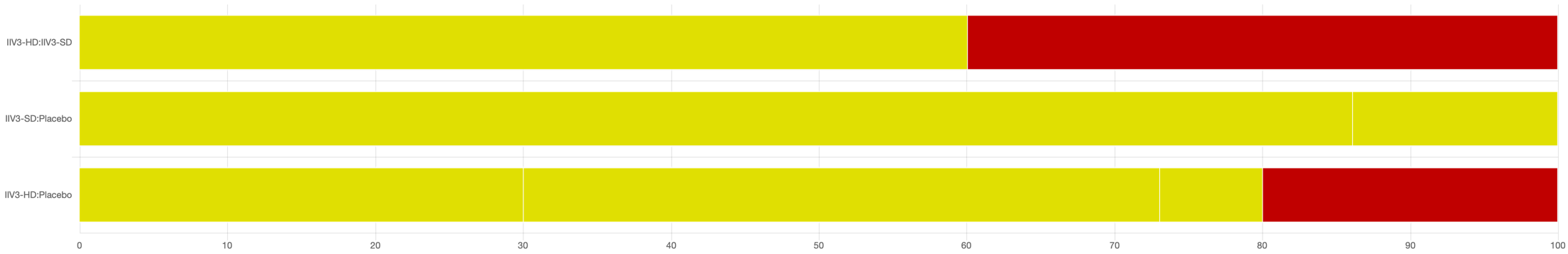

**Abbreviations-** IIV3: Trivalent **i**nactivated influenza vaccine; SD: Standard dosage; HD: High dosage

**Table 3.** CINeMA assessment providing credibility for each treatment comparison for outpatient visits under null minimally important differences.

| **Treatment Comparison** | **Number of studies** | **Within-study bias** | **Reporting bias** | **Indirectness** | **Imprecision** | **Heterogeneity** | **Incoherence** | **Confidence rating** | **Reason(s) for downgrading** |
| --- | --- | --- | --- | --- | --- | --- | --- | --- | --- |
| **Direct Evidence** | | | | | | | | | |
| IIV3-HD:IIV3-SD | 2 | Some concerns | Some concerns | No concerns | Major concerns | No concerns | Major concerns | Very low | ["Within-study bias","Imprecision","Incoherence"] |
| IIV3-SD:Placebo | 2 | Some concerns | Some concerns | No concerns | Major concerns | No concerns | Major concerns | Very low | ["Within-study bias","Imprecision","Incoherence"] |
| **Indirect Evidence** | | | | | | | | | |
| IIV3-HD:Placebo | 0 | Some concerns | Some concerns | No concerns | Major concerns | No concerns | Major concerns | Very low | ["Within-study bias","Imprecision","Incoherence"] |
| **Abbreviations-** IIV3: Trivalent **i**nactivated influenza vaccine; SD: Standard dosage; HD: High dosage | | | | | | | | | |

**All-cause mortality**

**Figure 4.** Contribution Matrix plot illustrating the risk of bias for each treatment comparison for all-cause mortality

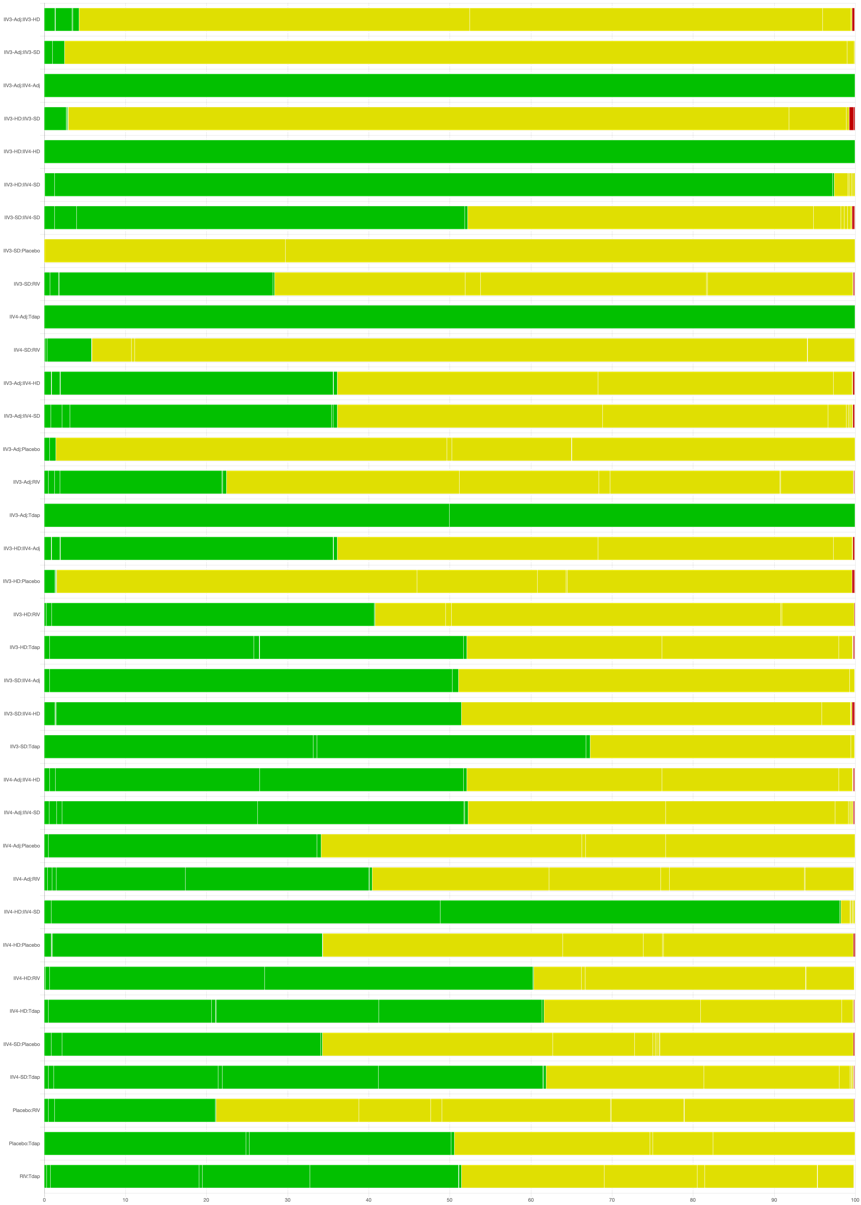

**Abbreviations-** IIV3: Trivalent **i**nactivated influenza vaccine; IIV4: Quadrivalent **i**nactivated influenza vaccine; Adj: Adjuvanted; SD: Standard dosage; HD: High dosage; RIV: Recombinant influenza vaccine; Tdap: Tetanus, diphtheria, pertussis

**Table 4.** CINeMA assessment providing credibility for each treatment comparison for all-cause mortality under null minimally important differences.

| **Treatment Comparison** | **Number of studies** | **Within-study bias** | **Reporting bias** | **Indirectness** | **Imprecision** | **Heterogeneity** | **Incoherence** | **Confidence rating** | **Reason(s) for downgrading** |
| --- | --- | --- | --- | --- | --- | --- | --- | --- | --- |
| **Direct Evidence** | | | | | | | | | |
| IIV3-Adj:IIV3-HD | 1 | Some concerns | No concerns | No concerns | Major concerns | No concerns | No concerns | Low | ["Within-study bias","Imprecision"] |
| IIV3-Adj:IIV3-SD | 2 | Some concerns | No concerns | No concerns | Major concerns | No concerns | No concerns | Low | ["Within-study bias","Imprecision"] |
| IIV3-Adj:IIV4-Adj | 1 | No concerns | No concerns | No concerns | Major concerns | No concerns | No concerns | Moderate | ["Imprecision"] |
| IIV3-HD:IIV3-SD | 6 | Some concerns | No concerns | No concerns | Major concerns | No concerns | No concerns | Low | ["Within-study bias","Imprecision"] |
| IIV3-HD:IIV4-HD | 1 | No concerns | No concerns | No concerns | Major concerns | No concerns | No concerns | Moderate | ["Imprecision"] |
| IIV3-HD:IIV4-SD | 1 | No concerns | No concerns | Some concerns | Major concerns | No concerns | No concerns | Low | ["Indirectness","Imprecision"] |
| IIV3-SD:IIV4-SD | 3 | No concerns | No concerns | No concerns | Major concerns | No concerns | No concerns | Moderate | ["Imprecision"] |
| IIV3-SD:Placebo | 2 | Some concerns | No concerns | Some concerns | Major concerns | No concerns | No concerns | Very low | ["Within-study bias","Indirectness","Imprecision"] |
| IIV3-SD:RIV | 1 | Some concerns | No concerns | No concerns | Major concerns | No concerns | No concerns | Low | ["Within-study bias","Imprecision"] |
| IIV4-Adj:Tdap | 1 | No concerns | No concerns | No concerns | Major concerns | No concerns | No concerns | Moderate | ["Imprecision"] |
| IIV4-SD:RIV | 1 | Some concerns | No concerns | No concerns | Major concerns | No concerns | No concerns | Low | ["Within-study bias","Imprecision"] |
| **Indirect Evidence** | | | | | | | | | |
| IIV3-Adj:IIV4-HD | 0 | Some concerns | No concerns | No concerns | Major concerns | No concerns | No concerns | Low | ["Within-study bias","Imprecision"] |
| IIV3-Adj:IIV4-SD | 0 | Some concerns | No concerns | No concerns | Major concerns | No concerns | No concerns | Low | ["Within-study bias","Imprecision"] |
| IIV3-Adj:Placebo | 0 | Some concerns | No concerns | No concerns | Major concerns | No concerns | No concerns | Low | ["Within-study bias","Imprecision"] |
| IIV3-Adj:RIV | 0 | Some concerns | No concerns | No concerns | Major concerns | No concerns | No concerns | Low | ["Within-study bias","Imprecision"] |
| IIV3-Adj:Tdap | 0 | No concerns | No concerns | No concerns | Major concerns | No concerns | No concerns | Moderate | ["Imprecision"] |
| IIV3-HD:IIV4-Adj | 0 | Some concerns | No concerns | No concerns | Major concerns | No concerns | No concerns | Low | ["Within-study bias","Imprecision"] |
| IIV3-HD:Placebo | 0 | Some concerns | No concerns | No concerns | Major concerns | No concerns | No concerns | Low | ["Within-study bias","Imprecision"] |
| IIV3-HD:RIV | 0 | Some concerns | No concerns | No concerns | Major concerns | No concerns | No concerns | Low | ["Within-study bias","Imprecision"] |
| IIV3-HD:Tdap | 0 | No concerns | No concerns | No concerns | Major concerns | No concerns | No concerns | Moderate | ["Imprecision"] |
| IIV3-SD:IIV4-Adj | 0 | No concerns | No concerns | No concerns | Major concerns | No concerns | No concerns | Moderate | ["Imprecision"] |
| IIV3-SD:IIV4-HD | 0 | No concerns | No concerns | No concerns | Major concerns | No concerns | No concerns | Moderate | ["Imprecision"] |
| IIV3-SD:Tdap | 0 | No concerns | No concerns | No concerns | Major concerns | No concerns | No concerns | Moderate | ["Imprecision"] |
| IIV4-Adj:IIV4-HD | 0 | No concerns | No concerns | No concerns | Major concerns | No concerns | No concerns | Moderate | ["Imprecision"] |
| IIV4-Adj:IIV4-SD | 0 | No concerns | No concerns | No concerns | Major concerns | No concerns | No concerns | Moderate | ["Imprecision"] |
| IIV4-Adj:Placebo | 0 | Some concerns | No concerns | No concerns | Major concerns | No concerns | No concerns | Low | ["Within-study bias","Imprecision"] |
| IIV4-Adj:RIV | 0 | Some concerns | No concerns | No concerns | Major concerns | No concerns | No concerns | Low | ["Within-study bias","Imprecision"] |
| IIV4-HD:IIV4-SD | 0 | No concerns | No concerns | No concerns | Major concerns | No concerns | No concerns | Moderate | ["Imprecision"] |
| IIV4-HD:Placebo | 0 | Some concerns | No concerns | No concerns | Major concerns | No concerns | No concerns | Low | ["Within-study bias","Imprecision"] |
| IIV4-HD:RIV | 0 | No concerns | No concerns | No concerns | Major concerns | No concerns | No concerns | Moderate | ["Imprecision"] |
| IIV4-HD:Tdap | 0 | No concerns | No concerns | No concerns | Major concerns | No concerns | No concerns | Moderate | ["Imprecision"] |
| IIV4-SD:Placebo | 0 | Some concerns | No concerns | Some concerns | Major concerns | No concerns | No concerns | Very low | ["Within-study bias","Indirectness","Imprecision"] |
| IIV4-SD:Tdap | 0 | No concerns | No concerns | No concerns | Major concerns | No concerns | No concerns | Moderate | ["Imprecision"] |
| Placebo:RIV | 0 | Some concerns | No concerns | No concerns | Major concerns | No concerns | No concerns | Low | ["Within-study bias","Imprecision"] |
| Placebo:Tdap | 0 | No concerns | No concerns | No concerns | Major concerns | No concerns | No concerns | Low | ["Imprecision"] |
| RIV:Tdap | 0 | No concerns | No concerns | No concerns | Major concerns | No concerns | No concerns | Low | ["Imprecision"] |
| **Abbreviations-** IIV3: Trivalent **i**nactivated influenza vaccine; IIV4: Quadrivalent **i**nactivated influenza vaccine; Adj: Adjuvanted; SD: Standard dosage; HD: High dosage; RIV: Recombinant influenza vaccine; Tdap: Tetanus, diphtheria, pertussis | | | | | | | | | |

### **Appendix 21: Grading of Recommendations, Assessment, Development, and Evaluations (GRADE) Assessments**

**Comparison 1: High Dose Trivalent Vaccine compared to Standard Dose Trivalent Vaccine for preventing influenza.**

| **Certainty assessment** | | | | | | | **Number of participants** | | **Effect** | | **Certainty** | **Importance** |
| --- | --- | --- | --- | --- | --- | --- | --- | --- | --- | --- | --- | --- |
| **Number of studies** | **Study design** | **Risk of bias** | **Inconsistency** | **Indirectness** | **Imprecision** | **Other considerations** | **High Dose Trivalent Vaccine** | **Standard Dose Trivalent Vaccine** | **Relative (95% CI)** | **Absolute (95% CI)** |  |  |
| **Influenza-like illness (ILI) cases** | | | | | | | | | | | | |
| 2 | RCTs | Serious^a^ | Not serious | Not serious | Serious^b^ | None | 10/614 (1.6%) | 17/240 (7.1%) | **OR 0.39** (0.15 to 1.02) | **42 fewer per 1,000** (from 60 fewer to 1 more) | ⨁⨁◯◯ Low |  |
| **Hospitalization due to acute respiratory infection (ARI)** | | | | | | | | | | | | |
| 2 | RCTs | Serious^c^ | Not serious | Not serious | Not serious | None | 890/42629 (2.1%) | 1013/42362 (2.4%) | **OR 0.87** (0.79 to 0.95) | **3 fewer per 1,000** (from 5 fewer to 1 fewer) | ⨁⨁⨁◯ Moderate |  |
| **ER Visit due to Influenza-like illness (ILI)** | | | | | | | | | | | | |
| 2 | RCTs | Not serious^d^ | not serious^e^ | Not serious | Serious^f^ | None | 149/22098 (0.7%) | 138/19043 (0.7%) | **OR 0.94** (0.74 to 1.19) | **0 fewer per 1,000** (from 2 fewer to 1 more) | ⨁⨁⨁◯ Moderate |  |
| **Hospitalization due to Influenza-like illness (ILI)** | | | | | | | | | | | | |
| 2 | RCTs | Not serious^g^ | Not serious | Not serious | Not serious | None | 127/22098 (0.6%) | 155/19043 (0.8%) | **OR 0.72** (0.57 to 0.92) | **2 fewer per 1,000** (from 3 fewer to 1 fewer) | ⨁⨁⨁⨁ High |  |
| **Inpatient Hospitalization (any cause)** | | | | | | | | | | | | |
| 3 | RCTs | Serious^h^ | Not serious^i^ | Not serious | Serious^j^ | None | 1728/20039 (8.6%) | 1945/18777 (10.4%) | **OR 0.76** (0.40 to 1.42) | **23 fewer per 1,000** (from 59 fewer to 37 more) | ⨁⨁◯◯ Low |  |

**Abbreviations –** RCTs: Randomized controlled trials; CI: Confidence interval; OR**:** Odds ratio

**Explanations**

a. Study that carried large weight for the overall effect estimated rated as some concerns due to bias arising from the randomization process and bias in selection of the reported result. Another study that carried small weight for the overall effect estimated rated as high risk of bias due to missing outcome data

b. Serious imprecision. 95% CI is consistent with the possibility for important benefit and large harm, including only 27 events in total.

c. Study that carried large weight for the overall effect estimated rated as high risk of bias due to some concerns in the timing of identification or recruitment of participants in a cluster trial, bias due to deviations from intended interventions and bias due to missing outcome data.

d. Study that carried large weight for the overall effect estimated was rated as low risk of bias, and the other study, that carried small weight for the overall effect estimated, was rated as high risk of bias due to deviations from intended interventions.

e. I^2^ value is 87%, suggesting some heterogeneity; We could not find the reasons for this high heterogeneity

f. Serious imprecision. 95% CI is consistent with the possibility for important benefit and large harm, including only 287 events in total

g. Study that carried large weight for the overall effect estimated rated as some concerns due to bias in selection of the reported result. No rating down due to RoB

h. Study that carried large weight for the overall effect estimated rated as high risk of bias due to missing outcome data

i. I^2^=87%, suggesting substantial heterogeneity, but of questionable clinical importance, because the two studies with more weight in the meta-analysis show a significant effect. We did not rate down due to inconsistency

j. Serious imprecision. 95% CI is consistent with the possibility for important benefit and large harm.

**Comparison 2: Standard Dose Trivalent vaccine compared to Placebo for preventing influenza.**

| **Certainty assessment** | | | | | | | **Number of participants** | | **Effect** | | **Certainty** | **Importance** |
| --- | --- | --- | --- | --- | --- | --- | --- | --- | --- | --- | --- | --- |
| **Number of studies** | **Study design** | **Risk of bias** | **Inconsistency** | **Indirectness** | **Imprecision** | **Other considerations** | **Standard Dose Trivalent vaccine** | **Placebo** | **Relative (95% CI)** | **Absolute (95% CI)** |  |  |
| **Influenza-like illness cases** | | | | | | | | | | | | |
| 2 | RCTs | Not serious | Not serious | Not serious | Not serious | None | 5065/22098 (22.9%) | 4489/19043 (23.6%) | **OR 0.98** (0.93 to 1.02) | **4 fewer per 1,000** (from 13 fewer to 4 more) | ⨁⨁⨁⨁ High |  |

**Abbreviations –** RCTs: Randomized controlled trials; CI: Confidence interval; OR**:** Odds rati

### **Appendix 22: Pairwise Meta-Analyses**

**Table 1.** Pairwise meta-analysis results for each outcome

| **Primary Outcomes** | | | | | | | | | | | | | | | |
| --- | --- | --- | --- | --- | --- | --- | --- | --- | --- | --- | --- | --- | --- | --- | --- |
| ***Outcome*** | ***Comparison*** | ***Number of studies*** | ***Number of participants*** | ***OR*** | ***lower CI*** | ***upper CI*** | ***lower PI*** | ***upper PI*** | ***tau*** | ***tau^2^ lower CI*** | ***tau^2^ upper CI*** | ***Q*** | ***Q df*** | ***Q P-value*** | ***I^2^*** |
| LCI | IIV3-HD : IIV3-SD | 3 | 41753 | 0.75 | 0.63 | 0.88 | 0.25 | 2.19 | 0.00 | 0.00 | 2.72 | 1.00 | 2 | 0.61 | 0% |
|  | IIV3-SD : Placebo | 2 | 330 | 0.31 | 0.13 | 0.75 | . | . | 0.33 | . | . | 1.4 | 1 | 0.25 | 26% |
| ILI | IIV3-HD : IIV3-SD | 2 | 41141 | 0.98 | 0.93 | 1.02 | . | . | 0.00 | . | . | 0.14 | 1 | 0.71 | 0% |
|  | IIV3-SD : Placebo | 2 | 854 | 0.39 | 0.15 | 1.02 | . | . | 0.21 | . | . | 1.08 | 1 | 0.3 | 7.6% |
| **Secondary Outcomes** | | | | | | | | | | | | | | | |
| ***Outcome*** | ***Comparison*** | ***Number of studies*** | ***Number of participants*** | ***OR*** | ***lower CI*** | ***upper CI*** | ***lower PI*** | ***upper PI*** | ***tau^2^*** | ***tau^2^ lower CI*** | ***tau2 upper CI*** | ***Q*** | ***Q df*** | ***Q P-value*** | ***I^2^*** |
| ER Visit for ILI | IIV3-HD : IIV3-SD | 2 | 41141 | 0.94 | 0.74 | 1.19 | . | . | 0.00 | . | . | 0.20 | 1 | 0.66 | 0% |
| Hospitalization for ILI | IIV3-HD : IIV3-SD | 2 | 41141 | 0.72 | 0.57 | 0.92 | . | . | 0.00 | . | . | 0.40 | 1 | 0.52 | 0% |
| Hospitalization for ARI | IIV3-HD : IIV3-SD | 2 | 84991 | 0.87 | 0.79 | 0.95 | . | . | 0.00 | . | . | 0.00 | 1 | 0.98 | 0% |
| Number of Vascular Events | IIV3-Adj : IIV3-SD | 2 | 7577 | 0.83 | 0.54 | 1.27 | . | . | 0.00 | . | . | 0.00 | 1 | 0.89 | 0% |
|  | IIV3-HD : IIV3-SD | 4 | 45656 | 0.74 | 0.43 | 1.29 | 0.21 | 2.58 | 0.23 | 0.00 | 2.20 | 4.90 | 3 | 0.18 | 39% |
| Inpatient Hospitalization (any cause) | IIV3-HD : IIV3-SD | 3 | 38816 | 0.76 | 0.40 | 1.42 | 0.01 | 50.27 | 0.27 | 0.01 | 3.22 | 14.00 | 2 | 0.00 | 86% |
| Outpatient Visit | IIV3-HD : IIV3-SD | 2 | 41141 | 1.04 | 0.99 | 1.09 | . | . | 0 | . | . | 0.80 | 1 | 0.38 | 0% |
|  | IIV3-SD : Placebo | 2 | 814 | 0.40 | 0.07 | 2.14 | . | . | 1.09 | . | . | 4.30 | 1 | 0.04 | 77% |
| All-cause Death | IIV3-Adj : IIV3-SD | 2 | 7577 | 1.09 | 0.73 | 1.62 | . | . | 0.00 | . | . | 0.50 | 1 | 0.47 | 0% |
|  | IIV3-HD : IIV3-SD | 6 | 101187 | 0.97 | 0.92 | 1.02 | 0.90 | 1.04 | 0.00 | 0.00 | 0.02 | 1.20 | 5 | 0.95 | 0% |
|  | IIV4-SD : IIV3-SD | 3 | 3864 | 1.08 | 0.38 | 3.09 | 0.00 | 970.40 | 0.00 | 0.00 | 15.12 | 0.70 | 2 | 0.70 | 0% |
|  | IIV3-SD : Placebo | 2 | 854 | 1.47 | 0.43 | 5.06 | . | . | 0.00 | . | . | 0.20 | 1 | 0.66 | 0% |
| Influenza-related death | IIV3-Adj:IIV3-SD | 1 | 6961 | 0.75 | 0.17 | 3.36 |  |  |  |  |  |  |  |  |  |
|  | IIV4-SD:IIV3-SD | 1 | 1741 | 3.01 | 0.12 | 73.19 |  |  |  |  |  |  |  |  |  |
| **Abbreviations-** IIV3: Trivalent **i**nactivated influenza vaccine; IIV4: Quadrivalent **i**nactivated influenza vaccine; Adj: Adjuvanted; SD: Standard dosage; HD: High dosage; ARI: Acute respiratory infection; LC-ARI: Laboratory-confirmed acute respiratory infection; ER: Emergency room; LCI: Laboratory-confirmed influenza; ILI: Influenza-like illness; OR: Odds ratio; CI: Confidence interval; PI: Prediction interval; df: Degrees of freedom | | | | | | | | | | | | | | | |

**Laboratory-confirmed Influenza (LCI)**

**Figure 1.** Forest plot of pairwise meta-analysis comparing IIV3-HD (intervention) vs. IIV3-SD (control)

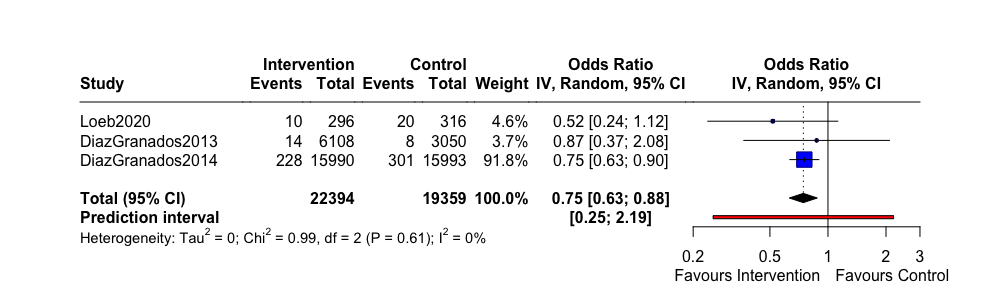

**Abbreviations-** IIV3: Trivalent **i**nactivated influenza vaccine; SD: Standard dosage; HD: High dosage; CI: Confidence interval

**Figure 2.** Forest plot of pairwise meta-analysis comparing IIV3-SD (intervention) vs. Placebo (control)

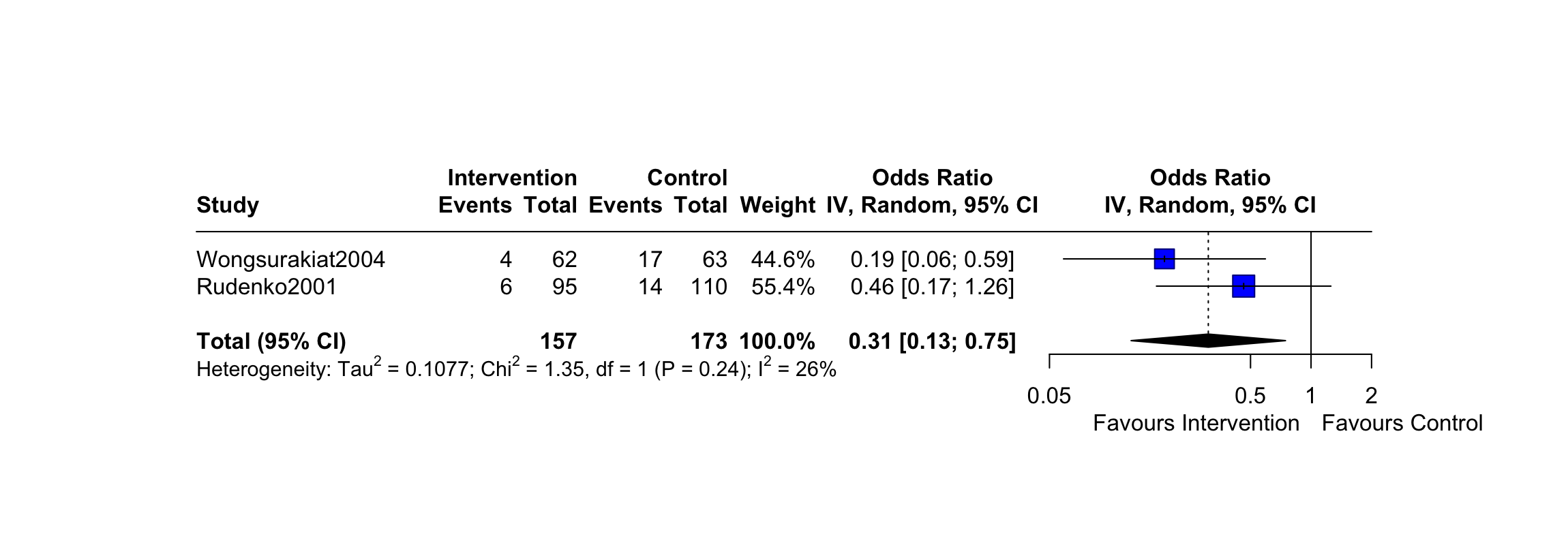

**Abbreviations-** IIV3: Trivalent **i**nactivated influenza vaccine; SD: Standard dosage; CI: Confidence interva

**Table 2**. Vaccine efficacy from pairwise meta-analysis results for laboratory-confirmed influenza with original coding of interventions

| ***Comparison*** | ***OR*** | ***RR*** | ***VE*** |
| --- | --- | --- | --- |
| IIV3-HD : IIV3-SD | 0.75 (0.63,0.88) | 0.79 (0.68,0.90) | 21.08 (9.65,32.00) |
| IIV3-SD : Placebo | 0.31 (0.13,0.75) | 0.36 (0.16,0.79) | 64.08 (21.08,84.29) |
| **Abbreviations-** IIV3: Trivalent **i**nactivated influenza vaccine; SD: Standard dosage; HD: High dosage; OR: odds ratio; RR: relative risk ; VE: vaccine efficacy | | | |

**Influenza-like illness (ILI)**

**Figure 3.** Forest plot of pairwise meta-analysis comparing IIV3-HD (intervention) vs. IIV3-SD (control)

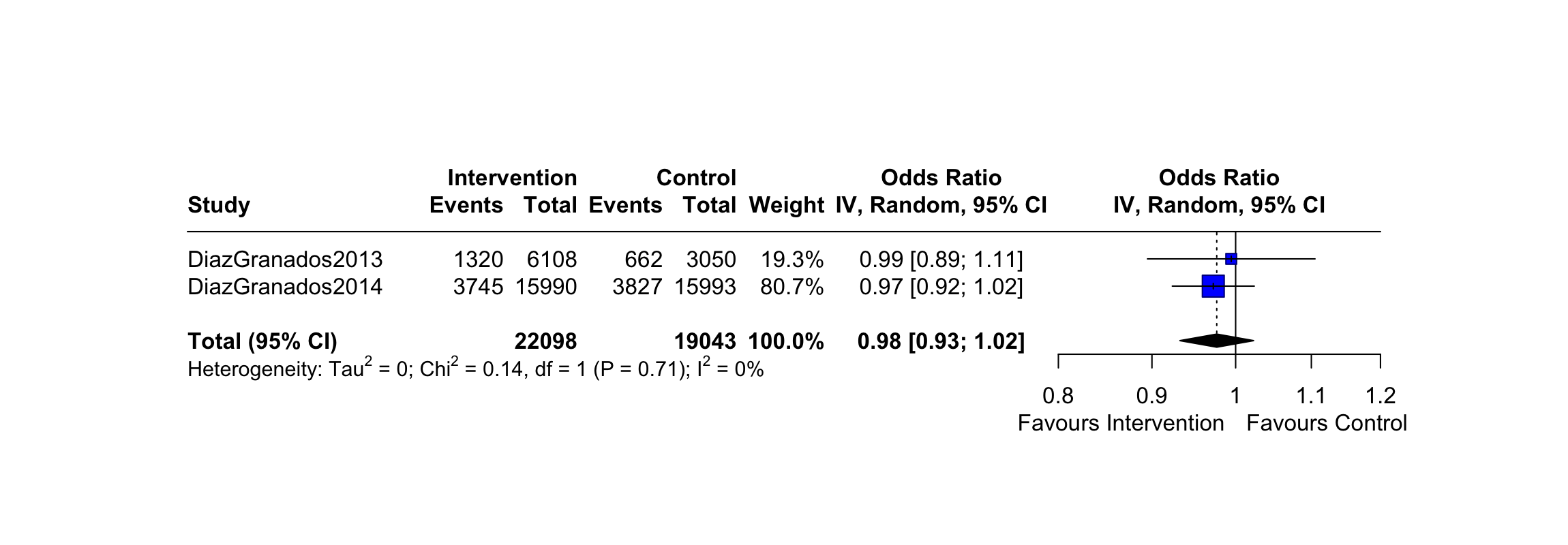

**Abbreviations-** IIV3: Trivalent **i**nactivated influenza vaccine; SD: Standard dosage; HD: High dosage; CI: Confidence interval

**Figure 4.** Forest plot of pairwise meta-analysis comparing IIV3-SD (intervention) vs. Placebo (control)

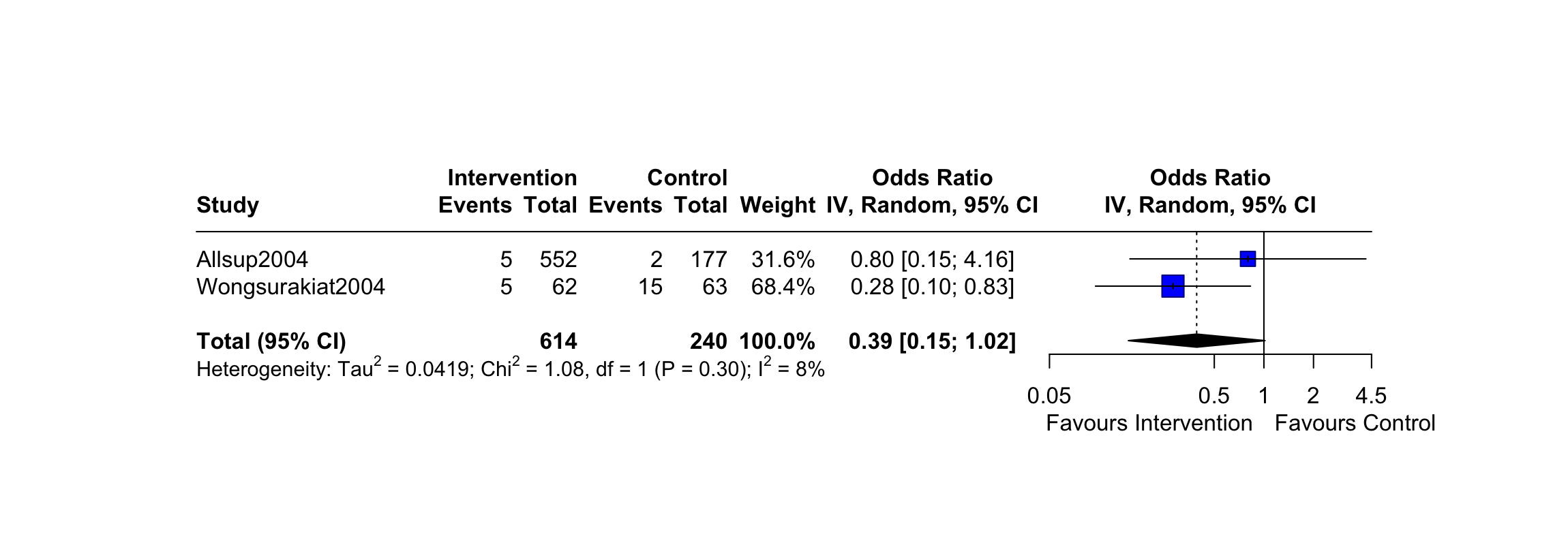

**Abbreviations-** IIV3: Trivalent **i**nactivated influenza vaccine; SD: Standard dosage; CI: Confidence interval

**Table 3**. Vaccine efficacy from pairwise meta-analysis results for influenza-like illness with original coding of interventions

| ***Comparison*** | ***OR*** | ***RR*** | ***VE*** |
| --- | --- | --- | --- |
| IIV3-HD : IIV3-SD | 0.98 (0.93,1.02) | 0.98 (0.93,1.02) | 1.76 (-1.75,6.18) |
| IIV3-SD : Placebo | 0.39 (0.15,1.02) | 0.42 (0.17,1.02) | 57.79 (-1.75,83.22) |
| **Abbreviations-** IIV3: Trivalent **i**nactivated influenza vaccine; SD: Standard dosage; HD: High dosage; OR: odds ratio; RR: relative risk ; VE: vaccine efficacy | | | |

**ER Visit for ILI**

**Figure 5.** Forest plot of pairwise meta-analysis comparing IIV3-HD (intervention) vs. IIV3-SD (control)

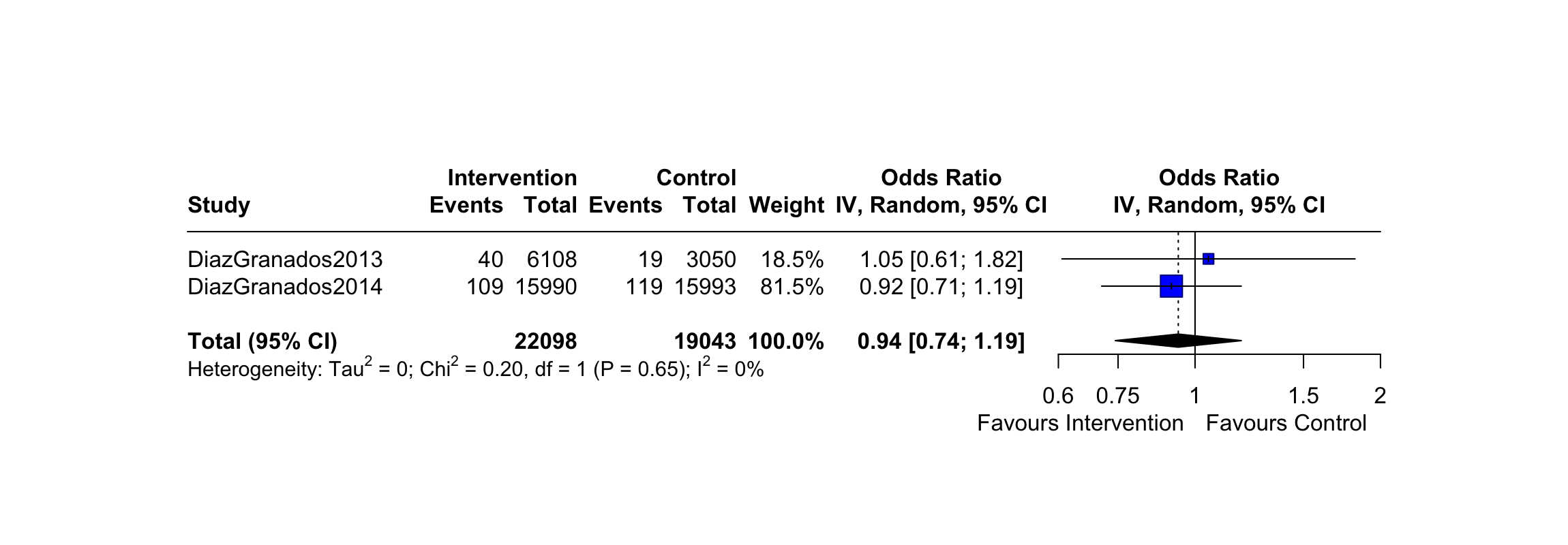

**Abbreviations-** IIV3: Trivalent **i**nactivated influenza vaccine; SD: Standard dosage; HD: High dosage; CI: Confidence interval

**Hospitalization for ILI**

**Figure 6.** Forest plot of pairwise meta-analysis comparing IIV3-HD (intervention) vs. IIV3-SD (control)

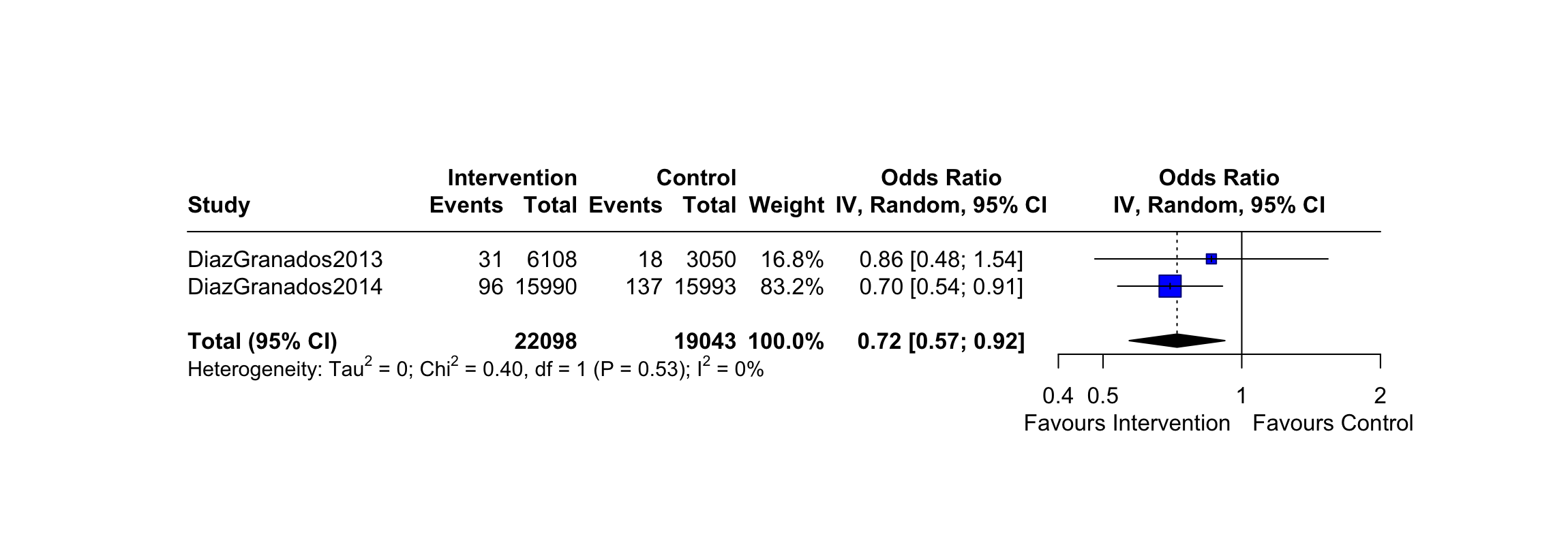

**Abbreviations-** IIV3: Trivalent **i**nactivated influenza vaccine; SD: Standard dosage; HD: High dosage; CI: Confidence interval

**Hospitalization for acute respiratory infection (ARI)**

**Figure 7.** Forest plot of pairwise meta-analysis comparing IIV3-HD (intervention) vs. IIV3-SD (control)

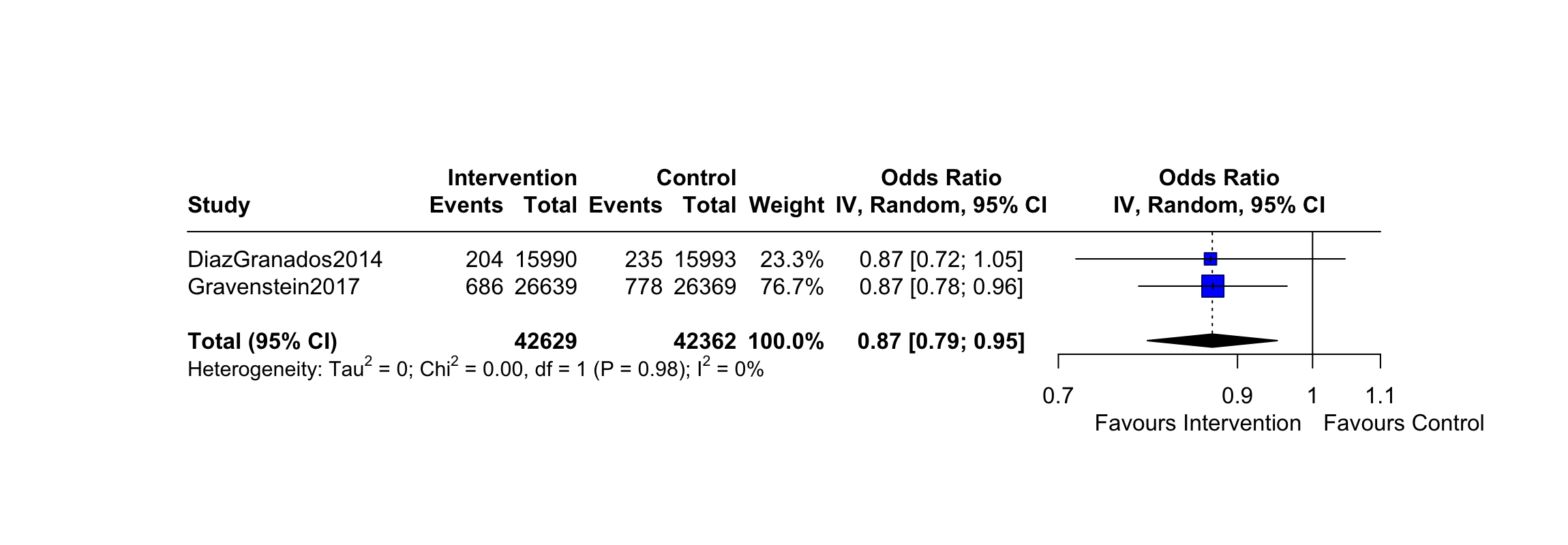

**Abbreviations-** IIV3: Trivalent **i**nactivated influenza vaccine; SD: Standard dosage; HD: High dosage; CI: Confidence interval

**Number of vascular adverse events**

**Figure 8.** Forest plot of pairwise meta-analysis comparing IIV3-Adj (intervention) vs. IIV3-SD (control)

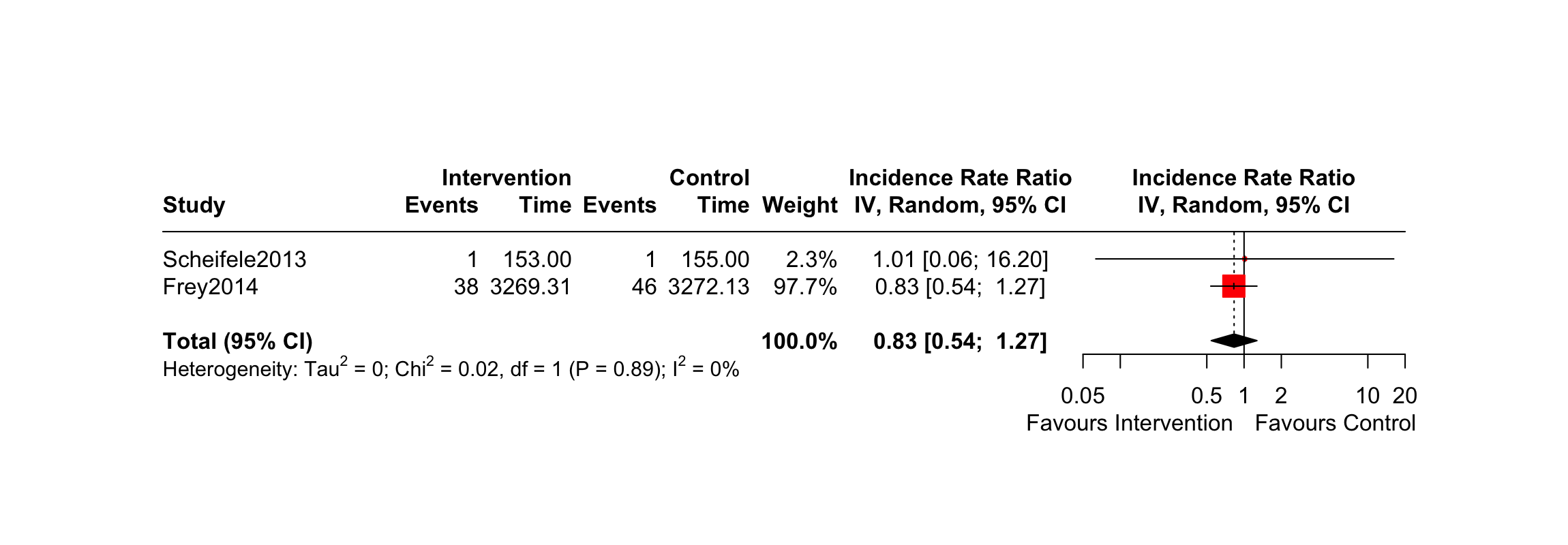

**Abbreviations-** IIV3: Trivalent **i**nactivated influenza vaccine; SD: Standard dosage; Adj: Adjuvanted; CI: Confidence interval

**Figure 9.** Forest plot of pairwise meta-analysis comparing IIV3-HD (intervention) vs. IIV3-SD (control)

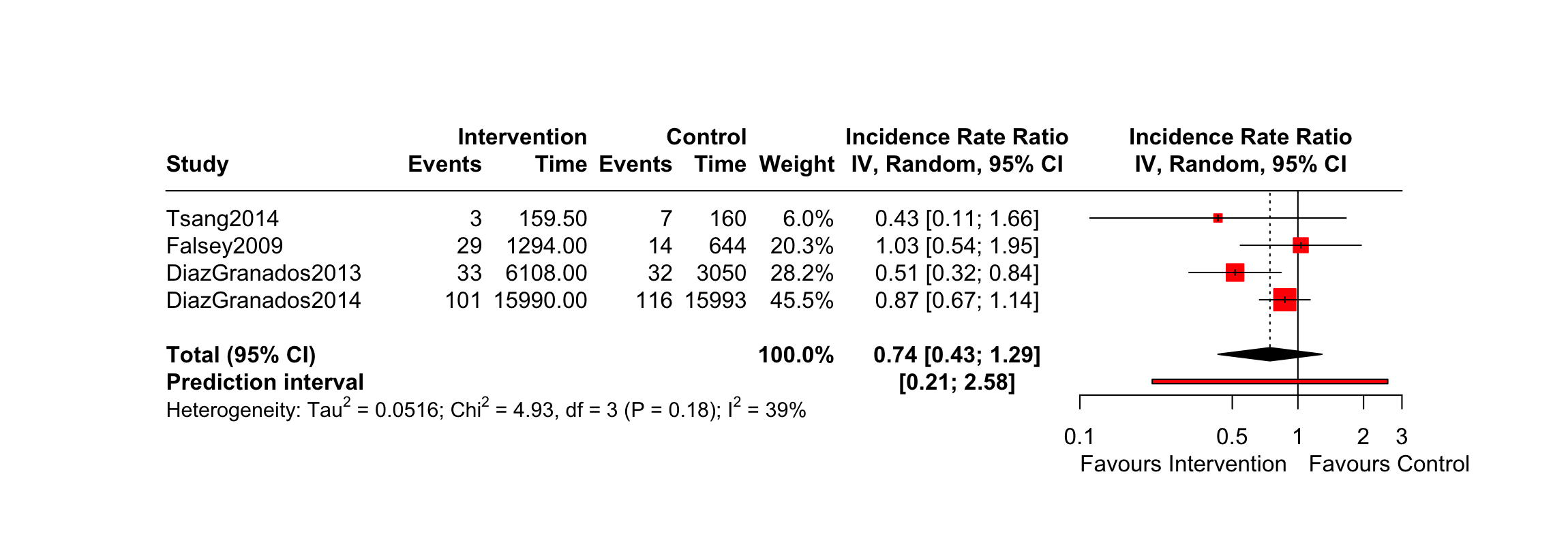

**Abbreviations-** IIV3: Trivalent **i**nactivated influenza vaccine; SD: Standard dosage; HD: High dosage; CI: Confidence interval

**Inpatient Hospitalization**

**Figure 10.** Forest plot of pairwise meta-analysis comparing IIV3-HD (intervention) vs. IIV3-SD (control)

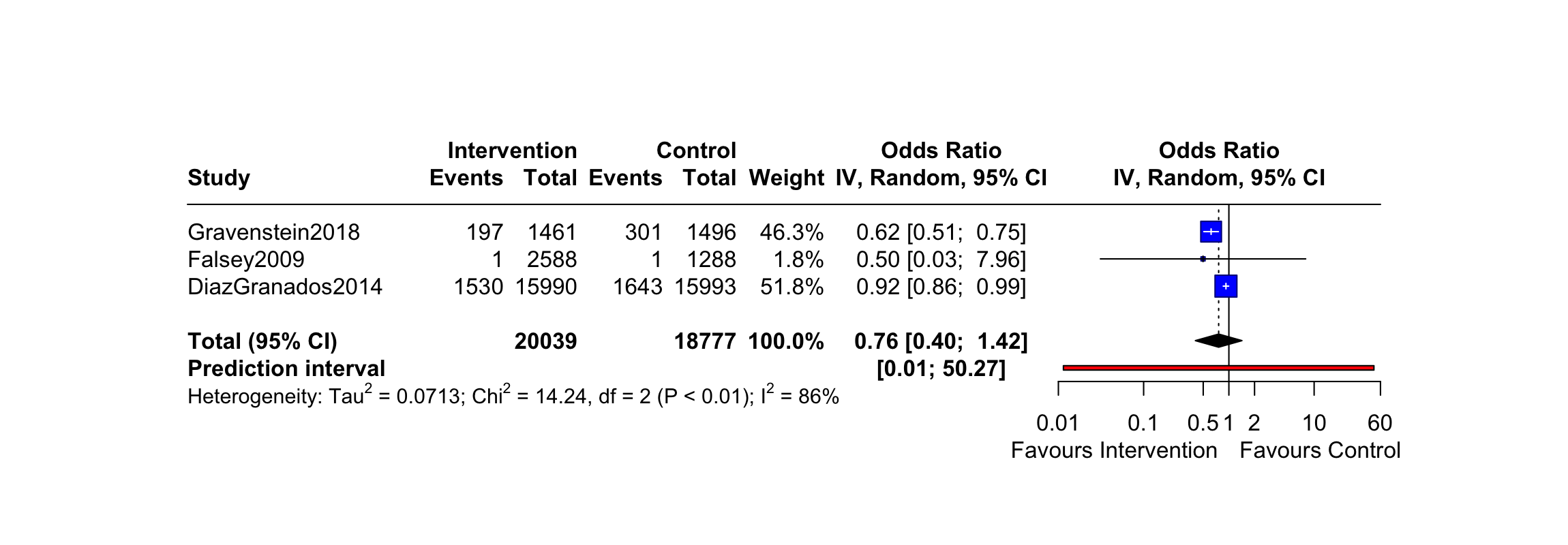

**Abbreviations-** IIV3: Trivalent **i**nactivated influenza vaccine; SD: Standard dosage; HD: High dosage; CI: Confidence interval

**Figure 11.** Forest plot of pairwise meta-analysis comparing IIV3-HD (intervention) vs. IIV3-SD (control) restricted to studies with a low overall risk of bias

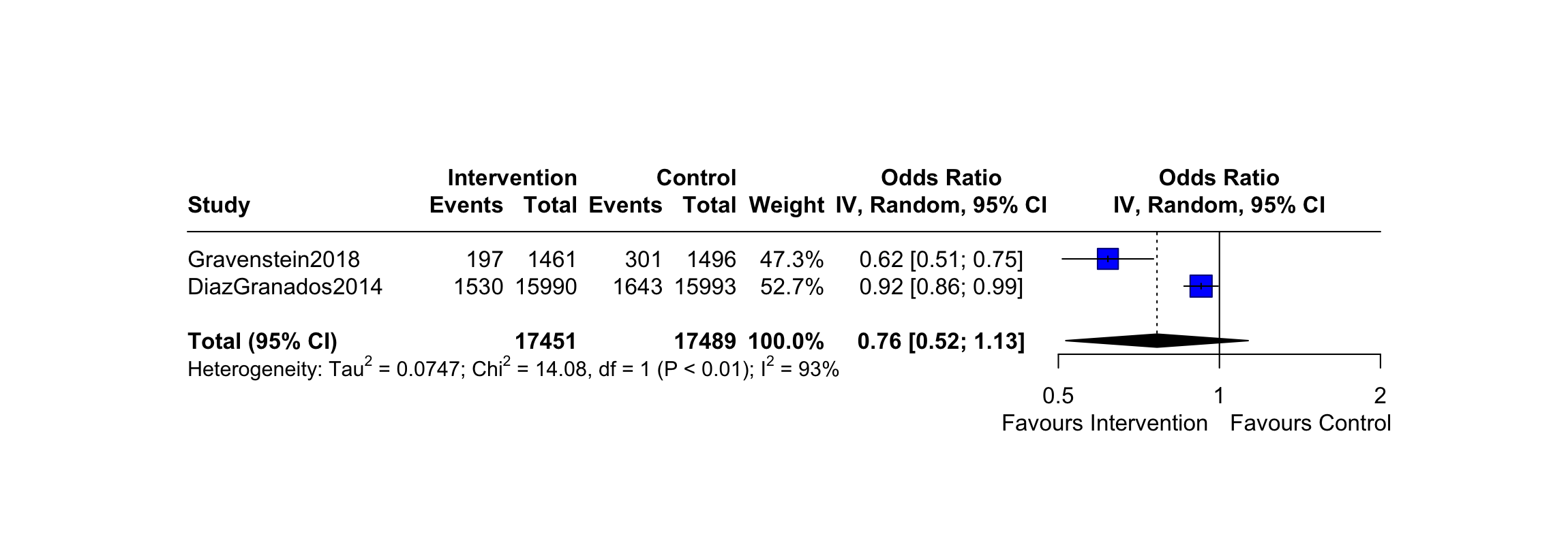

**Abbreviations-** IIV3: Trivalent **i**nactivated influenza vaccine; SD: Standard dosage; HD: High dosage; CI: Confidence interval

**Figure 12.** Forest plot of pairwise meta-analysis comparing IIV3-HD (intervention) vs. IIV3-SD (control) restricted to studies whose mean age is over 80 years of age.

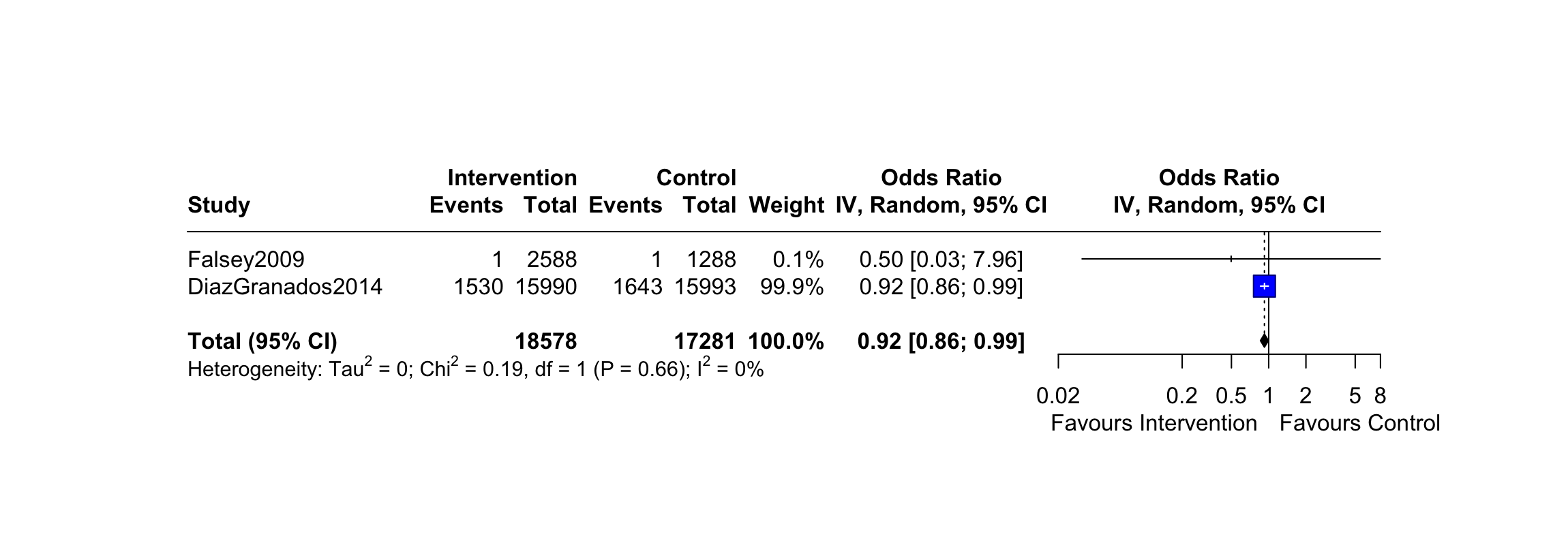

**Abbreviations-** IIV3: Trivalent **i**nactivated influenza vaccine; SD: Standard dosage; HD: High dosage; CI: Confidence interval

**Outpatient visit**

**Figure 13.** Forest plot of pairwise meta-analysis comparing IIV3-HD (intervention) vs. IIV3-SD (control)

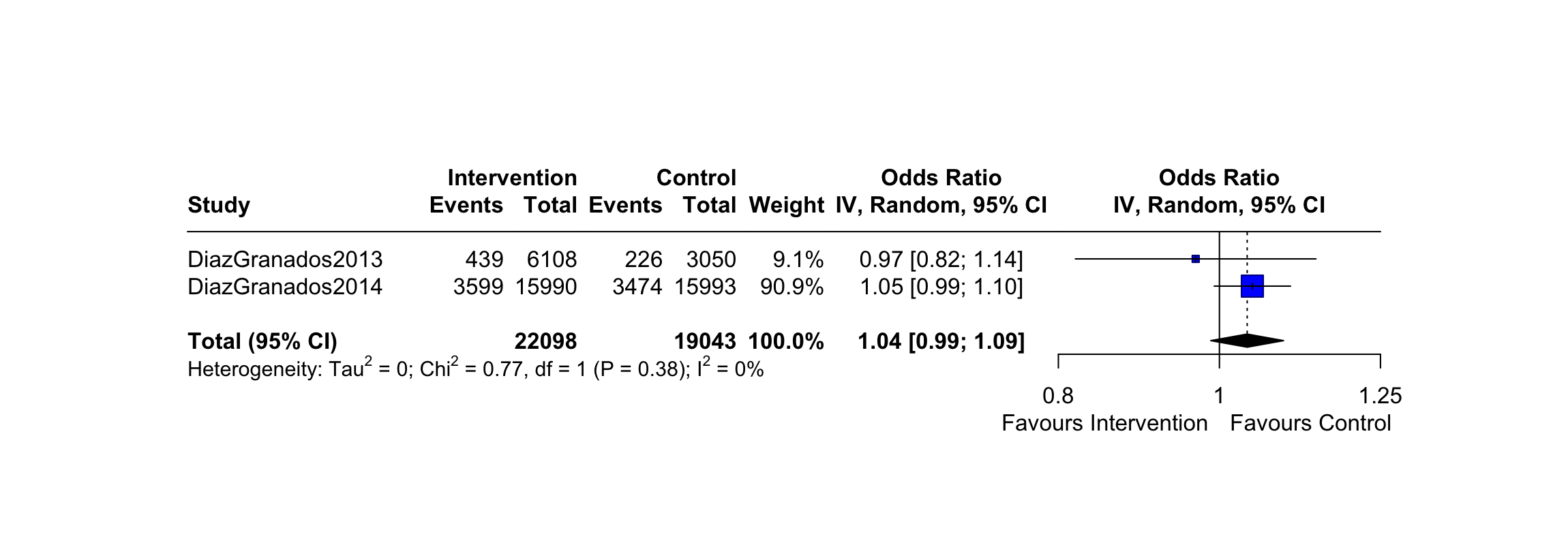

**Abbreviations-** IIV3: Trivalent **i**nactivated influenza vaccine; SD: Standard dosage; HD; High dosage; CI: Confidence interval

**Figure 14.** Forest plot of pairwise meta-analysis comparing IIV3-SD (intervention) vs. Placebo (control)

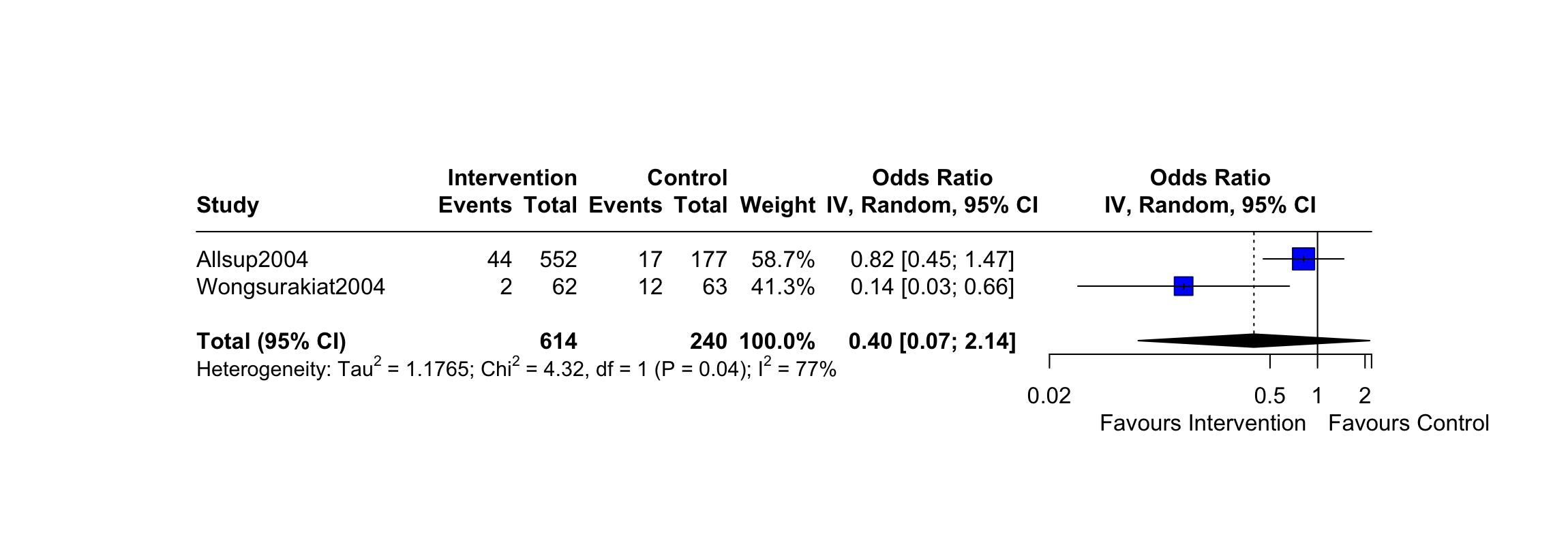

**Abbreviations-** IIV3: Trivalent **i**nactivated influenza vaccine; SD: Standard dosage; CI: Confidence interval

**All-cause mortality**

**Figure 15.** Forest plot of pairwise meta-analysis comparing IIV3-Adj (intervention) vs. IIV3-SD (control)

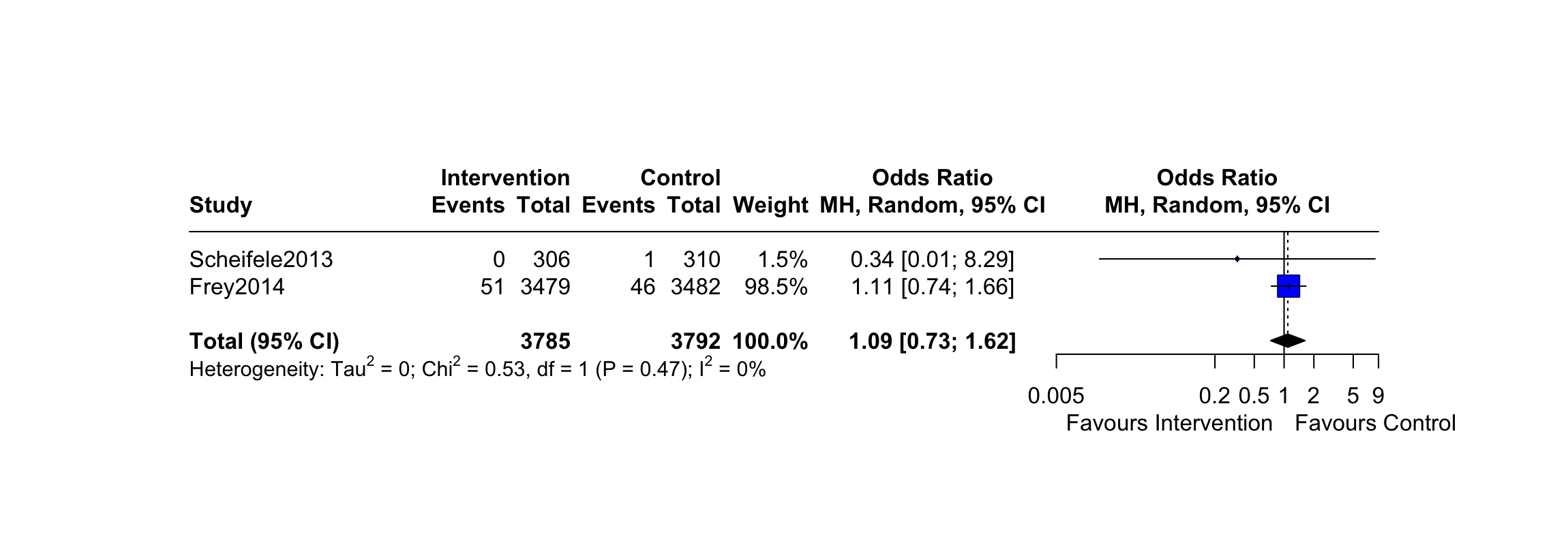
**Abbreviations-** IIV3: Trivalent **i**nactivated influenza vaccine; SD: Standard dosage; Adj: Adjuvanted; CI: Confidence interval

**Figure 16.** Forest plot of pairwise meta-analysis comparing IIV3-HD (intervention) vs. IIV3-SD (control)

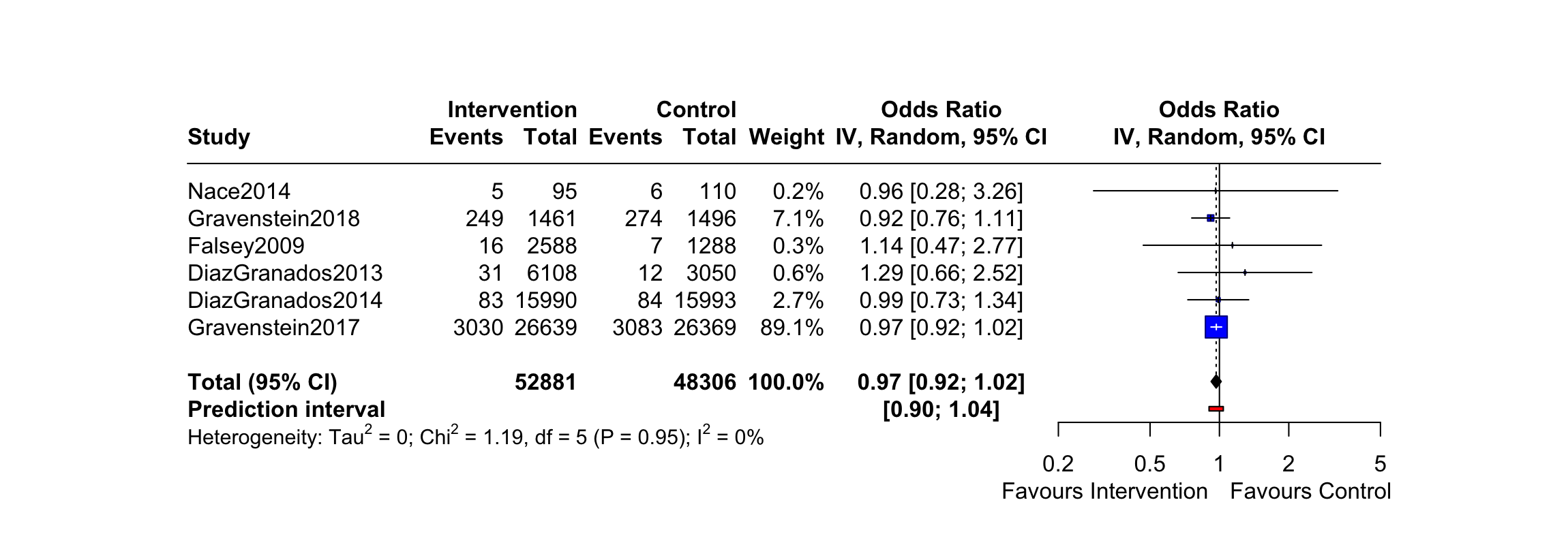

**Abbreviations-** IIV3: Trivalent **i**nactivated influenza vaccine; SD: Standard dosage; HD: High dosage; CI: Confidence interval

**Figure 17.** Forest plot of pairwise meta-analysis comparing IIV4-SD (intervention) vs. IIV3-SD (control)

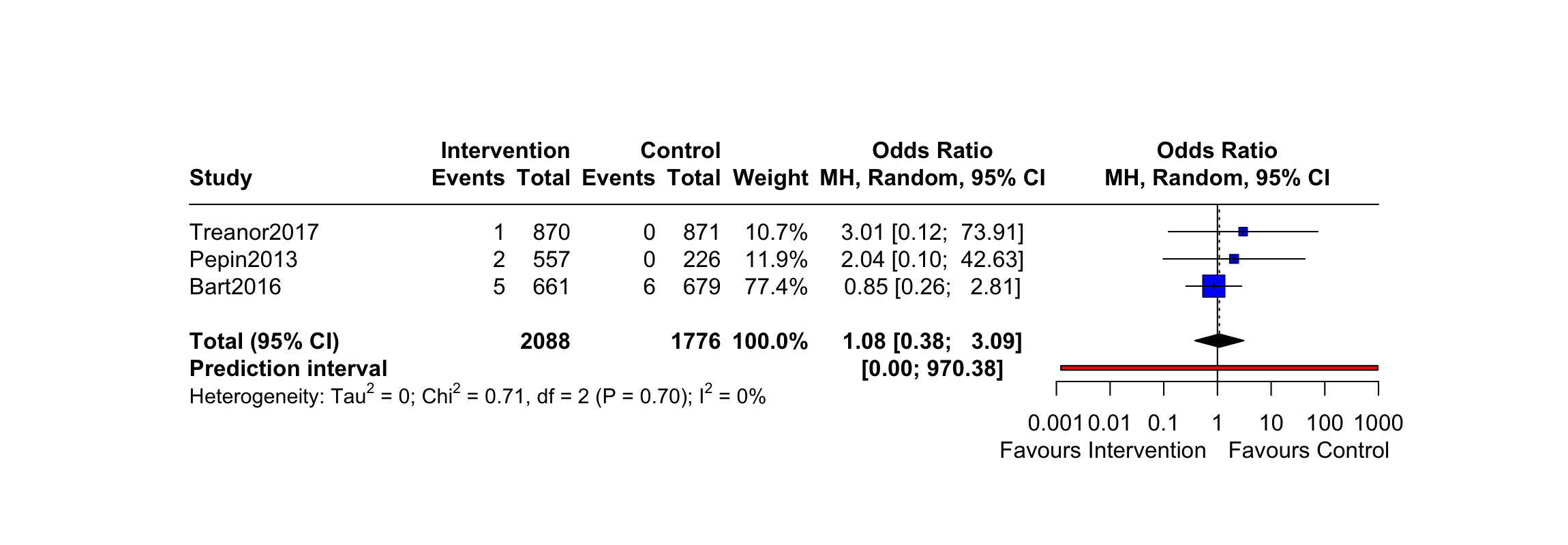

**Abbreviations-** IIV3: Trivalent **i**nactivated influenza vaccine; IIV4: Quadrivalent **i**nactivated influenza vaccine; SD: Standard dosage; CI: Confidence interval

**Figure 18.** Forest plot of pairwise meta-analysis comparing IIV3-SD (intervention) vs. Placebo (control)

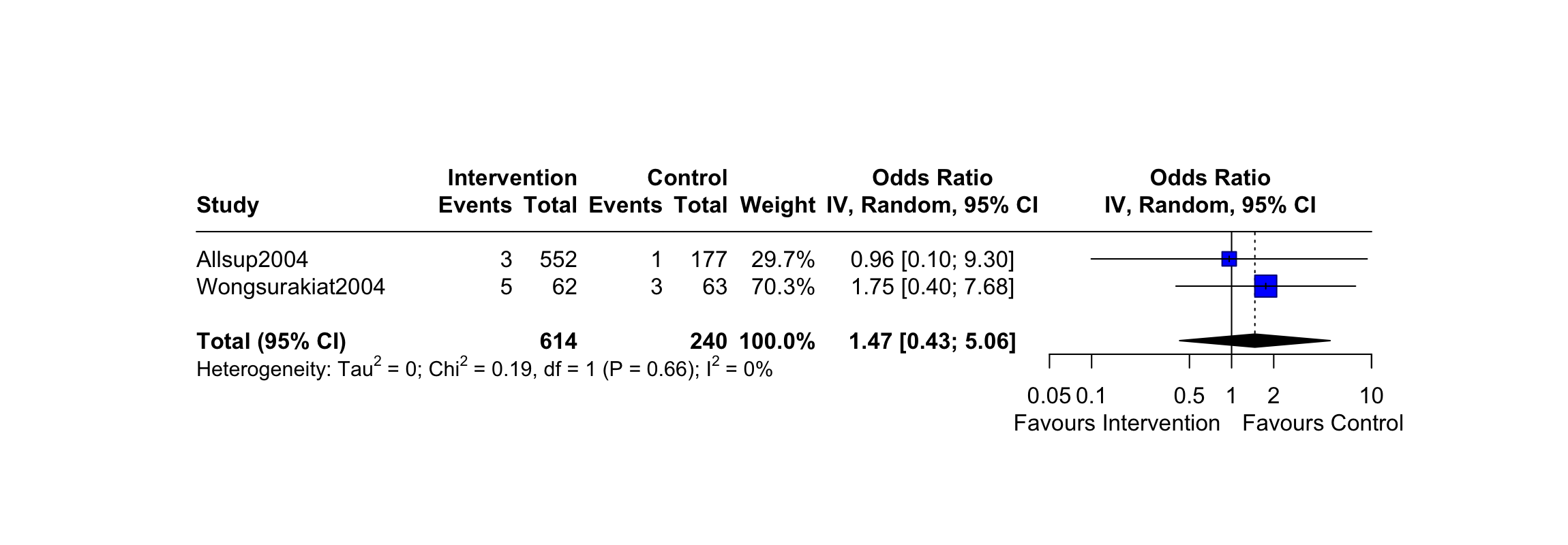

**Abbreviations-** IIV3: Trivalent **i**nactivated influenza vaccine; SD: Standard dosage; CI: Confidence interval

### **Appendix 23: Definitions of influenza-like illness (ILI)**

| **Author, Year** | **Source of Definition** | **Definition** |
| --- | --- | --- |
| DiazGranados, 2013^15^ | Determined by Investigator | “A new onset (or exacerbation of a pre-existing condition) of at least one of the following systemic symptoms: temperature >37.2 ◦C (>99.0 ◦F), feverishness (feeling of warmth), chills, tiredness, headaches or myalgia; and at least one of the following respiratory symptoms: nasal congestion or rhinorhea, sore throat, cough, sputum production, wheezing, chest tightness, shortness of breath, or chest pain with breathing.” |
| DiazGados, 2014^20^ | Determined by Investigator | “A respiratory illness with sore throat, cough, sputum production, wheezing, or difficulty breathing, concurrent with one or more of the following: temperature above 37.2°C, chills, tiredness, headaches, or myalgia.” |
| Frey, 2014^19^ | Determined by Investigator | “Temperature of ≥37.2◦C or feverishness and at least two of the following symptoms: headache, myalgia, cough, or sore throat.” |
| Beran, 2021^32^ | Determined by Investigator | “At least one respiratory symptom (sore throat, cough, sputum production, wheezing, or difficulty breathing) concurrently with at least one systemic symptom (temperature >37·2°C, chills, tiredness, headache, or myalgia).” |
| Dunkle, 2017^24^ | Determined by Investigator | “At least one symptom in both the respiratory and systemic illness categories, regardless of severity.” |
| Keitel, 2010^14^ | Determined by Investigator | “An influenza symptoms score of 2 or greater on the Flu Symptoms Card (based on presence of fever, cough, sore throat, and runny nose/stuffy nose, muscle or joint aches, headache, chills/sweats, and tiredness/malaise), or if they sought medical care for their acute illness.” |
| Wongsurakiat, 2004^11^ | Determined by Investigator | “Patients had symptoms of generalized aches, fever, and headache with or without upper respiratory tract symptoms.” |
| Rudenko, 2001^10^ | Determined by Investigator | “A temperature of ≥37.28C, and at least one respiratory symptom: cough, coryza, or sore throat.” |
| Teh, 2021^34^ | Determined by Investigator | “Presence of fever (≥38°C) and at least 1 respiratory symptom.” |
| Allsup, 2004^12^ | Determined by Investigator | “An illness of sudden onset with features of cough, feverishness, prostration, myalgia and widespread aches and pains.” |
| **Abbreviations:** None. | | |

### **Appendix 24: Definitions of vascular adverse events**

| **Author, Year** | **Source of Definition** | **Definition** |
| --- | --- | --- |
| DiazGranados, 2013^15^ | Results from trial registry | “Acute myocardial infarction, myocardial infarction, cardiac arrest, cardiac failure, cardiac failure acute, cardiac failure congestive, cardiac failure chronic, cerebral ischaemia, haemorrhagic stroke, ischaemic stroke.” |
| DiazGranados, 2014^20^ | Results from trial registry | “Acute myocardial infarction, cardiac arrest, myocardial infarction, myocardial ischaemia, cardiac failure, cardiac failure acute, cardiac failure chronic, cardiac failure congestive, cerebral infarction, cerebral ischaemia, ischaemic stroke, vascular infarction, vascular ischaemia, haemorrhagic stroke.” |
| Frey, 2014^19^ | Results from trial registry | “Acute myocardial infarction, myocardial infarction, myocardial ischemia, cardiac arrest, cardiac failure, cardiac failure congestive, cerebral infraction, cerebral ischaemia, ishaemic stroke.” |
| Beran, 2021^32^ | Results from trial registry | “Acute myocardial infarction, myocardial infarction, cardiac arrest, cardiac failure, cardiac failure acute, cardiac failure congestive, cardiac failure chronic, cerebral infarction, ischaemic stroke, haemorrhagic stroke.” |
| Dunkle, 2017^32^ | Results from trial registry | “Myocardial infarction, cardiac arrest, cardiac failure, acute myocardial infarction, embolic stroke, hemorrhagic stroke.” |
| Essink, 2020^31^ | Results from trial registry | “Myocardial infarction, Ischaemic cerebral infarction.” |
| Chang, 2019^27^ | Results from trial registry | “Acute Myocardial Infarction, cardiac failure congestive, myocardial infarction, cerebrovascular accident, ischaemic cerebral infarction.” |
| Falsey, 2009^13^ | Results from trial registry | “Acute coronary syndrome, Acute myocardial infarction, Cardiac arrest, Cardiac Failure, Cardiac failure congestive, Cardio-respiratory arrest, Myocardial infarction, Cerebrovascular accident, Transient ischaemic attack.” |
| Novartis Vaccines and Diagnostics, 2016^44^ | Results from trial registry | “Myocardial Infarction, Cerebral Vascular Accident.” |
| Schmader, 2021^33^ | Results from trial registry | “Congestive heart failure, Transient ischemic attack.” |
| Pepin, 2013^17^ | Results from trial registry | “Cardiac arrest, myocardial infarction.” |
| Szymczakiewicz-Multanowska, 2009^38^ | Results from trial registry | “Acute myocardial infraction and cerebral infraction, Transient ischaemic attack.” |
| Szymczakiewicz-Multanowska, 2012^a39^ | Results from trial registry | “Acute myocardial infraction and cerebral infraction, Transient ischaemic attack.” |
| Szymczakiewicz-Multanowska, 2012^b40^ | Results from trial registry | “Acute myocardial infraction, cardiac arrest, cardiac failure, myocardial infraction, myocardial ischaemia, ischaemic stroke, cerebrovascular accident.” |
| Tsang, 2014^18^ | Results from trial registry | “Acute myocardial infarction, cardiac arrest, cardiac failure, cardiac failure congestive & thrombotic stroke; death reported in text as: "no treatment-related serious adverse events or treatment-related deaths occurred during the study." |
| Scheifele, 2013^16^ | SAE determined by investigator | “Vascular”. |
| **Abbreviations**: SAE: Serious Adverse Event. | | |

### **Appendix 25: Vaccine composition and circulating strains**

| **Author, Year** | **Setting** | **Influenza season** | **Tx** | **Vaccine type** | **Vaccine name** | **Vaccine strain** | **Circulating strain** |
| --- | --- | --- | --- | --- | --- | --- | --- |
| **Studies included in quantitative and descriptive analysis (n = 26)** | | | | | | | |
| **Rudenko, 2001^10^** | Nursing home | N 1996/1997 | Tx1 | IIV3-SD | Trivalent inactivated, split virus influenza vaccine | A/Texas/36/91 (H1N1), A/Nanchang/933/95 (H3N2), B/Harbin/07/94 | Other circulating strains:  A/H1N1 and influenza B |
|  |  |  | Tx2 | Placebo | Placebo | NA |  |
| **Wongsurakiat, 2004^11^** | Clinic | S 1997/1998 | Tx1 | IIV3-SD | Trivalent inactivated, split virus influenza vaccine SD | A/Texas/36/91 (H1N1), A/Nanchang/933/95 (H3N2), B/Harbin/07/94 | Predominant circulating strain:  A/Sydney/5/97 (H3N2) |
|  |  |  | Tx2 | Placebo | Placebo (Vitamin B1) | NA |  |
| **Allsup, 2004^12^** | Community | N 1999/2000 | Tx1 | IIV3-SD | Trivalent inactivated, split virion influenza vaccine SD | A/Beijing/262/95(H1N1), A/Sydney/5/97 (H3N2), B/Beijing/184/93 | Predominant circulating strain:  A/H3N2 |
|  |  |  | Tx2 | Placebo | Placebo | NA |  |
| **Falsey, 2009^13^** | Community | N 2006/2007 | Tx1 | IIV3-HD | Fluzone HD | A/New Caledonia/20/99 (H1N1), A/Wisconsin/67/2005 (H3N2), B/Malaysia/2506/04 | Predominant circulating strain: A/H1  Other circulating strain: A/H3 |
|  |  |  | Tx2 | IIV3-SD | Fluzone SD | A/New Caledonia/20/99 (H1N1), A/Wisconsin/67/2005 (H3N2), B/Malaysia/2506/04 |  |
| **Keitel, 2010^14^** | Community | N 2006/2007 | Tx1 | RIV3 | FluBlok | A/Wisconsin (H3N2); A/New Caledonia (H1N1), B/Ohio | Predominant circulating strain: A/H1  Other circulating strain: A/H3 |
|  |  |  | Tx2 | IIV3-SD | Fluzone SD | A/Wisconsin (H3N2); A/New Caledonia (H1N1), B/Malaysia |  |
| **DiazGranados, 2013^15^** | Clinic | N 2009/2010 | Tx1 | IIV3-HD | Fluzone HD | A/Brisbane/59/07 (H1N1), A/Uruguay/716/2007 X-175C (H3N2), and B/Brisbane/60/2008 strains | Predominant circulating strain:  H1N1 |
|  |  |  | Tx2 | IIV3-SD | Fluzone SD | A/Brisbane/59/07 (H1N1), A/Uruguay/716/2007 X-175C (H3N2), B/Brisbane/60/2008 |  |
| **Scheifele, 2013^16^** | Community or living assistance facility | N 2011/2012 | Tx1 | IIV3-Adj | Fluad, adjuvanted | A/ California/7/2009 (H1N1)-like, A/Perth/16/2009 (H3N2)-like and B/Brisbane/60/2008 | Circulating strains:  A/H3N2, pH1N1 and influenza B |
|  |  |  | Tx2 | IIV3-SD | Agriflu SD | A/ California/7/2009 (H1N1)-like, A/Perth/16/2009 (H3N2)-like and B/Brisbane/60/2008 |  |
| **Pepin, 2013^17^** | Community | N 2011/2012 | Tx1 | IIV4-SD | Quadrivalent inactivated influenza vaccine SD | A/California/07/2009(H1N1), A/Victoria/210/2009 (H3N2), B/Brisbane/60/2008 (Victoria lineage), and B/Florida/04/2006 (Yamagata lineage) strains | Predominant circulating Strain: A/H3N2  Other circulating strains: B viruses and A/H1N1pdm09 |
|  |  |  | Tx2 | IIV3-SD | Trivalent inactivated influenza vaccine SD (pooled) | A/California/07/2009 (H1N1),A/Victoria/210/2009 (H3N2), and B/Brisbane/60/2008 (Victoria lineage) strains OR B/Florida/04/2006 (Yamagata lineage) strains |  |
| **Tsang, 2014^18^** | NR | N 2007/2008 | Tx1 | IIV3-SD | Fluzone SD | A/Solomon Islands/3/2006 (H1N1), A/Wisconsin/67/2005 (H3N2), and B/Malaysia/2506/2004 | Circulating strains: A/H1N1, A/H3N2 and B viruses |
|  |  |  | Tx2 | IIV3-HD | Fluzone HD | A/Solomon Islands/3/2006 (H1N1), A/Wisconsin/67/2005 (H3N2), and B/Malaysia/2506/2004 |  |
| **Frey, 2014^19^** | NR | N 2010/2011  and S 2010/2011  *Note: This study was conducted in 4 different countries in different hemisphere but in methods they only mention northern hemisphere vaccine | Tx1 | IIV3-Adj | Fluad, adjuvanted | A/California/7/2009 (H1N1), A/Perth/16/2009 (H3N2), B/Brisbane/60/2008 | Predominant circulating strain: A/H3N2  Other circulating strain: A/H1N1 |
|  |  |  | Tx2 | IIV3-SD | Agriflu SD | A/California/7/2009 (H1N1), A/Perth/16/2009 (H3N2), B/Brisbane/60/2008 |  |
| **DiazGranados, 2014^20^** | Research centers | N 2011/2012 and  N 2012/2013 | Tx1 | IIV3-HD | Fluzone HD | 2011/2012:  A/California/ 7/2009 (H1N1), A/Victoria/210/2009 (H3N2), B/Brisbane/60/2008;  2012/2013:  A/California/7/2009 (H1N1), A/Victoria/361/2011 (H3N2), B/Texas/6/2011 (B/Wisconsin/1/2010-like virus) | Predominating circulating strain: A/H3N2 |
|  |  |  | Tx2 | IIV3-SD | Fluzone SD | 2011/2012:  A/California/ 7/2009 (H1N1),A/Victoria/210/2009 (H3N2), B/Brisbane/60/2008;  2012/2013:  A/California/7/2009 (H1N1), A/Victoria/361/2011 (H3N2), B/Texas/6/2011 (B/Wisconsin/1/2010-like virus) |  |
| **Nace, 2014^21^** | Long term care home | N 2011/2012 and N 2012/2013 | Tx1 | IIV3-HD | Fluzone HD | 2011/2012: A/California/7/2009(H1N1), A/Victoria/210/2009 (H3N2), B/Brisbane/60/2008;  2012/2013: A/California/7/2009(H1N1), A/Victoria/361/2011(H3N2), B/Texas/6/2011 | Predominant circulating strain:  A/H3N2 |
|  |  |  | Tx2 | IIV3-SD | Fluzone SD | 2011/2012: A/California/7/2009(H1N1), A/Victoria/210/2009 (H3N2), B/Brisbane/60/2008;  2012/2013: A/California/7/2009(H1N1), A/Victoria/361/2011(H3N2), B/Texas/6/2011 |  |
| **Bart, 2016^22^** | Community | N 2013/2014 | Tx1 | IIV3-CC-SD | Flucelvax/Optaflu SD | A/Brisbane/10/2010 (H1N1);  A/Texas/50/2012 (H3N2);  B/Massachusetts/2/2012 (Yamagata lineage) OR B/Brisbane/60/2008 (Victoria lineage) | Predominant circulating strain:  A/H1N1pdm09 |
|  |  |  | Tx2 | IIV4-CC-SD | Quadrivalent inactivated influenza vaccine, cell-cultured, SD | A/Brisbane/10/2010 (H1N1);  A/Texas/50/2012 (H3N2);  B/Massachusetts/2/2012;  B/Brisbane/60/2008 |  |
| **Gravenstein, 2017^23^** | Nursing home | N 2013/2014 | Tx1 | IIV3-HD | Fluzone HD | A/California/7/2009 (H1N1)pdm09-like virus; A/Victoria/361/2011 (H3N2); B/Massachusetts/2/2012-like virus | Predominant circulating strain: A/H1N1pdm09 |
|  |  |  | Tx2 | IIV3-SD | Fluzone SD | A/California/7/2009 (H1N1)pdm09-like virus; A/Victoria/361/2011 (H3N2); B/Massachusetts/2/2012-like virus |  |
| **Dunkle, 2017^24^** | Outpatient centers | N 2014/2015 | Tx1 | RIV4 | Flublok | A/California/7/2009 (H1N1)-like, A/Texas/50/2012 (H3N2), B/Massachusetts/2/2012, B/Brisbane/60/2008 | Predominant circulating strain:  A/H3N2 |
|  |  |  | Tx2 | IIV4-SD | Fluarix SD | A/California/7/2009 (H1N1)-like, A/Texas/50/2012 (H3N2), B/Massachusetts/2/2012, B/Brisbane/60/2008 |  |
| **Treanor, 2017^25^** | NR | N 2014/2015 | Tx1 | IIV4-SD | Afluria Quadrivalent SD | A/California/7/2009 (H1N1) pdm09-like virus, A/Texas/50/2012 (H3N2)-like virus, B/Massachusetts/2/2012- like virus (B/Yamagata lineage), B/Brisbane/60/2008-like virus (B/Victoria lineage) | Predominant circulating strain:  A/H3N2 |
|  |  |  | Tx2 | IIV3-SD | Trivalent inactivated influenza vaccine (pooled) SD | A/California/7/2009 (H1N1) pdm09-like virus, A/Texas/50/2012 (H3N2)-like virus, B/Massachusetts/2/2012-like virus (B/Yamagata lineage)  A/California/7/2009 (H1N1) pdm09-like virus, A/Texas/50/2012 (H3N2)-like virus, B/Brisbane/ 60/ 2008-like virus (B/Victoria lineage) |  |
| **Gravenstein, 2018^26^** | Nursing home | N 2012/2013 | Tx1 | IIV3-HD | Fluzone HD | A/California/7/2009-like (pH1N1), A/Victoria/361/2011-like (H3N2), and B/Wisconsin/1/2010-like (B/Yamagata lineage) | Predominant circulating strain:  A/H3N2 for the  2012–2013 season |
|  |  |  | Tx2 | IIV3-SD | Fluzone SD | A/California/7/2009-like (pH1N1), A/Victoria/361/2011-like (H3N2), and B/Wisconsin/1/2010-like (B/Yamagata lineage) |  |
| **Chang, 2019^27^** | Community | N 2017/2018 | Tx1 | IIV4-HD | Quadrivalent inactivated split virion influenza vaccine HD | A/Michigan/45/2015 X-275, A/Hong Kong/4801/2014 [NYMC X-263B], B/Brisbane/60/2008, B/Phuket/3073/2013 | Predominant circulating strain:  A/H3N2 |
|  |  |  | Tx2 | IIV3-HD | Fluzone (pooled) HD | A/Michigan/45/2015 X-275, A/Hong Kong/4801/2014 [NYMC X-263B], B/Brisbane/60/2008 OR B/Phuket/3073/2013 |  |
| **Loeb, 2020^28^** | Community | N 2014/2015, N 2015/2016, N 2016/2017 and N 2017/2018 | Tx1 | IIV3-SD | Fluzone SD | 2014/2015: A/California/7/2009-like (2009 H1N1) virus, an A/Texas/50/2012-like (H3N2) virus, and a B/Massachusetts/2/2012-like (B/Yamagata lineage) virus  2015/2016: A/California/7/2009 (H1N1)pdm09-like virus, an A/Switzerland/9715293/2013 (H3N2)-like virus, and a B/Phuket/3073/2013-like (B/Yamagata lineage) virus  2016/2017: A/California/7/2009 (H1N1)pdm09-like virus, an A/Hong Kong/4801/2014 (H3N2)-like virus, and a B/Brisbane/60/2008-like virus (B/Victoria lineage)  2017/2018: A/Michigan/45/2015 (H1N1)pdm09-like virus, an A/Hong Kong/4801/2014 (H3N2)-like virus, and a B/Brisbane/60/2008-like (B/Victoria lineage) virus | Predominant circulating strains in  USA: 2014/2015: A/H3N2; 2015/2016: A/H1N1pdm09; 2016/2017: A/H3N2; 2017/2018: A/H3N2 |
|  |  |  | Tx2 | IIV3-HD | Fluzone HD | 2014/2015: A/California/7/2009-like (2009 H1N1) virus, an A/Texas/50/2012-like (H3N2) virus, and a B/Massachusetts/2/2012-like (B/Yamagata lineage) virus  2015/2016: A/California/7/2009 (H1N1)pdm09-like virus, an A/Switzerland/9715293/2013 (H3N2)-like virus, and a B/Phuket/3073/2013-like (B/Yamagata lineage) virus  2016/2017: A/California/7/2009 (H1N1)pdm09-like virus, an A/Hong Kong/4801/2014 (H3N2)-like virus, and a B/Brisbane/60/2008-like virus (B/Victoria lineage)  2017/2018: A/Michigan/45/2015 (H1N1)pdm09-like virus, an A/Hong Kong/4801/2014 (H3N2)-like virus, and a B/Brisbane/60/2008-like (B/Victoria lineage) virus |  |
| **Belongia, 2020^29^** | Clinic | N 2017/2018 | Tx1 | IIV3-HD | Fluzone HD | Not fully defined: "The A(H3N2) recommended vaccine component was identical in 2016–17 and 2017/18:  A/Hong Kong/4801/2014–like (clade 3C.2a)." | Predominant circulating strain:  circulating A(H3N2) viruses: (A/Singapore/INFIMH-16–0019/2016 and A/Kentucky/29/2017)  Other circulating strain: A/Kansas/14/2017 (KS14), an antigenically advanced virus that circulated at a low level during 2017–18. |
|  |  |  | Tx2 | IIV3-Adj | Fluad, adjuvanted | Not fully defined: "The A(H3N2) recommended vaccine component was identical in 2016–17 and 2017/18: A/Hong Kong/4801/2014–like (clade 3C.2a)." |  |
|  |  |  | Tx3 | RIV4 | FluBlok | Not fully defined: "The A(H3N2) recommended vaccine component was identical in 2016–17 and 2017/18:  A/Hong Kong/4801/2014–like (clade 3C.2a)." |  |
| **Cowling, 2020^30^** | Community | N 2017/2018 | Tx1 | IIV4-SD | FluQuadri SD | A/Michigan/45/2015(H1N1)-like virus (clade 6B.1), A/Hong Kong/4801/2014(H3N2)-like virus (clade 3C.2a), B/Brisbane/60/2008-like virus (Victoria lineage; clade 1A), B/Phuket/3073/2013-like virus (Yamagata lineage; clade 3) | Predominant circulating strain:  A(H1N1)pdm09 |
|  |  |  | Tx2 | IIV3-Adj | Fluad, adjuvanted | A/Michigan/45/2015(H1N1)-like virus (clade 6B.1), A/Hong Kong/4801/2014(H3N2)-like virus (clade 3C.2a), B/Brisbane/60/2008-like virus (Victoria lineage; clade 1A) |  |
|  |  |  | Tx3 | IIV3-HD | Fluzone HD | A/Michigan/45/2015(H1N1)-like virus (clade 6B.1), A/Hong Kong/4801/2014(H3N2)-like virus (clade 3C.2a), B/Brisbane/60/2008-like virus (Victoria lineage; clade 1A) |  |
|  |  |  | Tx4 | RIV4 | Flublok | A/Michigan/45/2015(H1N1)-like virus (clade 6B.1), A/Hong Kong/4801/2014(H3N2)-like virus (clade 3C.2a), B/Brisbane/60/2008-like virus (Victoria lineage; clade 1A), B/Phuket/3073/2013-like virus (Yamagata lineage; clade 3) |  |
| **Essink, 2020^31^** | NR | N 2017/2018 | Tx1 | IIV4-Adj | Quadrivalent inactivated influenza vaccine, adjuvanted | A/Michigan/45/2015 (H1N1)-like virus, A/Hong Kong/4801/2014 (H3N2)-like virus, B/Phuket/3073/2013-like virus (Yamagata lineage), B/Brisbane/60/2008-like virus (Victoria lineage) | Predominant circulating strain:  A/H3N2 |
|  |  |  | Tx2 | IIV3-Adj | Fluad (pooled), adjuvanted | A/Michigan/45/2015 (H1N1)-like virus, A/Hong Kong/4801/2014 (H3N2)-like virus, B/Brisbane/60/2008-like virus (Victoria lineage) OR B/Phuket/3073/2013-like virus (Yamagata lineage) |  |
| **Beran, 2021^32^** | Community | N 2016/2017 and  S 2017 | Tx1 | IIV4-Adj | Quadrivalent inactivated influenza vaccine, adjuvanted | N 2016/17:  A California/7/2009pdm NYMCX-181 (A H1N1), A Hong Kong/4801/2014 NYMCX-263B (A H3N2), B Brisbane/9/2014 (B Yamagata), B Brisbane/60/2008 (B Victoria)  S 2017:  A Singapore/GP1908/2015 IVR-180 (A H1N1), A Hong Kong/4801/2014 NYMCX-263B (A H3N2), B Brisbane/9/2014 (B Yamagata), B Brisbane/60/2008 (B Victoria) | Unspecified A/H3N2 strains |
|  |  |  | Tx2 | Tdap | Boostrix | NA |  |
| **Schmader, 2021^33^** | Community | N 2017/2018 and N 2018/2019 | Tx1 | IIV3-Adj | Fluad,, adjuvanted | 2017/2018: A/Michigan/45/2015 (H1N1)pdm09-like virus, an A/Hong Kong/4801/2014 (H3N2)-like virus, and a B/Brisbane/60/2008-like (B/Victoria lineage) virus  2018/2019: A/Michigan/45/2015 (H1N1)pdm09-like virus, A/Singapore/INFIMH-16-0019/2016 A(H3N2)-like virus (updated), B/Colorado/06/2017-like (Victoria lineage) virus (updated) | Predominant circulating strains:  A/H3N2 and A/H1N1pdm09, A/H3N2 |
|  |  |  | Tx2 | IIV3-HD | Fluzone HD | 2017/2018: A/Michigan/45/2015 (H1N1)pdm09-like virus, an A/Hong Kong/4801/2014 (H3N2)-like virus, and a B/Brisbane/60/2008-like (B/Victoria lineage) virus  2018/2019: A/Michigan/45/2015 (H1N1)pdm09-like virus, A/Singapore/INFIMH-16-0019/2016 A(H3N2)-like virus (updated), B/Colorado/06/2017-like (Victoria lineage) virus (updated) |  |
| **Teh, 2021^34^** | Community | S 2019 | Tx1 | IIV3-HD | Fluzone HD | A/Michigan/45/2015 (H1N1)pdm09–like virus, A/Switzerland/8060/2017 (H3N2)–like virus, B/Phuket/3073/2013-like virus (Yamagata lineage) | Circulating strains:  Influenza A viruses (A/H3N2, A/H1N1) and Influenza B viruses |
|  |  |  | Tx2 | IIV4-SD | FluQuadri SD | A/Michigan/45/2015 (H1N1)pdm09–like virus, A/Switzerland/8060/2017 (H3N2)–like virus, B/Phuket/3073/2013-like virus (Yamagata lineage), B/Colorado/06/2017-like virus (Victoria lineage) |  |
| **Vardeny, 2021^35^** | NR | N 2016/2017,  N 2017/2018, and N 2018/2019 | Tx1 | IIV3-HD | Fluzone HD | 2017/2018: A/Michigan/45/2015 (H1N1)pdm09-like virus, an A/Hong Kong/4801/2014 (H3N2)-like virus, and a B/Brisbane/60/2008-like (B/Victoria lineage) virus  2018/2019: A/Michigan/45/2015 (H1N1)pdm09-like virus, A/Singapore/INFIMH-16-0019/2016 A(H3N2)-like virus (updated), B/Colorado/06/2017-like (Victoria lineage) virus (updated) | Predominant circulating strain:  influenza A strain viruses (specifically influenza A/New York/55/2004 [H3N2] and novel influenza A [H1N1])  Other circulating strain: B/Yamagata strain |
|  |  |  | Tx2 | IIV4-SD | Fluzone Quadrivalent SD | 2017/2018: A/Michigan/45/2015 (H1N1)pdm09-like virus, an A/Hong Kong/4801/2014 (H3N2)-like virus, and a B/Brisbane/60/2008-like (B/Victoria lineage) virus  2018/2019: A/Michigan/45/2015 (H1N1)pdm09-like virus, A/Singapore/INFIMH-16-0019/2016 A(H3N2)-like virus (updated), B/Colorado/06/2017-like (Victoria lineage) virus (updated) |  |
| **Studies included only in descriptive analysis (n= 15)** | | | | | | | |
| **Treanor, 1994^36^** | Community | NR | Tx1 | IIV3-SD | Trivalent inactivated influenza vaccine SD | A/H1N1; A/H3N2; B/Ann Arbor/86 strain (1989 to 1990 flu vaccine) OR B/Yamgata/88 | NR |
|  |  |  | Tx3 | Placebo | Placebo | NA |  |
| **de Bruijn, 2006^37^** | Community | N 2004/2005 | Tx1 | IIV3-Adj | Fluad, adjuvanted | A/New Caledonia/20/99(H1N1)-like virus, an A/Fujian/411/2002(H3N2)-like virus and a B/Shanghai/361/2002-like virus | Predominant circulating strain:  A/H3N2 |
|  |  |  | Tx2 | IIV3-SD | Influvac subunit vaccine SD | A/New Caledonia/20/99(H1N1)-like virus, an A/Fujian/411/2002(H3N2)-like virus and a B/Shanghai/361/2002-like virus |  |
| **Szymczakiewicz-Multanowska, 2009^38^** | Community | N 2004/2005 | Tx1 | IIV3-SD | Agrippal SD | A/New Caledonia/20/99(H1N1)-like, A/Fujian/411/2002(H3N2)-like, and B/Shanghai/361/2002(B)-like | Predominant circulating strain:  A/H3N2 |
|  |  |  | Tx2 | IIV3-SD-CC | Trivalent inactivated influenza vaccine, cell-cultured, SD | A/New Caledonia/20/99(H1N1)-like, A/Fujian/411/2002(H3N2)-like, and B/Shanghai/361/2002(B)-like |  |
| **Szymczakiewicz-Multanowska, 2012 (a)^39^** | Community | N 2005/2006 | Tx1 | IIV3-SD-CC | Optaflu, cell-cultured SD | A/New Caledonia/20/99-like, A/California/7/2004-like, B/Shanghai/361/2002-like | Predominant circulating strain: Influenza B viruses  Other circulating strains: A/H1N1 and A/H3N2 |
|  |  |  | Tx2 | IIV3-SD | Agrippal SD | A/Solomon Islands/3/2006-like, A/Wisconsin/67/2005-like, B/Malaysia/2506/2004-like |  |
| **Szymczakiewicz-Multanowska, 2012 (b)^40^** | Community | N 2007/2008 | Tx1 | IIV3-SD-CC | Optaflu, cell-cultured, SD | A/Solomon Islands/3/2006-like, A/Wisconsin/67/2005-like, B/Malaysia/2506/2004-like | Predominant circulating strain: A/H1N1  Other circulating strains: A/H3N2, and B viruses |
|  |  |  | Tx2 | IIV3-SD | Agrippal SD | A/Solomon Islands/3/2006-like, A/Wisconsin/67/2005-like, B/Malaysia/2506/2004-like |  |
| **Della Cioppa, 2012^41^** | NR | N 2008/2009 | Tx1 | IIV3-Adj | Trivalent inactivated influenza vaccine, adjuvanted | A/Brisbane/59/2007 (H1N1), A/Uruguay/716/ 2007 (H3N2) and B/Florida/4/2006 | Predominant circulating strain: A/H3 and A/H3N2  Other circulating strains: Type B viruses |
|  |  |  | Tx2 | IIV3-Other-Adj | Trivalent inactivated influenza vaccine, adjuvanted | A/Brisbane/59/2007 (H1N1), A/Uruguay/716/ 2007 (H3N2) and B/Florida/4/2006 |  |
|  |  |  | Tx3 | IIV3-Adj | Trivalent inactivated influenza vaccine, adjuvanted | A/Brisbane/59/2007 (H1N1), A/Uruguay/716/ 2007 (H3N2) and B/Florida/4/2006 |  |
|  |  |  | Tx4 | IIV3-Other-Adj | Trivalent inactivated influenza vaccine, adjuvanted | A/Brisbane/59/2007 (H1N1), A/Uruguay/716/ 2007 (H3N2) and B/Florida/4/2006 |  |
| **Della Cioppa, 2014^42^** | NR | N 2008/2009 | Tx1 | IIV3-SD | Trivalent inactivated influenza vaccine SD | A/Brisbane/59/2007 (H1N1), A/Uruguay/716/2007 (H3N2), B/Florida/4/2006 | Predominant circulating strain: A/H3 and A/H3N2  Other circulating strains: Type B viruses |
|  |  |  | Tx2 | IIV3-Other | Trivalent inactivated influenza vaccine | A/Brisbane/59/2007 (H1N1), A/Uruguay/716/2007 (H3N2), B/Florida/4/2006 |  |
|  |  |  | Tx3 | IIV3-Adj | Trivalent inactivated influenza vaccine, adjuvanted | A/Brisbane/59/2007 (H1N1), A/Uruguay/716/2007 (H3N2), B/Florida/4/2006 |  |
|  |  |  | Tx4 | IIV3-Other-Adj | Trivalent inactivated influenza vaccine, adjuvanted | A/Brisbane/59/2007 (H1N1), A/Uruguay/716/2007 (H3N2), B/Florida/4/2006 |  |
| **Izikson, 2015^43^** | NR | N 2012/2013 | Tx1 | RIV3 | Flublok | A/H1N1:A/California/07/2009, A/H3N2:A/Victoria/361/2011, B/Wisconsin/1/2010 | Predominant circulating strain:  A/H3N2 |
|  |  |  | Tx2 | IIV3-SD | Afluria SD | A/H1N1:A/California/07/2009, A/H3N2:A/Victoria/361/2011, B/Wisconsin/1/2010 |  |
| **Novartis Vaccines and Diagnostics, 2016^44^** | NR | NR | Tx1 | IIV3-SD | Agriflu SD | NR | NR |
|  |  |  | Tx2 | IIV3-SD | Fluvirin SD | NR |  |
| **Trial registry, 2017^45^** | NR | N 2011/2012 | Tx1 | IIV3-SD | Fluzone SD | A/California/7/2009-like (H1N1), A/Perth/16/2009-like (H3N2), and B/Brisbane/60/2008-like viruses | Predominant circulating strain:  A/H3N2 |
|  |  |  | Tx2 | IIV3-HD | Fluzone HD | A/California/7/2009-like (H1N1), A/Perth/16/2009-like (H3N2), and B/Brisbane/60/2008-like viruses |  |
| **Otten, 2020^46^** | Community | N 2014/2015 | Tx1 | IIV3-Adj | Agrippal/Fluad, adjuvanted | A/California/7/2009 (H1N1) pdm09-like virus, A/Texas/50/2012 (H3N2)-like virus, and B/Massachusetts/2/2012-like virus | Predominant circulating strain: A/H3N2 |
|  |  |  | Tx2 | IIV3-Adj | Agrippal/Fluad, adjuvanted | A/California/7/2009 (H1N1) pdm09-like virus, A/Texas/50/2012 (H3N2)-like virus, and B/Massachusetts/2/2012-like virus |  |
|  |  |  | Tx3 | IIV3-Other-Adj | Agrippal/Fluad, adjuvanted | A/California/7/2009 (H1N1) pdm09-like virus, A/Texas/50/2012 (H3N2)-like virus, and B/Massachusetts/2/2012-like virus |  |
|  |  |  | Tx4 | IIV3-Other-Adj – bilateral | Agrippal/Fluad, adjuvanted | A/California/7/2009 (H1N1) pdm09-like virus, A/Texas/50/2012 (H3N2)-like virus, and B/Massachusetts/2/2012-like virus |  |
|  |  |  | Tx5 | IIV3-Adj-bilataeral + saline | Agrippal/Fluad, adjuvanted | A/California/7/2009 (H1N1) pdm09-like virus, A/Texas/50/2012 (H3N2)-like virus, and B/Massachusetts/2/2012-like virus |  |
|  |  |  | Tx6 | IIV3-Adj- bilateral + saline | Agrippal/Fluad, adjuvanted | A/California/7/2009 (H1N1) pdm09-like virus, A/Texas/50/2012 (H3N2)-like virus, and B/Massachusetts/2/2012-like virus |  |
|  |  |  | Tx7 | IIV3-Adj-bilateral | Agrippal/Fluad, adjuvanted | A/California/7/2009 (H1N1) pdm09-like virus, A/Texas/50/2012 (H3N2)-like virus, and B/Massachusetts/2/2012-like virus |  |
| **McConeghy, 2020^47^** | Nursing home | N 2016/2017 | Tx1 | IIV3-Adj | Fluad, adjuvanted | A/California/7/2009 (H1N1)–like virus, an A/Hong Kong/ 4801/2014 (H3N2)–like virus, and a B/Brisbane/60/2008–like virus (Victoria lineage) | Predominant circulating strain: A/H3N2 |
|  |  |  | Tx2 | IIV3-SD | Fluvirin SD | A/California/7/2009 (H1N1)–like virus, an A/Hong Kong/ 4801/2014 (H3N2)–like virus, and a B/Brisbane/60/2008–like virus (Victoria lineage) |  |
| **Sanchez, 2020^48^** | Clinic | N 2017/2018 | Tx1 | IIV4-HD | Quadrivalent inactivated influenza vaccine HD | A/Michigan/45/2015 (NYMC X-275), A/Hong Kong/4801/2014 (NYMC X-263B), B/Phuket/3073/2013, B/Brisbane/60/2008 | Predominant circulating strain:  A/H3N2 |
|  |  |  | Tx2 | IIV4-HD | Quadrivalent inactivated influenza vaccine HD | A/Michigan/45/2015 (NYMC X-275), A/Hong Kong/4801/2014 (NYMC X-263B), B/Phuket/3073/2013, B/Brisbane/60/2008 |  |
|  |  |  | Tx3 | IIV4-SD | Quadrivalent inactivated influenza vaccine SD | A/Singapore/GP1908/ 2015 (IVR-180) pdm09 (A/H1N1-like), A/Hong Kong/4801/2014 (NYMC X-263) (A/H3N2-like), B/Phuket/3073/2013, B/Texas/2/2013 (B/Victoria lineage-like) |  |
| **McLean, 2021^49^** | Community | N 2016/2017 and  N 2017/2018 | Tx1 | IIV3-HD (2015/2016, 2016/2017) | Fluzone HD in 2015/2016 and 2016/2017 | 2016/2017: A/California/7/2009(H1N1)-like, A/Hong Kong/4801/2014(H3N2)-like virus, B/Brisbane/60/2008(B/Victoria)-like virus.  2017/2018:  A/Michigan/45/2015(H1N1)-like virus, A/Hong Kong/4801/2014(H3N2)-like virus, /Brisbane/60/2008(B/Victoria)-like virus | Predominant circulating strain:  A/H3N2; B/Yamagata-lineage |
|  |  |  | Tx2 | IIV3-Adj (2015/2016, 2016/2017) | Fluad in 2015/2016 and 2016/2017 | 2016/2017: A/California/7/2009(H1N1)-like, A/Hong Kong/4801/2014(H3N2)-like virus, B/Brisbane/60/2008(B/Victoria)-like virus.  2017/2018:  A/Michigan/45/2015(H1N1)-like virus, A/Hong Kong/4801/2014(H3N2)-like virus, /Brisbane/60/2008(B/Victoria)-like virus |  |
|  |  |  | Tx3 | IIV3-SD (2015/2016) + IIV3-HD (2016/2017) | Fluvirin SD in 2015/2016 and Fluzone HD in 2016/2017 | 2016/2017: A/California/7/2009(H1N1)-like, A/Hong Kong/4801/2014(H3N2)-like virus, B/Brisbane/60/2008(B/Victoria)-like virus.  2017/2018:  A/Michigan/45/2015(H1N1)-like virus, A/Hong Kong/4801/2014(H3N2)-like virus, /Brisbane/60/2008(B/Victoria)-like virus |  |
|  |  |  | Tx4 | IIV3-SD (2015/2016) + IIV3-Adj (2016/2017) | Fluvirin SD in 2015/2016 and Fluad, adjuvanted in 2016/2017 | 2016/2017: A/California/7/2009(H1N1)-like, A/Hong Kong/4801/2014(H3N2)-like virus, B/Brisbane/60/2008(B/Victoria)-like virus.  2017/2018:  A/Michigan/45/2015(H1N1)-like virus, A/Hong Kong/4801/2014(H3N2)-like virus, /Brisbane/60/2008(B/Victoria)-like virus |  |
| **Trial registry, 2021^50^** | Community | N 2020/2021 | Tx1 | IIV4-HD | Quadrivalent inactivated influenza vaccine HD | A/Guangdong-Maonan/SWL1536/2019 CNIC-1909 (H1N1), A/Hong Kong/2671/2019 IVR-208 (H3N2), B/Washington/02/2019 wt, B/Phuket/3073/2013 wt | NR |
|  |  |  | Tx2 | IIV4-SD | Quadrivalent inactivated influenza vaccine SD | A/(H1N1)-like strain, A/(H3N2)-like strain, B/(Victoria lineage)-like strain, B/(Yamagata lineage)-like strain |  |
| Note: The term "-Other" in the vaccine name indicates that there are varying dosages available for different strains. As such, it is not categorized as standard dose nor high dose as per guidelines.  **Abbreviations**- NR: Not reported; N: Northern hemisphere; S: Southern hemisphere; IIV3: Trivalent **i**nactivated influenza vaccine; IIV4: Quadrivalent **i**nactivated influenza vaccine; Tdap: Tetanus, diphtheria, pertussis; Adj: Adjuvanted; SD: Standard dosage; HD: High dosage; CC: Cell-cultured; RIV: Recombinant influenza vaccine; LAIV: Live attenuated influenza vaccine | | | | | | | |

### **Appendix 26: Antigenic characterization of viral strains for studies reporting laboratory-confirmed influenza**

| **Author, year** | **Vaccine type** | **Vaccine composition** | **Type of laboratory-confirmed influenza used in the analysis** | **Circulating strains** | **Antigenic characterization as per author** | **Classification of laboratory-confirmed influenza viral strains as being matched** | **Classification of laboratory-confirmed influenza viral strains as being mismatched** |
| --- | --- | --- | --- | --- | --- | --- | --- |
| Rudenko, 2001^10^ | IIV3-SD | LAIV: A/Leningrad/134/17/57 (H3N2) or A/Texas/36/91(H1N1), A/Nanchang/933/95 (H3N2), B/Harbin/07/94 (Yamagata) and B/Ann Arbor/1/86 (Victoria);  TIV: A/Texas/36/91 (H1N1), A/Nanchang/933/95 (H3N2), B/Harbin/07/94 (Yamagata) | Culture or HI assay (secondary analysis) | Viral isolates similar to A/Texas/36/91 (H1N1), or B/Harbin/07/94 | Typed by HI (cut-off: fourfold rise) to vaccine strains | A/Texas/36/91 (H1N1), B/Harbin/07/94 (Yamagata)  (reported as matched) | NA |
| Wongsurakiat, 2004^11^ | IIV3-SD | A/Texas/36/91 (H1N1), A/Nanchang/933/95 (H3N2), and B/Harbin/  07/94 | HI test | A/Sydney/5/97 (H3N2) | Typed by HI (cut-off: fourfold rise) to vaccine strains | A/Sydney/5/97 (H3N2)  (closely related to the influenza  A/Nanchang/933/96 (H3N2))  (reported as matched) | NA |
| Keitel, 2010^14^ | IIV3-SD, RIV3 | IIV3-SD: A/Wisconsin (H3N2); A/New Caledonia (H1N1) and B/Malaysia;  RIV3: A/Wisconsin (H3N2); A/New Caledonia (H1N1) and B/Ohio | Culture | A/New Caledonia/20/99 (H1N1) (From CDC) | Typed by HI (cut-off: fourfold rise) to vaccine strains | A/New Caledonia/20/99  (Classified as matched) | NA |
| DiazGranados, 2013^15^ | IIV3 | A/Brisbane/59/07 (H1N1), A/Uruguay/716/2007 X-175C (H3N2), and B/Brisbane/60/2008 strains | PCR | A/California/7/2009 (H1N1) | Seroprotection was  considered as a post-vaccination HAI titer ≥1:40 | NA | A/California/7/2009 (H1N1) (reported as genetically or antigenically distant) |
| DiazGranados, 2014^20^ | IIV3 | 2011/2012:  A/California/ 7/2009 (H1N1), A/Victoria/210/2009 (H3N2) and B/Brisbane/60/2008;  2012/2013:  A/California/7/2009 (H1N1), A/Victoria/361/2011 (H3N2), B/Texas/6/2011 (B/Wisconsin/1/2010-like virus) | Culture or PCR assay or both | 2011/2012 and 2012/2013: A/H3N2  (From CDC) | Typed by HI (cut-off: fourfold rise) to vaccine strains (from appendix) | 2011/2012: A/Perth/16/2009-like (Reported as matched) | 2012/2013: A/Victoria/361/2011 (Reported as mismatch) |
| Dunkle, 2017^24^ | RIV4, IIV4 | A/California/  7/2009 (H1N1)-like, A/Texas/50/2012 (H3N2), B/ Massachusetts/2/2012, and B/Brisbane/60/2008 | RT-PCR assay and culture | influenza A subtype  H3N2 viruses | Antigenic characterization not actively performed; based on predominant epidemic viruses  or HAI titers ≥40 | NA | A/Switzerland/9715293/2013 (Classified as mismatched) |
| Loeb, 2020^28^ | IIV3 | 2014/2015: A/Texas/50/2012 (H3N2), A/California/7/2009 (H1N1), and  B/Massachusetts/2/2012  2015/2016: A/Switzerland/9715292-2013,  A/California/7/2009 (H1N1), and B/Phuket/3073/2013  2016/2017: A/Hong Kong/4801-2014 (H3N2), A/California/7/2009  NYNC X-179A (H1N1), and B/Brisbane/60/2008  2017/2018: A/HongKong/4801/2014 (H3N2), A/Michigan/45/2015  (H1N1), and B/Brisbane/60/2008 | PCR | 2014/2015: A/H3N2; 2015/2016: A/H1N1pdm09; 2016/2017: A/H3N2; 2017/2018: A/H3N2 | Typed by HI (cut-off: fourfold rise) to vaccine strains | 2015/2016: A/California/7/2009 (Classified as matched)  2016/2017: A/Hong Kong/4801/2014 (Classified as matched)  2017/2018: A/Hong Kong/4801/2014 (the A(H3N2) (Classified as matched) | 2014/2015: A/Texas/50/2012-like (Classified as mismatched) |
| Belongia, 2020^29^ | HD-IIV3, aIIV3, RIV4 | The A(H3N2) recommended vaccine component was identical in 2016–17 and 2017–18: A/Hong Kong/4801/2014–like (clade 3C.2a) | RT-PCR | A/Singapore/INFIMH-16–0019/2016 and A/Kentucky/29/2017 and A/Kansas/14/2017 (KS14) | Typed by HI (cut-off: fourfold rise) to vaccine strains | Circulating A(H3N2) viruses were antigenically similar to the cell-grown vaccine  reference virus. (reported as matched) | NA |
| Teh, 2021^34^ | IIV3, IIV4 | TIV: A/Michigan/45/2015 (H1N1)pdm09–  like virus, A/Switzerland/8060/2017 (H3N2)–like virus and B/  Phuket/3073/2013-like virus (Yamagata lineage)  QIV: A/Michigan/45/2015 (H1N1)pdm09–  like virus, A/Switzerland/8060/2017 (H3N2)–like virus, B/  Phuket/3073/2013-like virus (Yamagata lineage) and B/Colorado/06/2017-like virus (Victoria lineage) | PCR | A(H3N2) | Typed by HI (cut-off: fourfold rise) to vaccine strains | A(H1N1)pdm09 viruses and viruses from both influenza B lineages (Classified as matched) | NA |
| McLean, 2021^49^ | IIV3-HD, IIV3-SD, IIV3-Adj | 2016/2017  A/California/7/2009(H1N1)-like, A/Hong Kong/4801/2014(H3N2)-like, and B/Brisbane/60/2008(B/Victoria)-like viruses  2017/2018 A/Michigan/45/2015(H1N1)-like virus, A/Hong Kong/4801/2014(H3N2)-like, and  B/Brisbane/60/2008(B/Victoria)-like viruses | RT-PCR | A/H3N2; B/Yamagata-lineage | Typed by HI (cut-off: fourfold rise) to vaccine strains | NA | 2016/2017 and 2017 and 2018  (reported as mismatch) |
| *****The process of characterizing matched and mismatched vaccine strains involved two steps. In Step 1, viral strains from laboratory-confirmed influenza cases within the included RCTs were characterized using HI assays and reference vaccine strains. Strains were considered matched if they displayed antigenic similarity to the vaccine strain. In Step 2, when antigenic data was lacking, surveillance data from sources such as WHO and CDC were used to identify prevalent strains during the trial period. For multiple years, if there was a clear prevalence of matched over mismatched (or vice versa) - for instance a 3 year long study in which 1 year was mismatched and 2 years were matched, the study was characterized as matched. If a study where year 1 was mismatched and year 2 was matched, we characterized it as matched and mismatched in two separate analyses. When the matched or mismatched strains were presented in the study by the authors, they are listed as ‘reported’. When the matched or mismatched strains were determined from surveillance data sources they are listed as ‘classified’.  **Abbreviations-** NA: Not applicable; IIV3: Trivalent **i**nactivated influenza vaccine; IIV4: Quadrivalent **i**nactivated influenza vaccine; SD: Standard dosage; HD: High dosage; Adj: Adjuvanted; RIV: Recombinant influenza vaccine; LAIV: Live attenuated influenza vaccine; HI: Hemagglutination-inhibition; PCR: Polymerase chain reaction; RT-PCR: Reverse transcription polymerase chain reaction; CDC: Centers for disease control and prevention | | | | | | | |

### **Appendix 27: Antigenic characterization of viral strains for studies reporting influenza-like illness**

| **Author, year** | **Vaccine type** | **Vaccine composition** | **Influenza like illness definition** | **Circulating strains** | **Classification of influenza viral strains as being matched** | **Classification of viral strains as being mismatched** |
| --- | --- | --- | --- | --- | --- | --- |
| Wongsurakiat, 2004^11^ | IIV3-SD | A/Texas/36/91 (H1N1), A/Nanchang/933/95 (H3N2), and B/Harbin/  07/94 | Defined when the patients had symptoms of generalized aches, fever, and headache with or without upper respiratory tract symptoms. | A/Sydney/5/97 (H3N2) | A/Sydney/5/97 (H3N2)  (closely related to the influenza  A/Nanchang/933/96 (H3N2))  (reported as matched) | NA |
| Allsup, 2004^12^ | IIV3 | A/Beijing/262/95(H1N1), A/Sydney/5/97 (H3N2) and B/Beijing/184/93 | Defined as an illness of sudden onset with features of cough, feverishness, prostration, myalgia and widespread aches and pains. | A/H3N2 | A (H3N2) (Reported as match) | NA |
| Keitel, 2010^14^ | IIV3-SD, RIV3 | IIV3-SD: A/Wisconsin (H3N2); A/New Caledonia (H1N1) and B/Malaysia;  RIV3: A/Wisconsin (H3N2); A/New Caledonia (H1N1) and B/Ohio | Subject met CDC-ILI definition (fever with cough and/or sore throat) and/or if the subject had sought medical care at another location (medically attended acute respiratory illness, or MAARI) | A/New Caledonia/20/99 (H1N1) (From CDC) | A/New Caledonia/20/99  (Classified as matched) | NA |
| DiazGranados, 2013^15^ | IIV3 | A/Brisbane/59/07 (H1N1), A/Uruguay/716/2007 X-175C (H3N2), and B/Brisbane/60/2008 strains | Defined as a new onset (or exacerbation of a pre-existing condition) of at least one of the following systemic symptoms: temperature >37.2 ◦C (>99.0 ◦F), feverishness (feeling of warmth), chills, tiredness, headaches or myalgia; and at least one of the following respiratory symptoms: nasal congestion or rhinorhea, sore throat, cough, sputum production, wheezing, chest tightness, shortness of breath, or chest pain with breathing. | A/California/7/2009 (H1N1) | NA | A/California/7/2009 (H1N1) (reported as genetically or antigenically distant) |
| Frey, 2014^19^ | IIV3 | A/California/7/2009 (H1N1), A/Perth/16/2009 (H3N2) and B/Brisbane/60/2008 | Defined as temperature of ≥37.2 ◦C or feverishness and at least two of the following symptoms: headache, myalgia, cough, or sore throat | A/H3N2 and A/H1N1 | A/California/7/2009-like and A/Perth/16/2009-like (Classified as matched) | NA |
| DiazGranados, 2014^20^ | IIV3 | 2011/2012:  A/California/ 7/2009 (H1N1), A/Victoria/210/2009 (H3N2) and B/Brisbane/60/2008;  2012/2013:  A/California/7/2009 (H1N1), A/Victoria/361/2011 (H3N2) and B/Texas/6/2011 (B/Wisconsin/1/2010-like virus) | Defined as a respiratory illness with sore throat, cough, sputum production, wheezing, or difficulty breathing, concurrent with one or more of the following: temperature above 37.2°C, chills, tiredness, headaches, or myalgia. | 2011/2012 and 2012/2013: A/H3N2  (From CDC) | 2011/2012: A/Perth/16/2009-like (Reported as matched) | 2012/2013: A/Victoria/361/2011 (Reported as mismatch) |
| Dunkle, 2017^24^ | RIV4, IIV4 | A/California/  7/2009 (H1N1)-like, A/Texas/50/2012 (H3N2), B/ Massachusetts/2/2012, and B/Brisbane/60/2008 | Defined in the protocol as at least one symptom in both the respiratory and systemic illness categories, regardless of severity. | influenza A subtype  H3N2 viruses | NA | A/Switzerland/9715293/2013 (Classified as mismatched) |
| Teh, 2021^34^ | IIV3, IIV4 | TIV: A/Michigan/45/2015 (H1N1)pdm09–  like virus, A/Switzerland/8060/2017 (H3N2)–like virus and B/  Phuket/3073/2013-like virus (Yamagata lineage)  QIV: A/Michigan/45/2015 (H1N1)pdm09–  like virus, A/Switzerland/8060/2017 (H3N2)–like virus, B/  Phuket/3073/2013-like virus (Yamagata lineage) and B/Colorado/06/2017-like virus (Victoria lineage) | Defined by the presence of fever (≥38°C) and at least 1 respiratory symptom. | A(H3N2) | A(H1N1)pdm09 viruses and viruses from both influenza B lineages (Classified as matched) | NA |
| *****The process of characterizing matched and mismatched vaccine strains involved two steps. In Step 1, viral strains from influenza cases within the included RCTs were characterized using HI assays and reference vaccine strains. Strains were considered matched if they displayed antigenic similarity to the vaccine strain. In Step 2, when antigenic data was lacking, surveillance data from sources such as WHO and CDC were used to identify prevalent strains during the trial period. For multiple years, if there was a clear prevalence of matched over mismatched (or vice versa) - for instance a 3 year long study in which 1 year was mismatched and 2 years were matched, the study was characterized as matched. If a study where year 1 was mismatched and year 2 was matched, we characterized it as matched and mismatched in two separate analyses. When the matched or mismatched strains were presented in the study by the authors, they are listed as ‘reported’. When the matched or mismatched strains were determined from surveillance data sources they are listed as ‘classified’.  **Abbreviations-** NA: Not applicable; IIV3: Trivalent **i**nactivated influenza vaccine; IIV4: Quadrivalent **i**nactivated influenza vaccine; SD: Standard dosage; HD: High dosage; Adj: Adjuvanted; RIV: Recombinant influenza vaccine; LAIV: Live attenuated influenza vaccine; HI: Hemagglutination-inhibition; PCR: Polymerase chain reaction; RT-PCR: Reverse transcription polymerase chain reaction; CDC: Centers for disease control and prevention; MAARI: medically attended acute respiratory illness | | | | | | |

### **Appendix 28: Additional Analyses**

**Figure 1.** Network plots for the network meta-analyses with combined coding of interventions of a) laboratory-confirmed influenza, b) influenza-like illness.

**a)**
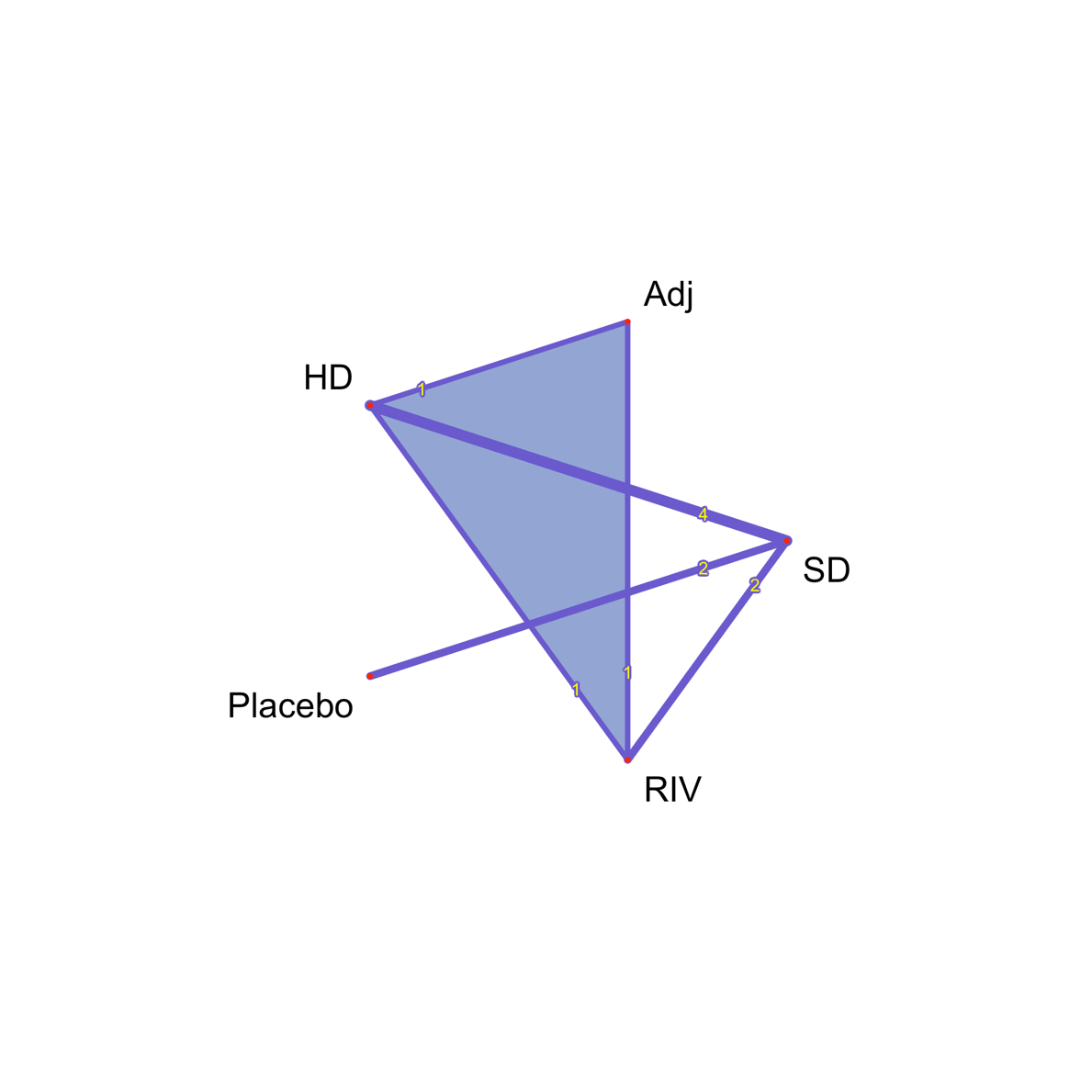
 **b)**
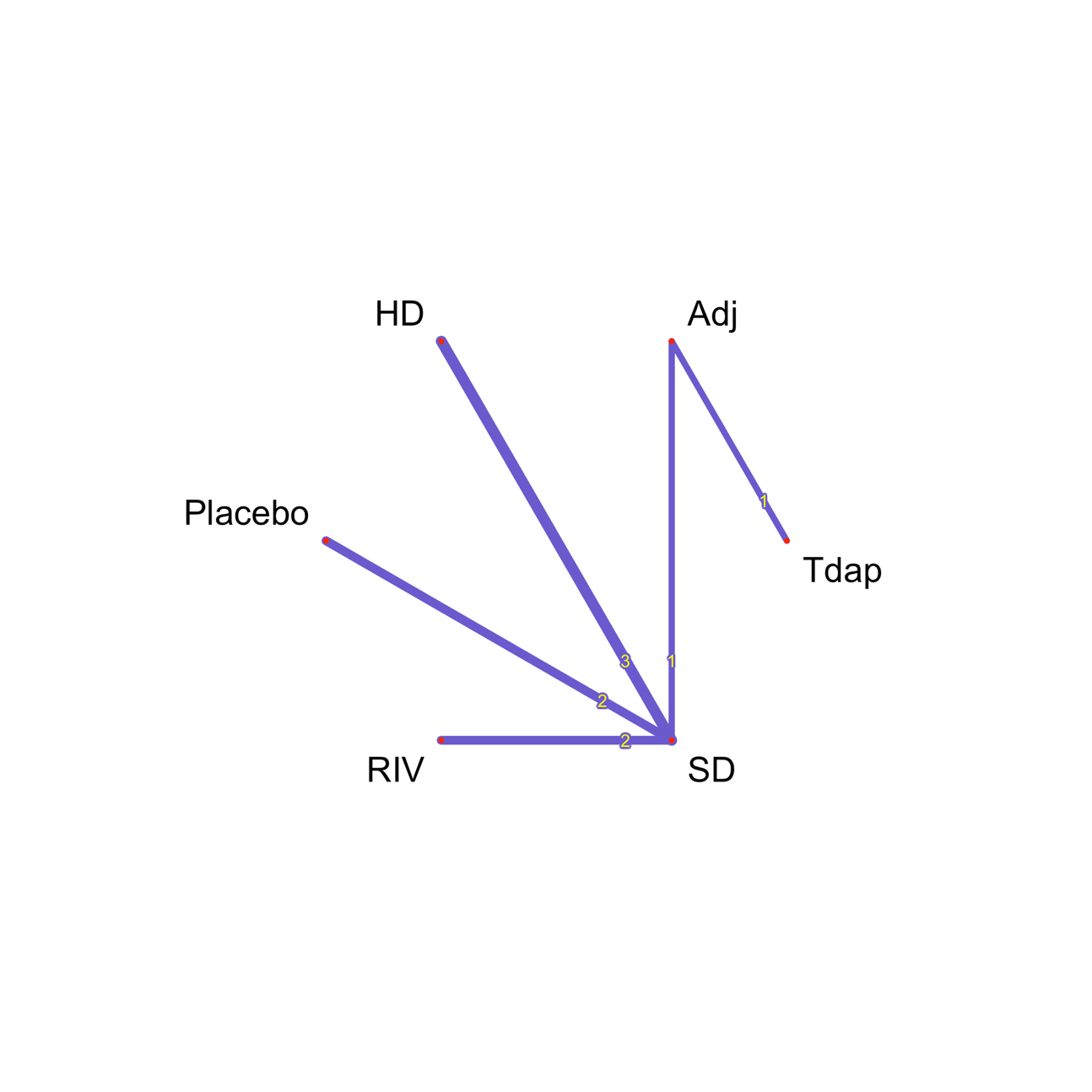

**Abbreviations-** Adj: Adjuvanted; SD: Standard dosage; HD: High dosage; RIV: Recombinant influenza vaccine; Tdap: Tetanus, diphtheria, pertussis

#### **Appendix 28A: Sensitivity Analyses**

**Laboratory-confirmed influenza**

**Table 1.** Sensitivity Analysis: Summary of network meta-analysis results for laboratory-confirmed influenza with original coding of interventions, restricted to studies in which the proportion of females was at least 50%.

| ***Network Meta-analysis results; 7 studies, 52,009 patients, 6 treatments*** | | | | | | | | |
| --- | --- | --- | --- | --- | --- | --- | --- | --- |
| ***Comparison*** | ***OR*** | ***seOR*** | ***Lower CI*** | ***Upper CI*** | ***Statistic*** | ***p-value*** | ***Lower PI*** | ***Upper PI*** |
| IIV3-Adj: IIV3-HD | 1.15 | 2.32 | 0.22 | 5.98 | 0.17 | 0.86 | 0.03 | 42.79 |
| IIV3-Adj: IIV3-SD | 0.85 | 2.31 | 0.17 | 4.41 | -0.19 | 0.85 | 0.02 | 31.46 |
| IIV3-Adj: IIV4-SD | 0.66 | 2.20 | 0.14 | 3.08 | -0.53 | 0.59 | 0.02 | 19.59 |
| IIV3-Adj: Placebo | 0.39 | 2.67 | 0.06 | 2.70 | -0.95 | 0.34 | 0.01 | 26.88 |
| IIV3-Adj: RIV | 0.95 | 2.18 | 0.21 | 4.37 | -0.06 | 0.95 | 0.03 | 27.01 |
| IIV3-HD: IIV3-SD | 0.74 | 1.09 | 0.63 | 0.87 | -3.56 | 0.00 | 0.51 | 1.07 |
| IIV3-HD: IIV4-SD | 0.57 | 1.56 | 0.24 | 1.36 | -1.27 | 0.20 | 0.08 | 3.84 |
| IIV3-HD: Placebo | 0.34 | 1.68 | 0.12 | 0.94 | -2.08 | 0.04 | 0.04 | 3.16 |
| IIV3-HD: RIV | 0.83 | 1.53 | 0.36 | 1.89 | -0.46 | 0.65 | 0.13 | 5.08 |
| IIV3-SD: IIV4-SD | 0.77 | 1.55 | 0.33 | 1.81 | -0.60 | 0.55 | 0.12 | 5.05 |
| IIV3-SD: Placebo | 0.46 | 1.66 | 0.17 | 1.26 | -1.51 | 0.13 | 0.05 | 4.14 |
| IIV3-SD: RIV | 1.11 | 1.52 | 0.49 | 2.52 | 0.26 | 0.79 | 0.19 | 6.68 |
| IIV4-SD: Placebo | 0.60 | 1.96 | 0.16 | 2.24 | -0.76 | 0.45 | 0.03 | 10.81 |
| IIV4-SD: RIV | 1.45 | 1.14 | 1.11 | 1.89 | 2.76 | 0.01 | 0.81 | 2.59 |
| RIV: Placebo | 0.41 | 1.93 | 0.11 | 1.51 | -1.34 | 0.18 | 0.02 | 7.04 |
| Common within-network between-study SD (tau) | <0.0001 |  |  |  |  |  |  |  |
| I-square | 0% |  |  |  |  |  |  |  |
| **Abbreviations-** IIV3: Trivalent **i**nactivated influenza vaccine; IIV4: Quadrivalent **i**nactivated influenza vaccine; Adj: Adjuvanted; SD: Standard dosage; HD: High dosage; RIV: Recombinant influenza vaccine; OR: Odds ratio; CI: Confidence interval; PI: Prediction interval; se: Standard error | | | | | | | | |

**Figure 1.** Treatment ranking of interventions using P-score for laboratory-confirmed influenza with original coding of interventions, restricted to studies in which the proportion of females was at least 50%.

| ***Treatment Ranking*** | |
| --- | --- |
| ***Treatment*** | ***P-score*** |
| IIV3-HD | 0.82 |
| RIV | 0.66 |
| IIV3-Adj | 0.61 |
| IIV3-SD | 0.50 |
| IIV4-SD | 0.29 |
| Placebo | 0.11 |

**Abbreviations-** IIV3: Trivalent **i**nactivated influenza vaccine; IIV4: Quadrivalent **i**nactivated influenza vaccine; Adj: Adjuvanted; SD: Standard dosage; HD: High dosage; RIV: Recombinant influenza vaccine

**Table 2.** Sensitivity Analysis: Summary of network meta-analysis results for laboratory-confirmed influenza with combined coding of interventions.

| ***Network Meta-analysis results; 9 studies, 52,202 patients, 5 treatments*** | | | | | | | | |
| --- | --- | --- | --- | --- | --- | --- | --- | --- |
| ***Comparison*** | ***OR*** | ***seOR*** | ***Lower CI*** | ***Upper CI*** | ***Statistic*** | ***p-value*** | ***Lower PI*** | ***Upper PI*** |
| Adj: HD | 0.96 | 2.18 | 0.21 | 4.42 | -0.06 | 0.95 | 0.13 | 7.12 |
| Adj: Placebo | 0.22 | 2.38 | 0.04 | 1.22 | -1.73 | 0.08 | 0.02 | 2.08 |
| Adj: RIV | 1.00 | 2.17 | 0.22 | 4.56 | 0.00 | 1.00 | 0.14 | 7.31 |
| Adj: SD | 0.71 | 2.18 | 0.15 | 3.27 | -0.44 | 0.66 | 0.10 | 5.25 |
| HD: Placebo | 0.23 | 1.48 | 0.11 | 0.50 | -3.69 | 0.00 | 0.08 | 0.64 |
| HD: RIV | 1.05 | 1.17 | 0.78 | 1.42 | 0.32 | 0.75 | 0.71 | 1.55 |
| HD: SD | 0.74 | 1.09 | 0.63 | 0.88 | -3.53 | 0.00 | 0.60 | 0.92 |
| RIV: Placebo | 0.22 | 1.50 | 0.10 | 0.49 | -3.70 | 0.00 | 0.08 | 0.63 |
| SD: Placebo | 0.31 | 1.47 | 0.15 | 0.67 | -3.01 | 0.00 | 0.12 | 0.85 |
| RIV: SD | 0.71 | 1.14 | 0.55 | 0.91 | -2.70 | 0.01 | 0.51 | 0.98 |
| Common within-network between-study SD (tau) | <0.0001 |  |  |  |  |  |  |  |
| I-square | 0% |  |  |  |  |  |  |  |
| **Abbreviations-** Adj: Adjuvanted; SD: Standard dosage; HD: High dosage; RIV: Recombinant influenza vaccine; OR: Odds ratio; CI: Confidence interval; PI: Prediction interval; se: Standard error | | | | | | | | |

**Figure 2.** Treatment ranking of interventions using P-score for laboratory-confirmed influenza with combined coding of interventions..

| ***Treatment Ranking*** | |
| --- | --- |
| ***Treatment*** | ***P-score*** |
| RIV | 0.78 |
| HD | 0.71 |
| Adj | 0.66 |
| SD | 0.33 |
| Placebo | 0.01 |

**Abbreviations-** Adj: Adjuvanted; SD: Standard dosage; HD: High dosage; RIV: Recombinant influenza vaccine

**Table 3.** Sensitivity Analysis: Summary of network meta-analysis results for laboratory-confirmed influenza with original coding of interventions, restricted to matched studies

| ***Network Meta-analysis results; 7 studies, 52,009 patients, 6 treatments*** | | | | | | | | |
| --- | --- | --- | --- | --- | --- | --- | --- | --- |
| ***Comparison*** | ***OR*** | ***seOR*** | ***Lower CI*** | ***Upper CI*** | ***Statistic*** | ***p-value*** | ***Lower PI*** | ***Upper PI*** |
| IIV3-Adj:IIV3-HD | 1.23 | 2.37 | 0.23 | 6.72 | 0.24 | 0.81 | 0.03 | 54.51 |
| IIV3-Adj:IIV3-SD | 0.85 | 2.36 | 0.16 | 4.60 | -0.18 | 0.85 | 0.02 | 36.84 |
| IIV3-Adj:IIV4-SD | 2.54 | 4.61 | 0.13 | 50.89 | 0.61 | 0.54 | 0.00 | 1899.21 |
| IIV3-Adj:Placebo | 0.27 | 2.58 | 0.04 | 1.71 | -1.39 | 0.16 | 0.00 | 16.81 |
| IIV3-Adj: RIV | 0.94 | 2.21 | 0.20 | 4.46 | -0.07 | 0.94 | 0.03 | 30.79 |
| IIV3-HD: IIV3-SD | 0.69 | 1.19 | 0.50 | 0.97 | -2.17 | 0.03 | 0.25 | 1.92 |
| IIV3-HD: IIV4-SD | 2.06 | 3.53 | 0.17 | 24.40 | 0.57 | 0.57 | 0.01 | 489.55 |
| IIV3-HD: Placebo | 0.22 | 1.55 | 0.09 | 0.51 | -3.50 | 0.00 | 0.03 | 1.61 |
| IIV3-HD: RIV | 0.76 | 1.59 | 0.31 | 1.90 | -0.58 | 0.56 | 0.09 | 6.36 |
| IIV3-SD: IIV4-SD | 0.34 | 3.57 | 0.03 | 4.06 | -0.86 | 0.39 | 0.00 | 83.59 |
| IIV3-SD: Placebo | 3.21 | 1.50 | 1.45 | 7.06 | 2.89 | 0.00 | 0.49 | 20.89 |
| IIV3-SD: RIV | 0.90 | 1.55 | 0.38 | 2.15 | -0.23 | 0.82 | 0.12 | 6.86 |
| IIV4-SD: Placebo | 0.10 | 3.80 | 0.01 | 1.43 | -1.69 | 0.09 | 0.00 | 34.04 |
| IIV4-SD: RIV | 0.37 | 3.83 | 0.03 | 5.15 | -0.74 | 0.46 | 0.00 | 125.18 |
| RIV:Placebo | 3.54 | 1.82 | 1.10 | 11.42 | 2.12 | 0.03 | 0.25 | 51.00 |
| Common within-network between-study SD (tau) | 0.17 |  |  |  |  |  |  |  |
| I-square | 16.8% |  |  |  |  |  |  |  |
| **Abbreviations-** IIV3: Trivalent **i**nactivated influenza vaccine; IIV4: Quadrivalent **i**nactivated influenza vaccine; Adj: Adjuvanted; SD: Standard dosage; HD: High dosage; RIV: Recombinant influenza vaccine; OR: Odds ratio; CI: Confidence interval; PI: Prediction interval; se: Standard error | | | | | | | | |

Note: This analysis incorporates DiazGranados' 2014 study categorized as matched ^20^.

**Figure 3.** Treatment ranking of interventions using P-score for laboratory-confirmed influenza with original coding of interventions, restricted to matched studies

| ***Treatment Ranking*** | |
| --- | --- |
| ***Treatment*** | ***P-score*** |
| IIV4-SD | 0.80 |
| IIV3-HD | 0.72 |
| IIV3-Adj | 0.54 |
| RIV | 0.51 |
| IIV3-SD | 0.41 |
| Placebo | 0.03 |

**Abbreviations-** IIV3: Trivalent **i**nactivated influenza vaccine; IIV4: Quadrivalent **i**nactivated influenza vaccine; Adj: Adjuvanted; SD: Standard dosage; HD: High dosage; RIV: Recombinant influenza vaccine

Note: This analysis incorporates DiazGranados' 2014 study categorized as matched ^20^.

**Table 4.** Sensitivity Analysis: Summary of network meta-analysis results for laboratory-confirmed influenza with original coding of interventions, restricted to matched studies

| ***Network Meta-analysis results; 7 studies, 52,009 patients, 6 treatments*** | | | | | | | | |
| --- | --- | --- | --- | --- | --- | --- | --- | --- |
| ***Comparison*** | ***OR*** | ***seOR*** | ***Lower CI*** | ***Upper CI*** | ***Statistic*** | ***p-value*** | ***Lower PI*** | ***Upper PI*** |
| IIV3-Adj:IIV3-HD | 1.63 | 2.49 | 0.27 | 9.75 | 0.24 | 0.81 | 0.00 | 233156.46 |
| IIV3-Adj:IIV3-SD | 0.75 | 2.41 | 0.13 | 4.20 | -0.18 | 0.85 | 0.00 | 72688.61 |
| IIV3-Adj:IIV4-SD | 3.37 | 4.76 | 0.16 | 71.72 | 0.61 | 0.54 | 0.00 | 1626520650.37 |
| IIV3-Adj:Placebo | 0.23 | 2.64 | 0.03 | 1.57 | -1.39 | 0.16 | 0.00 | 70720.59 |
| IIV3-Adj: RIV | 0.87 | 2.24 | 0.18 | 4.23 | -0.07 | 0.94 | 0.00 | 33527.26 |
| IIV3-HD: IIV3-SD | 0.46 | 1.52 | 0.20 | 1.05 | -2.17 | 0.03 | 0.00 | 176.28 |
| IIV3-HD: IIV4-SD | 2.06 | 3.55 | 0.17 | 24.68 | 0.57 | 0.57 | 0.00 | 24787969.68 |
| IIV3-HD: Placebo | 0.14 | 1.80 | 0.04 | 0.45 | -3.50 | 0.00 | 0.00 | 397.02 |
| IIV3-HD: RIV | 0.53 | 1.78 | 0.17 | 1.65 | -0.58 | 0.56 | 0.00 | 1283.40 |
| IIV3-SD: IIV4-SD | 0.22 | 3.80 | 0.02 | 3.04 | -0.86 | 0.39 | 0.00 | 6280021.74 |
| IIV3-SD: Placebo | 3.21 | 1.51 | 1.43 | 7.21 | 2.89 | 0.00 | 0.01 | 1123.64 |
| IIV3-SD: RIV | 0.86 | 1.58 | 0.35 | 2.10 | -0.23 | 0.82 | 0.00 | 502.06 |
| IIV4-SD: Placebo | 0.07 | 4.04 | 0.00 | 1.07 | -1.69 | 0.09 | 0.00 | 4275629.92 |
| IIV4-SD: RIV | 0.26 | 4.02 | 0.02 | 3.96 | -0.74 | 0.46 | 0.00 | 15005749.72 |
| RIV:Placebo | 3.74 | 1.85 | 1.12 | 12.52 | 2.12 | 0.03 | 0.00 | 14333.37 |
| Common within-network between-study SD (tau) | 0.21 |  |  |  |  |  |  |  |
| I-square | 3.7% |  |  |  |  |  |  |  |
| **Abbreviations-** IIV3: Trivalent **i**nactivated influenza vaccine; IIV4: Quadrivalent **i**nactivated influenza vaccine; Adj: Adjuvanted; SD: Standard dosage; HD: High dosage; RIV: Recombinant influenza vaccine; OR: Odds ratio; CI: Confidence interval; PI: Prediction interval; se: Standard error | | | | | | | | |

Note: Note: This analysis incorporates DiazGranados' 2014 study categorized as mismatched ^20^.

**Figure 4.** Treatment ranking of interventions using P-score for laboratory-confirmed influenza with original coding of interventions, restricted to matched studies

| ***Treatment Ranking*** | |
| --- | --- |
| ***Treatment*** | ***P-score*** |
| IIV4-SD | 0.83 |
| IIV3-HD | 0.76 |
| IIV3-Adj | 0.53 |
| RIV | 0.47 |
| IIV3-SD | 0.38 |
| Placebo | 0.02 |

**Abbreviations-** IIV3: Trivalent **i**nactivated influenza vaccine; IIV4: Quadrivalent **i**nactivated influenza vaccine; Adj: Adjuvanted; SD: Standard dosage; HD: High dosage;

RIV: Recombinant influenza vaccine

Note: This analysis incorporates DiazGranados' 2014 study categorized as mismatched ^20^.

**Influenza-like Illness**

**Note:** NMA of original coding of interventions was not conducted because the number of studies was smaller than the number of nodes.

**Table 5.** Sensitivity Analysis: Summary of network meta-analysis results for influenza-like illness with combined coding of interventions.

| ***Network Meta-analysis results; 9 studies, 65,658 participants, 6 treatments*** | | | | | | | | |
| --- | --- | --- | --- | --- | --- | --- | --- | --- |
| ***Comparison*** | ***OR*** | ***seOR*** | ***Lower CI*** | ***Upper CI*** | ***Statistic*** | ***p-value*** | ***Lower PI*** | ***Upper PI*** |
| Adj: HD | 1.05 | 1.09 | 0.89 | 1.25 | 0.60 | 0.55 | 0.80 | 1.39 |
| Adj: Placebo | 0.40 | 1.60 | 0.16 | 0.99 | -1.97 | 0.05 | 0.09 | 1.77 |
| Adj: RIV | 1.04 | 1.10 | 0.86 | 1.26 | 0.44 | 0.66 | 0.76 | 1.42 |
| Adj: SD | 1.03 | 1.09 | 0.87 | 1.21 | 0.34 | 0.73 | 0.79 | 1.34 |
| Adj: Tdap | 0.98 | 1.06 | 0.87 | 1.10 | -0.36 | 0.72 | 0.82 | 1.17 |
| HD: Placebo | 0.38 | 1.59 | 0.15 | 0.93 | -2.12 | 0.03 | 0.09 | 1.63 |
| HD: RIV | 0.99 | 1.06 | 0.89 | 1.11 | -0.17 | 0.87 | 0.83 | 1.19 |
| HD: SD | 0.98 | 1.02 | 0.93 | 1.02 | -0.99 | 0.32 | 0.90 | 1.05 |
| HD: Tdap | 0.93 | 1.11 | 0.76 | 1.14 | -0.70 | 0.49 | 0.67 | 1.30 |
| Placebo: RIV | 0.38 | 1.59 | 0.15 | 0.94 | -2.09 | 0.04 | 0.60 | 11.59 |
| Placebo: SD | 0.38 | 1.59 | 0.16 | 0.95 | -2.07 | 0.04 | 0.60 | 11.25 |
| Placebo: Tdap | 0.40 | 1.60 | 0.16 | 1.02 | -1.92 | 0.06 | 0.55 | 11.13 |
| RIV: SD | 0.99 | 1.05 | 0.89 | 1.09 | -0.27 | 0.78 | 0.84 | 1.16 |
| RIV: Tdap | 0.94 | 1.12 | 0.75 | 1.17 | -0.56 | 0.58 | 0.66 | 1.35 |
| SD: Tdap | 0.95 | 1.11 | 0.78 | 1.16 | -0.49 | 0.63 | 0.69 | 1.31 |
| Common within-network between-study SD (tau) | <0.0001 |  |  |  |  |  |  |  |
| I-square | 0.00% |  |  |  |  |  |  |  |
| **Abbreviations-** Adj: Adjuvanted; SD: Standard dosage; HD: High dosage; RIV: Recombinant influenza vaccine; Tdap: Tetanus, diphtheria, pertussis; OR: Odds ratio; CI: Confidence interval; PI: Prediction interval; se: Standard error | | | | | | | | |

**Figure 5.** Treatment ranking of interventions using P-score for influenza-like illness with combined coding of interventions.

| ***Treatment Ranking*** | |
| --- | --- |
| ***Treatment*** | ***P-score*** |
| Adj | 0.52 |
| HD | 0.77 |
| Placebo | 0.02 |
| RIV | 0.68 |
| SD | 0.57 |
| Tdap | 0.44 |

**Abbreviations-** Adj: Adjuvanted; SD: Standard dosage; HD: High dosage; RIV: Recombinant influenza vaccine; Tdap: Tetanus, diphtheria, pertussis

**Number of vascular adverse events**

**Table 6.** Summary of network meta-analysis results for number of vascular adverse events with original coding of interventions, restricted to studies in which the overall risk of bias was low.

| ***Network Meta-analysis results; 4 studies, 37,043 patients, 5 treatments*** | | | | | | | | |
| --- | --- | --- | --- | --- | --- | --- | --- | --- |
| ***Comparison*** | ***IRR*** | ***seOR*** | ***Lower CI*** | ***Upper CI*** | ***Statistic*** | ***p-value*** | ***Lower PI*** | ***Upper PI*** |
| IIV3-Adj: IIV3-HD | 1.16 | 4.15 | 0.07 | 18.94 | 0.11 | 0.91 | . | . |
| IIV3-Adj: IIV3-SD | 1.01 | 4.12 | 0.06 | 16.27 | 0.01 | 0.99 | . | . |
| IIV3-Adj: IIV4-Adj | 5.01 | 4.71 | 0.24 | 104.53 | 1.04 | 0.30 | . | . |
| IIV3-Adj: IIV4-HD | 1.75 | 4.58 | 0.09 | 34.60 | 0.37 | 0.71 | . | . |
| IIV3-HD: IIV3-SD | 0.87 | 1.15 | 0.67 | 1.14 | -1.02 | 0.31 | . | . |
| IIV3-HD: IIV4-Adj | 4.30 | 8.20 | 0.07 | 265.99 | 0.69 | 0.49 | . | . |
| IIV3-HD: IIV4-HD | 1.50 | 1.72 | 0.52 | 4.34 | 0.75 | 0.45 | . | . |
| IIV3-SD: IIV4-Adj | 0.20 | 8.16 | 0.00 | 12.39 | -0.76 | 0.45 | . | . |
| IIV3-SD: IIV4-HD | 0.58 | 1.75 | 0.19 | 1.73 | -0.98 | 0.33 | . | . |
| IIV4-Adj: IIV4-HD | 0.35 | 8.78 | 0.00 | 24.69 | -0.48 | 0.63 | . | . |
| Common within-network between-study SD | 0 |  |  |  |  |  |  |  |
| I-square | 0.00% |  |  |  |  |  |  |  |
| **Abbreviations-** IIV3: Trivalent **i**nactivated influenza vaccine; IIV4: Quadrivalent **i**nactivated influenza vaccine; Adj: Adjuvanted; SD: Standard dosage; HD: High dosage; IRR: Incidence rate ratio; CI: Confidence interval; PI: Prediction interval; se: Standard error | | | | | | | | |

**Figure 6.** Treatment ranking of interventions using P-score for number of vascular adverse events with original coding of interventions, restricted to studies in which the overall risk of bias was low.

| ***Treatment Ranking*** | |
| --- | --- |
| ***Treatment*** | ***P-score*** |
| IIV4-Adj | 0.77 |
| IIV4-HD | 0.64 |
| IIV3-HD | 0.46 |
| IIV3-Adj | 0.36 |
| IIV3-SD | 0.26 |

**Abbreviations-** IIV3: Trivalent **i**nactivated influenza vaccine; IIV4: Quadrivalent **i**nactivated influenza vaccine; Adj: Adjuvanted; SD: Standard dosage; HD: High dosage

**All-cause mortality**

**Table 7.** Summary of network meta-analysis results for all-cause mortality with original coding of interventions, restricted to studies in which the overall risk of bias was low.

| ***Network Meta-analysis results; 9 studies, 52,902 patients, 7 treatments*** | | | | | | | | | |
| --- | --- | --- | --- | --- | --- | --- | --- | --- | --- |
| ***Comparison*** | ***OR*** | ***seOR*** | ***Lower CI*** | ***Upper CI*** | ***Statistic*** | | ***p-value*** | ***Lower PI*** | ***Upper PI*** |
| IIV3-Adj:IIV3-HD | 0.63 | 2.92 | 0.08 | 5.17 | -0.43 | | 0.67 | 0.01 | 63.79 |
| IIV3-Adj:IIV3-SD | 0.62 | 2.93 | 0.08 | 5.10 | -0.44 | | 0.66 | 0.01 | 63.17 |
| IIV3-Adj:IIV4-Adj | 0.20 | 4.71 | 0.01 | 4.16 | -1.04 | | 0.30 | 0.00 | 157.13 |
| IIV3-Adj:IIV4-HD | 0.84 | 4.09 | 0.05 | 13.34 | -0.12 | | 0.90 | 0.00 | 361.86 |
| IIV3-Adj:IIV4-SD | 0.64 | 2.93 | 0.08 | 5.24 | -0.42 | | 0.67 | 0.01 | 65.23 |
| IIV3-Adj:Tdap | 0.19 | 4.80 | 0.01 | 4.20 | -1.05 | | 0.30 | 0.00 | 165.84 |
| IIV3-HD:IIV3-SD | 0.98 | 1.16 | 0.73 | 1.32 | -0.11 | | 0.91 | 0.52 | 1.87 |
| IIV3-HD:IIV4-Adj | 0.32 | 6.58 | 0.01 | 12.69 | -0.61 | | 0.54 | 0.00 | 1049.28 |
| IIV3-HD:IIV4-HD | 1.33 | 2.49 | 0.22 | 7.99 | 0.31 | | 0.75 | 0.03 | 67.99 |
| IIV3-HD:IIV4-SD | 1.00 | 1.10 | 0.83 | 1.22 | 0.05 | | 0.96 | 0.66 | 1.53 |
| IIV3-HD:Tdap | 0.31 | 6.69 | 0.01 | 12.71 | -0.62 | | 0.53 | 0.00 | 1091.33 |
| IIV3-SD:IIV4-Adj | 3.12 | 6.59 | 0.08 | 125.57 | 0.60 | | 0.55 | 0.00 | 10406.29 |
| IIV3-SD:IIV4-HD | 0.74 | 2.52 | 0.12 | 4.53 | -0.33 | | 0.74 | 0.01 | 39.65 |
| IIV3-SD:IIV4-SD | 0.98 | 1.19 | 0.70 | 1.38 | -0.12 | | 0.91 | 0.46 | 2.08 |
| IIV3-SD:Tdap | 3.21 | 6.70 | 0.08 | 133.40 | 0.61 | | 0.54 | 0.00 | 11475.22 |
| IIV4-Adj:IIV4-HD | 4.23 | 8.12 | 0.07 | 256.30 | 0.69 | | 0.49 | 0.00 | 34656.14 |
| IIV4-Adj:IIV4-SD | 3.18 | 6.60 | 0.08 | 128.57 | 0.61 | | 0.54 | 0.00 | 10692.71 |
| IIV4-Adj:Tdap | 0.97 | 1.28 | 0.60 | 1.57 | -0.12 | | 0.91 | 0.34 | 2.79 |
| IIV4-HD:IIV4-SD | 0.75 | 2.51 | 0.12 | 4.56 | -0.31 | | 0.76 | 0.01 | 39.29 |
| IIV4-HD:Tdap | 0.23 | 8.24 | 0.00 | 14.34 | -0.70 | | 0.49 | 0.00 | 2005.28 |
| IIV4-SD:Tdap | 0.31 | 6.71 | 0.01 | 12.71 | -0.62 | | 0.53 | 0.00 | 1097.64 |
| Common within-network between-study SD | | | | <0.0001 |  |  |  |  |  |
| I-square | | | | 0.00% |  |  |  |  |  |
| **Abbreviations-** IIV3: Trivalent **i**nactivated influenza vaccine; IIV4: Quadrivalent **i**nactivated influenza vaccine; Adj: Adjuvanted; SD: Standard dosage; HD: High dosage; Tdap: Tetanus, diphtheria, pertussis; OR: Odds ratio; CI: Confidence interval; PI: Prediction interval ; se: Standard error | | | | | | | | | |

**Figure 7.** Treatment ranking of interventions using P-score for outpatient visits with original coding of interventions, restricted to studies in which the overall risk of bias was low.

| ***Treatment Ranking*** | |
| --- | --- |
| ***Treatment*** | ***P-score*** |
| IIV3-Adj | 0.71 |
| IIV4-HD | 0.64 |
| IIV4-SD | 0.54 |
| IIV3-HD | 0.53 |
| IIV3-SD | 0.51 |
| IIV4-Adj | 0.29 |
| Tdap | 0.27 |

**Abbreviations-** IIV3: Trivalent **i**nactivated influenza vaccine; IIV4: Quadrivalent **i**nactivated influenza vaccine; Adj: Adjuvanted; SD: Standard dosage; HD: High dosage;

Tdap: Tetanus, diphtheria, pertussis

**Table 8.** Sensitivity Analysis**:** Summary of network meta-analysis results for all-cause mortality with original coding of interventions, restricted to studies in which the proportion of females was at least 50%.

| ***Network Meta-analysis results; 16 studies, 102,519 patients, 6 treatments*** | | | | | | | | | | | | |
| --- | --- | --- | --- | --- | --- | --- | --- | --- | --- | --- | --- | --- |
| ***Comparison*** | ***OR*** | ***seOR*** | ***Lower CI*** | ***Upper CI*** | ***Statistic*** | | ***p-value*** | | ***Lower PI*** | | | ***Upper PI*** |
| IIV3-Adj:IIV3-HD | 1.12 | 1.23 | 0.75 | 1.68 | 0.58 | | 0.56 | | 0.70 | | | 1.80 |
| IIV3-Adj:IIV3-SD | 1.09 | 1.22 | 0.73 | 1.61 | 0.42 | | 0.67 | | 0.68 | | | 1.73 |
| IIV3-Adj:IIV4-HD | 1.50 | 2.55 | 0.24 | 9.40 | 0.43 | | 0.67 | | 0.17 | | | 12.99 |
| IIV3-Adj:IIV4-SD | 1.12 | 1.25 | 0.72 | 1.74 | 0.50 | | 0.62 | | 0.66 | | | 1.88 |
| IIV3-Adj:Placebo | 1.60 | 1.94 | 0.44 | 5.85 | 0.71 | | 0.48 | | 0.35 | | | 7.36 |
| IIV3-Adj:RIV | 1.56 | 1.60 | 0.62 | 3.91 | 0.94 | | 0.35 | | 0.53 | | | 4.60 |
| IIV3-HD:IIV3-SD | 0.97 | 1.03 | 0.91 | 1.03 | -0.99 | | 0.32 | | 0.90 | | | 1.04 |
| IIV3-HD:IIV4-HD | 1.33 | 2.49 | 0.22 | 7.99 | 0.31 | | 0.75 | | 0.16 | | | 10.97 |
| IIV3-HD:IIV4-SD | 0.99 | 1.10 | 0.82 | 1.21 | -0.06 | | 0.95 | | 0.79 | | | 1.25 |
| IIV3-HD:Placebo | 1.42 | 1.88 | 0.41 | 4.90 | 0.55 | | 0.58 | | 0.33 | | | 6.10 |
| IIV3-HD:RIV | 1.38 | 1.53 | 0.60 | 3.18 | 0.77 | | 0.44 | | 0.52 | | | 3.68 |
| IIV3-SD:IIV4-HD | 1.38 | 2.50 | 0.23 | 8.27 | 0.35 | | 0.73 | | 0.17 | | | 11.35 |
| IIV3-SD:IIV4-SD | 1.03 | 1.11 | 0.84 | 1.26 | 0.26 | | 0.79 | | 0.81 | | | 1.31 |
| IIV3-SD:Placebo | 1.47 | 1.88 | 0.43 | 5.06 | 0.61 | | 0.54 | | 0.34 | | | 6.29 |
| IIV3-SD:RIV | 1.43 | 1.53 | 0.62 | 3.29 | 0.84 | | 0.40 | | 0.54 | | | 3.81 |
| IIV4-HD:IIV4-SD | 0.75 | 2.51 | 0.12 | 4.52 | -0.32 | | 0.75 | | 0.09 | | | 6.21 |
| IIV4-HD:Placebo | 1.06 | 3.04 | 0.12 | 9.40 | 0.06 | | 0.96 | | 0.08 | | | 13.81 |
| IIV4-HD:RIV | 1.04 | 2.74 | 0.14 | 7.48 | 0.04 | | 0.97 | | 0.10 | | | 10.60 |
| IIV4-SD:Placebo | 1.43 | 1.90 | 0.41 | 5.00 | 0.56 | | 0.58 | | 0.33 | | | 6.24 |
| IIV4-SD vs. RIV | 1.39 | 1.52 | 0.62 | 3.15 | 0.80 | | 0.43 | | 0.53 | | | 3.64 |
| RIV vs. Placebo | 1.03 | 2.14 | 0.23 | 4.55 | 0.03 | | 0.97 | | 0.18 | | | 5.93 |
| Common within-network between-study SD | | <0.0001 |  | | |  | |  | |  |  | |
| I-square | | 0.00% |  | | |  | |  | |  |  | |
| **Abbreviations-** IIV3: Trivalent **i**nactivated influenza vaccine; IIV4: Quadrivalent **i**nactivated influenza vaccine; Adj: Adjuvanted; SD: Standard dosage; HD: High dosage; RIV: Recombinant influenza vaccine; OR: Odds ratio; CI: Confidence interval; PI: Prediction interval; se: Standard error | | | | | | | | | | | | |

**Figure 8.** Treatment ranking of interventions using P-score for all-cause mortality with original coding of interventions, restricted to studies in which the proportion of females was at least 50%.

| ***Treatment Ranking*** | |
| --- | --- |
| ***Treatment*** | ***P-score*** |
| IIV3-HD | 0.82 |
| RIV | 0.66 |
| IIV3-Adj | 0.61 |
| IIV3-SD | 0.50 |
| IIV4-SD | 0.29 |
| Placebo | 0.11 |

**Abbreviations-** IIV3: Trivalent **i**nactivated influenza vaccine; IIV4: Quadrivalent **i**nactivated influenza vaccine; Adj: Adjuvanted; SD: Standard dosage; HD: High dosage; RIV: Recombinant influenza vaccine

**Laboratory-confirmed influenza**

**Figure 9.** Sensitivity Analysis: Forest Plots of network estimates relative to placebo for laboratory-confirmed influenza with original coding of interventions, restricted to studies in which the proportion of females was at least 50%.

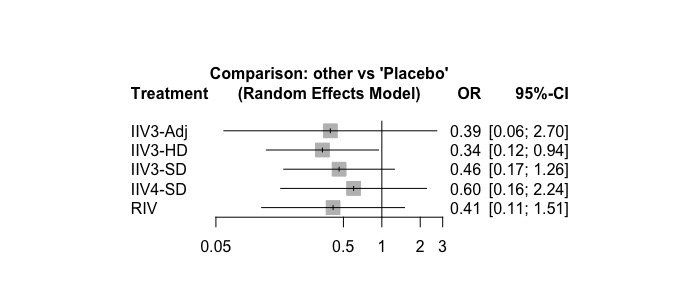

*OR>1 favours placebo; OR<1 favours intervention

**Abbreviations-** IIV3: Trivalent **i**nactivated influenza vaccine; IIV4: Quadrivalent **i**nactivated influenza vaccine; Adj: Adjuvanted; SD: Standard dosage; HD: High dosage; RIV: Recombinant influenza vaccine; OR: Odds ratio; CI: Confidence interval

**Figure 10.** Sensitivity Analysis: Forest Plots of network estimates relative to placebo for laboratory-confirmed influenza with combined coding of interventions.

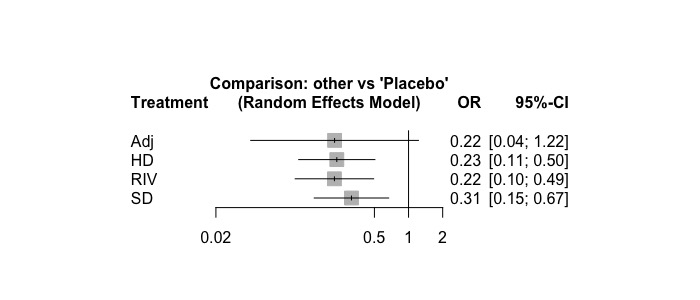

*OR>1 favours placebo; OR<1 favours intervention

**Abbreviations-** Adj: Adjuvanted; SD: Standard dosage; HD: High dosage; RIV: Recombinant influenza vaccine; OR: Odds ratio; CI: Confidence interval

**Influenza-like illness**

**Note:** NMA of original coding of interventions was not conducted because the number of studies was smaller than the number of nodes.

**Figure 11.** Sensitivity Analysis: Forest Plots of network estimates relative to placebo for influenza-like illness with combined coding of interventions.

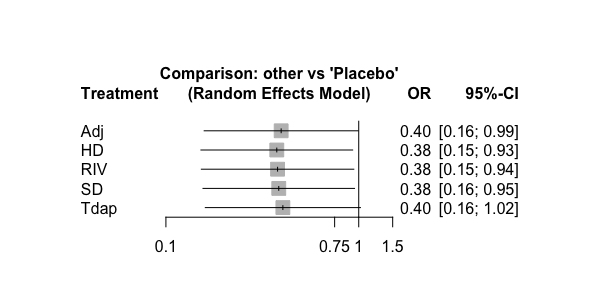

*OR>1 favours placebo; OR<1 favours intervention

**Abbreviations-** Adj: Adjuvanted; SD: Standard dosage; HD: High dosage; RIV: Recombinant influenza vaccine; Tdap: Tetanus, diphtheria, pertussis; OR: Odds ratio; CI: Confidence interval

**Number of vascular adverse events**

**Figure 12.** Forest Plot of network estimates relative to IIV3-SD for number of vascular adverse events with original coding of interventions, restricted to studies in which the overall risk of bias was low.

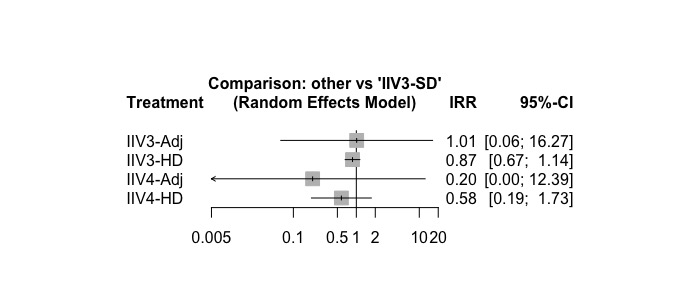

*IRR>1 favours IIV3-SD; IRR<1 favours intervention

**Abbreviations-** IIV3: Trivalent **i**nactivated influenza vaccine; IIV4: Quadrivalent **i**nactivated influenza vaccine; Adj: Adjuvanted; HD: High dosage; IRR: incidence rate ratio; CI: Confidence interval

**All-cause mortality**

**Figure 13.** Sensitivity Analysis: Forest Plots of network estimates relative to IIV3-SD for all-cause mortality with original coding of interventions, restricted to studies in which the overall risk of bias was low.

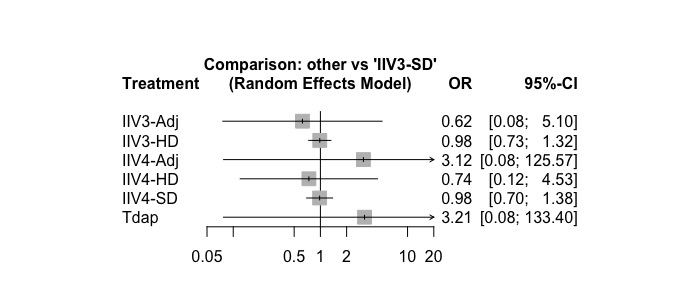

*OR>1 favours placebo; OR<1 favours intervention

**Abbreviations-** IIV3: Trivalent **i**nactivated influenza vaccine; IIV4: Quadrivalent **i**nactivated influenza vaccine; Adj: Adjuvanted; SD: Standard dosage; HD: High dosage; Tdap: Tetanus, diphtheria, pertussis; OR: Odds ratio; CI: Confidence interval

**Figure 14.** Sensitivity Analysis: Forest Plots of network estimates relative to placebo for all-cause mortality with original coding of interventions, restricted to studies in which the proportion of females was at least 50%.

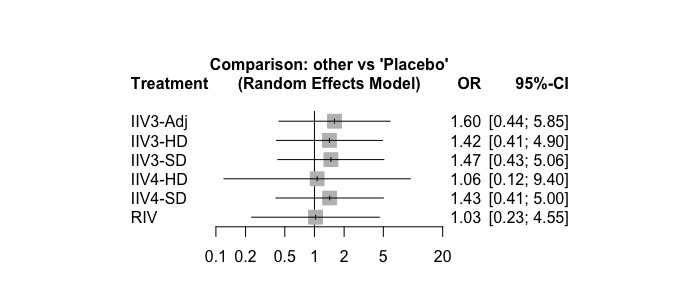

*OR>1 favours placebo; OR<1 favours intervention

**Abbreviations-** IIV3: Trivalent **i**nactivated influenza vaccine; IIV4: Quadrivalent **i**nactivated influenza vaccine; Adj: Adjuvanted; SD: Standard dosage; HD: High dosage; RIV: Recombinant influenza vaccine; Tdap: Tetanus, diphtheria, pertussis; OR: Odds ratio; CI: Confidence interva

#### **Appendix 28B: Confidence in Network Meta-Analysis (CINeMA) Assessments Under NACI-Recommended Minimally Important Differences.**

**Laboratory-confirmed influenza**

**Table 1.** CINeMA assessment providing credibility for each treatment comparison for laboratory-confirmed influenza under NACI-recommended minimally important differences.

| **Treatment Comparison** | **Number of studies** | **Within-study bias** | **Reporting bias** | **Indirectness** | **Imprecision** | **Heterogeneity** | **Incoherence** | **Confidence rating** | **Reason(s) for downgrading** |
| --- | --- | --- | --- | --- | --- | --- | --- | --- | --- |
| **Direct Evidence** | | | | | | | | | |
| IIV3-Adj:IIV3-HD | 1 | Some concerns | Some concerns | No concerns | Major concerns | No concerns | No concerns | Low | ["Within-study bias","Imprecision"] |
| IIV3-Adj:RIV | 1 | Major concerns | Some concerns | No concerns | Major concerns | No concerns | No concerns | Low | ["Within-study bias","Imprecision"] |
| IIV3-HD:IIV3-SD | 3 | No concerns | Some concerns | No concerns | No concerns | No concerns | No concerns | High | Not applicable |
| IIV3-HD:IIV4-SD | 1 | Some concerns | Some concerns | No concerns | Some concerns | Some concerns | No concerns | Very low | ["Within-study bias","Imprecision","Heterogeneity"] |
| IIV3-HD:RIV | 1 | Some concerns | Some concerns | No concerns | Major concerns | No concerns | No concerns | Low | ["Within-study bias","Imprecision"] |
| IIV3-SD:Placebo | 2 | Major concerns | Some concerns | Some concerns | No concerns | No concerns | No concerns | Low | ["Within-study bias","Indirectness"] |
| IIV3-SD:RIV | 1 | Some concerns | Some concerns | No concerns | Major concerns | No concerns | No concerns | Low | ["Within-study bias","Imprecision"] |
| IIV4-SD:RIV | 1 | Major concerns | Some concerns | No concerns | No concerns | Some concerns | No concerns | Low | ["Within-study bias","Heterogeneity"] |
| **Indirect Evidence** | | | | | | | | | |
| IIV3-Adj:IIV3-SD | 0 | Some concerns | Some concerns | No concerns | Major concerns | No concerns | No concerns | Low | ["Within-study bias","Imprecision"] |
| IIV3-Adj:IIV4-SD | 0 | Major concerns | Some concerns | No concerns | Major concerns | No concerns | No concerns | Low | ["Within-study bias","Imprecision"] |
| IIV3-Adj:Placebo | 0 | Some concerns | Some concerns | No concerns | Some concerns | Some concerns | No concerns | Very low | ["Within-study bias","Imprecision","Heterogeneity"] |
| IIV3-HD:Placebo | 0 | Some concerns | Some concerns | Some concerns | No concerns | No concerns | No concerns | Low | ["Within-study bias","Indirectness"] |
| IIV3-SD:IIV4-SD | 0 | Some concerns | Some concerns | No concerns | Major concerns | No concerns | No concerns | Low | ["Within-study bias","Imprecision"] |
| IIV4-SD:Placebo | 0 | Some concerns | Some concerns | No concerns | Some concerns | Some concerns | No concerns | Very low | ["Within-study bias","Imprecision","Heterogeneity"] |
| Placebo:RIV | 0 | Some concerns | Some concerns | No concerns | No concerns | Some concerns | No concerns | Low | ["Within-study bias","Heterogeneity"] |
| **Abbreviations-** IIV3: Trivalent **i**nactivated influenza vaccine; IIV4: Quadrivalent **i**nactivated influenza vaccine; Adj: Adjuvanted; SD: Standard dosage; HD: High dosage; RIV: Recombinant influenza vaccine | | | | | | | | | |

**Number of vascular adverse events**

**Table 2.** CINeMA assessment providing credibility for each treatment comparison for number of vascular adverse events under NACI-recommended minimally important differences.

| **Treatment Comparison** | **Number of studies** | **Within-study bias** | **Reporting bias** | **Indirectness** | **Imprecision** | **Heterogeneity** | **Incoherence** | **Confidence rating** | **Reason(s) for downgrading** |
| --- | --- | --- | --- | --- | --- | --- | --- | --- | --- |
| **Direct Evidence** | | | | | | | | | |
| IIV3-Adj:IIV3-SD | 2 | Major concerns | Some concerns | No concerns | No concerns | Major concerns | Major concerns | Very low | ["Within-study bias","Heterogeneity","Incoherence"] |
| IIV3-Adj:IIV4-Adj | 1 | No concerns | Some concerns | No concerns | No concerns | No concerns | Major concerns | Moderate | ["Incoherence"] |
| IIV3-HD:IIV3-SD | 4 | No concerns | Some concerns | No concerns | No concerns | Major concerns | Major concerns | Low | ["Heterogeneity","Incoherence"] |
| IIV3-HD:IIV4-HD | 1 | No concerns | Some concerns | No concerns | No concerns | Major concerns | Major concerns | Low | ["Heterogeneity","Incoherence"] |
| **Indirect Evidence** | | | | | | | | | |
| IIV3-Adj:IIV3-HD | 0 | Some concerns | Some concerns | No concerns | No concerns | Major concerns | Major concerns | Very low | ["Within-study bias","Heterogeneity","Incoherence"] |
| IIV3-Adj:IIV4-HD | 0 | Some concerns | Some concerns | No concerns | No concerns | Major concerns | Major concerns | Very low | ["Within-study bias","Heterogeneity","Incoherence"] |
| IIV3-HD:IIV4-Adj | 0 | Some concerns | Some concerns | No concerns | No concerns | Major concerns | Major concerns | Very low | ["Within-study bias","Heterogeneity","Incoherence"] |
| IIV3-SD:IIV4-Adj | 0 | Some concerns | Some concerns | No concerns | No concerns | Some concerns | Major concerns | Very low | ["Within-study bias","Heterogeneity","Incoherence"] |
| IIV3-SD:IIV4-HD | 0 | No concerns | Some concerns | No concerns | No concerns | Major concerns | Major concerns | Low | ["Heterogeneity","Incoherence"] |
| IIV4-Adj:IIV4-HD | 0 | Some concerns | Some concerns | No concerns | No concerns | Major concerns | Major concerns | Very low | ["Within-study bias","Heterogeneity","Incoherence"] |
| **Abbreviations-** IIV3: Trivalent **i**nactivated influenza vaccine; IIV4: Quadrivalent **i**nactivated influenza vaccine; Adj: Adjuvanted; SD: Standard dosage; HD: High dosage | | | | | | | | | |

**Outpatient visits**

**Table 3.** CINeMA assessment providing credibility for each treatment comparison for outpatient visits under recommended minimally important differences.

| **Treatment Comparison** | **Number of studies** | **Within-study bias** | **Reporting bias** | **Indirectness** | **Imprecision** | **Heterogeneity** | **Incoherence** | **Confidence rating** | **Reason(s) for downgrading** |
| --- | --- | --- | --- | --- | --- | --- | --- | --- | --- |
| **Direct Evidence** | | | | | | | | | |
| IIV3-HD:IIV3-SD | 2 | Some concerns | Some concerns | No concerns | No concerns | Major concerns | Major concerns | Very low | ["Within-study bias","Heterogeneity","Incoherence"] |
| IIV3-SD:Placebo | 2 | Some concerns | Some concerns | No concerns | Some concerns | Some concerns | Major concerns | Very low | ["Within-study bias","Imprecision","Heterogeneity","Incoherence"] |
| **Indirect Evidence** | | | | | | | | | |
| IIV3-HD:Placebo | 0 | Some concerns | Some concerns | No concerns | Some concerns | Some concerns | Major concerns | Very low | ["Within-study bias","Imprecision","Heterogeneity","Incoherence"] |
| **Abbreviations-** IIV3: Trivalent **i**nactivated influenza vaccine; IIV4: Quadrivalent **i**nactivated influenza vaccine; SD: Standard dosage; HD: High dosage | | | | | | | | | |

**All-cause mortality**

**Table 4.** CINeMA assessment providing credibility for each treatment comparison for all-cause mortality under recommended minimally important differences.

| **Treatment Comparison** | **Number of studies** | **Within-study bias** | **Reporting bias** | **Indirectness** | **Imprecision** | **Heterogeneity** | **Incoherence** | **Confidence rating** | **Reason(s) for downgrading** |
| --- | --- | --- | --- | --- | --- | --- | --- | --- | --- |
| **Direct Evidence** | | | | | | | | | |
| IIV3-Adj:IIV3-HD | 1 | Some concerns | No  concerns | No concerns | No concerns | No concerns | No concerns | Moderate | ["Within-study bias"] |
| IIV3-Adj:IIV3-SD | 2 | Some concerns | No  concerns | No concerns | No concerns | No concerns | No concerns | Moderate | ["Within-study bias"] |
| IIV3-Adj:IIV4-Adj | 1 | No concerns | No  concerns | No concerns | Major concerns | No concerns | No concerns | Moderate | ["Imprecision"] |
| IIV3-HD:IIV3-SD | 6 | Some concerns | No  concerns | No concerns | No concerns | No concerns | No concerns | Moderate | ["Within-study bias"] |
| IIV3-HD:IIV4-HD | 1 | No concerns | No  concerns | No concerns | Major concerns | No concerns | No concerns | Moderate | ["Imprecision"] |
| IIV3-HD:IIV4-SD | 1 | No concerns | No  concerns | Some concerns | No concerns | No concerns | No concerns | Moderate | ["Indirectness"] |
| IIV3-SD:IIV4-SD | 3 | No concerns | No  concerns | No concerns | No concerns | No concerns | No concerns | High | Not applicable |
| IIV3-SD:Placebo | 2 | Some concerns | No  concerns | Some concerns | Some concerns | No concerns | No concerns | Very low | ["Within-study bias","Indirectness","Imprecision"] |
| IIV3-SD:RIV | 1 | Some concerns | No  concerns | No concerns | Some concerns | No concerns | No concerns | Low | ["Within-study bias","Imprecision"] |
| IIV4-Adj:Tdap | 1 | No concerns | No  concerns | No concerns | No concerns | No concerns | No concerns | High | Not applicable |
| IIV4-SD:RIV | 1 | Some concerns | No  concerns | No concerns | Some concerns | No concerns | No concerns | Low | ["Within-study bias","Imprecision"] |
| **Indirect Evidence** | | | | | | | | | |
| IIV3-Adj:IIV4-HD | 0 | Some concerns | No  concerns | No concerns | Major concerns | No concerns | No concerns | Low | ["Within-study bias","Imprecision"] |
| IIV3-Adj:IIV4-SD | 0 | Some concerns | No  concerns | No concerns | No concerns | No concerns | No concerns | Moderate | ["Within-study bias"] |
| IIV3-Adj:Placebo | 0 | Some concerns | No  concerns | No concerns | Some concerns | No concerns | No concerns | Low | ["Within-study bias","Imprecision"] |
| IIV3-Adj:RIV | 0 | Some concerns | No  concerns | No concerns | Some concerns | No concerns | No concerns | Low | ["Within-study bias","Imprecision"] |
| IIV3-Adj:Tdap | 0 | No concerns | No  concerns | No concerns | Major concerns | No concerns | No concerns | Moderate | ["Imprecision"] |
| IIV3-HD:IIV4-Adj | 0 | Some concerns | No  concerns | No concerns | Major concerns | No concerns | No concerns | Low | ["Within-study bias","Imprecision"] |
| IIV3-HD:Placebo | 0 | Some concerns | No  concerns | No concerns | Some concerns | No concerns | No concerns | Low | ["Within-study bias","Imprecision"] |
| IIV3-HD:RIV | 0 | Some concerns | No  concerns | No concerns | Some concerns | No concerns | No concerns | Low | ["Within-study bias","Imprecision"] |
| IIV3-HD:Tdap | 0 | No concerns | No  concerns | No concerns | Major concerns | No concerns | No concerns | Moderate | ["Imprecision"] |
| IIV3-SD:IIV4-Adj | 0 | No concerns | No  concerns | No concerns | Major concerns | No concerns | No concerns | Moderate | ["Imprecision"] |
| IIV3-SD:IIV4-HD | 0 | No concerns | No  concerns | No concerns | Major concerns | No concerns | No concerns | Moderate | ["Imprecision"] |
| IIV3-SD:Tdap | 0 | No concerns | No  concerns | No concerns | Major concerns | No concerns | No concerns | Moderate | ["Imprecision"] |
| IIV4-Adj:IIV4-HD | 0 | No concerns | No  concerns | No concerns | Major concerns | No concerns | No concerns | Moderate | ["Imprecision"] |
| IIV4-Adj:IIV4-SD | 0 | No concerns | No  concerns | No concerns | Major concerns | No concerns | No concerns | Moderate | ["Imprecision"] |
| IIV4-Adj:Placebo | 0 | Some concerns | No  concerns | No concerns | Major concerns | No concerns | No concerns | Low | ["Within-study bias","Imprecision"] |
| IIV4-Adj:RIV | 0 | Some concerns | No  concerns | No concerns | Major concerns | No concerns | No concerns | Low | ["Within-study bias","Imprecision"] |
| IIV4-HD:IIV4-SD | 0 | No concerns | No  concerns | No concerns | Major concerns | No concerns | No concerns | Moderate | ["Imprecision"] |
| IIV4-HD:Placebo | 0 | Some concerns | No  concerns | No concerns | Major concerns | No concerns | No concerns | Low | ["Within-study bias","Imprecision"] |
| IIV4-HD:RIV | 0 | No concerns | No  concerns | No concerns | Major concerns | No concerns | No concerns | Moderate | ["Imprecision"] |
| IIV4-HD:Tdap | 0 | No concerns | No  concerns | No concerns | Major concerns | No concerns | No concerns | Moderate | ["Imprecision"] |
| IIV4-SD:Placebo | 0 | Some concerns | No  concerns | Some concerns | Some concerns | No concerns | No concerns | Very low | ["Within-study bias","Indirectness","Imprecision"] |
| IIV4-SD:Tdap | 0 | No concerns | No  concerns | No concerns | Major concerns | No concerns | No concerns | Moderate | ["Imprecision"] |
| Placebo:RIV | 0 | Some concerns | No  concerns | No concerns | Major concerns | No concerns | No concerns | Low | ["Within-study bias","Imprecision"] |
| Placebo:Tdap | 0 | No concerns | No  concerns | No concerns | Major concerns | No concerns | No concerns | Moderate | ["Imprecision"] |
| RIV:Tdap | 0 | No concerns | No  concerns | No concerns | Major concerns | No concerns | No concerns | Moderate | ["Imprecision"] |
| **Abbreviations-** IIV3: Trivalent **i**nactivated influenza vaccine; IIV4: Quadrivalent **i**nactivated influenza vaccine; Adj: Adjuvanted; SD: Standard dosage; HD: High dosage; RIV: Recombinant influenza vaccine; Tdap: Tetanus, diphtheria, pertussis | | | | | | | | | |

#### **Appendix 28C: Grading of Recommendations, Assessment, Development, and Evaluations (GRADE) Assessments of Subgroup Analysis**

**Comparison 1: High Dose Trivalent Vaccine compared to Standard Dose Trivalent Vaccine for preventing influenza.**

| **Certainty assessment** | | | | | | | | | | | | | **Number of participants** | | | | **Effect** | | | **Certainty** | **Importance** |
| --- | --- | --- | --- | --- | --- | --- | --- | --- | --- | --- | --- | --- | --- | --- | --- | --- | --- | --- | --- | --- | --- |
| **Number of studies** | **Study design** | | **Risk of bias** | | **Inconsistency** | | **Indirectness** | | **Imprecision** | | **Other considerations** | | **High Dose Trivalent Vaccine** | | **Standard Dose Trivalent Vaccine** | | **Relative (95% CI)** | | **Absolute (95% CI)** |  |  |
| **Inpatient hospitalization (any cause)- SUBGROUP: Low RoB** | | | | | | | | | | | | | | | | | | | | | |
| RCTs | Not serious | | Not serious^i^ | | Not serious | | Serious^j^ | | None | | 1727/17456 (9.9%) | | 1944/17489 (11.1%) | | **OR 0.70** (0.52 to 1.13) | | **31 fewer per 1,000** (from 50 fewer to 13 more) | | ⨁⨁⨁◯ Moderate | RCTs |  |
| **Inpatient hospitalization (any cause)- SUBGROUP >80y** | | | | | | | | | | | | | | | | | | | | | |
| RCTs | | Serious^h^ | | Not serious | | Not serious | | Not serious | | None | | 1531/18578 (8.2%) | | 1644/17281 (9.5%) | | **OR 0.92** (0.86 to 0.99) | | **7 fewer per 1,000** (from 12 fewer to 1 fewer) | | ⨁⨁⨁◯ Moderate |  |

**Abbreviations –** RCTs: Randomized controlled trials; CI: Confidence interval; OR**:** Odds ratio

**Explanations**

a. Study that carried large weight for the overall effect estimated rated as some concerns due to bias arising from the randomization process and bias in selection of the reported result. Another study that carried small weight for the overall effect estimated rated as high risk of bias due to missing outcome data

b. Serious imprecision. 95% CI is consistent with the possibility for important benefit and large harm, including only 27 events in total.

c. Study that carried large weight for the overall effect estimated rated as high risk of bias due to some concerns in the timing of identification or recruitment of participants in a cluster trial, bias due to deviations from intended interventions and bias due to missing outcome data.

d. Study that carried large weight for the overall effect estimated was rated as low risk of bias, and the other study, that carried small weight for the overall effect estimated, was rated as high risk of bias due to deviations from intended interventions.

e. I^2^ value is 87%, suggesting some heterogeneity; We could not find the reasons for this high heterogeneity

f. Serious imprecision. 95% CI is consistent with the possibility for important benefit and large harm, including only 287 events in total

g. Study that carried large weight for the overall effect estimated rated as some concerns due to bias in selection of the reported result. No rating down due to RoB

h. Study that carried large weight for the overall effect estimated rated as high risk of bias due to missing outcome data

i. I^2^=87%, suggesting substantial heterogeneity, but of questionable clinical importance, because the two studies with more weight in the meta-analysis show a significant effect. We did not rate down due to inconsistency

j. Serious imprecision. 95% CI is consistent with the possibility for important benefit and large harm.

#### **Appendix 28D: Pairwise Subgroup Meta-Analyses**

**Table 1.** Pairwise meta-analysis results for each outcome in which a subgroup analysis was conducted

| **Primary Outcomes** | | | | | | | | | | | | | | | |
| --- | --- | --- | --- | --- | --- | --- | --- | --- | --- | --- | --- | --- | --- | --- | --- |
| ***Outcome*** | ***Comparison*** | ***Number of studies*** | ***Number of participants*** | ***OR*** | ***lower CI*** | ***upper CI*** | ***lower PI*** | ***upper PI*** | ***tau*** | ***tau^2^ lower CI*** | ***tau^2^ upper CI*** | ***Q*** | ***Q df*** | ***Q P-value*** | ***I^2^*** |
| LCI – Matched strains (DiazGranados 2014 defined as Match) | IIV3-HD : IIV3-SD | 2 | 330 | 0.74 | 0.63 | 0.88 | . | . | 0 | . | . | 0.86 | 1 | 0.35 | 0 |
| LCI – Mismatched strains (DiazGranados 2014 defined as Match) | IIV3-HD : IIV3-SD | 1 | 9158 | 0.87 | 0.56 | 1.36 | . | . | . | . | . | . | . | . | . |
| LCI – Matched strains (DiazGranados 2014 defined as Mismatch) | IIV3-HD : IIV3-SD | 1 | 612 | 0.52 | 0.35 | 0.77 | . | . | . | . | . | . | . | . | . |
| LCI – Mismatched strains (DiazGranados 2014 defined as Mismatch) | IIV3-HD : IIV3-SD | 2 | 31,141 | 0.76 | 0.64 | 0.90 | . | . | 0 | . | . | 0.11 | 1 | 0.75 | 0 |
| **Secondary Outcomes** | | | | | | | | | | | | | | | |
| ***Outcome*** | ***Comparison*** | ***Number of studies*** | ***Number of participants*** | ***OR*** | ***lower CI*** | ***upper CI*** | ***lower PI*** | ***upper PI*** | ***tau*** | ***tau^2^ lower CI*** | ***tau^2^ upper CI*** | ***Q*** | ***Q df*** | ***Q P-value*** | ***I^2^*** |
| Inpatient Hospitalization (any cause) – Low RoB subgroup | IIV3-HD : IIV3-SD | 2 | 34940 | 0.76 | 0.52 | 1.13 | . | . | 0.27 | . | . | 14.00 | 1 | 0.00 | 93% |
| Inpatient Hospitalization (any cause) – Over 80 y.o. subgroup | IIV3-HD : IIV3-SD | 2 | 35859 | 0.92 | 0.86 | 0.99 | . | . | 0.00 | . | . | 0.20 | 1 | 0.66 | 0% |

**Laboratory confirmed influenza**

**Figure 1.** Forest plot of pairwise meta-analysis comparing IIV3-HD (intervention) vs. IIV3-SD (control), restricted to matched studies.

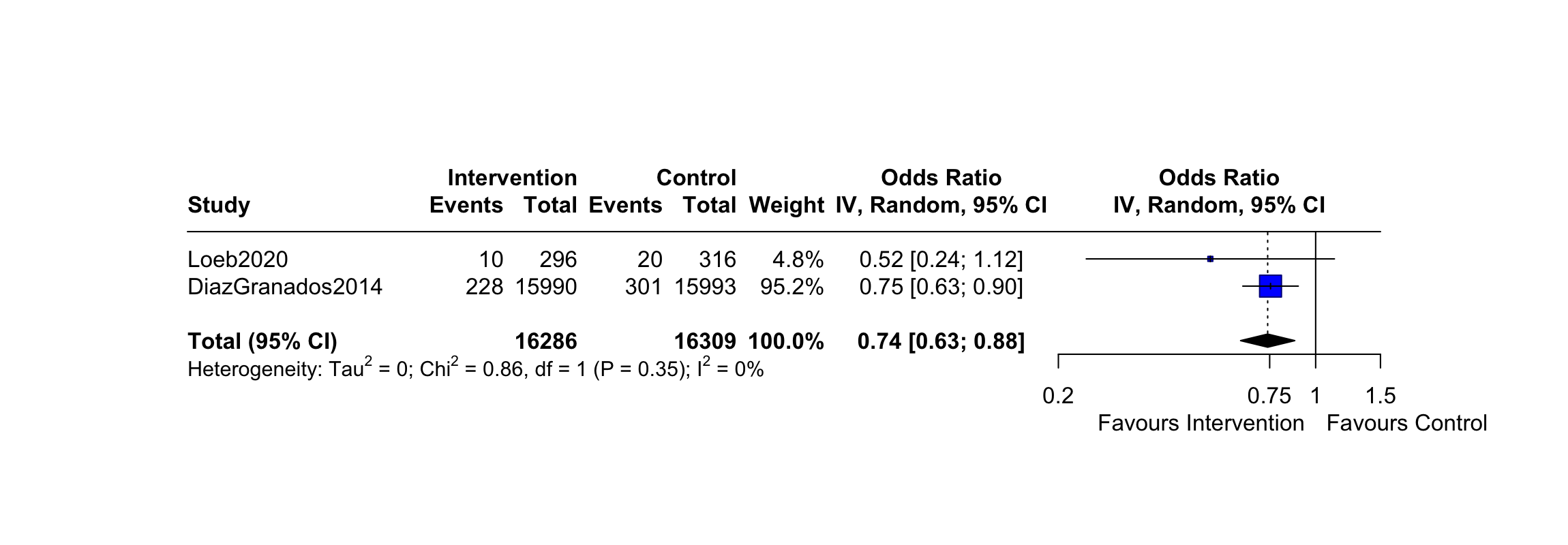

**Figure 2.** Forest plot of pairwise meta-analysis comparing IIV3-HD (intervention) vs. IIV3-SD (control), restricted to mismatched studies

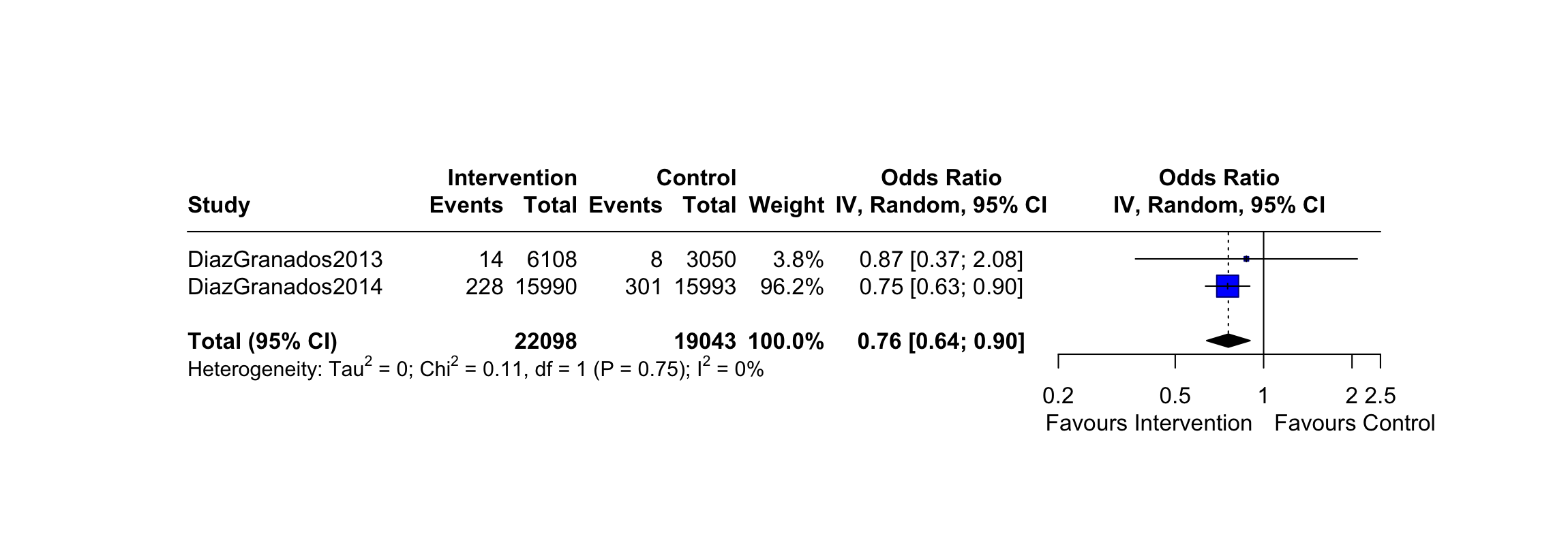
