## Additional file 3 for "Comparing trivalent and quadrivalent seasonal influenza vaccine efficacy in persons 60 years of age and older: A systematic review and network meta-analysis"

**GRIPP2 Checklist – Long Form**

| **Section and topic** | **Item** | **Reported on page No** |
| --- | --- | --- |
| Section 1: Abstract of paper | | |
| 1a: Aim | Report the aim of the study | 4 |
| 1b: Methods | Describe the methods used by which patients and the public were involved | 3 |
| 1c: Results | Report the impacts and outcomes of PPI in the study | N/A |
| 1d: Conclusions | Summarise the main conclusions of the study | 5 |
| 1e: Keywords | Include PPI, “patient and public involvement,” or alternative terms as keywords | 5 |
| Section 2: Background to paper | | |
| 2a: Definition | Report the definition of PPI used in the study and how it links to comparable studies | N/A |
| 2b: Theoretical underpinnings | Report the theoretical rationale and any theoretical influences relating to PPI in the study | N/A |
| 2c: Concepts and theory development | Report any conceptual or theoretical models, or influences, used in the study | N/A |
| Section 3: Aims of paper | | |
| 3: Aim | Report the aim of the study | 7 |
| Section 4: Methods of paper | | |
| 4a: Design | Provide a clear description of methods by which patients and the public were involved | 8 |
| 4b: People involved | Provide a description of patients, carers, and the public involved with the PPI activity in the study | 8 |
| 4c: Stages of involvement | Report on how PPI is used at different stages of the study | 8 |
| 4d: Level or nature of involvement | Report the level or nature of PPI used at various stages of the study | 8 |
| Section 5: Capture or measurement of PPI impact | | |
| 5a: Qualitative evidence of impact | If applicable, report the methods used to qualitatively explore the impact of PPI in the study | N/A |
| 5b: Quantitative evidence of impact | If applicable, report the methods used to quantitatively measure or assess the impact of PPI | N/A |
| 5c: Robustness of measure | If applicable, report the rigour of the method used to capture or measure the impact of PPI | N/A |
| Section 6: Economic assessment | | |
| 6: Economic assessment | If applicable, report the method used for an economic assessment of PPI | N/A |
| Section 7: Study results | | |
| 7a: Outcomes of PPI | Report the results of PPI in the study, including both positive and negative outcomes | N/A |
| 7b: Impacts of PPI | Report the positive and negative impacts that PPI has had on the research, the individuals involved (including patients and researchers), and wider impacts | N/A |
| 7c: Context of PPI | Report the influence of any contextual factors that enabled or hindered the process or impact of PPI | N/A |
| 7d: Process of PPI | Report the influence of any process factors, that enabled or hindered the impact of PPI | N/A |
| 7ei: Theory development | Report any conceptual or theoretical development in PPI that have emerged | N/A |
| 7eii: Theory development | Report evaluation of theoretical models, if any | N/A |
| 7f: Measurement | If applicable, report all aspects of instrument development and testing (eg, validity, reliability, feasibility, acceptability, responsiveness, interpretability, appropriateness, precision) | N/A |
| 7g: Economic assessment | Report any information on the costs or benefit of PPI | N/A |
| Section 8: Discussion and conclusions | | |
| 8a: Outcomes | Comment on how PPI influenced the study overall. Describe positive and negative effects | 21-22 |
| 8b: Impacts | Comment on the different impacts of PPI identified in this study and how they contribute to new knowledge | 21-22 |
| 8c: Definition | Comment on the definition of PPI used (reported in the Background section) and whether or not you would suggest any changes | N/A |
| 8d: Theoretical underpinnings | Comment on any way your study adds to the theoretical development of PPI | N/A |
| 8e: Context | Comment on how context factors influenced PPI in the study | N/A |
| 8f: Process | Comment on how process factors influenced PPI in the study | N/A |
| 8g: Measurement and capture of PPI impact | If applicable, comment on how well PPI impact was evaluated or measured in the study | N/A |
| 8h: Economic assessment | If applicable, discuss any aspects of the economic cost or benefit of PPI, particularly any suggestions for future economic modelling. | N/A |
| 8i: Reflections/critical perspective | Comment critically on the study, reflecting on the things that went well and those that did not, so that others can learn from this study | 22 |
