## Additional file 4 for "Comparing trivalent and quadrivalent seasonal influenza vaccine efficacy in persons 60 years of age and older: A systematic review and network meta-analysis"

**Plain Language Summary**

**Objective:** To assess how different types of influenza vaccines compare in adults aged 60 years and older.

To decide whether how well the influenza vaccines work varies by risk group i.e.: between sex, previous vaccinations and age greater or equal to 80 years old.

Many different flu vaccines are available for older individuals, and in various formulations. However, the lack of high-quality research analyzing these vaccines makes it hard for policy makers and healthcare providers to make decisions and recommendations on which vaccines to use for preventing influenza and its complications.

This systematic review, which summarized published research studies on this topic, included randomized controlled trials on adults 60 years and older who received an influenza vaccine that was approved for use in Canada or the United States.

The systematic review assessed several outcomes categorized as main and most critical measurements (primary) and additional measurements of interest (secondary):

1. Primary Outcomes: Laboratory confirmed Influenza (LCI) and Influenza like illnesses (ILI).
2. Secondary Outcomes: number of vascular events, hospitalization for acute respiratory infection (ARI) and ILI, inpatient hospitalization, emergency room (ER) visit for ILI, outpatient visit, and death, among others.

The research studies were screened by two independent reviewers. The reviewers excluded observational studies, experimental vaccines and vaccines not licensed in Canada or United States.

After the screening of articles, 41 were included and involved 206,032 people.

The 2 reviewers then collected the following information from the included studies: study design, country of the study, how long patients were followed, whether data was from one site or many, characteristics of patients (e.g., average age, sex, frailty), type of vaccine, dose and whether the vaccine was adjuvanted. Adjuvanted means the vaccine is boosted to provide greater immune boosting qualities. Influenza strains included in the vaccine were also collected. If there were 2 inflluenza A strains and 1 B strain in the vaccine that would be a Trivalent vaccine. Two influenza A strains and 2 B strains would be a Quadrivalent vaccine.

The final part of this data collection involved the outcomes of the different vaccines. These included number of patients who became infected with influenza and its complications.

The risk of bias was then assessed. This was confirmed by 2 researchers and in the case of disagreement a third opinion was given.

The data was then analysed using advanced statistical techniques and a summary of the outcomes and how certain the outcome was produced.

**Conclusion:**

Results found that influenza vaccines were effective in preventing lab confirmed influenza compared with placebo. In particular, RIV which is Recombinant Influenza Vaccine and a synthetically made vaccine, was among the most effective vaccines in preventing LCI and minimizing all-cause deaths.

High Dose Trivalent Inactivated Influenza Vaccine (IIV3-HD) was among the most efficacious vaccines for preventing LCI and can reduce hospitalizations for ILI and ARI in older adults. This vaccine is produced by using egg yolk as a medium to grow strains of influenza vaccines. People with egg allergies may use both the trivalent and quadrivalent vaccines.

The studies also showed the quadrivalent inactivated influenza vaccine (IIV4-Adj) may reduce number of vascular events compared with others.

However, vaccine efficacy may vary depending on population (e.g., frailty, comorbidities) virus strain matching and study characteristics. More high quality RCTs are necessary to evaluate the relationship between these factors and vaccine efficacy, such as seasonal vaccines specifically based on RIV and IIV4 formulations.
